# Intensive care unit admissions in Germany associated with atmospheric factors: a regional time-series analysis

**DOI:** 10.64898/2026.02.27.26347246

**Authors:** Katharina Sasse, Christian Merkenschlager, Michael Johler, Till Baldenius, Patrik Dröge, Christian Günster, Thomas Ruhnke, Pablo Escrihuela Branz, Lucas Pröll, Bastian Wein, Saskia Hettich, Yevgeniia Ignatenko, Taner Öksüz, Iñaki Soto-Rey, Elke Hertig

**Author notes:** **Correspondence:** Katharina Sasse.

## Abstract

**Introduction:** Atmospheric conditions under climate change constitute an increasing health burden and raise pressure on health care systems. Intensive care units (ICUs) are particularly vulnerable due to their low buffer capacity and high utilization rates. This study aimed to examine the association between atmospheric exposures and ICU admissions for intensive care relevant diseases across Germany.

**Methods:** We analyzed daily time-series data on ICU admissions from AOK health insurances and atmospheric data in the period 2009 to 2023. Using principal component analysis, we clustered Germany into eleven human-bioclimatic regions. For each atmospheric predictor, the optimal time lag was selected, and disease-specific predictor sets were defined while accounting for multiple testing. In each region, we fitted quasi-Poisson generalized additive models to estimate disease-specific ICU admission risks associated with atmospheric exposures. Models were stratified by age (<60, ≥60 years) and sex. Model performance was assessed using (weighted) explained deviance (WED).

**Results:** Over the 15-year study period, on average 2,526 ICU admissions per 100,000 AOK insured persons per year were recorded in Germany (44% women, 74.3% aged ≥60 years). Pressure-related variables were most frequently associated with ICU admissions (p ≤ 0.05). Sea-level pressure demonstrated a significant association in 29.3% of the models. Heat-related variables, including tropical nights (18.3%) and summer days (17.8%), were significantly associated with ICU admissions in Germany. Among air pollutants, ozone showed the highest proportion of significant associations (15.6%). Significant associations for heat-related atmospheric variables were observed more frequently among men and persons ≥60 years. Mean model performance across all models was 10% WED. The models for ICU admissions due to myocardial infarction (NSTEMI) in the East German Lowlands demonstrated the best model performance (WED: 49%).

**Discussion:** Atmospheric factors were associated with ICU admissions, and these associations differed by region, disease, and age-sex group. Pressure- and heat-related factors were the most relevant atmospheric factors. These findings indicate that atmospheric factors have the potential to adversely impact intensive health care preparedness and delivery.

## 1 Introduction

Urbanization, economic development, and industrial growth have contributed to increasing levels of air pollution and changes of the atmospheric environment over recent decades. At the same time, climate change is expected to increase the frequency, intensity, and duration of atmospheric hazards, including heat waves and other extreme weather events [1, 2]. These environmental changes represent important stressors for human health and are associated with substantial increases in morbidity and mortality [3, 4]. A growing body of evidence indicates that there is an influence on the occurrence and exacerbation of a wide range of diseases, including respiratory, cardiovascular, gastrointestinal, neurological, and trauma-related diseases [5]. As many of these diseases frequently require hospital-based treatment, climate change-related health impacts are expected to place an increasing burden on health care systems. Vulnerable populations, particularly older adults, children, and individuals with pre-existing medical conditions, are disproportionately affected by atmospheric hazards and extreme weather events [4, 6]. Consequently, a comprehensive understanding of atmosphere-health relationships is essential for quantifying future health care demands and supporting the development of climate-resilient health care systems and infrastructure [7].

Numerous studies have reported associations between atmospheric stressors and increased rates of mortality and hospital admissions [2, 8]. However, considerably less attention has been paid to the potential implications for intensive care medicine, despite intensive care units (ICUs) representing one of the most resource-intensive and capacity-limited sectors of health care. Patients requiring intensive care are often characterized by severe acute illnesses, multiple comorbidities, and heightened physiological vulnerability, making them particularly susceptible to environmental stressors. Moreover, ongoing climate change may translate into higher demand for critical care services, potentially exacerbating existing capacity constraints. In Germany, the average ICU utilization rate of 81.6% leaves limited reserve capacity to accommodate sudden increases in demand associated with atmospheric conditions [9]. This challenge is further intensified by the persistent shortage of specialized healthcare personnel [10]. Understanding how atmospheric factors are associated with ICU admissions has therefore a substantial clinical and public health relevance, as it may help identify vulnerable patient groups, anticipate future health care demands, and support strategic resource planning in intensive care.

Despite growing evidence on climate-related health effects, nationwide assessments of atmospheric influences on ICU admissions remain limited, particularly regarding regional and disease-specific differences. This study addresses this gap by linking region-specific daily ICU admissions with atmospheric data across Germany. The objectives are to analyze atmosphere-ICU associations across Germany, quantify regional heterogeneity, and identify diseases and age-sex-specific subgroups with heightened susceptibility to atmospheric conditions.

## 2 Materials and Methods

### 2.1 Study sample and health data

Daily time-series data of intensive care admissions derived from anonymized, nationwide routine data from the German health care system was collected for the years 2009-2023. The effects of the Covid-19 pandemic on ICU admissions were observed, particularly in respiratory diseases (see Supplementary Figure S1). However, the impact on intensive care as a whole was negligible (Figure 2). Moreover, a long-term downward trend in admissions and health care utilization has been observed [11]. Consequently, the pandemic years were not considered separately. The 15-year health data was provided by the AOK Research Institute (WIdO), derived from statutory health insurance data of eleven regional AOK insurances. The study sample was stratified by sex and two age groups (<60 and ≥60 years old). The choice for only two age groups was motivated by the need to standardize the health insurance-based subsample of about 30% of the population to the total population of Germany in a follow-up study. Thus, four subgroups were analyzed: female ≥60 years (FE), male ≥60 years (ME), female <60 years (FY), and male <60 years (MY). Repeated admissions of one individual were not accounted for. All admissions were treated as independent admissions.

The operationalization of intensive care required a broader definition, given that not all hospitals have dedicated ICU or fulfil the billing criteria for complex intensive care treatment. Therefore, the operation and procedure (OPS) code for intensive care complex treatment, the OPS code for intensive care patient monitoring, or the department code of ICU was used as a selection criterion (Supplementary, Table S1& S2). The study included admissions of the 13 most common diseases in the ICU, accounting for 46% of all ICU admissions [5, 12, 13]. The diseases were classified using the codes of the International Statistical Classification Of Diseases And Related Health Problems, 10th revision, German Modification (ICD-10-GM) (Supplementary, Table S3). They were grouped into the following categories: cardiovascular diseases (heart failure, non-ST elevation myocardial infarction (NSTEMI), ST elevation myocardial infarction (STEMI), tachycardia, pulmonary artery embolism), respiratory diseases (pneumonia, chronic obstructive pulmonary disease (COPD)), neurological diseases (ischemic insult, transient ischemic attack, non subdural hemorrhage, epilepsy), internal diseases (sepsis) and traumatic diseases (trauma). For a few disease categories, the term “ICU admissions” was defined more broadly than the ICU definition given above. For cardiovascular diseases, continuous rhythm monitoring is essential. Therefore, admissions treated in settings such as chest pain units were also classified as ICU admissions. Without this adjustment, a considerable proportion of cardiovascular admissions requiring intensive monitoring might not have been captured. Similarly, for respiratory diseases, admissions receiving ventilation outside of an ICU were included to ensure that all admissions requiring intensive care-level treatment were represented (Supplementary Table S3). For neurological diseases (except epilepsy), admissions were categorized separately according to whether treatment was provided in an ICU or in a specialized stroke unit (Supplementary, Table S4), resulting in the consideration of 16 outcome types in total. Prior to statistical model building, the daily health data was analyzed regarding erroneous values within the ICD-10-GM coding system, internal validity as well as with respect to completeness, homogeneity, variability, outliers and trends [14, 15].

### 2.2 Human-bioclimatic Regionalization

To account for regional differences and to define internally homogeneous exposure clusters, Germany was divided into human-bioclimatic regions (HBC-regions). The regionalization was based on seasonally resolved climatic variables, which are related to human health. Data was derived from the ERA5-Land reanalysis dataset provided by the Copernicus Climate Data Store [16]. For the classification of the HBC-regions, a spatial principal component analysis was used to identify the optimal number of regions, and the grid boxes were assigned to the determined number of HBC-regions by means of a k-means cluster analysis. The regionalization identified six main regions and, given the presence of still some noticeable intra-regional variations, three further subregions using the aforementioned methods were built. Overall, the regionalization resulted in eleven distinct, bioclimatic consistent HBC-regions, characterizing the different human-bioclimatic conditions in Germany on regional scales (Figure 1). The eleven HBC-regions were mapped to their corresponding ZIP codes, allowing the daily health data to be assigned to the respective regions.

**Figure 1.**
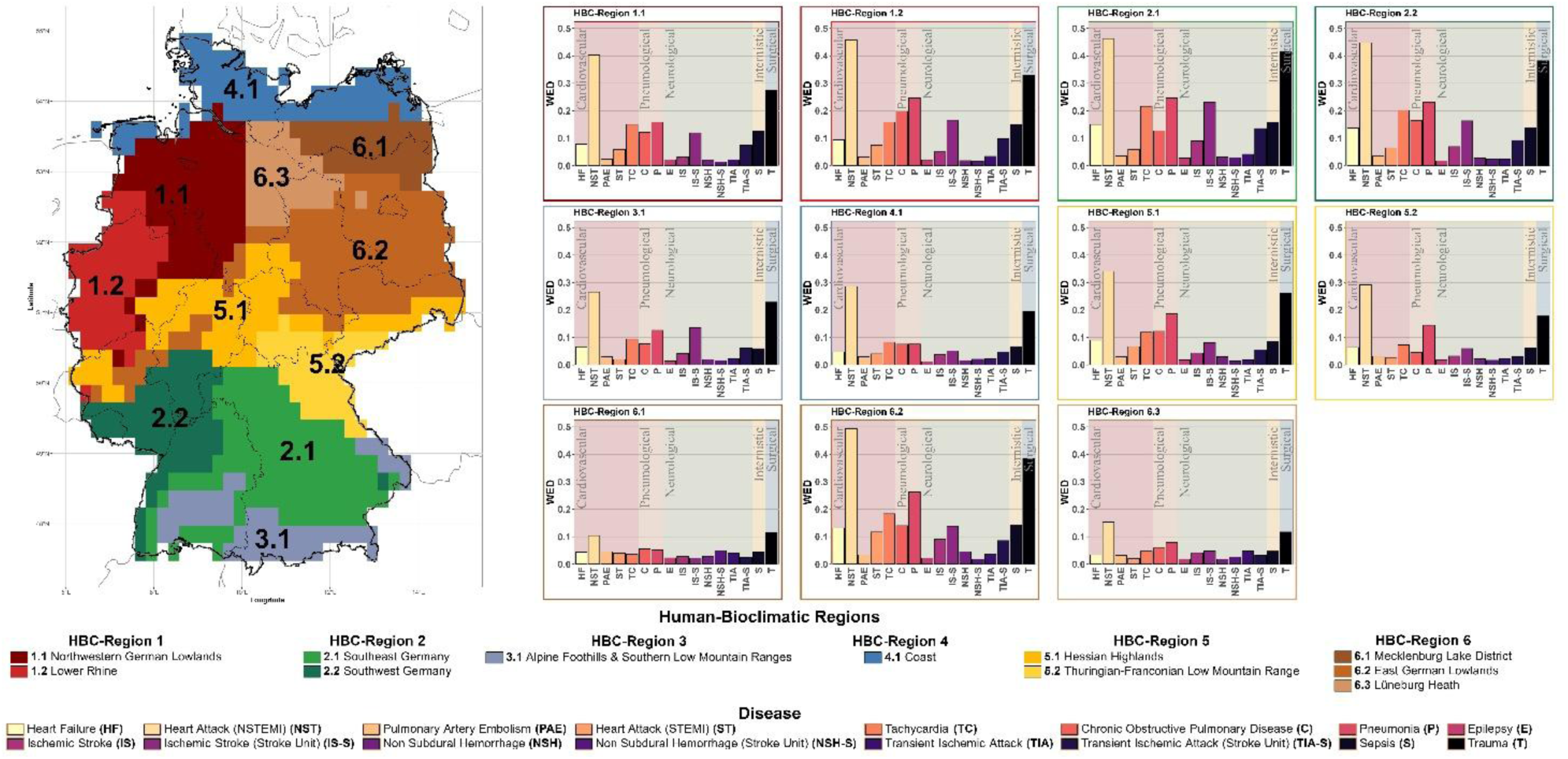
Human-bioclimatic (HBC) regions of Germany and model performance (weighted explained deviance) per HBC-region and disease **Alt text Figure 1:** Two-part figure. Left: Map of Germany divided into eleven human-bioclimatic (HBC) regions. Right: One bar chart per HBC-region showing model performance for each disease, grouped by disease category. Performance varies between diseases and HBC-regions.

### 2.3 Atmospheric data

For assessing the atmospheric impact on intensive care, daily data of multiple atmospheric variables and indices was collected. Daily meteorological data was obtained from the quality-controlled ERA5 reanalysis data of the European Centre for Medium-Range Weather Forecasts (ECMWF), and air pollution data from the Copernicus Atmosphere Monitoring Service (CAMS) [16–19]. In total, 31 atmospheric variables and indices were extracted (Supplementary, Table S5). Averaging atmospheric exposures within the HBC-regions inevitably led to a reduction in the spatial resolution of the atmospheric data as well as to a smoothing of extremes. This limitation particularly affected the representation of urban heat island effects as well as the small-scale variability of air quality variables. Thus, regional exposure characterization can be regarded as background exposure, giving an overall conservative exposure assignment.

### 2.4 Statistical analysis

To investigate the associations between daily ICU admissions and atmospheric data, we calculated generalized additive models (GAMs) with quasi-Poisson distribution to account for overdispersion [20, 21]. GAMs are particularly useful for time series analyses that investigate the relationship between climate effects and health, and allow nonparametric adjustments for nonlinear effects [20, 22–24]. In each HBC-region the ICU outcomes stratified according to sex and age were analyzed. In total, 704 different models (11 regions x 16 ICU outcome types x 4 subgroups) were created to establish the regional associations between the daily ICU admissions and atmospheric factors.

To determine the optimal lag in the exposure-outcome models, for all regions, outcome types and subgroups bivariate predictor-response models were built using no lag up to ten days lag for meteorological predictors and up to seven days for air quality predictors [23, 25]. The optimal lag for each atmospheric predictor was selected based on the relative risk (RR) and 95% confidence interval (CI) [26]. Subsequently, for each region, outcome type and subgroup GAMs were established using all predictor variables with their specific lags. Predictor variables remained in the models if the p-value was significant (α ≤ 0.05) after Bonferroni correction. The correction for multiple testing was applied separately within each region-, outcome-, and subgroup-specific model [27, 28]. In order to harmonize predictor sets according to outcome types and to facilitate interpretation, the final predictor set per outcome type consisted of all predictors which were at least significant in one GAM out of 44 in total (11 regions x 4 subgroups). The resulting outcome type-specific predictor sets were subsequently used to fit the final GAMs for each combination of outcome type, subgroup, and HBC-region.

For nonlinear associations, thin plate regression splines with shrinkage were used as the smoothing basis [29, 30]. Models were fitted using restricted maximum likelihood (REML), which provides robust estimation of smoothing parameters and is particularly suitable for models with overdispersed count data [30]. The shrinkage approach (select = TRUE) additionally allowed terms with little explanatory value to be penalized towards zero, thereby reducing the influence of irrelevant predictors [30]. Next to atmospheric predictors, the models controlled for seasonality effects through sinusoidal and cosinusoidal components, time trend as well as weekday versus weekend [26, 31–34]. Seasonality and time trend were represented in the models using thin plate regression splines.

Collinearity among the atmospheric predictors was assessed using Spearman’s rank correlation coefficient. Predictor pairs with an absolute correlation coefficient of |ρ| ≥ 0.7 were considered as collinear. In cases of collinearity, the predictor showing the stronger statistical association with the respective outcome, as indicated by the lower p-value in the regression analysis, was retained, while the less strongly associated predictor was excluded from the model [35].

Explained deviance (ED) was used to check model performance. To summarize subgroup-specific performance for each region and outcome type, weighted explained deviance (WED) was calculated by weighting the EDs of the subgroups according to their proportion in the case numbers of a region and outcome. Differences between subgroups were assessed descriptively by comparing the WED, ED and the proportion of atmospheric predictors with a significant association (p ≤ 0.05). These comparisons are therefore exploratory and do not allow conclusions on statistically significant differences between subgroups.

For the regional outcome type with the highest WED, predictor-specific log-RRs from the corresponding subgroup-specific models were compared assuming independence between subgroups. Heterogeneity across the four subgroups was assessed using Cochran’s Q test and quantified by I².

The resulting p-values were adjusted across predictors using the Benjamini-Hochberg procedure [36]. For predictors exhibiting significant heterogeneity (adjusted p < 0.05), pairwise z-tests were performed, comparing sexes within age groups and age groups within sexes. Results are reported as ratios of RRs with corresponding 95% CIs and p-values.

The main statistical analyses were performed using the package *mgcv* in R (Version 4.5.1) and RStudio (Version 2025.05.1.513) [20, 23]. The study was conducted in accordance to the Declaration of Helsinki and the reporting follows the RECORD guidelines for studies using routine health data [37].

## 3 Results

### 3.1 Health data

The sample comprised on average 2,526 standardized ICU admissions per 100,000 AOK insured persons per year over the 15-year study period (a total of 9,970,548 ICU admissions) (Figure 2, Supplementary: Standardized ICU admissions per outcome type, Figure S1). 44% of the admissions were female and 74.3% of the admitted persons were ≥60 years old. ICU admissions were most frequent in the ME subgroup and least frequent in the FY subgroup. Compared with the FY subgroup, the ME subgroup exhibited approximately three times as many ventilation days. In contrast, obesity was more common among people younger than 60 years than among those aged ≥60 years. The highest proportion of obese people was observed in the FY subgroup (Table 1). Trauma accounted for the highest number of ICU admissions among all outcome types. In the ≥60 year subgroups, ischemic stroke represented one of the most frequent outcome types, whereas pneumonia was more prevalent in the <60 year subgroups (Table 1). An almost continuous decrease in ICU admissions was observed across all HBC-regions (except 6.3) from 2017 onwards. Over the entire study period, HBC-region 6.1 (Mecklenburg Lake District in northeastern Germany) consistently showed the highest ICU admission rate per 100,000 AOK insured people (Figure 2).

**Figure 2.**
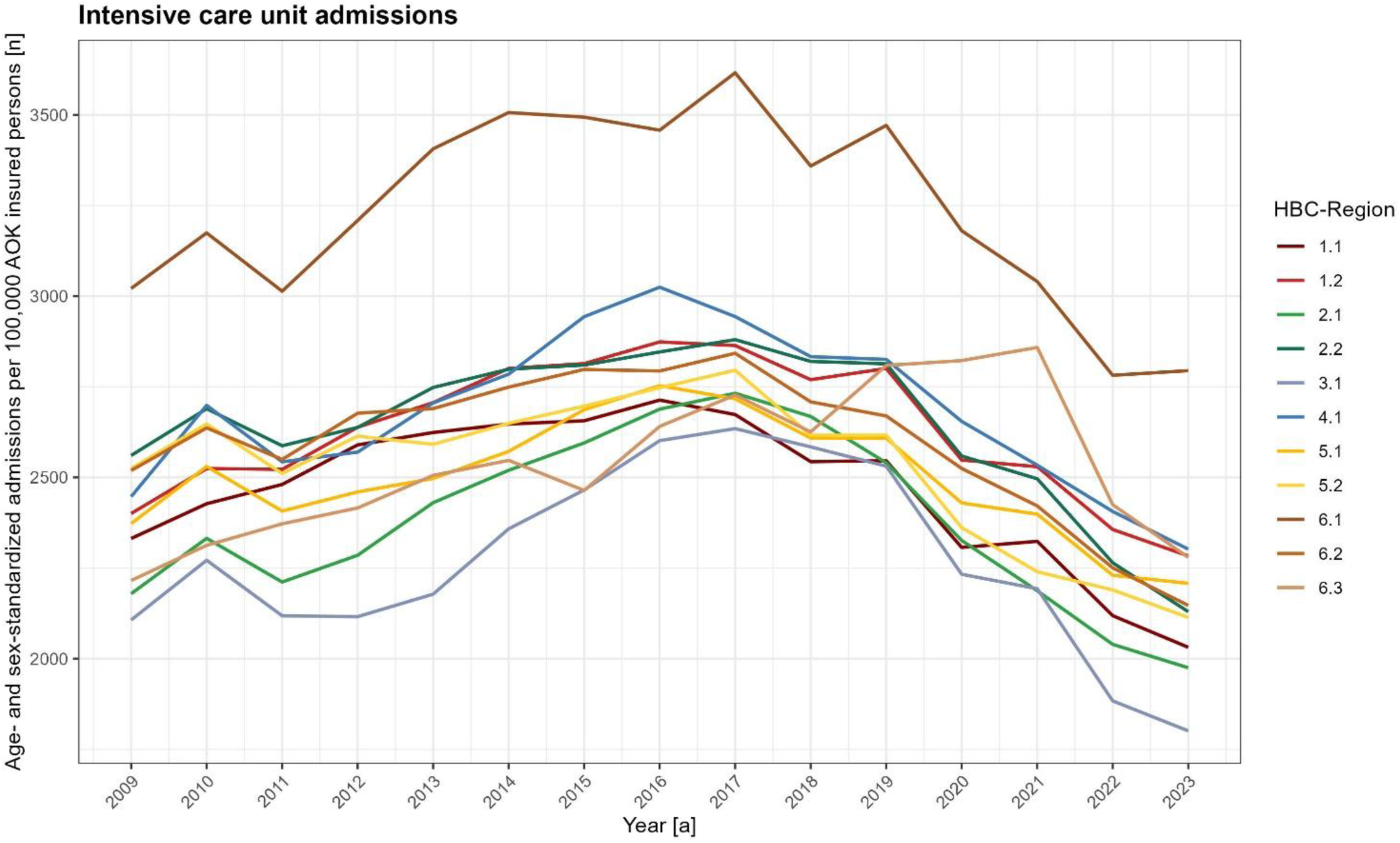
Annual intensive care unit (ICU) admissions per 100,000 AOK insured persons by human-bioclimatic (HBC) region, 2009–2023, standardized for age and sex to the AOK insured population **Alt text Figure 2:** Line chart with one line per human-bioclimatic region, showing annual intensive care unit admissions per 100,000 insured persons from 2009 to 2023. Admissions follow an inverted U-shaped trend in all regions.

**Table 1.**
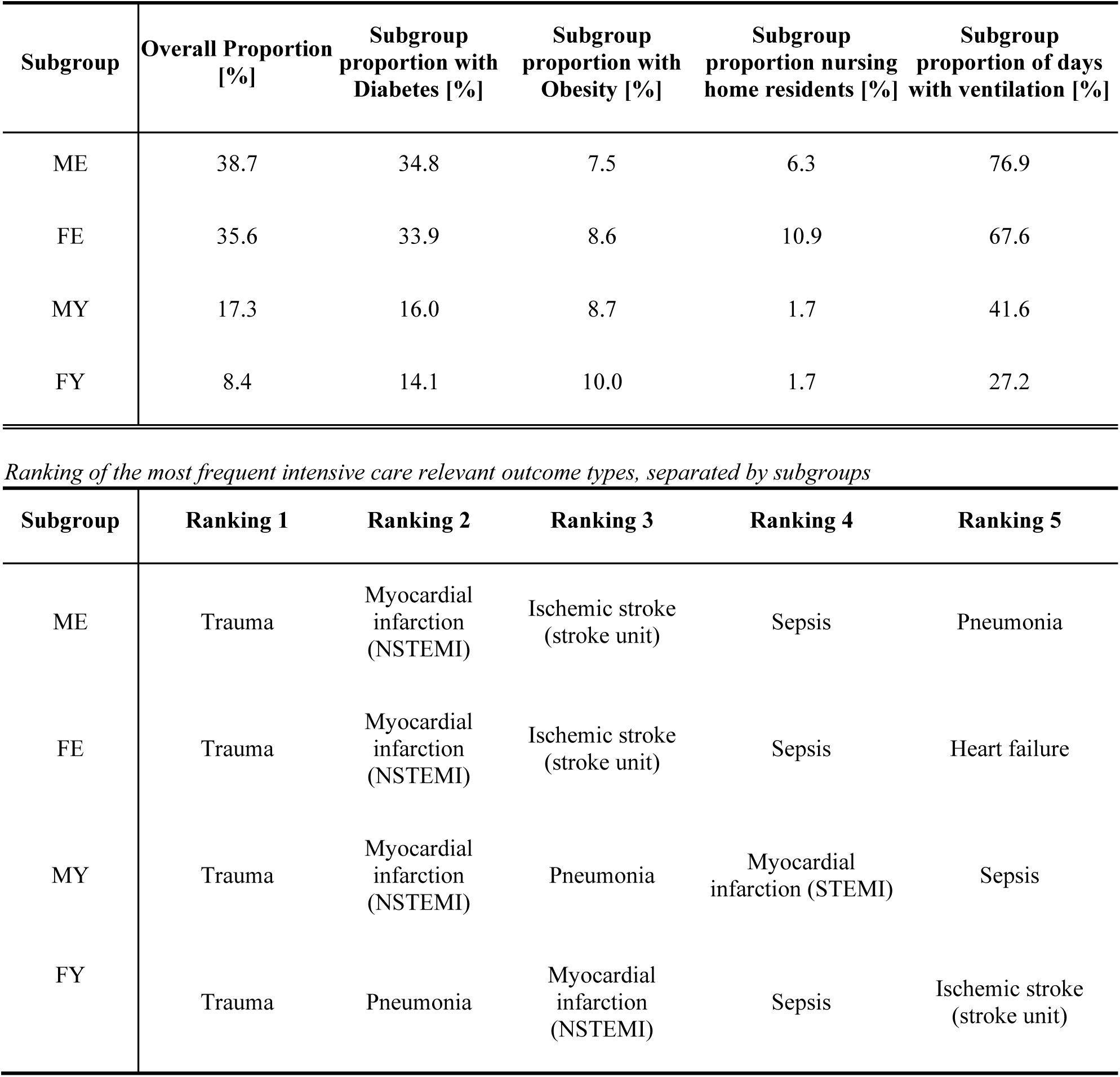
Descriptive statistics of the study sample and ranking of the most frequent intensive care related outcome types, separated by subgroups (ME: males ≥60 years, FE: females ≥60 years, MY: males <60 years, FY: females <60 years)

| Subgroup | Overall Proportion [%] | Subgroup proportion with Diabetes [%] | Subgroup proportion with Obesity [%] | Subgroup proportion nursing home residents [%] | Subgroup proportion of days with ventilation [%] |
|---|---|---|---|---|---|
| ME | 38.7 | 34.8 | 7.5 | 6.3 | 76.9 |
| FE | 35.6 | 33.9 | 8.6 | 10.9 | 67.6 |
| MY | 17.3 | 16.0 | 8.7 | 1.7 | 41.6 |
| FY | 8.4 | 14.1 | 10.0 | 1.7 | 27.2 |
| **Subgroup** | **Ranking 1** | **Ranking 2** | **Ranking 3** | **Ranking 4** | **Ranking 5** |
| ME | Trauma | Myocardial infarction (NSTEMI) | Ischemic stroke (stroke unit) | Sepsis | Pneumonia |
| FE | Trauma | Myocardial infarction (NSTEMI) | Ischemic stroke (stroke unit) | Sepsis | Heart failure |
| MY | Trauma | Myocardial infarction (NSTEMI) | Pneumonia | Myocardial infarction (STEMI) | Sepsis |
| FY | Trauma | Pneumonia | Myocardial infarction (NSTEMI) | Sepsis | Ischemic stroke (stroke unit) |

### 3.2 Atmospheric data

A comparison of regional atmospheric characteristics revealed pronounced differences across the eleven HBC-regions (Supplementary Figures S2-S8). Distinct regional patterns emerged, particularly regarding cold exposure, heat exposure, and air pollution. The most cold-exposed regions were observed in HBC-regions 5.2 (Thuringian-Franconian Low Mountain Range) and 3.1 (Alpine Foothills & Southern Low Mountain Ranges). HBC-region 5.2 exhibited the highest number of ice days (average: 27 days/year; daily T_max_ < 0°C) and the lowest mean wind chill index (WCI, mean ± standard deviation: 5.4 ± 7.4°C). Similarly, HBC-region 3.1 showed pronounced cold exposure, with the highest number of frost days (average: 95.2 days/year; daily T_min_ < 0°C) and an absence of tropical nights (daily T_min_ > 20°C). In contrast, HBC-region 2.2 (southwest Germany) represented the most heat-exposed region, with the highest number of tropical nights (average: 1.5 nights/year, maximum: 6 nights in 2018), summer days (average: 43.5 days/year; daily T_max_ > 25°C), and hot days (average: 9.6 days/year; daily T_max_ > 30°C), as well as the highest mean wet-bulb globe temperature (WBGT: 13.6 ± 7.5°C). The highest daily mean temperature was observed in HBC-region 1.2 (10.7 ± 6.8°C). Overall, the air quality in Germany improved over the study period. Only ozone levels continued to rise in all regions over the 15 years (Supplementary, Figure S2). Even though the air pollution exposure on the chosen regional scale does only reflect average background conditions, distinct air pollution characteristics could be noticed. Elevated air pollutant concentrations were primarily observed in Lower Rhine and southwest Germany (HBC-regions 1.2, 2.2). The Lower Rhine region exhibited the highest daily mean concentrations of nitrogen dioxide (NO₂) (8.2 ± 3.7ppbv) and particulate matter (PM_10_) (19.5 ± 6.6µg/m³), while southwest Germany showed the highest daily mean PM_2.5_ concentration (13.5 ± 6.5µg/m³). In contrast, the highest daily mean ozone concentration was observed in HBC-region 5.2 (36.8 ± 12.4ppbv). Northern regions exhibited the greatest exposure to atmospheric pressure variability, with HBC-regions 4.1 and 6.3 experiencing the highest frequency of day-to-day pressure changes exceeding 10hPa (approximately 15 days per year). Likewise, the greatest number of wind days was recorded in north-eastern Germany, particularly in HBC-regions 6.2, 4.1, and 1.1, with up to 18 wind days annually. Conversely, the highest mean sea-level pressure (SLP) was observed in southern Germany, namely in HBC-regions 3.1 (1017.4hPa) and 2.1 (1017.2hPa).

### 3.3 Health-atmosphere associations

The outcome type-specific predictor sets included between six and 18 atmospheric predictors (Table 2). Across all models, the three pressure-related variables (SLP, SLPD, wind day) showed most frequently statistically significant associations (p ≤ 0.05) with ICU admissions. SLP was significantly associated with ICU admissions in 29.3% of all models (206/704 models), wind day in 24.9% (175/704 models) and sea-level pressure difference (SLPD) in 20.2% (142/704 models). Heat-related predictors followed, in particular tropical nights (18.3% of models) and summer days (17.8%). Among air pollutants, ozone showed the highest proportion of significant associations (15.6% of models). Cold wave day was the only atmospheric predictor with no significant association in any model. Table 3 summarizes the significant associations between atmospheric predictors and ICU admissions. All outcome-specific results are provided in the supplement and show that the atmospheric factors act on specific outcomes. Pressure- and cold-related predictors were predominantly associated with respiratory outcomes, whereas heat-related variables were mainly associated with neurological outcomes. Cardiovascular outcomes were associated with a broad range of atmospheric factors (Supplementary, Table S6).

**Table 2.** Overview of the selected predictor set per outcome type.

| **Outcome type** | **Predictor set** |
|---|---|
| Myocardial infarction (STEMI) | Sea-level pressure (SLP), Tropical night (TN), Cold wave Duration (CWDur), Hot day (HD), ozone (O<sub>3</sub>), Summer day (SD) |
| Myocardial infarction (NSTEMI) | SLP, WCI, Wind day (WD), Frost day (FD), Heat stress wave duration (HSWDur), Heat stress wave intensity (HSWI), Heatwave (HW), Particulate matter (PM<sub>10</sub>) |
| Heart failure | SLP, Cold stress wave intensity (CSWI), SD, Cold stress wave day (CSWDay), Heat wave duration (HWDur), PM<sub>10</sub>, Daily mean temperature (T), TN, Wet-bulb globe temperature (WBGT) |
| Pulmonary artery embolism | WD, O<sub>3</sub>, TN, Heat wave intensity (HWI), Ice day (ID), SLP, Heat stress wave day (HSWDay), HSWI, HWDur, PM<sub>10</sub>, seal-level pressure difference (SLPD), SD, Wind chill index (WCI) |
| Tachycardia | TN, WD, CWDur, HD, Heat wave day (HWDay), HWI, O<sub>3</sub>, SLP, WCI |
| Pneumonia | T, TN, WCI, O<sub>3</sub>, Cold stress wave duration (CSWDur), CWDur, Cold wave intensity (CWI), Nitrogen dioxide (NO<sub>2</sub>), SLP, CSWI, Cold wave (CW), FD, HSWI, SLPD, WD |
| Chronic obstructive pulmonary disease (COPD) | O<sub>3</sub>, Cold stress wave (CSW), PM<sub>10</sub>, WBGT, CSWI, CW, CWDur, CWI, FD, Heat stress wave (HSW), HWI, ID, SLP, SLPD, T |
| Ischemic stroke | CWI, CSWDur, CSWI, HD, HSWI, O<sub>3</sub>, PM<sub>10</sub>, SLP, SD, T, TN, WD |
| Ischemic stroke (stroke unit) | SLPD, CSW, CSWDur, CSWI, CWDur, HSWDay, Heat stress wave intensity (HSWI), ID, TN, WBGT |
| Epilepsy | FD, O<sub>3</sub>, CSW, CSWI, CWDur, HSWI, HWDay, PM<sub>2.5</sub>, SLPD, WBGT, WD |
| Non subdural hemorrhage | T, WCI, WD, CSWDay, CWI, CSWI, HD, HW, HWDur, O<sub>3</sub>, PM<sub>2.5</sub>, SLP, SLPD, SD, TN, WBGT |
| Non subdural hemorrhage (stroke unit) | TN, O<sub>3</sub>, WD, HD, HWI, WBGT, WCI |
| Transient ischemic attack | HW, SLPD, TN, CSW, CSWDay, CW, O<sub>3</sub>, SD, WBGT |
| Transient ischemic attack (stroke unit) | NO<sub>2</sub>, CSWDur, FD, HSW, PM<sub>10</sub>, SLP, SD, WBGT, WCI, WD |
| Sepsis | CSWI, FD, WBGT, HWDay, SLP, SLPD, SD, T, WCI, WD |
| Trauma | WBGT, O<sub>3</sub>, CWDur, FD, SLP, T, CSWI, HD, HSWDay, HW, HWDay, HWI, ID, NO<sub>2</sub>, PM<sub>10</sub>, PM<sub>2.5</sub>, TN, WCI |

**Table 3.**
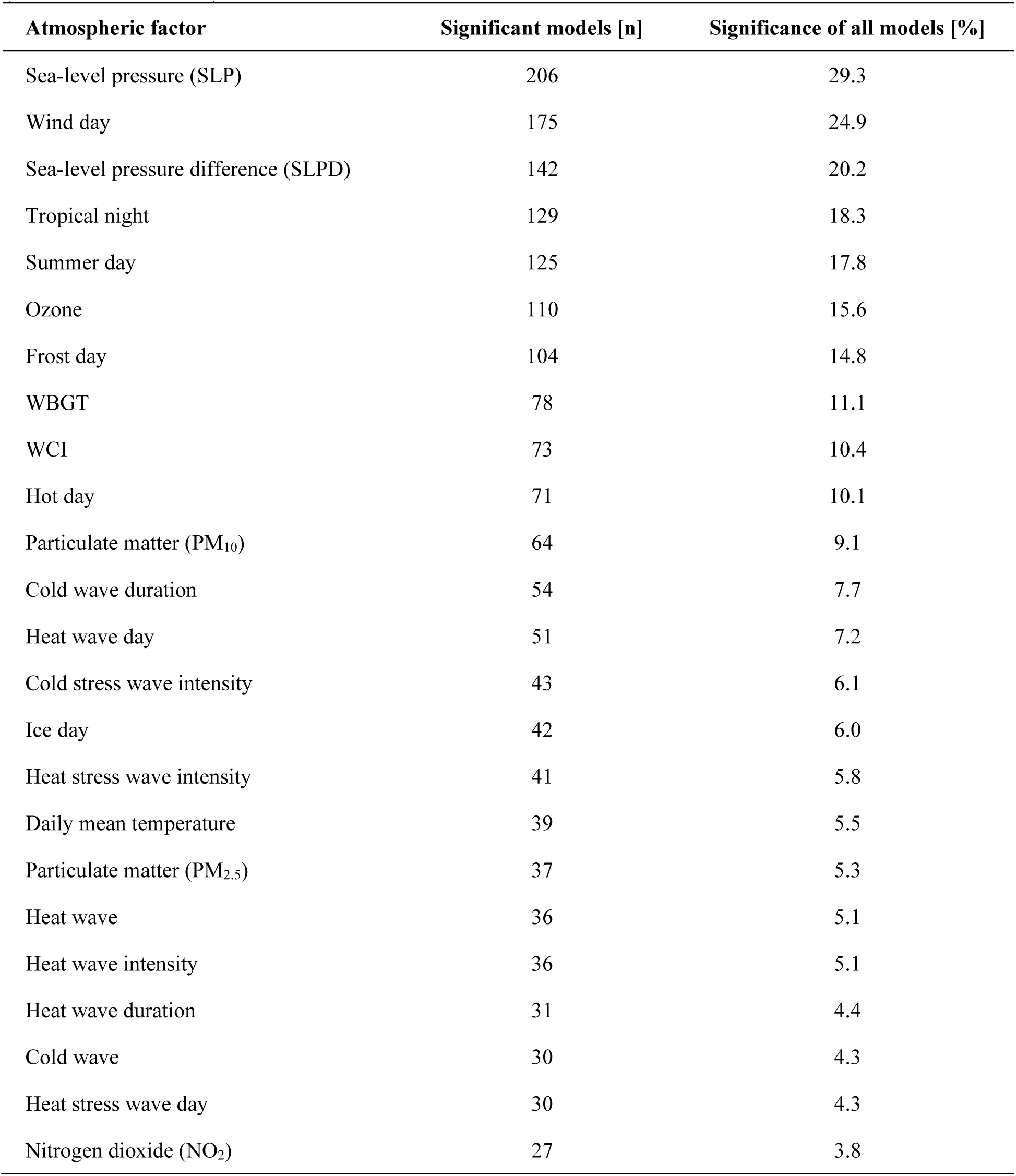
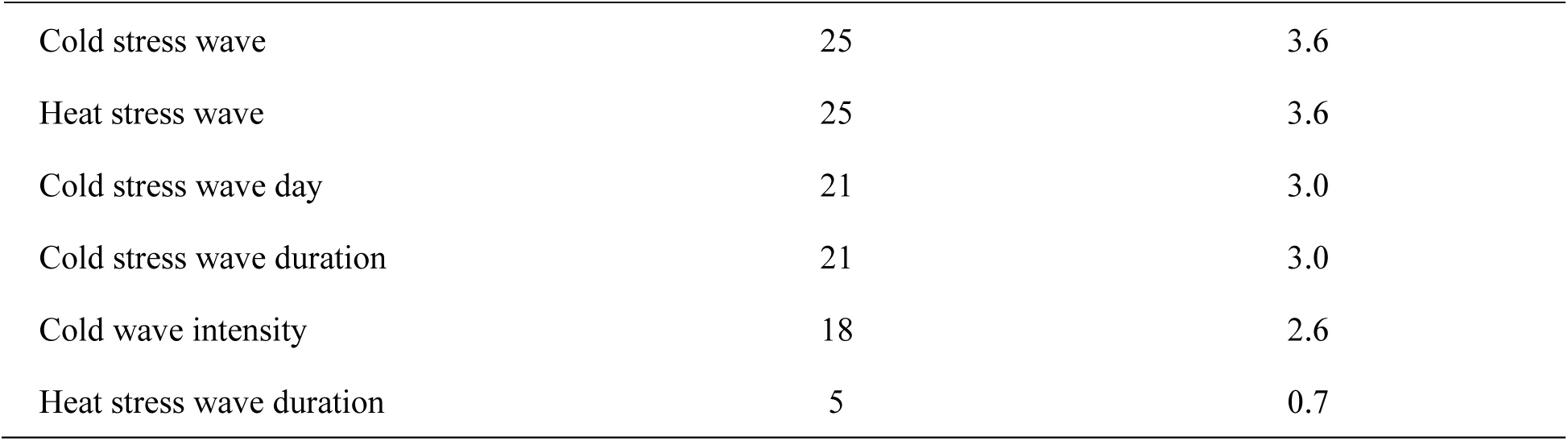
Significant associations (p ≤ 0.05) between daily intensive care unit admissions and atmospheric factors in Germany across outcome types, human-bioclimatic regions and subgroups (704 models in total)

| **Atmospheric factor** | **Significant models [n]** | **Significance of all models [%]** |
|---|---|---|
| Sea-level pressure (SLP) | 206 | 29.3 |
| Wind day | 175 | 24.9 |
| Sea-level pressure difference (SLPD) | 142 | 20.2 |
| Tropical night | 129 | 18.3 |
| Summer day | 125 | 17.8 |
| Ozone | 110 | 15.6 |
| Frost day | 104 | 14.8 |
| WBGT | 78 | 11.1 |
| WCI | 73 | 10.4 |
| Hot day | 71 | 10.1 |
| Particulate matter (PM<sub>10</sub>) | 64 | 9.1 |
| Cold wave duration | 54 | 7.7 |
| Heat wave day | 51 | 7.2 |
| Cold stress wave intensity | 43 | 6.1 |
| Ice day | 42 | 6.0 |
| Heat stress wave intensity | 41 | 5.8 |
| Daily mean temperature | 39 | 5.5 |
| Particulate matter (PM<sub>2.5</sub>) | 37 | 5.3 |
| Heat wave | 36 | 5.1 |
| Heat wave intensity | 36 | 5.1 |
| Heat wave duration | 31 | 4.4 |
| Cold wave | 30 | 4.3 |
| Heat stress wave day | 30 | 4.3 |
| Nitrogen dioxide (NO<sub>2</sub>) | 27 | 3.8 |
| Cold stress wave | 25 | 3.6 |
| Heat stress wave | 25 | 3.6 |
| Cold stress wave day | 21 | 3.0 |
| Cold stress wave duration | 21 | 3.0 |
| Cold wave intensity | 18 | 2.6 |
| Heat stress wave duration | 5 | 0.7 |

#### 3.3.1 Sex- and age-specific differences

Outcome-specific analyses showed differences between sexes (Supplementary Table S7), detailed analysis including statistical effect sizes are given in the Supplementary Tables S8-S23. The proportion of significant associations per atmospheric factor was almost the same for both sexes (differences < 2% for 21 of 30 atmospheric factors, Supplementary Table S7). SLP showed the highest proportion in both sexes (female 27.3%, male 31.2%). Sex differences were most pronounced for SLPD (female 22.7% vs. male 17.6%) and wet-bulb globe temperature (female 9.1% vs. male 13.1%). Heat-related predictors were consistently more frequently significant in males, with tropical nights being the exception (men: 17.0%; women: 19.9%). Conversely, cold stress wave indices, particulate matter (PM_10_) and ozone were more often significant in females. Outcome-specific analyses revealed further sex differences (Supplementary Table S24). For ischemic stroke treated in stroke units, neither cold wave duration nor heat stress wave days were significantly associated with ICU admissions in women, whereas both were significant in 13.6% of the models for men. For trauma, PM_10_ was significantly associated with ICU admissions in 22.7% of the models for women but in none of the models for men.

Overall, a larger proportion of models for persons aged ≥60 years showed significant associations with atmospheric factors. Age-stratified analyses showed that the proportion of significant associations differed mainly for heat-related and wind-related factors (Supplementary Table S7). Heat-related indices were more frequently significant in people aged ≥60 years (e.g., heat wave day: 10.8% vs. 3.7%; summer day: 20.7% vs. 14.8%), as well as for atmospheric pressure predictors (SLP: 31.8% vs. 26.7%; SLPD: 20.7% vs. 19.6%). In contrast, wind days (<60: 29.0% vs. ≥60: 20.7%) and cold wave indices (cold wave: <60 6.2% vs. ≥60 2.3%) were more often significant in people aged <60 years. An examination of outcome-specific differences revealed a significant association between tropical nights and neurological outcomes, particularly among people <60 years (e.g., ischemic stroke: 40.9% vs. 36.4%, non subdural hemorrhage: 45.5% vs. 22.7%), whereas hot days (T_max_ >30 °C) were significantly associated among persons ≥60 years (Supplementary Table S24). Cold-related predictors, such as cold waves, exhibited a significant association with respiratory outcomes more often in persons <60 years than in those ≥60 years (pneumonia: 59.1% vs. 9.1%, COPD: 31.8% vs. 18.2%).

#### 3.3.2 Best-fitting regional outcome type based on weighted explained deviance (WED)

Model performance was evaluated and compared across outcome types and HBC-regions using the WED. Averaged across all outcome types and HBC-regions, the WED was 10%. When averaged across all regions, WED ranged from 2% for epilepsy to 34% for NSTEMI (Table 4). NSTEMI was the best-performing outcome type in every HBC-region except the north-east (HBC-region 6.1), where trauma performed marginally better (WED: 11% for trauma vs. 10% for NSTEMI; Supplementary Table S25). At the level of individual regions, the highest WED was observed for NSTEMI in HBC-region 6.2 (WED = 49%). At subgroup level for NSTEMI in HBC-region 6.2, a particularly good model fit was observed in the older subgroups (FE and ME: ED = 56%). An overview of the ED values by region, outcome type, and subgroup are provided in Supplementary Table S26.

**Table 4.** Weighted explained deviance (WED, %) per outcome type across all human-bioclimatic regions.

| Category | Outcome type | Weighted explained deviance (%) |
|---|---|---|
| Cardiovascular | Non-ST elevation myocardial infarction (NSTEMI) | 33.6 |
|  | ST-elevation myocardial infarction (STEMI) | 5.6 |
|  | Heart failure | 8.5 |
|  | Pulmonary artery embolism | 3.1 |
|  | Tachycardia | 12.4 |
| Neurological | Ischemic stroke | 5.0 |
|  | Ischemic stroke (stroke unit) | 11.1 |
|  | Epilepsy | 1.9 |
|  | Non subdural hemorrhage | 2.6 |
|  | Non subdural hemorrhage (stroke unit) | 2.4 |
|  | Transient ischemic attack | 2.9 |
|  | Transient ischemic attack (stroke unit) | 6.8 |
| Respiratory | Pneumonia | 16.6 |
|  | Chronic obstructive pulmonary disease | 10.9 |
| Internal | Sepsis | 9.9 |
| Traumatic | Trauma | 26.0 |

To illustrate the associations between atmospheric factors and daily ICU admissions in more detail, we focus on the four subgroup-specific NSTEMI models for eastern Germany (HBC-region 6.2). The results (including lags, effect sizes and p-values) for all other models are given in the Supplementary (Tables S8-S23). SLP was significantly associated with NSTEMI admissions (p ≤ 0.05) in all subgroup models except for female <60 years. A 10hPa lower SLP was associated with a 1.1% to 1.6% higher risk of ICU admissions (FE: RR 1.011, 95% CI 1.003 – 1.019; FY: RR 1.000, 95% CI 0.999 – 1.000; ME: RR 1.012, 95% CI 1.005 – 1.021; MY: RR 1.016, 95% CI 1.003 – 1.028; Supplementary Figure S9). Other pressure-related variables, such as wind days, showed no collinearity with SLP and no relevant association (Figure 3). From the heat-related variables only heat waves showed a statistically significant association in males ≥60 years (RR 0.941, 95% CI 0.895 – 0.989, also see Figure 3 and Supplementary Figures S10, S11), corresponding to a 6% lower risk of ICU admissions on heat wave days. In contrast, cold-related variables such as frost days were associated with higher risk for ICU admissions in older persons, estimates in younger persons were similar but nonsignificant and less precise (FE: RR 1.022, 95% CI 1.001 – 1.043; ME: 1.024, 95% CI 1.004 – 1.044; FY: RR 1.038, 95% CI 0.982 – 1.098; MY: RR 1.022, 95% CI 0.987 – 1.058; Figure 3, WCI: Supplementary Figure S12). No air pollutant was significantly associated with NSTEMI admissions (p ≤ 0.05), and the effect estimates were close to zero (Figure 3, Supplementary, Figure S13). Tests for heterogeneity between subgroups indicated differences only for SLP (Cochran’s Q = 14.2, adjusted p = 0.016; I² = 79%). For all other predictors, there was no evidence of differences between subgroups (all adjusted p ≥ 0.53). The heterogeneity for SLP was driven by a weaker association in women <60 years compared with men <60 years (ratio of RRs 0.985, 95% CI 0.972 – 0.998, p = 0.05) and women ≥60 years (ratio of RRs 0.989, 95%CI 0.980 – 0.998, p = 0.05). In summary, lower SLP and cold-related conditions were associated with higher risk for NSTEMI-related ICU admissions in eastern Germany, whereas heat and air pollution showed little or no association.

**Figure 3.**
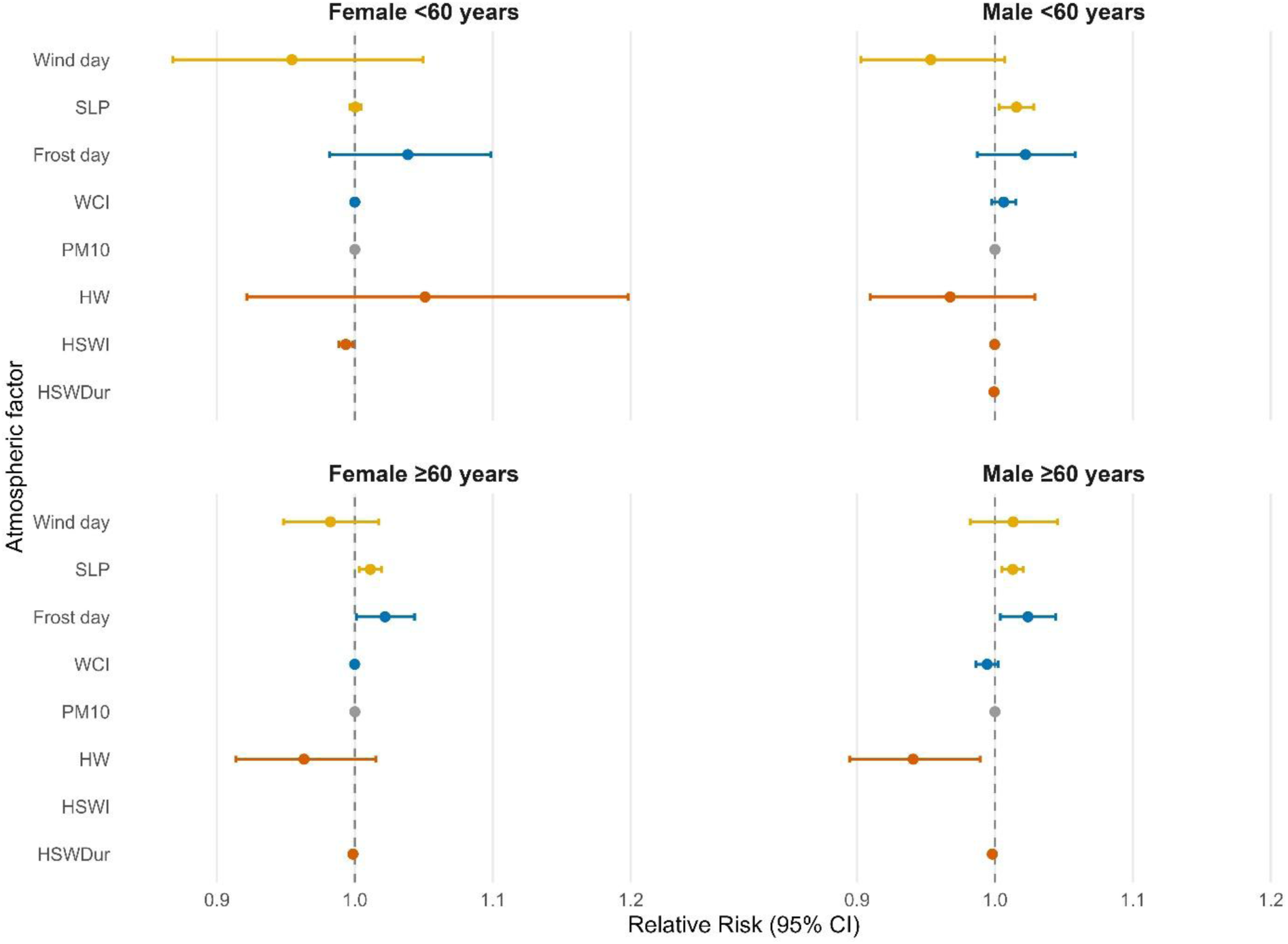
Forest plot of the association between myocardial infarction (NSTEMI) admissions and atmospheric predictors in human-bioclimatic region 6.2. Relative risks (RR) are shown with 95% confidence intervals (CI). RRs refer to the following changes in exposure: sea-level pressure (SLP), decrease of 10hPa (1015.9 vs. 1005.9hPa); wind chill index (WCI), decrease of 5°C (0 vs. -5°C); particulate matter (PM_10_), increase of 10µg/m³ (0 vs. 10µg/m³); heat stress wave intensity (HSWI), increase of 0.4°C (0 vs. 0.4°C); heat stress wave duration (HSWDur), increase of 0.3 days (0 vs. 0.3 days). For the binary predictors wind day, frost day and heat wave, RRs compare days with an event to days without an event. **Alt text Figure 3:** Four forest plots, one per age-sex subgroup, showing relative risks with 95% confidence intervals for atmospheric predictors and NSTEMI intensive care unit admissions in human-bioclimatic region 6.2. Lower sea-level pressure and frost days are associated with a significant higher risk in almost all subgroups.

## 4 Discussion

The present study indicates that atmospheric factors are associated with intensive care admissions, with pronounced differences by atmospheric predictor, outcome, region, age and sex. Pressure-related variables were most frequently significantly associated with ICU admissions, followed by heat-related predictors. Associations were dependent on outcome types and no atmospheric factor was associated with all outcome types. This suggests that diseases are influenced by different atmospheric exposures. The relevance of atmospheric pressure for an increased risk of cardiovascular and neurological diseases was also reported in earlier studies showing for example that drops in atmospheric pressure are related with stroke incidence [38–40].

Heat-related predictors, particularly tropical nights and summer days, were significantly associated with ICU admissions in a considerable proportion of models and were mainly linked to neurological outcomes. This is consistent with previous research. He et al. [41] reported that higher nighttime temperatures were associated with an increased stroke risk, and other studies have linked temperature to mortality and cardiopulmonary diseases [4, 42]. The relevance of both day- and nighttime heat for ICU admissions suggests that insufficient nocturnal cooling may contribute to thermal stress. In contrast, heat and cold (stress) wave related indices were less frequently associated with ICU admissions. Although earlier studies have reported associations between heat waves and various diseases, inconsistent definitions of spell-based indices and differences in the health outcomes studied make comparisons difficult [12, 43].

Thermo-physiological indices, which combine temperature, humidity and radiation, played an important role in many models. This supports the assumption that other thermal stress variables than temperature alone are relevant for ICU admissions in Germany. Also the German Weather Service uses perceived temperature to assess thermal stress and issues warnings to health authorities [44]. Beyond purely temperature-based predictors, thermo-physiological indices should therefore be considered in practical applications to describe thermal stress more accurately.

Among the air pollutants, ozone showed the highest proportion of significant associations with ICU admissions, whereas all other pollutants were significantly associated in fewer than 10 % of models. A possible explanation is the decline in concentrations of these pollutants over the 15-year study period, which reduced exposure and thus the ability to detect associations, whereas ozone exposure has steadily increased in Germany. Therefore, future studies and health adaptation tools should consider daily ozone levels to reduce health consequences and support the planning of health care capacities [45]. However, it also must be noted, that in the present study air pollution exposure was described by regional averages. This could have led to an exposure underestimation in certain areas such as larger urban areas and industrial hubs.

In the sex-stratified analyses, a larger proportion of models for males showed significant associations with atmospheric factors than for females. However, since models were fitted separately for each subgroup and comparisons are based on the proportion of significant results, these differences do not necessarily reflect true differences in susceptibility. Although slightly more ICU admissions occurred among men (56 %), which may have contributed to higher statistical power, this is unlikely to be the sole explanation. Heat-related factors were more frequently significantly associated with ICU admissions in men, whereas women showed more significant associations with air pollutants, particularly PM_10_ and ozone, which is in line with literature describing a higher vulnerability of women to air pollution [46]. The lower number of significant heat associations in women could be related to behavioral and occupational factors, such as higher health awareness, greater use of preventive care or lower occupational heat exposure [47, 48]. As these factors were not assessed in this study, this explanation remains speculative. Age-specific analyses showed more significant associations with atmospheric factors in people aged ≥60 years. Gronlund et al. [49] reported increased hospital admissions and emergency room visits among older people during extreme heat and heatwaves. The higher statistical power of the models for elderly may partly be explained by the high proportion of older adults admitted to ICUs. Nevertheless, heat-related factors were also significantly associated with ICU admissions in younger age groups, indicating that heat is relevant across all ages.

Overall, the best model performance based on WED was observed for myocardial infarction (NSTEMI) models. As NSTEMI was among the outcomes with the most frequent ICU admissions, the large number of admissions may have allowed higher statistical power. In eastern Germany (HBC-region 6.2), atmospheric pressure was significantly associated with NSTEMI ICU admissions in most subgroup models. A 10hPa drop in atmospheric pressure as well as cold-related events were associated with an increase in ICU admission risk. In this context, a sudden atmospheric pressure drop may act directly on the cardiovascular system or serve as a proxy for the associated changes of the weather conditions. For cold-related associations, increased blood viscosity and arterial pressure have been discussed as possible mechanisms, although neither was assessed in this study [50]. No clear association was found between heat and NSTEMI, which may be explained by less heat exposure in this region. The apparently protective effect of heat waves in older men was only marginally significant (upper confidence limit close to 1). It should therefore be interpreted with caution. Fine particulate matter has previously been linked to vascular damage and myocardial infarction [51]. The absence of such an association in these models may be due to the use of regional averages, which cannot capture small-scale variations in air pollutant concentrations and may therefore obscure localized exposure effects. Although the effects of the atmospheric factors were relatively small and their public health relevance at the population level is therefore limited, even small changes in ICU admissions can be detrimental to health care preparedness and delivery [52].

A strength of this study is the use of routine data, which reflect real-world healthcare provision and provide a large number of cases, enabling disease-specific and subgroup analyses [14]. Changes in ICD-10-GM coding were checked and incorporated into the case selection, and the ICU-relevant diseases were defined a priori. Furthermore, the consideration of different HBC-regions in Germany made it possible to capture regional differences in atmospheric-health associations across the whole country.

Several limitations should be considered. AOK data are claims data, and coding biases due to financial incentives may have affected data validity [14]. Also, the sample consisted exclusively of AOK insured individuals. The age and sex structure differs from that of the German population, which may limit external validity. However, AOK insured people account for almost one third of the German population [53]. In a follow-up study, which will assess the prediction suitability of the models, the results will be age-sex standardized to the German population. Thus, subgroup models were fitted separately in the present study, taking the loss that differences in the number of ICU admissions may have led to differences in statistical power. Regarding atmospheric predictors, the use of regional averages for exposure did not allow to capture small-scale variations, particularly relevant for air pollutants and the urban heat islands. Furthermore, Bonferroni correction was applied within each model but not across the large number of region-, outcome- and subgroup-specific models, so some significant associations may be chance findings. Collinear predictors (|ρ| ≥ 0.7) were handled by retaining the predictor with the lower p-value in the regional regression analysis. As a result, retained predictors may be favored, which should be considered when interpreting the proportion of significant models and comparing predictors. Also, as for any epidemiological time series study, the present study does not per se allow causal interpretation. Models were adjusted for seasonality, time trend and weekday, but not for other time-varying factors such as public holidays or influenza epidemics, which may be particularly relevant for respiratory outcomes. The study period of 15 years included the COVID-19 pandemic, as the impact of the pandemic on the intensive care was negligible and the data shown a long-term downward trend in admissions, suggesting a lasting change in health care structures [11]. Furthermore, given that intensive care constitutes critical infrastructure that is less affected by the postponement of elective procedures than general hospital admissions, the pandemic period was not analyzed separately [54].

In conclusion, this study indicates that atmospheric factors, most noticeably pressure-related ones, are associated with intensive care admissions in Germany. The study also extends previous research that focused mainly on mortality and general hospitalization. Associations differed by region, outcome, age and sex. Although the observed effects were small, they may be relevant for critical health care infrastructure such as intensive care and may inform regional ICU capacity planning. In a follow-up study, the identified associations will be used to assess short-term predictability of ICU admissions with the scope to develop a practically relevant tool.

## 5 Author Contributions

KS: Conceptualization, Data curation, formal analysis, methodology, software, visualization, writing – original draft, writing – review & editing; CM: Data curation, formal analysis, software, visualization, writing – review & editing; MJ: Writing – review & editing; TB: Data curation, writing – review & editing; PD: Data curation, writing – review & editing; CG: Data curation, writing – review & editing; TR: Data curation, writing – review & editing; PEB: Data curation, writing – review & editing; LP: Writing – review & editing; BW: Writing – review & editing; YI: Writing – review & editing; SH: Writing – review & editing; TÖ: Writing – review & editing; IS: Writing – review & editing; EH: Conceptualization, methodology, writing – review & editing

## 7 Acknowledgments

The present study is part of the ALERT-ITS project. ALERT-ITS is an acronym for “Development of a prediction and monitoring model for regional forecasting of intensive care and ventilation needs based on atmospheric factors”.

## 8 Ethical approval

The study protocol of ALERT-ITS was approved by the Ethics Committee of the Medical Faculty of Ludwig Maximilian University of Munich (LMU). The study was registered in the German Clinical Trials Register (DRKS) under the registration number DRKS00034441, with registration being completed on December 17, 2024. The analysis used anonymized secondary data, the need to obtain the informed consent was waived by the Ethics Committee of the Medical Faculty of Ludwig Maximilian University of Munich (LMU). The committee decided that informed consent was not required. A data protection concept was adopted in collaboration with the AOK Research Institute (WIdO).

## 9 Data Availability Statement

The atmospheric datasets supporting the conclusion of this article are available in a public, open access repository. The reanalysis data for the atmospheric conditions is available on the ECMWF website, https://doi.org/10.24381/cds.adbb2d47 and the air hygiene data on the CAMS website, https://doi.org/10.24381/d58bbf47 [17, 19, 55, 56]. The authors confirm that the health data utilized in this study cannot be made available in the manuscript, the supplemental files, or in a public repository due to German data protection laws (‘Bundesdatenschutzgesetz’, BDSG). Therefore, they are stored on a secure drive in the WIdO, to facilitate replication of the results. Generally, access to data of statutory health insurance funds for research purposes is possible only under the conditions defined in German Social Law (SGB V § 287). Requests for data access can be sent as a formal proposal specifying the recipient and purpose of the data transfer to the appropriate data protection agency. Access to the data used in this study can only be provided to external parties under the conditions of the cooperation contract of this research project and after written approval by the sickness fund. For assistance in obtaining access to the data, please contact. The data and figures regarding time lag selection (including relative risks and its 95% confidence intervals) are available upon request.

