## Supplementary for "Intensive care unit admissions in Germany associated with atmospheric factors: a regional time-series analysis"

### Supplementary Material

#### Table of content

**Supplementary Tables**


**Table S1.** Definition of the intensive care unit according to Operation and Procedure (OPS) codes…… 5

**Table S2.** Definition of intensive care unit according to specialist department code………… 5

**Table S3.** Definition of the intensive care unit (ICU) relevant diseases including International Statistical Classification Of Diseases And Related Health Problems, 10th revision, German Modification (ICD-10-GM) codes and the pick-up criterion………………. 6

**Table S4.** Definition of stroke unit cases according to Operation and Procedure (OPS) codes. 7

**Table S5.** Overview of meteorological and air pollution variables and the corresponding calculation…… 8

**Table S6.1.** Significant associations between daily intensive care unit admissions and atmospheric factors in Germany per cardiovascular outcome type. Significant (%) and absolute (n) values of all 44 possible models (4 subgroups x 11 human-bioclimatic regions)... 11

**Table S6.2**. Significant associations between daily intensive care unit admissions and atmospheric factors in Germany per neurological outcome type. Significant (%) and absolute (n) values of all 44 possible models (4 subgroups x 11 human-bioclimatic regions)………… 14

**Table S6.3.** Significant associations between daily intensive care unit admissions and atmospheric factors in Germany per respiratory, internal and traumatic outcome type. Significant (%) and absolute (n) values of all 44 possible models (4 subgroups x 11 human-bioclimatic regions)……. 17

**Table S7.** Percentual sex- and age-distributed significant associations between daily intensive care unit admissions and atmospheric factors in Germany, compromised 352 models in for female and male groups as well as for <60 years and ≥60 years groups……….. 20

**Table S8.** Overview of time lags, effect sizes (effective degree of freedom (EDF) or relative risk (95% confidence interval)) and p-values per human-bioclimatic region, atmospheric factor and subgroup (pink = female <60 years, orange = females ≥60 years, green = males <60 years, blue = males ≥60 years) for chronic obstructive pulmonary disease (COPD), significant p-values (≤ 0.05) are bold…….. 22

**Table S9.** Overview of time lags, effect sizes (effective degree of freedom (EDF) or relative risk (95% confidence interval)) and p-values per human-bioclimatic region, atmospheric factor and subgroup (pink = female <60 years, orange = females ≥60 years, green = males <60 years, blue = males ≥60 years) for epilepsy, significant p-values (≤ 0.05) are bold……… 27

**Table S10.** Overview of time lags, effect sizes (effective degree of freedom (EDF) or relative risk (95% confidence interval)) and p-values per human-bioclimatic region, atmospheric factor and subgroup (pink = female <60 years, orange = females ≥60 years, green = males <60 years, blue = males ≥60 years) for heart failure, significant p-values (≤ 0.05) are bold…………………………………………………………………………... 31

**Table S11.** Overview of time lags, effect sizes (effective degree of freedom (EDF) or relative risk (95% confidence interval)) and p-values per human-bioclimatic region, atmospheric factor and subgroup (pink = female <60 years, orange = females ≥60 years, green = males <60 years, blue = males ≥60 years) for ischemic stroke, significant p-values (≤ 0.05) are bold…………………………………………………………………………... 34

**Table S12.** Overview of time lags, effect sizes (effective degree of freedom (EDF) or relative risk (95% confidence interval)) and p-values per human-bioclimatic region, atmospheric factor and subgroup (pink = female <60 years, orange = females ≥60 years, green = males <60 years, blue = males ≥60 years) for ischemic stroke (stroke unit), significant p-values (≤ 0.05) are bold………………………………………………………….. 39

**Table S13.** Overview of time lags, effect sizes (effective degree of freedom (EDF) or relative risk (95% confidence interval)) and p-values per human-bioclimatic region, atmospheric factor and subgroup (pink = female <60 years, orange = females ≥60 years, green = males <60 years, blue = males ≥60 years) for pulmonary artery embolism, significant p-values (≤ 0.05) are bold………………………………………………………….. 43

**Table S14.** Overview of time lags, effect sizes (effective degree of freedom (EDF) or relative risk (95% confidence interval)) and p-values per human-bioclimatic region, atmospheric factor and subgroup (pink = female <60 years, orange = females ≥60 years, green = males <60 years, blue = males ≥60 years) for myocardial infarction (NSTEMI), significant p-values (≤ 0.05) are bold………………………………………………………….. 48

**Table S15.** Overview of time lags, effect sizes (effective degree of freedom (EDF) or relative risk (95% confidence interval)) and p-values per human-bioclimatic region, atmospheric factor and subgroup (pink = female <60 years, orange = females ≥60 years, green = males <60 years, blue = males ≥60 years) for non subdural hemorrhage, significant p-values (≤ 0.05) are bold………………………………………………………………… 51

**Table S16.** Overview of time lags, effect sizes (effective degree of freedom (EDF) or relative risk (95% confidence interval)) and p-values per human-bioclimatic region, atmospheric factor and subgroup (pink = female <60 years, orange = females ≥60 years, green = males <60 years, blue = males ≥60 years) for non subdural hemorrhage (stroke unit), significant p-values (≤ 0.05) are bold……………………………………………………….. 56

**Table S17.** Overview of time lags, effect sizes (effective degree of freedom (EDF) or relative risk (95% confidence interval)) and p-values per human-bioclimatic region, atmospheric factor and subgroup (pink = female <60 years, orange = females ≥60 years, green = males <60 years, blue = males ≥60 years) for pneumonia, significant p-values (≤ 0.05) are bold……………………………………………………………………………... 59

**Table S18.** Overview of time lags, effect sizes (effective degree of freedom (EDF) or relative risk (95% confidence interval)) and p-values per human-bioclimatic region, atmospheric factor and subgroup (pink = female <60 years, orange = females ≥60 years, green = males <60 years, blue = males ≥60 years) for myocardial infarction (STEMI), significant p-values (≤ 0.05) are bold……………………………………………………………………………………….. 64

**Table S19.** Overview of time lags, effect sizes (effective degree of freedom (EDF) or relative risk (95% confidence interval)) and p-values per human-bioclimatic region, atmospheric factor and subgroup (pink = female <60 years, orange = females ≥60 years, green = males <60 years, blue = males ≥60 years) for sepsis, significant p-values (≤ 0.05) are bold……………………………………………………………………………………….. 67

**Table S20.** Overview of time lags, effect sizes (effective degree of freedom (EDF) or relative risk (95% confidence interval)) and p-values per human-bioclimatic region, atmospheric factor and subgroup (pink = female <60 years, orange = females ≥60 years, green = males <60 years, blue = males ≥60 years) for transient ischemic attack, significant p-values (≤ 0.05) are bold…………………………………………………………………... 70

**Table S21.** Overview of time lags, effect sizes (effective degree of freedom (EDF) or relative risk (95% confidence interval)) and p-values per human-bioclimatic region, atmospheric factor and subgroup (pink = female <60 years, orange = females ≥60 years, green = males <60 years, blue = males ≥60 years) for transient ischemic attack (stroke unit), significant p-values (≤ 0.05) are bold……………………………………………………….. 74

**Table S22.** Overview of time lags, effect sizes (effective degree of freedom (EDF) or relative risk (95% confidence interval)) and p-values per human-bioclimatic region, atmospheric factor and subgroup (pink = female <60 years, orange = females ≥60 years, green = males <60 years, blue = males ≥60 years) for tachycardia, significant p-values (≤ 0.05) are bold……………………………………………………………………………... 78

**Table S23.** Overview of time lags, effect sizes (effective degree of freedom (EDF) or relative risk (95% confidence interval)) and p-values per human-bioclimatic region, atmospheric factor and subgroup (pink = female <60 years, orange = females ≥60 years, green = males <60 years, blue = males ≥60 years) for trauma, significant p-values (≤ 0.05) are bold………………………………………………………………………………… 81

**Table S24.1.** Sex- and age-specific significant associations between daily intensive care unit admissions and atmospheric factors in Germany per cardiovascular outcome type. Significant percentage (%) of all 22 possible models (2 sex/age subgroups x 11 human-bioclimatic regions), order of the presented values: Females, males, <60 years, ≥60 years…... 88

**Table S24.2.** Sex- and age-specific significant associations between daily intensive care unit admissions and atmospheric factors in Germany per neurological outcome type. Significant percentage (%) of all 22 possible models (2 sex/age subgroups x 11 human-bioclimatic regions), order of the presented values: Females, males, <60 years, ≥60 years……………….. 91

**Table S24.3.** Sex- and age-specific significant associations between daily intensive care unit admissions and atmospheric factors in Germany per respiratory, internal and traumatic outcome type. Significant percentage (%) of all 22 possible models (2 sex/age subgroups x 11 human-bioclimatic regions), order of the presented values: Females, males, <60 years, ≥60 years………………………………………………………………………………… 94

**Table S25.1.** Weighted explained deviance (WED, %) across sex-, age-specific subgroups per human-bioclimatic (HBC) region and cardiovascular outcome type……………………… 97

**Table S25.2.** Weighted explained deviance (WED, %) across subgroups per human-bioclimatic (HBC) region and neurological outcome type…………………………………….. 98

**Table S25.3.** Weighted explained deviance (WED, %) across subgroups per human-bioclimatic (HBC) region and respiratory, internal and traumatic outcome type……………... 99

**Table S26.1.** Explained deviance (ED, %) across sex-, age-specific subgroups per human-bioclimatic (HBC) region and cardiovascular outcome type, order of the presented values: Female <60 years, Female ≥60 years, Male <60 years, Male ≥60 years……………………… 100

**Table S26.2.** Explained deviance (ED, %) across subgroups per human-bioclimatic (HBC) region and neurological outcome type, order of the presented values: Female <60 years, Female ≥60 years, Male <60 years, Male ≥60 years…………………………………………... 101

**Table S26.3.** Explained deviance (ED, %) across subgroups per human-bioclimatic (HBC) region and respiratory, internal and traumatic outcome type, order of the presented values: Female <60 years, Female ≥60 years, Male <60 years, Male ≥60 years……………………… 102

**Supplementary Figures**

**Figure S1.** Annual intensive care unit (ICU) admissions per 100,000 AOK insured persons by outcome type and human-bioclimatic (HBC) region, 2009–2023, standardized for age and sex to the pooled AOK-insured population. Each panel shows one outcome type, lines represent HBC-regions…………………………………………………………………… 103

**Figure S2.** Annually air pollution factors per human-bioclimatic (HBC) region……………... 104

**Figure S3.** Annually heat-related factors (raw, binary or indices) per human-bioclimatic (HBC) region………………………………………………………………………………... 105

**Figure S4.** Annually heat wave-related factors per human-bioclimatic (HBC) region……….. 105

**Figure S5**. Annually heat stress wave-related factors per human-bioclimatic (HBC) region… 106

**Figure S6.** Annually cold-related factors (binary or indices) per human-bioclimatic (HBC) region…………………………………………………………………………………... 106

**Figure S7.** Annually cold wave-related factors per human-bioclimatic (HBC) region……….. 107

**Figure S8**. Annually cold stress wave-related factors per human-bioclimatic (HBC) region… 108

**Figure S9.** Subgroup-specific exposure-response curve between sea-level pressure (SLP) and myocardial infarction (NSTEMI) intensive care unit admissions in human-bioclimatic (HBC) region 6.2, dashed line = point of comparison (relative risk calculation)……………... 109

**Figure S10.** Subgroup-specific exposure-response curve between heat stress wave intensity (°C) and myocardial infarction (NSTEMI) intensive care unit admissions in human-bioclimatic (HBC) region 6.2, dashed line = point of comparison (relative risk calculation)… 110

**Figure S11**. Subgroup-specific exposure-response curve between heat stress wave duration (days) and myocardial infarction (NSTEMI) intensive care unit admissions in human-bioclimatic (HBC) region 6.2, dashed line = point of comparison (relative risk calculation)… 111

**Figure S12.** Subgroup-specific exposure-response curve between Wind chill index (WCI) and myocardial infarction (NSTEMI) intensive care unit admissions in human-bioclimatic (HBC) region 6.2, dashed line = point of comparison (relative risk calculation)……………... 112

**Figure S13.** Subgroup-specific exposure-response curve between Particulate matter ($PM_{10}$) and myocardial infarction (NSTEMI) intensive care unit admissions in human-bioclimatic (HBC) region 6.2, dashed line = point of comparison (relative risk calculation)……………... 113

#### Supplementary Tables

**Table S1.** Definition of the intensive care unit according to Operation and Procedure (OPS) codes

| Category | OPS-Code | Description | Remark |
|---|---|---|---|
| Intensive care complex treatment | 8-98f | Complex intensive care treatment (basic procedure) | Relevant since 2013 |
| Intensive care complex treatment | 8-980 | Intensive care treatment (basic procedure) | |
| Patient monitoring | 8-920 | EEG monitoring (at least 2 channels) for more than 24 h | |
| Patient monitoring | 8-921 | Monitoring by means of evoked potentials | |
| Patient monitoring | 8-923 | Monitoring of cerebral venous oxygen saturation | Since 2019 differentiation between invasive and non-invasive |
| Patient monitoring | 8-924 | Invasive neurological monitoring | |
| Patient monitoring | 8-930 | Monitoring of respiration, heart and circulation without measurement of pulmonary artery pressure and central venous pressure | |
| Patient monitoring | 8-931 | Monitoring of respiration, heart and circulation with measurement of central venous pressure | |
| Patient monitoring | 8-932 | Monitoring of respiration, heart and circulation with measurement of pulmonary artery pressure | |

**Table S2.** Definition of intensive care unit according to specialist department code

| Specialist department code | Description | Remark |
|---|---|---|
| 36XX | Intensive care medicine | All department codes in the range 3600 to 3699 are included |
| 0436 | Nephrology/Intensive care medicine | |

| | |
|---|---|
| 1536 | General surgery/ Intensive care medicine |
| 2036 | Thoracic surgery/ Intensive care medicine |
| 2050 | Thoracic surgery/ Focus on cardiac surgery Intensive care medicine |
| 2136 | Cardiac surgery/ Intensive care medicine |
| 2150 | Cardiac surgery/ Focus on thoracic surgery Intensive care medicine |

**Table S3.** Definition of the intensive care unit (ICU) relevant diseases including International Statistical Classification Of Diseases And Related Health Problems, 10th revision, German Modification (ICD-10-GM) codes and the pick-up criterion

| Category | Disease | ICD-10-GM code | Pick-up criterion |
|---|---|---|---|
| **Cardiovascular** | Heart failure | I11.0, I13.0, I13.2, I50 | ICU |
| | Coronary heart diseases (ST-segment elevation myocardial infarction - STEMI) | I21.0, I21.1, I21.2, I21.3 | Independent of ICU |
| | Coronary heart disease (Non ST-segment elevation myocardial infarction – NSTEMI) | 1: I21.4, I21.9, I20.0 | 1: Independent of ICU |
| | | 2: I20.1, I20.8, I20.9, I25.6, I25.8, I25.9 | 2: ICU |
| | | 3: I25.11, I25.12, I25.13, I25.14, I25.15, I25.16 | 3: ICU exclusive OPS code of 5-361 |
| | Tachycardia | I47.0, I47.1, I47.2, I47.9, I48.00, I48.01, I48.10, I48.11, I47.0, I47.1, I47.2, I47.9, I48.0, I48.2, I48.3, I48.4 | ICU |

| | | | |
|---|---|---|---|
| | Pulmonary artery embolism | I26 | 1: ICU independent of OPS code 8-020.8<br><br>2: Independent of ICU with the condition of OPS code 8-020.8 |
| **Respiratory** | Pneumonia | J10, J11, J12, J13, J14, J15, J16, J17, J18, J20, J21, J22 | 1: ICU<br><br>2: Independent of ICU but with ventilation (OPS code 8-71 and/or 8-70) |
| | Chronic obstructive pulmonary disease (COPD) | J44 | 1: ICU<br><br>2: Independent of ICU but with ventilation (OPS code 8-71 and/or 8-70) |
| **Neurological** | Ischemic insult | I63 | Independent of ICU or<br><br>Stroke Unit OPS code |
| | Transient ischemic attack | G45 | Independent of ICU or<br><br>Stroke Unit OPS code |
| | Non-Subdural hemorrhage | I60, I61 | Independent of ICU or<br><br>Stroke Unit OPS code |
| | Epilepsy | G40, G41 | ICU |
| **General internal** | Sepsis | A40, A41, R65* | ICU |
| **Traumatic** | Trauma | S*, T00 – T14 | ICU |

**Table S4.** Definition of stroke unit cases according to Operation and Procedure (OPS) codes

| **OPS code** | **Description** |
|---|---|
| 8-981 | Neurological complex treatment |
| 8-981.2 | Stroke unit without thrombectomy/intracranial procedures |
| 8-981.3 | Stroke unit with thrombectomy/intracranial procedures |

| | |
|---|---|
| 8-98b | Other complex neurological treatment of acute stroke |
| 8-98b.2 | Without telemedicine |
| 8-98b.3 | With telemedicine |

**Table S5.** Overview of meteorological and air pollution variables and the corresponding calculation

| Variable | Long name | Unit | Calculation |
|---|---|---|---|
| T | Mean temperature | °C | $T_D = \frac{\sum_{h=1}^{24} T_{D_h}}{24}$ |
| SLP | Mean sea-level pressure | hPa | $SLP_D = \frac{\sum_{h=1}^{24} SLP_{D_h}}{24}$ |
| SLPD | Substantial change in sea-level pressure compared to the previous day | -1\|0\|1 | $SLPD_D = \begin{cases} \|SLP_D - SLP_{D-1}\| > +10 = +1 \\ -10 \leq \|SLP_D - SLP_{D-1}\| \leq +10 = 0 \\ \|SLP_D - SLP_{D-1}\| < -10 = -1 \end{cases}$ |
| WD | Wind day | 0\|1 | $WS_D = \frac{\sum_{h=1}^{24} WS_{D_h}}{24}$ <br> $WD_D = \begin{cases} WS_D \leq WS_{Q95} = 0 \\ WS_D > WS_{Q95} = 1 \end{cases}$ |
| WBGT | Wet-bulb globe temperature | °C | $according\ to\ Lilijegren:$ <br> $WBGT_{D_h} = 0.7T_{W_h} + 0.2T_{G_h} + 0.1T_{A_h}$ <br> $WBGT_D = max\{WBGT_{D_h}\}$ |
| WCI | Windchill index | °C | $WCI_{D_h} = 13.12 + 0.6215T_{D_h} - 11.37WS_{D_h}^{0.16} + 0.3965T_{D_h}WS_{D_h}^{0.16}$ <br> $WCI_D = min\{WCI_{D_h}\}$ |
| $PM_{2.5}$ | Particular matter < 2.5µm | µg/m$^3$ | $PM_{2.5_D} = \frac{\sum_{h=1}^{24} PM_{2.5_{D_h}}}{24}$ |
| $PM_{10}$ | Particular matter < 10µm | µg/m$^3$ | $PM_{10_D} = \frac{\sum_{h=1}^{24} PM_{10_{D_h}}}{24}$ |

| | | | |
|---|---|---|---|
| $NO_2$ | Nitrogen dioxide | ppbv | $NO_{2_D} = \frac{\sum_{h=1}^{24} NO_{2_{D_h}}}{24}$ |
| $O_3$ | Ozone | pbbv | $O_{3_D} = max\left\{\frac{\sum_{h=-1}^{-7} O_{3_{D_h}}}{8}\right\}$ |
| ID | Ice day | 0\|1 | $if\ max\{T_{D_h}\} < 0°C, 1\ else\ 0$ |
| FD | Frost day | 0\|1 | $if\ min\{T_{D_h}\} < 0°C, 1\ else\ 0$ |
| SD | Summer day | 0\|1 | $if\ max\{T_{D_h}\} > 25°C, 1\ else\ 0$ |
| HD | Hot Day | 0\|1 | $if\ max\{T_{D_h}\} > 30°C, 1\ else\ 0$ |
| TN | Tropical night | 0\|1 | $if\ min\{T_{D_h}\} > 20°C, 1\ else\ 0$ |
| HWI | Heatwave intensity | °C | $HWI_D = \begin{cases} if\ T_D > T_{Q95}, \sum_{i=1}^{n}(T_D - T_{Q95}) \\ if\ T_D \leq T_{Q95}, \sum_{i=1}^{n}(T_D - T_{Q95}) \cdot N_D \end{cases}$ <br> $if\ HWI_D < 0 \rightarrow HWI_D = 0$ |
| HWDay | Heatwave day | 0\|1 | $if\ HWI > \ 0, 1\ else\ 0$ |
| HWDur | Heatwave duration | N | $Number\ of\ consecutive\ days\ with\ HWI > 0$ |
| HW | Heatwave | 0\|1 | $if\ HWDur > 2, 1\ else\ 0$ |
| HSWI | Heatstresswave intensity | °C | $see\ HWI, but\ for\ WBGT\ instead\ of\ T$ |
| HSWDay | Heatstresswave day | 0\|1 | $if\ HSWI > 0, 1\ else\ 0$ |
| HSWDur | Heatstresswave duration | N | $Number\ of\ consecutive\ days\ with\ HSWI > 0$ |
| HSW | Heatstresswave | 0\|1 | $if\ HSWDur > 2,1\ else\ 0$ |
| CWI | Coldwave intensity | °C | $CWI_D = \begin{cases} if\ T_D < T_{Q5}, \sum_{i=1}^{n}(T_D - T_{Q5}) \\ if\ T_D \geq T_{Q5}, \sum_{i=1}^{n}(T_D - T_{Q5}) \cdot N_D \end{cases}$ |

| | | | |
|---|---|---|---|
| | | | $if\ CWI_D > 0 \rightarrow CWI_D = 0$ |
| CWDay | Coldwave day | 0\|1 | $if\ CWI < 0, 1\ else\ 0$ |
| CWDur | Coldwave duration | N | $Number\ of\ consecutive\ days\ with\ CWI < 0$ |
| CW | Coldwave | 0\|1 | $if\ CWDur > 2{,}1\ else\ 0$ |
| CSWI | Coldstresswave intensity | °C | $see\ CWI, but\ for\ WCI\ instead\ of\ T$ |
| CSWDay | Coldstresswave day | 0\|1 | $if\ CSWI < 0, 1\ else\ 0$ |
| CSWDur | Coldstresswave duration | N | $Number\ of\ consecutive\ days\ with\ CSWI < 0$ |
| CSW | Coldstresswave | 0\|1 | $if\ CSWDur > 2{,}1\ else\ 0$ |

**Table S6.1.** Significant associations between daily intensive care unit admissions and atmospheric factors in Germany per cardiovascular outcome type. Significant (%) and absolute (n) values of all 44 possible models (4 subgroups x 11 human-bioclimatic regions)

| Atmospheric factor | Myocardial infarction (STEMI) | Myocardial infarction (NSTEMI) | Heart failure | Pulmonary artery embolism | Tachycardia |
|---|---|---|---|---|---|
| Sea-level pressure (SLP) | 45.5% (20) | 59.1% (26) | 43.2% (19) | 36.4% (16) | 40.9% (18) |
| Sea-level pressure difference (SLPD) | - | - | - | 34.1% (15) | - |
| Wind day | - | 36.4% (16) | - | 45.5% (20) | 50.0% (22) |
| Tropical night | 31.8% (14) | - | 22.7% (10) | 36.4% (16) | 20.5% (9) |
| Hot day | 29.5% (13) | - | - | - | 29.5% (13) |
| Summer day | 45.5% (20) | - | 45.5% (20) | 20.5% (9) | - |
| Wet-bulb globe temperature | - | - | 11.4% (5) | - | - |
| Daily mean temperature | - | - | 15.9% (7) | - | - |
| Ice day | - | - | - | 31.8% (14) | - |
| Frost day | - | - | - | - | - |
| Wind chill index | - | 34.1% (15) | - | 18.2% (8) | 22.7% (10) |
| Heat wave | - | 34.1% (15) | - | - | - |

| | | | | | |
|---|---|---|---|---|---|
| Heat wave day | - | - | - | - | 31.8% (14) |
| Heat wave duration | - | - | 38.6% (17) | 22.7% (10) | - |
| Heat wave intensity | - | - | - | 13.6% (6) | 18.2% (8) |
| Heat stress wave | - | - | - | - | - |
| Heat stress wave day | - | - | - | 27.3% (12) | - |
| Heat stress wave duration | - | 11.4% (5) | - | - | - |
| Heat stress wave intensity | - | 20.5% (9) | - | 13.6% (6) | - |
| Cold wave | - | - | - | - | - |
| Cold wave day | - | - | - | - | - |
| Cold wave duration | 15.9% (7) | - | - | - | 20.5% (9) |
| Cold wave intensity | - | - | - | - | - |
| Cold stress wave | - | - | - | - | - |
| Cold stress wave day | - | - | 13.6% (6) | - | - |
| Cold stress wave duration | - | - | - | - | - |

| | | | | | |
|---|---|---|---|---|---|
| Cold stress wave intensity | - | - | 9.1 \| 18.2 \| 9.1 \| 18.2 | - | - |
| Ozone | 40.9 \| 27.3 \| 36.4 \| 31.8 | - | - | 54.5 \| 54.5 \| 50.0 \| 59.1 | 27.3 \| 13.6 \| 27.3 \| 13.6 |
| Nitrogen dioxide ($NO_2$) | - | - | - | - | - |
| Particular matter ($PM_{10}$) | - | 31.8 \| 27.3 \| 31.8 \| 27.3 | 13.6 \| 22.7 \| 27.3 \| 9.1 | 27.3 \| 40.9 \| 27.3 \| 40.9 | - |
| Particular matter ($PM_{2.5}$) | - | - | - | - | - |

**Table S6.2.** Significant associations between daily intensive care unit admissions and atmospheric factors in Germany per neurological outcome type. Significant (%) and absolute (n) values of all 44 possible models (4 subgroups x 11 human-bioclimatic regions)

| Atmospheric factor | Ischemic stroke | Ischemic stroke (stroke unit) | Non subdural hemorrhage | Non subdural hemorrhage (stroke unit) | Transient ischemic attack | Transient ischemic attack (stroke unit) | Epilepsy |
|---|---|---|---|---|---|---|---|
| Sea-level pressure (SLP) | 47.7% (21) | - | 34.1% (15) | - | - | 25.0% (11) | - |
| Sea-level pressure difference (SLPD) | - | 34.1% (15) | 31.8% (14) | - | 54.5% (24) | - | 38.6% (17) |
| Wind day | 50.0% (22) | - | 40.9% (18) | 52.3% (23) | - | 36.4% (16) | 40.9% (18) |
| Tropical night | 36.4% (16) | 27.3% (12) | 31.8% (14) | 20.5% (9) | 22.7% (10) | - | - |
| Hot day | 27.3% (12) | - | 18.2% (8) | 27.3% (12) | - | - | - |
| Summer day | 27.3% (12) | - | 25.0% (11) | - | 36.4% (16) | 52.3% (23) | - |
| Wet-bulb globe temperature | - | 25.0% (11) | 6.8% (3) | 40.9% (18) | 13.6% (6) | 9.1% (4) | 9.1% (4) |
| Daily mean temperature | 22.7% (10) | - | 13.6% (6) | - | - | - | - |
| Ice day | - | 11.4% (5) | - | - | - | - | - |
| Frost day | - | - | - | - | - | - | - |

| | | | | | | | |
|---|---|---|---|---|---|---|---|
| Wind chill index | - | - | 20.5% (9) | 34.1% (15) | - | 15.9% (7) | - |
| Heat wave | - | - | 13.6% (6) | - | 20.5% (9) | - | - |
| Heat wave day | - | - | - | - | - | - | 34.1% (15) |
| Heat wave duration | - | - | 9.1% (4) | - | - | - | - |
| Heat wave intensity | - | - | - | 15.9% (7) | - | - | - |
| Heat stress wave | - | - | - | - | - | 22.7% (10) | - |
| Heat stress wave day | - | 34.1% (15) | - | - | - | - | - |
| Heat stress wave duration | - | - | - | - | - | - | - |
| Heat stress wave intensity | 15.9% (7) | 15.9% (7) | - | - | - | - | 9.1% (4) |
| Cold wave | - | - | - | - | 9.1% (4) | - | - |
| Cold wave day | - | - | - | - | - | - | - |
| Cold wave duration | - | 6.8% (3) | - | - | - | - | 9.1% (4) |
| Cold wave intensity | 15.9% (7) | - | 6.8% (3) | - | - | - | - |

| | | | | | | | |
|---|---|---|---|---|---|---|---|
| Cold stress wave | - | 9.1% (4) | - | - | 6.8% (3) | - | 20.5% (9) |
| Cold stress wave day | - | - | 27.3% (12) | - | 6.8% (3) | - | - |
| Cold stress wave duration | 9.1% (4) | 6.8% (3) | - | - | - | 13.6% (6) | - |
| Cold stress wave intensity | 4.5% (2) | 4.5% (2) | 15.9% (7) | - | - | - | 9.1% (4) |
| Ozone | 20.5% (9) | - | 13.6% (6) | 27.3% (12) | 6.8% (3) | - | 18.2% (8) |
| Nitrogen dioxide ($NO_2$) | - | - | - | - | - | 13.6% (6) | - |
| Particular matter ($PM_{10}$) | 20.5% (9) | - | - | - | - | 18.2% (8) | - |
| Particular matter ($PM_{2.5}$) | - | - | 36.4% (16) | - | - | - | 27.3% (12) |

**Table S6.3.** Significant associations between daily intensive care unit admissions and atmospheric factors in Germany per respiratory, internal and traumatic outcome type. Significant (%) and absolute (n) values of all 44 possible models (4 subgroups x 11 human-bioclimatic regions)

| Atmospheric factor | Pneumonia | Chronic obstructive pulmonary disease (COPD) | Sepsis | Trauma |
|---|---|---|---|---|
| Sea-level pressure (SLP) | 36.4% (16) | 29.5% (13) | 29.5% (13) | 40.9% (18) |
| Sea-level pressure difference (SLPD) | 36.4% (16) | 47.7% (21) | 45.5% (20) | - |
| Wind day | 15.9% (7) | - | 29.5% (13) | - |
| Tropical night | 45.5% (20) | - | - | 20.5% (9) |
| Hot day | - | - | - | 29.5% (13) |
| Summer day | - | - | 31.8% (14) | - |
| Wet-bulb globe temperature | - | 20.5% (9) | 15.9% (7) | 25.0% (11) |
| Daily mean temperature | 11.4% (5) | 11.4% (5) | 9.1% (4) | 4.5% (2) |
| Ice day | - | 22.7% (10) | - | 29.5% (13) |
| Frost day | - | - | - | - |
| Wind chill index | 9.1% (4) | - | 6.8% (3) | 4.5% (2) |
| Heat wave | - | - | - | 13.6% (6) |

| | | | | |
|---|---|---|---|---|
| Heat wave day | - | - | 36.4% (16) | 13.6% (6) |
| Heat wave duration | - | - | - | - |
| Heat wave intensity | - | 15.9% (7) | - | 18.2% (8) |
| Heat stress wave | - | 34.1% (15) | - | - |
| Heat stress wave day | - | - | - | 6.8% (3) |
| Heat stress wave duration | - | - | - | - |
| Heat stress wave intensity | 18.2% (8) | - | - | - |
| Cold wave | 34.1% (15) | 25.0% (11) | - | - |
| Cold wave day | - | - | - | - |
| Cold wave duration | 20.5% (9) | 13.6% (6) | - | 36.4% (16) |
| Cold wave intensity | 6.8% (3) | 11.4% (5) | - | - |
| Cold stress wave | - | 20.5% (9) | - | - |
| Cold stress wave day | - | - | - | - |
| Cold stress wave duration | 18.2% (8) | - | - | - |
| Cold stress wave intensity | 18.2% (8) | 9.1% (4) | 11.4% (5) | 11.4% (5) |
| Ozone | 29.5% (13) | 9.1% (4) | - | 15.9% (7) |

| | | | | |
|---|---|---|---|---|
| Nitrogen dioxide ($NO_2$) | 22.7% (10) | - | - | 25.0% (11) |
| Particular matter ($PM_{10}$) | - | 13.6% (6) | - | 11.4% (5) |
| Particular matter ($PM_{2.5}$) | - | - | - | 20.5% (9) |

**Table S7.** Percentual sex- and age-distributed significant associations between daily intensive care unit admissions and atmospheric factors in Germany, compromised 352 models in for female and male groups as well as for <60 years and ≥60 years groups

| **Atmospheric factor** | **Female (%)** | **Male (%)** | **<60 years (%)** | **≥60 years (%)** |
|---|---|---|---|---|
| Sea-level pressure (SLP) | 27.3 | 31.2 | 26.7 | 31.8 |
| Sea-level pressure difference (SLPD) | 22.7 | 17.6 | 19.6 | 20.7 |
| Wind day | 24.7 | 25.0 | 29.0 | 20.7 |
| Tropical night | 19.9 | 17.0 | 19.3 | 17.6 |
| Hot day | 9.9 | 10.2 | 9.1 | 11.1 |
| Summer day | 17.3 | 18.2 | 14.8 | 20.7 |
| Wet-bulb globe temperature | 9.1 | 13.1 | 12.5 | 9.7 |
| Daily mean temperature | 6.8 | 4.3 | 6.2 | 4.8 |
| Ice day | 5.7 | 6.2 | 6.0 | 6.0 |
| Frost day | 15.6 | 13.9 | 14.8 | 14.8 |
| Wind chill index | 9.9 | 10.8 | 9.9 | 10.8 |
| Heat wave | 4.5 | 5.7 | 3.1 | 7.1 |
| Heat wave day | 6.0 | 8.5 | 3.7 | 10.8 |
| Heat wave duration | 3.4 | 5.4 | 2.8 | 6.0 |
| Heat wave intensity | 4.0 | 6.2 | 4.5 | 5.7 |
| Heat stress wave | 3.4 | 3.7 | 2.6 | 4.5 |
| Heat stress wave day | 4.3 | 4.3 | 5.1 | 3.4 |
| Heat stress wave duration | 0.9 | 0.6 | 0.6 | 0.9 |
| Heat stress wave intensity | 6.5 | 5.1 | 5.1 | 6.5 |
| Cold wave | 4.0 | 4.5 | 6.2 | 2.3 |
| Cold wave duration | 7.4 | 8.0 | 8.0 | 7.4 |
| Cold wave intensity | 2.8 | 2.3 | 2.8 | 2.3 |
| Cold stress wave | 4.3 | 2.8 | 2.8 | 4.3 |
| Cold stress wave day | 3.7 | 2.3 | 4.0 | 2.0 |
| Cold stress wave duration | 2.8 | 3.1 | 2.8 | 3.1 |

| | | | | |
|---|---|---|---|---|
| Cold stress wave intensity | 6.0 | 6.2 | 7.1 | 5.1 |
| Ozone | 17.0 | 14.2 | 15.6 | 15.6 |
| Nitrogen dioxide ($NO_2$) | 3.1 | 4.5 | 4.8 | 2.8 |
| Particular matter ($PM_{10}$) | 10.2 | 8.0 | 11.1 | 7.1 |
| Particular matter ($PM_{2.5}$) | 5.1 | 5.4 | 5.7 | 4.8 |

**Table S8.** Overview of time lags, effect sizes (effective degree of freedom (EDF) or relative risk (95% confidence interval)) and p-values per human-bioclimatic region, atmospheric factor and subgroup (pink = female <60 years, orange = females ≥60 years, green = males <60 years, blue = males ≥60 years) for chronic obstructive pulmonary disease (COPD), significant p-values (≤ 0.05) are bold

| **Atmospheric factor** | **1.1** | **1.2** | **2.1** | **2.2** | **3.1** | **4.1** | **5.1** | **5.2** | **6.1** | **6.2** | **6.3** |
|---|---|---|---|---|---|---|---|---|---|---|---|
| Sea-level pressure (SLP) | lag 9: EDF=0.51, p=0.171<br>**lag 9: EDF=0.79, p=0.024**<br>lag 8: EDF=0.51, p=0.158<br>**lag 1: EDF=0.84, p=0.010** | lag 10: EDF=0.72, p=0.052<br>**lag 0: EDF=0.87, p=0.005**<br>lag 3: EDF=0.37, p=0.193<br>lag 4: EDF=0.05, p=0.293 | lag 7: EDF=0.00, p=0.874<br>lag 0: EDF=0.00, p=0.832<br>lag 6: EDF=0.50, p=0.168<br>lag 5: EDF=0.00, p=0.528 | lag 0: EDF=0.00, p=0.682<br>lag 6: EDF=0.00, p=0.799<br>lag 10: EDF=0.70, p=0.066<br>lag 1: EDF=0.00, p=0.389 | lag 2: EDF=0.39, p=0.353<br>lag 3: EDF=0.00, p=0.900<br>lag 10: EDF=0.00, p=0.637<br>lag 6: EDF=0.09, p=0.274 | lag 3: EDF=0.01, p=0.430<br>**lag 2: EDF=0.90, p=0.003**<br>**lag 10: EDF=0.89, p=0.018**<br>**lag 10: EDF=0.91, p=0.002** | **lag 9: EDF=0.85, p=0.024**<br>lag 2: EDF=0.59, p=0.105<br>lag 6: EDF=0.00, p=0.669<br>lag 10: EDF=0.21, p=0.240 | lag 9: EDF=0.82, p=0.113<br>**lag 6: EDF=0.87, p=0.011**<br>lag 10: EDF=0.84, p=0.053<br>lag 0: EDF=0.00, p=0.734 | lag 2: EDF=0.45, p=0.462<br>**lag 8: EDF=0.94, p=0.003**<br>lag 6: EDF=0.72, p=0.257<br>lag 10: EDF=0.47, p=0.205 | **lag 10: EDF=2.04, p=0.025**<br>**lag 10: EDF=0.75, p=0.040**<br>**lag 3: EDF=0.85, p=0.013**<br>lag 7: EDF=0.49, p=0.148 | lag 6: EDF=0.51, p=0.343<br>lag 7: EDF=0.00, p=0.483<br>**lag 5: EDF=0.92, p=0.035**<br>lag 2: EDF=0.00, p=0.355 |
| Sea-level pressure difference (SLPD) | **lag 4: 0.87 (0.76–1.00), p=0.050**<br>lag 9: 0.96 (0.89–1.03), p=0.217<br>**lag 9: 0.87 (0.77–0.99), p=0.035**<br>**lag 0: 0.93 (0.88–0.99), p=0.030** | lag 6: 1.06 (0.96–1.17), p=0.268<br>lag 5: 1.03 (0.97–1.09), p=0.298<br>lag 9: 1.02 (0.93–1.12), p=0.676<br>**lag 0: 0.95 (0.90–1.00), p=0.035** | lag 10: 0.85 (0.71–1.02), p=0.087<br>**lag 0: 1.14 (1.05–1.24), p=0.001**<br>**lag 1: 0.84 (0.72–0.99), p=0.035**<br>lag 10: 1.02 (0.95–1.10), p=0.572 | **lag 10: 0.85 (0.73–0.98), p=0.026**<br>**lag 5: 0.92 (0.86–0.99), p=0.019**<br>lag 6: 0.88 (0.77–1.01), p=0.061<br>lag 1: 1.03 (0.96–1.09), p=0.442 | lag 6: 0.80 (0.58–1.10), p=0.176<br>lag 8: 1.09 (0.95–1.25), p=0.225<br>**lag 9: 0.72 (0.53–0.98), p=0.037**<br>lag 10: 0.91 (0.81–1.03), p=0.135 | lag 9: 1.12 (0.92–1.37), p=0.247<br>**lag 10: 0.89 (0.81–0.98), p=0.020**<br>**lag 0: 0.78 (0.65–0.93), p=0.005**<br>**lag 3: 0.87 (0.80–0.95), p=0.002** | lag 3: 1.12 (0.94–1.34), p=0.191<br>lag 5: 1.08 (0.99–1.18), p=0.091<br>lag 3: 0.87 (0.75–1.01), p=0.067<br>lag 10: 0.94 (0.87–1.01), p=0.098 | **lag 1: 0.71 (0.52–0.96), p=0.026**<br>**lag 5: 1.17 (1.01–1.35), p=0.034**<br>lag 3: 1.23 (0.95–1.58), p=0.110<br>lag 3: 0.89 (0.79–1.01), p=0.074 | lag 1: 1.25 (0.84–1.88), p=0.274<br>**lag 2: 0.76 (0.64–0.91), p=0.003**<br>lag 6: 0.76 (0.55–1.06), p=0.107<br>**lag 1: 0.76 (0.66–0.89), p=<0.001** | **lag 0: 0.87 (0.76–1.00), p=0.044**<br>lag 1: 1.07 (1.00–1.14), p=0.057<br>lag 1: 1.11 (0.99–1.25), p=0.070<br>lag 10: 1.02 (0.96–1.08), p=0.480 | **lag 9: 1.41 (1.06–1.88), p=0.020**<br>**lag 0: 0.87 (0.75–1.00), p=0.045**<br>**lag 7: 0.68 (0.52–0.90), p=0.006**<br>**lag 6: 1.13 (1.00–1.29), p=0.048** |
| Wet-bulb globe temperature | lag 0: EDF=0.24, p=0.222 | **lag 0: EDF=0.85, p=0.011**<br>**lag 0: 0.99 (0.98–0.99), p=<0.001** | lag 6: EDF=0.00, p=0.380<br>lag 0: EDF=0.00, p=0.670 | - | lag 10: EDF=0.01, p=0.547<br>lag 0: EDF=0.00, p=0.589<br>lag 10: | lag 7: EDF=0.66, p=0.067 | lag 7: EDF=0.01, p=0.370<br>**lag 6: EDF=0.84, p=0.017** | **lag 6: EDF=3.75, p=0.047**<br>lag 2: EDF=0.00, p=0.792<br>**lag 5:** | **lag 9: EDF=1.10, p=0.027**<br>**lag 9: EDF=4.14, p=0.001** | lag 8: EDF=0.00, p=0.682 | **lag 1: EDF=5.54, p=0.010**<br>lag 2: EDF=0.84, p=0.096<br>**lag 9:** |

| Atmospheric factor | 1.1 | 1.2 | 2.1 | 2.2 | 3.1 | 4.1 | 5.1 | 5.2 | 6.1 | 6.2 | 6.3 |
|---|---|---|---|---|---|---|---|---|---|---|---|
| | | lag 0: EDF=0.00, p=0.563 | | | EDF=0.00, p=0.452 | | | **EDF=0.84, p=0.014** | | | **EDF=0.80, p=0.030** |
| Daily mean temperature | lag 0: EDF=0.00, p=0.398<br>lag 2: EDF=1.96, p=0.088<br>**lag 1: EDF=2.97, p=<0.001** | lag 0: EDF=0.66, p=0.081<br>lag 0: EDF=0.00, p=0.747 | lag 0: EDF=0.65, p=0.091<br>lag 2: EDF=0.29, p=0.233 | lag 0: EDF=0.00, p=0.625<br>lag 2: EDF=0.00, p=0.837<br>lag 0: EDF=0.17, p=0.259 | lag 5: EDF=0.39, p=0.180 | lag 7: EDF=0.00, p=0.748<br>lag 0: EDF=0.00, p=0.744<br>lag 3: EDF=0.00, p=0.425 | **lag 4: EDF=0.94, p=0.003**<br>lag 1: EDF=0.00, p=0.699 | lag 0: EDF=2.21, p=0.163 | **lag 10: EDF=2.92, p=0.009**<br>**lag 9: 0.96 (0.92–0.99), p=0.017**<br>lag 4: EDF=0.00, p=0.365 | lag 0: EDF=0.29, p=0.213<br>lag 1: EDF=0.04, p=0.271 | **lag 0: EDF=0.74, p=0.046** |
| Ice day | lag 5: 0.93 (0.68–1.28), p=0.667<br>lag 1: 0.95 (0.84–1.07), p=0.362<br>**lag 0: 0.74 (0.58–0.95), p=0.017**<br>lag 2: 0.91 (0.80–1.03), p=0.137 | lag 5: 0.89 (0.71–1.12), p=0.316<br>lag 10: 1.00 (0.89–1.14), p=0.961<br>lag 5: 0.90 (0.73–1.11), p=0.335<br>lag 1: 0.93 (0.84–1.03), p=0.176 | **lag 7: 1.29 (1.01–1.64), p=0.044**<br>lag 0: 1.08 (0.97–1.21), p=0.167<br>lag 6: 1.06 (0.97–1.16), p=0.203 | **lag 10: 0.72 (0.55–0.94), p=0.014**<br>**lag 1: 0.83 (0.73–0.93), p=0.002**<br>lag 8: 1.14 (0.95–1.36), p=0.172<br>**lag 6: 0.89 (0.81–0.99), p=0.032** | lag 4: 1.07 (0.75–1.53), p=0.709<br>lag 3: 0.99 (0.85–1.16), p=0.895<br>lag 5: 1.32 (1.00–1.75), p=0.051<br>lag 4: 0.98 (0.85–1.14), p=0.801 | lag 10: 1.13 (0.77–1.67), p=0.536<br>lag 5: 0.98 (0.82–1.17), p=0.802<br>lag 6: 1.18 (0.84–1.66), p=0.334<br>lag 4: 0.87 (0.73–1.04), p=0.129 | lag 8: 1.03 (0.92–1.16), p=0.569<br>**lag 4: 0.68 (0.55–0.83), p=<0.001**<br>lag 2: 1.04 (0.94–1.15), p=0.452 | lag 2: 1.21 (0.88–1.66), p=0.245<br>**lag 3: 0.79 (0.66–0.94), p=0.008**<br>lag 7: 1.03 (0.78–1.35), p=0.852<br>lag 3: 1.01 (0.89–1.14), p=0.925 | lag 2: 0.63 (0.24–1.66), p=0.353<br>lag 8: 0.81 (0.58–1.11), p=0.185<br>lag 7: 1.31 (0.75–2.28), p=0.343<br>**lag 2: 1.34 (1.07–1.68), p=0.011** | lag 7: 1.18 (0.94–1.49), p=0.155<br>**lag 2: 0.90 (0.81–0.99), p=0.034**<br>lag 4: 0.87 (0.71–1.06), p=0.169<br>lag 4: 0.95 (0.87–1.03), p=0.221 | lag 9: 0.83 (0.45–1.56), p=0.572<br>lag 3: 0.82 (0.62–1.08), p=0.150<br>**lag 0: 0.46 (0.25–0.84), p=0.011**<br>lag 5: 0.90 (0.71–1.13), p=0.359 |
| Frost day | **lag 0: 1.14 (1.01–1.29), p=0.034**<br>**lag 7: 1.10 (1.04–1.17), p=0.002**<br>lag 0: 1.13 | lag 2: 1.08 (0.98–1.19), p=0.116<br>lag 1: 1.05 (1.00–1.10), p=0.074<br>lag 0: 1.01 | lag 3: 1.10 (0.97–1.25), p=0.126<br>lag 0: 1.06 (0.99–1.13), p=0.084<br>lag 3: 1.10 | lag 5: 1.05 (0.95–1.17), p=0.327<br>lag 1: 1.01 (0.95–1.06), p=0.831<br>lag 1: 1.04 | lag 6: 1.19 (0.97–1.46), p=0.094<br>lag 9: 1.08 (1.00–1.18), p=0.061<br>lag 10: 0.99 | lag 9: 1.03 (0.83–1.27), p=0.796<br>lag 1: 1.06 (0.96–1.17), p=0.268<br>lag 2: 1.08 | lag 2: 1.03 (0.89–1.18), p=0.693<br>lag 2: 1.06 (1.00–1.13), p=0.071<br>lag 0: 1.11 | **lag 6: 1.38 (1.04–1.83), p=0.027**<br>lag 5: 1.06 (0.96–1.17), p=0.278<br>lag 0: 1.22 | lag 3: 0.89 (0.59–1.36), p=0.595<br>lag 6: 1.08 (0.92–1.28), p=0.336<br>lag 1: 1.21 | lag 8: 1.12 (1.00–1.26), p=0.057<br>**lag 1: 1.07 (1.01–1.13), p=0.014**<br>lag 2: 1.05 | **lag 1: 1.58 (1.16–2.16), p=0.004**<br>**lag 7: 1.21 (1.07–1.37), p=0.003**<br>lag 5: 1.23 (0.96–1.59), p=0.100 |

| Atmospheric factor | 1.1 | 1.2 | 2.1 | 2.2 | 3.1 | 4.1 | 5.1 | 5.2 | 6.1 | 6.2 | 6.3 |
|---|---|---|---|---|---|---|---|---|---|---|---|
| | (1.00–1.29), p=0.052<br>**lag 7: 1.11 (1.04–1.18), p=0.001** | (0.91–1.11), p=0.897<br>**lag 7: 1.09 (1.04–1.15), p=<0.001** | (0.98–1.22), p=0.101<br>lag 10: 1.05 (1.00–1.10), p=0.060 | (0.94–1.14), p=0.475<br>lag 1: 1.01 (0.96–1.06), p=0.640 | (0.80–1.22), p=0.913<br>lag 5: 1.05 (0.96–1.14), p=0.281 | (0.89–1.32), p=0.430<br>**lag 8: 1.12 (1.02–1.22), p=0.013** | (0.99–1.24), p=0.067<br>**lag 5: 1.07 (1.02–1.13), p=0.012** | (0.96–1.55), p=0.105<br>lag 10: 1.07 (0.98–1.16), p=0.111 | (0.89–1.63), p=0.222<br>lag 2: 1.09 (0.95–1.26), p=0.208 | (0.95–1.17), p=0.324<br>**lag 9: 1.08 (1.03–1.12), p=<0.001** | lag 3: 0.93 (0.83–1.05), p=0.256 |
| Heat wave intensity | lag 7: EDF=0.00, p=0.628<br>lag 7: EDF=0.00, p=0.925<br>lag 5: EDF=0.00, p=0.431<br>lag 9: EDF=0.19, p=0.250 | lag 0: EDF=0.00, p=0.444<br>lag 3: EDF=0.39, p=0.180<br>lag 3: EDF=0.62, p=0.101<br>lag 1: EDF=0.34, p=0.199 | lag 3: EDF=0.00, p=0.776<br>lag 8: EDF=0.68, p=0.062<br>lag 8: EDF=0.17, p=0.281<br>lag 6: EDF=0.72, p=0.054 | lag 5: EDF=0.07, p=0.306<br>**lag 3: EDF=2.71, p=0.010**<br>lag 2: EDF=0.62, p=0.093<br>lag 9: EDF=0.00, p=0.335 | lag 8: EDF=0.40, p=0.356<br>lag 6: EDF=0.60, p=0.118<br>lag 10: EDF=0.62, p=0.204<br>lag 9: EDF=0.00, p=0.346 | lag 4: EDF=0.00, p=0.595<br>lag 10: EDF=0.64, p=0.094<br>**lag 3: EDF=0.83, p=0.045**<br>**lag 2: EDF=0.75, p=0.041** | lag 8: EDF=0.00, p=0.985<br>**lag 10: EDF=0.77, p=0.025**<br>lag 8: EDF=0.00, p=0.847<br>lag 1: EDF=0.00, p=0.629 | lag 9: EDF=0.01, p=0.499<br>lag 8: EDF=0.00, p=0.566<br>lag 1: EDF=0.70, p=0.130<br>**lag 4: EDF=2.57, p=<0.001** | lag 0: EDF=4.54, p=0.181<br>lag 10: EDF=0.77, p=0.087<br>**lag 5: EDF=0.95, p=0.005**<br>lag 3: EDF=0.00, p=0.901 | lag 4: EDF=0.75, p=0.059<br>lag 5: EDF=0.46, p=0.161<br>lag 3: EDF=0.00, p=0.618<br>**lag 10: EDF=0.75, p=0.042** | lag 3: EDF=0.10, p=0.483<br>lag 6: EDF=0.00, p=0.601<br>lag 6: EDF=0.00, p=0.765<br>lag 10: EDF=0.33, p=0.243 |
| Heat stress wave | lag 3: 0.84 (0.65–1.07), p=0.160<br>**lag 7: 1.17 (1.06–1.30), p=0.003**<br>**lag 5: 0.78 (0.63–0.98), p=0.034**<br>lag 8: 0.94 (0.84–1.05), p=0.249 | **lag 5: 1.19 (1.02–1.38), p=0.024**<br>**lag 0: 1.19 (1.09–1.30), p=<0.001**<br>lag 2: 0.96 (0.81–1.14), p=0.618<br>**lag 3: 1.09 (1.01–1.18), p=0.023** | **lag 3: 0.69 (0.52–0.91), p=0.008**<br>lag 7: 0.90 (0.80–1.00), p=0.057<br>lag 7: 1.21 (1.00–1.48), p=0.053<br>lag 4: 0.95 (0.86–1.05), p=0.287 | lag 6: 0.91 (0.74–1.11), p=0.348<br>**lag 1: 1.12 (1.02–1.23), p=0.013**<br>**lag 5: 0.77 (0.63–0.94), p=0.009**<br>lag 0: 0.99 (0.91–1.09), p=0.899 | lag 10: 0.62 (0.35–1.13), p=0.117<br>lag 8: 1.20 (1.00–1.44), p=0.056<br>lag 9: 1.38 (0.93–2.06), p=0.109<br>**lag 4: 1.21 (1.04–1.41), p=0.012** | lag 3: 1.21 (0.93–1.58), p=0.164<br>lag 1: 1.15 (0.99–1.34), p=0.060<br>lag 10: 0.90 (0.66–1.24), p=0.522<br>lag 4: 1.00 (0.87–1.15), p=0.999 | lag 8: 0.80 (0.59–1.07), p=0.132<br>lag 7: 0.91 (0.80–1.04), p=0.152<br>lag 5: 0.87 (0.68–1.11), p=0.248<br>lag 1: 0.91 (0.82–1.02), p=0.120 | lag 10: 0.59 (0.30–1.13), p=0.110<br>lag 2: 0.94 (0.74–1.18), p=0.584<br>lag 3: 1.24 (0.85–1.81), p=0.259<br>**lag 3: 0.73 (0.58–0.91), p=0.006** | **lag 0: 2.69 (1.16–6.26), p=0.021**<br>lag 4: 1.06 (0.79–1.44), p=0.693<br>lag 0: 0.46 (0.21–1.01), p=0.054<br>lag 4: 0.83 (0.63–1.09), p=0.184 | **lag 6: 1.27 (1.02–1.58), p=0.031**<br>lag 4: 0.94 (0.84–1.05), p=0.286<br>lag 1: 1.12 (0.94–1.33), p=0.215<br>lag 7: 0.92 (0.84–1.01), p=0.073 | lag 3: 1.28 (0.83–1.98), p=0.270<br>**lag 6: 1.23 (1.00–1.51), p=0.045**<br>**lag 10: 1.50 (1.03–2.18), p=0.033**<br>**lag 2: 1.37 (1.15–1.63), p=<0.001** |
| Cold wave | lag 3: 1.00 (0.90–1.10), | lag 3: 1.17 (0.96–1.42), | lag 4: 0.86 (0.63–1.19), | lag 10: 1.23 (0.99–1.52), | lag 0: 1.25 (0.83–1.87), | lag 9: 1.29 (0.90–1.85), | **lag 7: 1.33 (1.01–1.76),** | lag 8: 1.16 (0.97–1.39), | **lag 10: 1.48 (1.16–1.90),** | **lag 6: 1.26 (1.01–** | lag 0: 1.49 (0.81–2.73), p=0.197 |

| Atmospheric factor | 1.1 | 1.2 | 2.1 | 2.2 | 3.1 | 4.1 | 5.1 | 5.2 | 6.1 | 6.2 | 6.3 |
|---|---|---|---|---|---|---|---|---|---|---|---|
| | p=0.930<br>**lag 2: 1.50 (1.21–1.86), p=<0.001**<br>lag 7: 1.06 (0.97–1.17), p=0.212 | p=0.120<br>lag 10: 1.05 (0.95–1.15), p=0.339<br>**lag 10: 1.23 (1.07–1.41), p=0.003** | p=0.374<br>**lag 1: 1.42 (1.07–1.88), p=0.016**<br>**lag 3: 1.10 (1.01–1.21), p=0.037** | p=0.061<br>**lag 7: 1.11 (1.01–1.21), p=0.023**<br>**lag 10: 1.25 (1.07–1.46), p=0.004**<br>lag 2: 1.06 (0.97–1.16), p=0.213 | p=0.280<br>**lag 9: 1.39 (1.01–1.90), p=0.041**<br>lag 3: 1.11 (0.93–1.33), p=0.257 | p=0.164<br>lag 0: 1.12 (0.82–1.53), p=0.474<br>lag 4: 1.02 (0.87–1.19), p=0.842 | **p=0.045**<br>**lag 6: 1.17 (1.03–1.32), p=0.016**<br>lag 10: 1.00 (0.80–1.24), p=0.989 | p=0.114<br>lag 9: 1.11 (0.79–1.56), p=0.536<br>lag 5: 1.05 (0.89–1.24), p=0.567 | **p=0.002**<br>lag 10: 1.33 (0.82–2.15), p=0.245 | **1.58), p=0.039** | lag 10: 0.94 (0.76–1.16), p=0.541 |
| Cold wave duration | lag 0: EDF=0.00, p=0.752 | **lag 3: EDF=0.71, p=0.046**<br>lag 5: EDF=0.00, p=0.892<br>lag 3: EDF=0.00, p=0.924<br>lag 9: EDF=0.00, p=0.864 | lag 1: EDF=0.00, p=0.541<br>lag 1: EDF=0.75, p=0.054<br>lag 2: EDF=0.00, p=0.666 | lag 10: EDF=0.11, p=0.290<br>lag 0: EDF=0.15, p=0.257<br>lag 10: EDF=0.00, p=0.860<br>lag 6: EDF=0.00, p=0.656 | lag 1: EDF=0.00, p=0.748<br>**lag 10: EDF=0.84, p=0.009**<br>lag 10: EDF=0.00, p=0.730<br>lag 3: EDF=0.00, p=0.724 | **lag 3: EDF=0.83, p=0.041**<br>lag 8: EDF=0.00, p=0.555 | lag 10: EDF=0.00, p=0.621 | lag 5: EDF=0.77, p=0.141<br>lag 9: EDF=0.15, p=0.383<br>**lag 8: EDF=0.78, p=0.026** | **lag 2: EDF=2.20, p=0.036**<br>**lag 8: EDF=0.94, p=<0.001** | lag 0: EDF=0.00, p=0.638 | lag 0: EDF=0.80, p=0.138<br>lag 2: EDF=0.00, p=0.702 |
| Cold wave intensity | lag 10: EDF=0.00, p=0.729<br>**lag 4: EDF=0.89, p=0.003** | - | **lag 0: EDF=0.89, p=0.001**<br>lag 9: EDF=0.01, p=0.367<br>lag 3: EDF=0.00, p=0.933 | lag 8: EDF=0.00, p=0.727<br>lag 2: EDF=0.00, p=0.362 | lag 8: EDF=0.57, p=0.120 | lag 6: EDF=0.00, p=0.604<br>lag 10: EDF=0.00, p=0.821<br>lag 2: EDF=0.00, p=0.923 | lag 6: EDF=0.10, p=0.272<br>lag 7: EDF=0.34, p=0.205 | **lag 4: EDF=0.84, p=0.017** | lag 0: EDF=0.78, p=0.234<br>**lag 3: EDF=0.80, p=0.038** | **lag 3: EDF=0.83, p=0.019**<br>lag 1: EDF=0.00, p=0.781 | lag 0: EDF=0.00, p=0.788 |
| Cold stress wave | **lag 3: 0.70 (0.52–0.95), p=0.023** | lag 8: 0.92 (0.79–1.07), p=0.260 | lag 0: 1.07 (0.86–1.34), p=0.535 | **lag 7: 1.23 (1.01–1.51), p=0.040** | **lag 10: 3.55 (1.69–7.47), p=<0.001** | lag 9: 0.91 (0.78–1.06), p=0.246 | **lag 0: 1.12 (1.00–1.26), p=0.042** | lag 3: 1.58 (0.99–2.53), p=0.056 | lag 0: 0.80 (0.30–2.17), p=0.666 | **lag 10: 1.18 (1.09–1.28), p=<0.001** | lag 7: 0.70 (0.38–1.29), p=0.247<br>lag 0: 0.97 |

| Atmospheric factor | 1.1 | 1.2 | 2.1 | 2.2 | 3.1 | 4.1 | 5.1 | 5.2 | 6.1 | 6.2 | 6.3 |
|---|---|---|---|---|---|---|---|---|---|---|---|
| | lag 10: 1.00 (0.91–1.10), p=0.975<br>lag 4: 1.08 (0.86–1.35), p=0.522<br>lag 1: 0.89 (0.80–1.00), p=0.056 | lag 5: 0.93 (0.79–1.09), p=0.388<br>lag 7: 0.99 (0.92–1.07), p=0.827 | lag 0: 1.04 (0.92–1.18), p=0.488 | **lag 1: 1.24 (1.13–1.37), p=<0.001**<br>lag 0: 0.99 (0.91–1.09), p=0.879 | lag 2: 1.14 (0.95–1.36), p=0.152<br>lag 5: 1.05 (0.89–1.23), p=0.561 | **lag 6: 1.46 (1.10–1.94), p=0.010** | lag 7: 1.11 (0.89–1.38), p=0.371<br>lag 0: 1.11 (1.00–1.23), p=0.058 | **lag 3: 1.35 (1.08–1.70), p=0.010**<br>lag 8: 0.92 (0.74–1.15), p=0.459 | lag 1: 0.81 (0.56–1.17), p=0.259<br>lag 1: 0.77 (0.42–1.40), p=0.387<br>**lag 9: 0.72 (0.55–0.96), p=0.024** | lag 0: 0.92 (0.76–1.12), p=0.405<br>lag 1: 1.03 (0.95–1.12), p=0.522 | (0.76–1.25), p=0.835<br>lag 6: 0.92 (0.73–1.15), p=0.449 |
| Cold stress wave intensity | lag 3: EDF=0.67, p=0.089<br>lag 4: EDF=0.01, p=0.342<br>lag 1: EDF=0.00, p=0.964 | **lag 1: EDF=0.89, p=0.002**<br>lag 10: EDF=0.00, p=0.905<br>lag 5: EDF=0.00, p=0.627 | lag 4: EDF=1.62, p=0.116 | **lag 8: EDF=0.71, p=0.048**<br>**lag 1: EDF=0.86, p=0.005** | **lag 10: EDF=6.30, p=0.002**<br>lag 7: EDF=0.00, p=0.805<br>lag 4: EDF=0.00, p=0.608 | lag 10: EDF=0.00, p=0.708 | lag 7: EDF=0.80, p=0.051<br>lag 2: EDF=0.00, p=0.487 | lag 10: EDF=4.85, p=0.075<br>lag 10: EDF=0.00, p=0.389<br>lag 0: EDF=0.58, p=0.187 | lag 2: EDF=0.72, p=0.245 | lag 2: EDF=0.16, p=0.257 | lag 8: EDF=0.33, p=0.408<br>lag 6: EDF=0.00, p=0.682 |
| Ozone | lag 7: EDF=0.27, p=0.255<br>lag 0: EDF=0.00, p=0.372<br>lag 3: EDF=0.00, p=0.201<br>lag 7: EDF=0.00, p=0.338 | lag 5: EDF=0.50, p=0.139<br>lag 2: EDF=0.00, p=0.873<br>lag 4: EDF=0.00, p=0.685 | lag 2: EDF=0.36, p=0.194<br>lag 4: EDF=0.00, p=0.798 | lag 0: EDF=0.00, p=0.570<br>lag 7: EDF=0.00, p=0.523 | lag 3: EDF=2.44, p=0.250<br>lag 7: EDF=0.00, p=0.550<br>lag 2: EDF=0.46, p=0.304 | lag 7: EDF=0.68, p=0.136<br>**lag 7: EDF=0.74, p=0.049**<br>**lag 6: EDF=0.90, p=0.011**<br>**lag 1: EDF=2.09, p=0.001** | lag 4: EDF=0.41, p=0.171<br>lag 0: EDF=0.00, p=0.711<br>lag 4: EDF=0.00, p=0.969 | - | lag 2: EDF=0.89, p=0.126<br>lag 7: EDF=0.00, p=0.902<br>lag 0: EDF=3.66, p=0.340<br>lag 5: EDF=0.65, p=0.120 | lag 1: EDF=0.01, p=0.590<br>lag 6: EDF=0.00, p=0.700<br>lag 7: EDF=0.00, p=0.388 | **lag 4: 1.02 (1.00–1.03), p=0.004**<br>lag 6: EDF=0.75, p=0.057<br>lag 7: EDF=0.00, p=0.490<br>lag 1: EDF=0.44, p=0.173 |
| Particulate matter ($PM_{10}$) | lag 5: EDF=0.53, p=0.166<br>lag 2: EDF=0.07, | lag 0: EDF=0.00, p=0.403<br>**lag 7: EDF=0.74,** | lag 3: EDF=0.00, p=0.493<br>lag 7: EDF=0.27, | lag 3: EDF=0.06, p=0.308<br>lag 4: EDF=0.00, | lag 6: EDF=0.43, p=0.332<br>lag 4: EDF=0.69, | **lag 3: EDF=2.80, p=<0.001**<br>lag 4: EDF=0.00, | lag 2: EDF=0.42, p=0.235<br>lag 6: EDF=0.08, | lag 5: EDF=0.58, p=0.289<br>lag 4: EDF=0.01, | **lag 2: EDF=0.94, p=0.041**<br>lag 4: EDF=0.00, | lag 3: EDF=0.70, p=0.076<br>lag 7: EDF=0.36, | **lag 4: EDF=0.89, p=0.047**<br>lag 0: EDF=0.21, |

| Atmospheric factor | 1.1 | 1.2 | 2.1 | 2.2 | 3.1 | 4.1 | 5.1 | 5.2 | 6.1 | 6.2 | 6.3 |
|---|---|---|---|---|---|---|---|---|---|---|---|
| | p=0.279<br>lag 2: EDF=0.00, p=0.372<br>lag 3: EDF=0.00, p=0.421 | **p=0.044**<br>lag 1: EDF=0.00, p=0.970<br>lag 5: EDF=0.32, p=0.210 | p=0.224<br>lag 5: EDF=0.37, p=0.218<br>lag 6: EDF=0.00, p=0.614 | p=0.342<br>lag 7: EDF=0.00, p=0.891<br>lag 4: EDF=0.00, p=0.874 | p=0.072<br>lag 7: EDF=0.62, p=0.220<br>lag 6: EDF=0.00, p=0.717 | p=0.798<br>lag 2: EDF=0.48, p=0.232<br>lag 6: EDF=0.00, p=0.559 | p=0.283<br>**lag 1: EDF=0.85, p=0.013**<br>lag 3: EDF=0.00, p=0.858 | p=0.442<br>lag 7: EDF=0.42, p=0.298<br>lag 7: EDF=0.04, p=0.294 | p=0.710<br>lag 2: EDF=0.22, p=0.490<br>lag 4: EDF=0.65, p=0.120 | p=0.192<br>lag 2: EDF=0.00, p=0.426<br>lag 3: EDF=0.00, p=0.639 | p=0.290<br>**lag 0: EDF=4.66, p=0.005**<br>lag 3: EDF=0.00, p=0.910 |

**Table S9**. Overview of time lags, effect sizes (effective degree of freedom (EDF) or relative risk (95% confidence interval)) and p-values per human-bioclimatic region, atmospheric factor and subgroup (pink = female <60 years, orange = females ≥60 years, green = males <60 years, blue = males ≥60 years) for epilepsy, significant p-values (≤ 0.05) are bold

| Atmospheric factor | 1.1 | 1.2 | 2.1 | 2.2 | 3.1 | 4.1 | 5.1 | 5.2 | 6.1 | 6.2 | 6.3 |
|---|---|---|---|---|---|---|---|---|---|---|---|
| Sea-level pressure difference (SLPD) | lag 5: 0.91 (0.81–1.03), p=0.154<br>**lag 7: 0.87 (0.79–0.97), p=0.015**<br>lag 5: 0.96 (0.87–1.06), p=0.405<br>**lag 0: 0.88 (0.79–0.98), p=0.023** | lag 5: 0.90 (0.81–1.00), p=0.058<br>lag 6: 1.08 (0.98–1.18), p=0.128<br>**lag 7: 0.91 (0.84–0.99), p=0.024**<br>lag 10: 1.07 (0.97–1.18), p=0.186 | **lag 1: 0.86 (0.77–0.97), p=0.011**<br>**lag 2: 1.10 (1.00–1.21), p=0.047**<br>**lag 4: 0.91 (0.84–1.00), p=0.047**<br>lag 0: 0.96 (0.87–1.06), p=0.421 | lag 9: 0.88 (0.77–1.01), p=0.066<br>lag 2: 1.07 (0.97–1.19), p=0.182<br>lag 2: 1.08 (0.97–1.20), p=0.147<br>**lag 8: 0.88 (0.79–0.98), p=0.017** | lag 0: 0.84 (0.67–1.06), p=0.137<br>lag 1: 0.87 (0.73–1.04), p=0.129<br>**lag 10: 0.82 (0.69–0.97), p=0.018**<br>lag 7: 0.91 (0.75–1.09), p=0.303 | **lag 9: 0.80 (0.66–0.98), p=0.030**<br>lag 6: 0.88 (0.76–1.02), p=0.096<br>lag 9: 1.15 (0.99–1.33), p=0.074<br>lag 2: 0.89 (0.76–1.06), p=0.187 | **lag 1: 0.85 (0.74–0.98), p=0.027**<br>**lag 6: 0.85 (0.75–0.96), p=0.008**<br>lag 5: 1.10 (0.98–1.23), p=0.110<br>lag 8: 0.89 (0.79–1.01), p=0.064 | lag 5: 0.85 (0.70–1.03), p=0.090<br>lag 1: 1.16 (0.98–1.37), p=0.092<br>**lag 6: 1.21 (1.04–1.41), p=0.014**<br>lag 2: 0.84 (0.71–1.00), p=0.051 | lag 0: 0.81 (0.57–1.14), p=0.220<br>lag 4: 1.14 (0.90–1.43), p=0.273<br>lag 3: 0.85 (0.66–1.09), p=0.193<br>lag 0: 1.25 (0.97–1.61), p=0.083 | **lag 9: 0.81 (0.72–0.91), p=<0.001**<br>lag 9: 1.06 (0.97–1.15), p=0.196<br>lag 0: 0.95 (0.87–1.04), p=0.287<br>lag 7: 1.09 (0.99–1.19), p=0.068 | **lag 1: 1.42 (1.09–1.84), p=0.010**<br>**lag 10: 0.79 (0.63–0.99), p=0.043**<br>**lag 0: 1.31 (1.07–1.60), p=0.009**<br>**lag 3: 1.34 (1.07–1.68), p=0.010** |
| Wind day | **lag 1: 1.17 (1.01–1.36), p=0.035**<br>lag 7: 1.11 (0.97–1.27), p=0.115<br>lag 6: 0.88 | **lag 10: 1.16 (1.02–1.32), p=0.023**<br>lag 1: 1.06 (0.94–1.20), p=0.314<br>lag 5: 0.91 | **lag 2: 1.13 (1.00–1.27), p=0.047**<br>lag 5: 0.91 (0.81–1.01), p=0.071<br>lag 5: 1.08 | lag 4: 0.93 (0.80–1.08), p=0.354<br>**lag 3: 1.13 (1.01–1.26), p=0.038**<br>**lag 5: 1.13** | lag 0: 0.84 (0.65–1.09), p=0.191<br>lag 8: 1.14 (0.96–1.36), p=0.141<br>lag 9: 0.85 | **lag 0: 0.73 (0.54–0.99), p=0.045**<br>lag 10: 1.17 (0.97–1.41), p=0.094<br>**lag 3: 1.24** | lag 1: 1.15 (0.98–1.35), p=0.086<br>lag 9: 1.13 (0.98–1.29), p=0.086<br>lag 9: 0.89 | lag 10: 1.19 (0.96–1.47), p=0.108<br>**lag 3: 0.79 (0.63–0.98), p=0.033**<br>lag 6: 0.91 | **lag 0: 1.71 (1.18–2.47), p=0.005**<br>lag 8: 1.17 (0.88–1.55), p=0.274<br>**lag 4: 0.64** | **lag 7: 1.19 (1.05–1.35), p=0.008**<br>lag 8: 1.06 (0.96–1.17), p=0.250<br>**lag 3: 1.15** | **lag 2: 0.63 (0.41–0.97), p=0.035**<br>lag 10: 1.30 (0.99–1.71), p=0.057<br>**lag 0: 1.56** |

| Atmospheric factor | 1.1 | 1.2 | 2.1 | 2.2 | 3.1 | 4.1 | 5.1 | 5.2 | 6.1 | 6.2 | 6.3 |
|---|---|---|---|---|---|---|---|---|---|---|---|
| | (0.78–1.01), p=0.068<br>lag 9: 0.86 (0.74–1.00), p=0.056 | (0.82–1.02), p=0.116<br>lag 7: 1.08 (0.96–1.23), p=0.208 | (0.98–1.19), p=0.106<br>**lag 1: 0.87 (0.77–0.98), p=0.021** | **(1.01–1.26), p=0.027**<br>lag 1: 1.10 (0.98–1.23), p=0.119 | (0.70–1.03), p=0.092<br>**lag 0: 1.24 (1.04–1.48), p=0.017** | **(1.03–1.50), p=0.025**<br>lag 2: 1.14 (0.93–1.41), p=0.216 | (0.78–1.03), p=0.112<br>**lag 10: 1.20 (1.05–1.38), p=0.010** | (0.76–1.10), p=0.331<br>lag 6: 1.14 (0.94–1.38), p=0.182 | **(0.43–0.96), p=0.030**<br>**lag 0: 1.46 (1.09–1.96), p=0.011** | **(1.04–1.28), p=0.005**<br>lag 10: 1.06 (0.96–1.18), p=0.265 | **(1.23–1.97), p=<0.001**<br>lag 4: 1.07 (0.80–1.44), p=0.634 |
| Wet-bulb globe temperature | lag 9: EDF=0.71, p=0.058<br>lag 4: EDF=0.00, p=0.400<br>lag 2: EDF=0.00, p=0.347<br>lag 4: EDF=0.00, p=0.360 | lag 4: EDF=0.01, p=0.244<br>lag 2: EDF=0.00, p=0.420<br>lag 2: EDF=0.67, p=0.051 | lag 6: EDF=0.00, p=0.994<br>lag 9: EDF=0.00, p=0.874<br>lag 4: EDF=0.00, p=0.411<br>lag 2: EDF=0.00, p=0.804 | **lag 10: EDF=0.89, p=0.003**<br>lag 1: EDF=0.00, p=0.452<br>lag 9: EDF=0.36, p=0.179 | lag 5: EDF=0.01, p=0.371<br>lag 3: EDF=0.00, p=0.985<br>**lag 8: EDF=0.75, p=0.048** | lag 9: EDF=0.00, p=0.837<br>lag 3: EDF=0.24, p=0.268<br>lag 7: EDF=0.00, p=0.429<br>lag 10: EDF=0.00, p=0.563 | lag 0: EDF=0.00, p=0.402<br>lag 0: EDF=0.02, p=0.271<br>**lag 1: EDF=0.76, p=0.026** | lag 2: EDF=0.00, p=0.420<br>lag 0: EDF=0.00, p=0.492<br>lag 3: EDF=0.00, p=0.621 | lag 6: EDF=0.04, p=0.506<br>lag 2: EDF=0.00, p=0.575<br>lag 10: EDF=0.00, p=0.542<br>lag 3: EDF=0.00, p=0.732 | lag 6: EDF=0.00, p=0.590<br>lag 1: EDF=0.00, p=0.449<br>**lag 10: EDF=0.70, p=0.049**<br>lag 1: EDF=0.00, p=0.332 | lag 2: EDF=0.00, p=0.874<br>lag 2: EDF=0.64, p=0.192 |
| Frost day | **lag 7: 0.87 (0.77–0.99), p=0.035**<br>lag 7: 0.95 (0.85–1.06), p=0.326<br>lag 5: 0.91 (0.83–1.00), p=0.062<br>lag 8: 1.10 (0.99–1.21), p=0.065 | lag 3: 0.96 (0.86–1.07), p=0.423<br>lag 10: 0.98 (0.89–1.07), p=0.612<br>**lag 1: 0.90 (0.83–0.97), p=0.007**<br>lag 5: 0.96 (0.87–1.05), p=0.363 | lag 5: 0.95 (0.88–1.02), p=0.129<br>**lag 9: 1.07 (1.01–1.13), p=0.020**<br>**lag 6: 0.91 (0.86–0.97), p=0.002**<br>lag 2: 1.02 (0.95–1.09), p=0.635 | lag 1: 0.93 (0.85–1.02), p=0.133<br>lag 8: 0.95 (0.88–1.03), p=0.231<br>**lag 5: 0.93 (0.86–1.00), p=0.049**<br>lag 4: 1.05 (0.97–1.14), p=0.244 | **lag 9: 0.76 (0.65–0.88), p=<0.001**<br>lag 5: 0.97 (0.87–1.08), p=0.584<br>lag 0: 0.99 (0.89–1.09), p=0.801<br>**lag 6: 0.84 (0.75–0.95), p=0.004** | **lag 5: 1.26 (1.03–1.54), p=0.023**<br>lag 6: 0.97 (0.83–1.12), p=0.641<br>lag 3: 1.10 (0.93–1.30), p=0.264<br>**lag 0: 0.74 (0.62–0.88), p=<0.001** | lag 5: 0.94 (0.84–1.05), p=0.278<br>**lag 3: 0.90 (0.82–0.98), p=0.021**<br>lag 0: 0.94 (0.87–1.01), p=0.101<br>**lag 10: 1.16 (1.06–1.26), p=0.001** | **lag 0: 0.81 (0.72–0.91), p=<0.001**<br>lag 3: 1.05 (0.95–1.17), p=0.333<br>lag 1: 0.94 (0.84–1.05), p=0.263<br>lag 9: 1.11 (1.00–1.23), p=0.057 | lag 10: 0.71 (0.49–1.01), p=0.059<br>lag 6: 0.85 (0.69–1.04), p=0.114<br>lag 2: 0.85 (0.68–1.05), p=0.127<br>lag 1: 0.84 (0.68–1.05), p=0.128 | **lag 6: 0.86 (0.78–0.94), p=0.002**<br>lag 6: 1.05 (0.98–1.12), p=0.153<br>lag 0: 0.97 (0.90–1.04), p=0.340<br>lag 1: 1.06 (0.99–1.14), p=0.104 | **lag 5: 0.71 (0.55–0.92), p=0.009**<br>lag 3: 0.86 (0.69–1.07), p=0.166<br>lag 2: 0.91 (0.74–1.12), p=0.356<br>**lag 0: 0.61 (0.48–0.78), p=<0.001** |
| Heat wave day | lag 6: 1.01 (0.88–1.16), | lag 1: 0.96 (0.84–1.10), | lag 10: 1.04 (0.93–1.15), | lag 5: 0.88 (0.77–1.00), | lag 9: 1.11 (0.92–1.34), | lag 4: 1.24 (0.99–1.54), | lag 3: 1.08 (0.94–1.24), | lag 0: 1.09 (0.92–1.31), | lag 7: 1.29 (0.87–1.91), | lag 10: 1.06 (0.95–1.19), | lag 10: 0.84 (0.56–1.27), |

| Atmospheric factor | 1.1 | 1.2 | 2.1 | 2.2 | 3.1 | 4.1 | 5.1 | 5.2 | 6.1 | 6.2 | 6.3 |
|---|---|---|---|---|---|---|---|---|---|---|---|
| | p=0.880<br>**lag 0: 1.21 (1.05–1.39), p=0.009**<br>lag 5: 1.05 (0.94–1.17), p=0.357<br>**lag 1: 0.85 (0.75–0.97), p=0.017** | p=0.565<br>**lag 2: 0.88 (0.79–0.99), p=0.035**<br>lag 2: 1.09 (1.00–1.18), p=0.058<br>**lag 10: 0.86 (0.77–0.97), p=0.011** | p=0.492<br>**lag 1: 0.84 (0.77–0.92), p=<0.001**<br>lag 0: 1.07 (0.99–1.15), p=0.110<br>**lag 0: 0.83 (0.74–0.92), p=<0.001** | p=0.055<br>**lag 1: 0.89 (0.80–1.00), p=0.048**<br>lag 5: 1.06 (0.97–1.17), p=0.203<br>**lag 1: 0.83 (0.74–0.93), p=0.001** | p=0.280<br>**lag 4: 0.74 (0.61–0.89), p=0.002**<br>lag 10: 1.13 (0.98–1.30), p=0.085<br>**lag 10: 0.68 (0.55–0.85), p=<0.001** | p=0.058<br>lag 6: 0.85 (0.71–1.03), p=0.104<br>lag 5: 0.84 (0.68–1.03), p=0.094<br>**lag 0: 0.79 (0.64–0.98), p=0.029** | p=0.265<br>**lag 1: 0.86 (0.75–0.98), p=0.020**<br>lag 1: 1.07 (0.96–1.19), p=0.234<br>lag 7: 0.96 (0.82–1.12), p=0.593 | p=0.326<br>lag 8: 1.05 (0.86–1.26), p=0.647<br>**lag 7: 0.71 (0.59–0.85), p=<0.001**<br>lag 3: 0.88 (0.72–1.07), p=0.191 | p=0.197<br>lag 10: 0.78 (0.58–1.04), p=0.093<br>lag 5: 1.15 (0.87–1.51), p=0.330<br>lag 8: 1.13 (0.85–1.49), p=0.408 | p=0.313<br>lag 0: 1.00 (0.90–1.10), p=0.945<br>lag 4: 1.07 (0.98–1.18), p=0.140<br>**lag 0: 0.86 (0.77–0.95), p=0.005** | p=0.419<br>lag 4: 1.30 (0.99–1.70), p=0.056<br>lag 3: 1.05 (0.83–1.33), p=0.666<br>**lag 6: 1.34 (1.03–1.75), p=0.028** |
| Heat stress wave intensity | lag 9: EDF=0.00, p=0.810<br>**lag 0: EDF=0.84, p=0.010**<br>lag 8: EDF=0.52, p=0.118<br>lag 5: EDF=0.01, p=0.330 | lag 0: EDF=0.71, p=0.053<br>lag 0: EDF=0.71, p=0.051<br>lag 8: EDF=0.00, p=0.534<br>lag 1: EDF=0.53, p=0.128 | lag 2: EDF=0.45, p=0.155<br>lag 0: EDF=0.00, p=0.315<br>lag 0: EDF=0.00, p=0.738<br>lag 0: EDF=0.00, p=0.606 | lag 10: EDF=0.53, p=0.115<br>lag 2: EDF=0.68, p=0.052<br>lag 10: EDF=0.50, p=0.120<br>lag 1: EDF=0.00, p=0.377 | lag 0: EDF=0.65, p=0.106<br>**lag 8: EDF=0.83, p=0.015**<br>**lag 2: EDF=0.82, p=0.018**<br>lag 10: EDF=0.35, p=0.214 | lag 8: EDF=0.67, p=0.126<br>lag 3: EDF=0.15, p=0.303<br>lag 7: EDF=0.65, p=0.118<br>lag 1: EDF=0.03, p=0.356 | lag 5: EDF=0.00, p=0.519<br>lag 1: EDF=0.00, p=0.552<br>lag 1: EDF=0.00, p=0.631<br>lag 7: EDF=0.60, p=0.105 | lag 2: EDF=0.00, p=0.887<br>lag 8: EDF=0.72, p=0.061<br>**lag 8: EDF=0.91, p=<0.001**<br>lag 0: EDF=0.70, p=0.092 | lag 1: EDF=0.83, p=0.149<br>lag 2: EDF=0.00, p=0.856<br>lag 1: EDF=1.56, p=0.060<br>lag 6: EDF=0.28, p=0.366 | lag 3: EDF=0.00, p=0.722<br>lag 8: EDF=0.46, p=0.139<br>lag 1: EDF=0.65, p=0.075<br>lag 0: EDF=0.00, p=0.369 | lag 10: EDF=1.58, p=0.100<br>lag 10: EDF=0.67, p=0.189<br>lag 9: EDF=0.78, p=0.067<br>lag 6: EDF=0.00, p=0.501 |
| Cold wave duration | lag 7: EDF=0.00, p=0.561<br>lag 4: EDF=0.00, p=0.601<br>lag 0: EDF=0.00, p=0.581<br>lag 9: EDF=0.35, p=0.180 | lag 1: EDF=0.00, p=0.636<br>lag 10: EDF=0.00, p=0.541<br>lag 8: EDF=0.00, p=0.367 | lag 6: EDF=0.00, p=0.329<br>lag 4: EDF=0.00, p=0.490<br>lag 4: EDF=0.51, p=0.129 | **lag 5: EDF=0.75, p=0.034**<br>lag 3: EDF=0.43, p=0.157<br>lag 7: EDF=0.00, p=0.472<br>lag 10: EDF=0.31, p=0.199 | lag 0: EDF=0.38, p=0.248<br>lag 2: EDF=0.00, p=0.995<br>lag 3: EDF=0.01, p=0.345<br>lag 8: EDF=0.00, p=0.490 | lag 10: EDF=0.00, p=0.678<br>lag 1: EDF=0.47, p=0.176<br>lag 2: EDF=0.76, p=0.059 | lag 3: EDF=0.52, p=0.131<br>lag 3: EDF=0.00, p=0.455<br>lag 2: EDF=0.01, p=0.424 | lag 9: EDF=0.09, p=0.334<br>lag 0: EDF=0.46, p=0.178<br>lag 3: EDF=0.01, p=0.332<br>lag 1: EDF=0.00, p=0.343 | lag 10: EDF=0.64, p=0.330<br>**lag 0: EDF=0.95, p=<0.001**<br>lag 3: EDF=0.40, p=0.313 | lag 2: EDF=0.00, p=0.453<br>**lag 1: EDF=0.86, p=0.005**<br>lag 9: EDF=0.00, p=0.912<br>lag 7: EDF=0.00, p=0.779 | lag 9: EDF=0.00, p=0.810<br>lag 7: EDF=0.17, p=0.387<br>**lag 1: EDF=0.88, p=0.016** |

| **Atmospheric factor** | **1.1** | **1.2** | **2.1** | **2.2** | **3.1** | **4.1** | **5.1** | **5.2** | **6.1** | **6.2** | **6.3** |
|---|---|---|---|---|---|---|---|---|---|---|---|
| Cold stress wave | **lag 0: 1.21 (1.03–1.43), p=0.023**<br>lag 6: 0.95 (0.81–1.12), p=0.555<br>lag 0: 0.91 (0.79–1.05), p=0.209<br>lag 9: 1.12 (0.95–1.33), p=0.182 | lag 1: 0.90 (0.74–1.11), p=0.320<br>lag 10: 0.95 (0.82–1.10), p=0.457<br>lag 10: 0.93 (0.83–1.04), p=0.190<br>lag 10: 0.98 (0.85–1.12), p=0.732 | lag 6: 0.94 (0.82–1.08), p=0.370<br>lag 10: 0.91 (0.81–1.01), p=0.082<br>lag 2: 0.92 (0.82–1.02), p=0.118<br>lag 5: 1.00 (0.88–1.14), p=0.957 | lag 5: 1.08 (0.90–1.29), p=0.388<br>**lag 9: 1.14 (1.01–1.29), p=0.035**<br>**lag 8: 0.88 (0.78–1.00), p=0.047**<br>lag 1: 1.04 (0.91–1.19), p=0.568 | **lag 6: 1.57 (1.22–2.01), p=<0.001**<br>lag 0: 0.94 (0.76–1.16), p=0.555<br>lag 6: 1.15 (0.97–1.36), p=0.113<br>**lag 7: 1.25 (1.03–1.53), p=0.025** | lag 1: 0.88 (0.66–1.17), p=0.375<br>lag 10: 0.96 (0.78–1.20), p=0.743<br>lag 1: 1.04 (0.77–1.41), p=0.796<br>lag 7: 1.20 (0.94–1.53), p=0.146 | lag 2: 1.17 (0.97–1.42), p=0.102<br>lag 1: 0.96 (0.82–1.12), p=0.597<br>lag 10: 1.07 (0.94–1.21), p=0.316<br>lag 3: 1.04 (0.88–1.22), p=0.643 | lag 10: 0.98 (0.76–1.26), p=0.864<br>lag 0: 1.02 (0.78–1.33), p=0.906<br>lag 3: 1.15 (0.97–1.37), p=0.105<br>lag 10: 0.84 (0.67–1.05), p=0.129 | lag 10: 1.10 (0.54–2.24), p=0.786<br>**lag 8: 1.46 (1.07–1.99), p=0.016**<br>lag 9: 0.67 (0.44–1.01), p=0.058<br>**lag 3: 1.86 (1.30–2.66), p=<0.001** | lag 0: 1.16 (1.00–1.34), p=0.051<br>lag 3: 1.11 (0.98–1.26), p=0.096<br>**lag 2: 1.12 (1.00–1.25), p=0.049**<br>lag 4: 0.90 (0.80–1.01), p=0.079 | **lag 8: 1.75 (1.26–2.42), p=<0.001**<br>lag 7: 0.79 (0.55–1.14), p=0.210<br>lag 2: 0.78 (0.52–1.17), p=0.230<br>lag 5: 0.69 (0.47–1.02), p=0.065 |
| Cold stress wave intensity | lag 10: EDF=0.58, p=0.104<br>lag 6: EDF=0.00, p=0.430<br>**lag 5: EDF=0.84, p=0.009** | **lag 3: EDF=1.79, p=0.024**<br>lag 0: EDF=0.00, p=0.685 | lag 10: EDF=0.01, p=0.305<br>lag 10: EDF=0.00, p=0.734<br>lag 5: EDF=0.00, p=0.911 | lag 9: EDF=0.01, p=0.289<br>lag 0: EDF=0.00, p=0.522<br>lag 5: EDF=0.46, p=0.133 | lag 10: EDF=0.40, p=0.252<br>lag 8: EDF=0.00, p=0.996<br>lag 5: EDF=0.00, p=0.901 | lag 0: EDF=0.00, p=0.495<br>**lag 4: EDF=1.72, p=0.001**<br>lag 10: EDF=0.00, p=0.395 | lag 9: EDF=0.00, p=0.620<br>lag 8: EDF=0.00, p=0.453<br>lag 8: EDF=0.64, p=0.088 | **lag 8: EDF=0.80, p=0.036**<br>lag 0: EDF=0.00, p=0.414<br>lag 4: EDF=0.00, p=0.633 | lag 4: EDF=0.52, p=0.263<br>lag 9: EDF=0.80, p=0.092 | lag 2: EDF=0.00, p=0.650<br>lag 4: EDF=0.00, p=0.408 | lag 10: EDF=0.00, p=0.765<br>lag 0: EDF=0.05, p=0.430 |
| Ozone | lag 7: EDF=0.00, p=0.403<br>lag 7: EDF=0.63, p=0.083<br>lag 3: EDF=0.56, p=0.111<br>lag 2: EDF=0.01, p=0.303 | lag 4: EDF=0.00, p=0.433<br>lag 5: EDF=0.50, p=0.127<br>lag 6: EDF=0.38, p=0.174<br>lag 5: EDF=0.00, p=0.448 | lag 0: EDF=0.00, p=0.617<br>lag 1: EDF=0.00, p=0.782<br>**lag 2: 0.99 (0.99–1.00), p=<0.001** | lag 2: EDF=0.00, p=0.905<br>lag 0: EDF=0.51, p=0.122<br>lag 1: EDF=0.00, p=0.275<br>lag 2: EDF=0.00, p=0.475 | lag 0: EDF=1.78, p=0.092<br>lag 3: EDF=0.49, p=0.167<br>**lag 1: EDF=1.62, p=0.001**<br>lag 6: EDF=0.00, p=0.623 | lag 7: EDF=1.85, p=0.098<br>lag 0: EDF=0.00, p=0.734<br>lag 5: EDF=0.75, p=0.056<br>lag 6: EDF=0.00, p=0.382 | **lag 6: EDF=0.82, p=0.020**<br>lag 6: EDF=0.00, p=0.388<br>lag 4: EDF=0.57, p=0.104 | lag 1: EDF=0.00, p=0.746 | **lag 4: EDF=6.78, p=0.015**<br>**lag 0: EDF=3.40, p=0.007**<br>lag 2: EDF=0.00, p=0.707<br>lag 7: EDF=0.00, p=0.668 | lag 0: EDF=0.47, p=0.147<br>lag 0: EDF=0.66, p=0.070<br>**lag 6: EDF=0.73, p=0.031**<br>lag 6: EDF=0.15, p=0.243 | **lag 1: EDF=2.22, p=0.009**<br>**lag 3: EDF=0.86, p=0.021**<br>lag 1: EDF=0.00, p=0.800 |

| Atmospheric factor | 1.1 | 1.2 | 2.1 | 2.2 | 3.1 | 4.1 | 5.1 | 5.2 | 6.1 | 6.2 | 6.3 |
|---|---|---|---|---|---|---|---|---|---|---|---|
| Particulate matter ($PM_{2.5}$) | lag 5: EDF=0.00, p=0.443<br>lag 0: EDF=0.25, p=0.223<br>lag 1: EDF=0.00, p=0.710<br>**lag 7: EDF=0.75, p=0.039** | lag 4: EDF=0.00, p=0.439<br>lag 2: EDF=0.00, p=0.522<br>lag 5: EDF=0.00, p=0.375<br>**lag 4: EDF=0.90, p=<0.001** | lag 4: EDF=0.16, p=0.242<br>lag 2: EDF=0.61, p=0.084<br>lag 0: EDF=0.00, p=0.648<br>lag 7: EDF=0.70, p=0.052 | lag 0: EDF=0.00, p=0.843<br>lag 3: EDF=0.63, p=0.075<br>lag 3: EDF=0.00, p=0.450<br>**lag 6: EDF=0.75, p=0.032** | lag 5: EDF=0.00, p=0.982<br>**lag 4: EDF=0.78, p=0.036**<br>**lag 4: EDF=0.79, p=0.024**<br>lag 5: EDF=0.45, p=0.186 | lag 0: EDF=0.48, p=0.234<br>lag 7: EDF=0.73, p=0.066<br>lag 0: EDF=0.07, p=0.317<br>lag 1: EDF=0.00, p=0.497 | lag 0: EDF=0.64, p=0.089<br>lag 7: EDF=0.62, p=0.089<br>lag 0: EDF=2.76, p=0.057<br>lag 5: EDF=0.57, p=0.109 | **lag 5: EDF=0.87, p=0.010**<br>**lag 0: EDF=0.91, p=0.002**<br>lag 7: EDF=0.00, p=0.493<br>lag 2: EDF=0.00, p=0.529 | lag 4: EDF=0.00, p=0.639<br>lag 5: EDF=0.00, p=0.908<br>**lag 1: EDF=0.94, p=0.005**<br>lag 3: EDF=0.59, p=0.244 | **lag 6: EDF=0.90, p=0.001**<br>**lag 1: EDF=0.82, p=0.012**<br>lag 6: EDF=0.00, p=0.387<br>**lag 6: EDF=0.79, p=0.021** | lag 0: EDF=0.66, p=0.217<br>lag 0: EDF=0.00, p=0.795<br>**lag 0: EDF=6.28, p=<0.001**<br>lag 5: EDF=0.07, p=0.421 |

**Table S10.** Overview of time lags, effect sizes (effective degree of freedom (EDF) or relative risk (95% confidence interval)) and p-values per human-bioclimatic region, atmospheric factor and subgroup (pink = female <60 years, orange = females ≥60 years, green = males <60 years, blue = males ≥60 years) for heart failure, significant p-values (≤ 0.05) are bold

| Atmospheric factor | 1.1 | 1.2 | 2.1 | 2.2 | 3.1 | 4.1 | 5.1 | 5.2 | 6.1 | 6.2 | 6.3 |
|---|---|---|---|---|---|---|---|---|---|---|---|
| Sea-level pressure (SLP) | lag 2: EDF=0.64, p=0.177<br>**lag 2: EDF=0.95, p=<0.001**<br>**lag 0: EDF=1.82, p=0.039**<br>lag 10: EDF=0.00, p=0.490 | lag 6: EDF=0.00, p=0.418<br>lag 1: EDF=0.33, p=0.203<br>lag 7: EDF=0.72, p=0.051<br>**lag 0: EDF=0.73, p=0.048** | lag 1: EDF=0.76, p=0.080<br>lag 9: EDF=0.71, p=0.059<br>**lag 2: EDF=0.79, p=0.030**<br>lag 5: EDF=0.54, p=0.128 | **lag 0: EDF=0.89, p=0.012**<br>**lag 6: EDF=0.76, p=0.038**<br>**lag 4: EDF=0.87, p=0.006**<br>lag 0: EDF=0.00, p=0.746 | lag 8: EDF=0.78, p=0.205<br>**lag 0: EDF=0.86, p=0.006**<br>lag 4: EDF=0.71, p=0.144<br>lag 6: EDF=0.00, p=0.807 | **lag 4: EDF=3.74, p=0.008**<br>**lag 0: EDF=0.94, p=<0.001**<br>lag 4: EDF=0.77, p=0.075<br>**lag 0: EDF=0.74, p=0.039** | lag 10: EDF=0.81, p=0.084<br>**lag 3: EDF=0.93, p=<0.001**<br>lag 2: EDF=0.64, p=0.115<br>lag 9: EDF=0.30, p=0.210 | lag 1: EDF=0.72, p=0.219<br>lag 9: EDF=0.59, p=0.094<br>lag 9: EDF=0.60, p=0.187<br>**lag 7: EDF=0.74, p=0.036** | lag 2: EDF=0.86, p=0.213<br>**lag 1: EDF=0.95, p=<0.001**<br>**lag 8: EDF=2.55, p=0.029**<br>lag 10: EDF=0.00, p=0.454 | lag 7: EDF=0.37, p=0.263<br>**lag 3: EDF=0.82, p=0.016**<br>**lag 10: EDF=0.82, p=0.020**<br>lag 3: EDF=0.67, p=0.073 | lag 3: EDF=2.35, p=0.398<br>lag 0: EDF=0.24, p=0.225<br>**lag 1: EDF=5.80, p=0.001**<br>**lag 10: EDF=0.84, p=0.012** |
| Tropical night | lag 0: 1.47 (0.36–6.04), p=0.595<br>lag 4: 0.73 (0.45–1.21), | lag 3: 1.89 (0.96–3.70), p=0.064<br>lag 4: 0.87 (0.66–1.14), | lag 9: 2.74 (0.67–11.26), p=0.162<br>lag 6: 0.60 (0.30–1.23), | lag 2: 0.21 (0.03–1.57), p=0.128<br>lag 1: 0.87 (0.70–1.07), | - | lag 0: 0.00 (0.00–Inf), p=1.000<br>lag 1: 1.30 (0.48–3.53), p=0.606 | lag 5: 4.72 (0.63–35.48), p=0.131<br>**lag 1: 1.89 (1.04–3.43),** | lag 6: 5.84 (0.79–43.19), p=0.084<br>lag 5: 1.63 (0.79–3.32), | **lag 4: 7.15 (1.62–31.54), p=0.009**<br>lag 3: 1.71 (0.80–3.67), | lag 7: 1.25 (0.46–3.37), p=0.665<br>lag 6: 0.85 (0.64–1.12), | **lag 4: 6.25 (1.31–29.85), p=0.022**<br>**lag 1: 0.23 (0.06–0.95),** |

| Atmospheric factor | 1.1 | 1.2 | 2.1 | 2.2 | 3.1 | 4.1 | 5.1 | 5.2 | 6.1 | 6.2 | 6.3 |
|---|---|---|---|---|---|---|---|---|---|---|---|
| | p=0.224<br>lag 5: 1.64 (0.74–3.67), p=0.225<br>lag 3: 0.65 (0.38–1.12), p=0.124 | p=0.303<br>lag 10: 1.46 (0.93–2.29), p=0.099<br>**lag 3: 0.71 (0.53–0.96), p=0.028** | p=0.165<br>**lag 9: 3.70 (1.72–7.95), p=<0.001**<br>lag 6: 0.50 (0.22–1.15), p=0.101 | p=0.191<br>**lag 2: 1.73 (1.12–2.65), p=0.013**<br>lag 4: 0.78 (0.61–1.00), p=0.051 | | **lag 1: 5.48 (1.29–23.32), p=0.021**<br>lag 8: 1.76 (0.73–4.28), p=0.211 | **p=0.038**<br>**lag 3: 4.07 (1.28–12.96), p=0.018**<br>lag 1: 1.64 (0.80–3.36), p=0.178 | p=0.183<br>lag 8: 3.65 (0.91–14.62), p=0.067<br>lag 5: 1.09 (0.45–2.63), p=0.847 | p=0.170<br>lag 0: 1.94 (0.26–14.25), p=0.517<br>lag 1: 1.34 (0.50–3.63), p=0.564 | p=0.253<br>lag 6: 0.34 (0.11–1.12), p=0.076<br>lag 0: 0.74 (0.54–1.02), p=0.062 | **p=0.042**<br>lag 2: 1.81 (0.44–7.35), p=0.409<br>lag 10: 0.14 (0.02–1.01), p=0.051 |
| Summer day | lag 4: 0.85 (0.64–1.13), p=0.268<br>**lag 0: 0.90 (0.84–0.97), p=0.003**<br>**lag 1: 0.77 (0.66–0.91), p=0.002**<br>**lag 5: 0.89 (0.83–0.96), p=0.003** | lag 5: 0.90 (0.74–1.09), p=0.260<br>**lag 5: 0.89 (0.85–0.94), p=<0.001**<br>lag 3: 0.94 (0.83–1.06), p=0.290<br>**lag 0: 0.94 (0.89–1.00), p=0.048** | **lag 8: 0.76 (0.62–0.94), p=0.011**<br>lag 0: 0.97 (0.92–1.02), p=0.252<br>lag 4: 1.08 (0.97–1.21), p=0.154<br>**lag 1: 0.91 (0.86–0.95), p=<0.001** | lag 3: 0.96 (0.76–1.22), p=0.746<br>lag 0: 0.97 (0.92–1.02), p=0.252<br>**lag 6: 0.86 (0.76–0.97), p=0.017**<br>**lag 0: 0.90 (0.86–0.95), p=<0.001** | lag 5: 1.44 (0.96–2.16), p=0.081<br>lag 7: 0.93 (0.86–1.01), p=0.097<br>lag 9: 0.77 (0.58–1.02), p=0.069<br>**lag 6: 0.87 (0.79–0.95), p=0.003** | lag 9: 0.76 (0.41–1.42), p=0.396<br>lag 2: 0.95 (0.83–1.08), p=0.402<br>**lag 0: 0.65 (0.46–0.93), p=0.019**<br>**lag 6: 0.87 (0.77–1.00), p=0.043** | lag 6: 0.95 (0.64–1.40), p=0.780<br>**lag 0: 0.93 (0.88–1.00), p=0.039**<br>lag 10: 0.89 (0.74–1.08), p=0.238<br>**lag 5: 0.92 (0.86–0.99), p=0.029** | lag 8: 0.69 (0.45–1.04), p=0.079<br>lag 8: 0.94 (0.86–1.02), p=0.139<br>lag 5: 0.79 (0.59–1.05), p=0.107<br>**lag 0: 0.89 (0.81–0.98), p=0.015** | **lag 0: 1.88 (1.19–2.97), p=0.007**<br>**lag 6: 0.81 (0.71–0.92), p=0.002**<br>lag 5: 0.80 (0.53–1.20), p=0.275<br>**lag 0: 0.77 (0.66–0.90), p=0.001** | **lag 9: 0.82 (0.68–0.98), p=0.034**<br>lag 1: 0.97 (0.93–1.02), p=0.215<br>lag 4: 1.09 (0.96–1.24), p=0.173<br>lag 2: 0.97 (0.93–1.02), p=0.312 | lag 4: 0.62 (0.33–1.16), p=0.135<br>lag 3: 0.94 (0.84–1.04), p=0.225<br>**lag 1: 0.65 (0.48–0.89), p=0.006**<br>lag 7: 0.97 (0.86–1.09), p=0.565 |
| Wet-bulb globe temperature | **lag 1: EDF=0.92, p=0.010**<br>**lag 10: EDF=0.94, p=<0.001**<br>lag 1: EDF=0.00, p=0.862<br>lag 5: EDF=0.58, p=0.095 | lag 9: EDF=0.53, p=0.182<br>lag 0: EDF=0.00, p=0.849 | lag 9: EDF=0.35, p=0.281<br>lag 0: EDF=0.00, p=0.440 | **lag 5: EDF=2.43, p=<0.001** | lag 7: EDF=0.02, p=0.424<br>lag 8: EDF=0.00, p=0.830 | lag 6: EDF=0.00, p=0.379 | lag 0: EDF=0.00, p=0.661<br>**lag 5: 0.98 (0.98–0.99), p=<0.001** | lag 7: EDF=0.00, p=0.350 | lag 5: EDF=2.00, p=0.550 | lag 3: EDF=0.13, p=0.339<br>lag 0: EDF=0.00, p=0.415<br>**lag 3: EDF=2.56, p=0.002** | lag 6: EDF=3.55, p=0.094<br>lag 8: EDF=0.71, p=0.052<br>lag 6: EDF=0.00, p=0.974<br>lag 6: EDF=0.68, p=0.067 |

| **Atmospheric factor** | **1.1** | **1.2** | **2.1** | **2.2** | **3.1** | **4.1** | **5.1** | **5.2** | **6.1** | **6.2** | **6.3** |
|---|---|---|---|---|---|---|---|---|---|---|---|
| Daily mean temperature | lag 0: EDF=0.00, p=0.624 | lag 2: EDF=0.06, p=0.285<br>**lag 0: EDF=0.78, p=0.028**<br>lag 0: EDF=0.00, p=0.657 | lag 0: EDF=0.63, p=0.081<br>lag 8: EDF=0.00, p=0.409 | lag 2: EDF=2.60, p=0.062<br>**lag 0: EDF=1.08, p=<0.001**<br>lag 9: EDF=0.00, p=0.367 | **lag 2: EDF=1.10, p=0.004**<br>lag 8: EDF=0.03, p=0.278 | lag 4: EDF=1.96, p=0.315<br>**lag 0: EDF=0.92, p=<0.001**<br>lag 6: EDF=0.00, p=0.408 | **lag 6: EDF=1.07, p=<0.001**<br>lag 5: EDF=0.00, p=0.776<br>lag 3: EDF=0.44, p=0.150 | lag 5: EDF=0.89, p=0.066<br>lag 1: EDF=0.00, p=0.721<br>**lag 6: EDF=2.15, p=0.042**<br>lag 0: EDF=0.00, p=0.697 | lag 6: EDF=0.00, p=0.320<br>lag 4: EDF=0.00, p=0.579<br>lag 2: EDF=0.68, p=0.066 | **lag 4: EDF=0.87, p=0.006**<br>lag 1: EDF=0.00, p=0.677 | - |
| Heat wave duration | lag 2: EDF=0.79, p=0.066<br>lag 0: EDF=0.65, p=0.083<br>lag 0: EDF=0.00, p=0.415<br>**lag 0: EDF=0.80, p=0.022** | lag 8: EDF=0.56, p=0.154<br>**lag 0: EDF=0.87, p=0.004**<br>lag 1: EDF=0.18, p=0.253<br>**lag 0: EDF=0.83, p=0.015** | **lag 0: EDF=0.86, p=0.019**<br>lag 0: EDF=0.72, p=0.052<br>lag 7: EDF=0.69, p=0.078<br>**lag 1: EDF=0.85, p=0.009** | lag 2: EDF=0.68, p=0.136<br>**lag 0: EDF=0.86, p=0.006**<br>lag 9: EDF=0.39, p=0.199<br>lag 0: EDF=0.59, p=0.110 | lag 6: EDF=0.90, p=0.074<br>lag 0: EDF=0.71, p=0.050<br>**lag 1: EDF=0.89, p=0.015**<br>lag 0: EDF=0.55, p=0.116 | lag 2: EDF=0.18, p=0.468<br>**lag 10: EDF=0.76, p=0.026**<br>**lag 6: EDF=0.88, p=0.018**<br>lag 5: EDF=0.56, p=0.111 | lag 4: EDF=0.69, p=0.194<br>**lag 10: EDF=0.87, p=0.005**<br>**lag 4: EDF=0.86, p=0.009**<br>**lag 4: EDF=0.73, p=0.045** | lag 7: EDF=0.00, p=0.634<br>lag 10: EDF=0.00, p=0.382<br>lag 2: EDF=0.00, p=0.461<br>lag 0: EDF=0.69, p=0.057 | lag 0: EDF=0.00, p=0.741<br>**lag 0: EDF=0.85, p=0.008**<br>lag 2: EDF=0.82, p=0.142<br>**lag 0: EDF=0.74, p=0.048** | lag 9: EDF=0.29, p=0.302<br>**lag 0: EDF=0.83, p=0.013**<br>**lag 1: EDF=0.84, p=0.013**<br>**lag 0: EDF=0.92, p=<0.001** | lag 7: EDF=0.85, p=0.151<br>lag 5: EDF=0.00, p=0.332<br>lag 0: EDF=0.00, p=0.559<br>lag 7: EDF=0.63, p=0.093 |
| Cold stress wave day | lag 4: 0.95 (0.73–1.24), p=0.710<br>lag 4: 1.00 (0.94–1.06), p=0.947<br>lag 10: 1.09 (0.93–1.28), p=0.266<br>lag 5: 1.01 (0.95– | **lag 1: 1.37 (1.14–1.64), p=<0.001**<br>lag 8: 1.04 (0.98–1.09), p=0.178<br>lag 8: 0.89 (0.78–1.01), p=0.080<br>lag 7: 1.05 (0.99– | lag 2: 0.76 (0.58–1.00), p=0.053<br>**lag 8: 1.07 (1.01–1.12), p=0.012**<br>lag 8: 0.89 (0.77–1.04), p=0.134<br>lag 3: 1.02 (0.96– | lag 3: 1.28 (1.00–1.65), p=0.050<br>lag 0: 0.98 (0.93–1.04), p=0.505<br>lag 5: 1.06 (0.93–1.22), p=0.384<br>lag 7: 1.03 (0.97– | lag 3: 0.51 (0.20–1.34), p=0.171<br>lag 10: 1.06 (0.98–1.15), p=0.144<br>**lag 9: 1.54 (1.17–2.03), p=0.002**<br>lag 6: 1.07 (0.97– | lag 2: 1.15 (0.71–1.85), p=0.574<br>lag 2: 1.04 (0.95–1.13), p=0.398<br>lag 5: 0.88 (0.66–1.17), p=0.374<br>lag 4: 1.07 (0.97– | **lag 5: 0.44 (0.29–0.67), p=<0.001**<br>lag 5: 1.04 (0.98–1.10), p=0.169<br>lag 8: 1.08 (0.89–1.31), p=0.438<br>lag 4: 0.98 (0.92– | **lag 10: 0.49 (0.29–0.83), p=0.008**<br>lag 8: 1.06 (0.98–1.16), p=0.137<br>lag 10: 1.19 (0.93–1.51), p=0.169<br>lag 9: 1.07 (0.98– | lag 7: 1.41 (0.75–2.64), p=0.289<br>lag 3: 1.06 (0.95–1.18), p=0.274<br>lag 6: 0.75 (0.52–1.09), p=0.135<br>lag 1: 1.05 (0.92– | lag 8: 1.09 (0.87–1.37), p=0.431<br>lag 5: 1.04 (0.99–1.09), p=0.116<br>lag 9: 1.10 (0.95–1.27), p=0.212<br>lag 3: 1.05 (0.98– | lag 1: 0.62 (0.19–2.06), p=0.438<br>lag 2: 1.10 (0.98–1.23), p=0.116<br>**lag 7: 0.64 (0.45–0.92), p=0.015**<br>lag 0: 1.05 (0.91– |

| Atmospheric factor | 1.1 | 1.2 | 2.1 | 2.2 | 3.1 | 4.1 | 5.1 | 5.2 | 6.1 | 6.2 | 6.3 |
|---|---|---|---|---|---|---|---|---|---|---|---|
| | 1.08),<br>p=0.698 | 1.11),<br>p=0.076 | 1.09),<br>p=0.448 | 1.09),<br>p=0.358 | 1.17),<br>p=0.176 | 1.18),<br>p=0.193 | 1.05),<br>p=0.567 | 1.17),<br>p=0.119 | 1.21),<br>p=0.480 | 1.12),<br>p=0.188 | 1.20),<br>p=0.507 |
| Cold stress wave intensity | lag 10:<br>EDF=0.70,<br>p=0.150<br>lag 4:<br>EDF=0.00,<br>p=0.347<br>lag 3:<br>EDF=0.62,<br>p=0.112<br>lag 5:<br>EDF=0.00,<br>p=0.899 | lag 0:<br>EDF=0.00,<br>p=0.931<br>lag 6:<br>EDF=0.00,<br>p=0.443<br>lag 10:<br>EDF=0.00,<br>p=0.801<br>lag 0:<br>EDF=0.00,<br>p=0.805 | lag 1:<br>EDF=0.67,<br>p=0.117<br>lag 0:<br>EDF=0.00,<br>p=0.458<br>lag 8:<br>EDF=0.00,<br>p=0.676<br>lag 3:<br>EDF=0.67,<br>p=0.070 | lag 5:<br>EDF=0.00,<br>p=0.613<br>**lag 10:**<br>**EDF=0.82,**<br>**p=0.015**<br>lag 7:<br>EDF=0.00,<br>p=0.426<br>**lag 5:**<br>**EDF=0.85,**<br>**p=0.006** | lag 3:<br>EDF=2.21,<br>p=0.414<br>lag 0:<br>EDF=0.52,<br>p=0.114<br>**lag 7:**<br>**EDF=0.88,**<br>**p=0.029**<br>lag 9:<br>EDF=0.52,<br>p=0.112 | lag 10:<br>EDF=0.80,<br>p=0.152<br>lag 3:<br>EDF=0.00,<br>p=0.881<br>lag 7:<br>EDF=0.65,<br>p=0.141<br>**lag 1:**<br>**EDF=0.71,**<br>**p=0.046** | lag 3:<br>EDF=0.82,<br>p=0.062<br>lag 10:<br>EDF=0.08,<br>p=0.280<br>lag 10:<br>EDF=0.57,<br>p=0.134<br>lag 4:<br>EDF=0.00,<br>p=0.523 | **lag 10:**<br>**EDF=0.94,**<br>**p=0.005**<br>lag 10:<br>EDF=0.00,<br>p=0.411<br>lag 7:<br>EDF=0.00,<br>p=0.591<br>lag 10:<br>EDF=0.00,<br>p=0.462 | lag 8:<br>EDF=0.00,<br>p=0.710<br>lag 3:<br>EDF=0.00,<br>p=0.479<br>lag 0:<br>EDF=0.00,<br>p=0.892<br>**lag 9:**<br>**EDF=0.72,**<br>**p=0.048** | lag 9:<br>EDF=0.64,<br>p=0.131<br>lag 4:<br>EDF=0.00,<br>p=0.995<br>lag 0:<br>EDF=0.66,<br>p=0.075<br>lag 5:<br>EDF=0.00,<br>p=0.799 | lag 1:<br>EDF=3.01,<br>p=0.533<br>lag 2:<br>EDF=0.00,<br>p=0.791<br>lag 5:<br>EDF=0.00,<br>p=0.754<br>lag 1:<br>EDF=0.49,<br>p=0.138 |
| Particulate matter ($PM_{10}$) | lag 2:<br>EDF=0.78,<br>p=0.072<br>lag 0:<br>EDF=0.55,<br>p=0.116<br>lag 7:<br>EDF=0.67,<br>p=0.084<br>lag 0:<br>EDF=0.11,<br>p=0.263 | lag 0:<br>EDF=0.76,<br>p=0.064<br>lag 1:<br>EDF=0.62,<br>p=0.090<br>lag 6:<br>EDF=0.58,<br>p=0.105<br>lag 6:<br>EDF=0.41,<br>p=0.178 | lag 5:<br>EDF=0.65,<br>p=0.138<br>lag 1:<br>EDF=0.71,<br>p=0.055<br>lag 7:<br>EDF=0.00,<br>p=0.940<br>lag 6:<br>EDF=0.04,<br>p=0.298 | **lag 0:**<br>**EDF=0.82,**<br>**p=0.039**<br>lag 0:<br>EDF=0.45,<br>p=0.162<br>**lag 2:**<br>**EDF=0.84,**<br>**p=0.010**<br>lag 0:<br>EDF=0.00,<br>p=0.489 | **lag 7:**<br>**EDF=0.95,**<br>**p=0.015**<br>lag 6:<br>EDF=0.00,<br>p=0.296<br>lag 6:<br>EDF=2.28,<br>p=0.070<br>**lag 6:**<br>**EDF=0.72,**<br>**p=0.044** | lag 4:<br>EDF=0.84,<br>p=0.105<br>lag 3:<br>EDF=0.00,<br>p=0.706<br>lag 7:<br>EDF=0.74,<br>p=0.090<br>**lag 3:**<br>**EDF=0.71,**<br>**p=0.047** | lag 5:<br>EDF=0.82,<br>p=0.069<br>lag 0:<br>EDF=0.00,<br>p=0.516<br>lag 5:<br>EDF=0.00,<br>p=0.590<br>lag 5:<br>EDF=0.25,<br>p=0.224 | lag 0:<br>EDF=0.84,<br>p=0.092<br>lag 0:<br>EDF=0.68,<br>p=0.057<br>lag 3:<br>EDF=0.00,<br>p=0.418<br>lag 0:<br>EDF=0.00,<br>p=0.449 | lag 5:<br>EDF=2.05,<br>p=0.315<br>lag 1:<br>EDF=0.30,<br>p=0.210<br>**lag 1:**<br>**EDF=6.48,**<br>**p=0.011**<br>lag 3:<br>EDF=0.62,<br>p=0.092 | lag 0:<br>EDF=0.72,<br>p=0.094<br>lag 4:<br>EDF=0.62,<br>p=0.094<br>**lag 1:**<br>**EDF=0.75,**<br>**p=0.041**<br>lag 7:<br>EDF=0.11,<br>p=0.273 | **lag 2:**<br>**EDF=0.95,**<br>**p=0.039**<br>lag 7:<br>EDF=0.43,<br>p=0.164<br>lag 6:<br>EDF=0.74,<br>p=0.147<br>lag 7:<br>EDF=0.00,<br>p=0.874 |

**Table S11.** Overview of time lags, effect sizes (effective degree of freedom (EDF) or relative risk (95% confidence interval)) and p-values per human-bioclimatic region, atmospheric factor and subgroup (pink = female <60 years, orange = females ≥60 years, green = males <60 years, blue = males ≥60 years) for ischemic stroke, significant p-values (≤ 0.05) are bold

| Atmospheric factor | 1.1 | 1.2 | 2.1 | 2.2 | 3.1 | 4.1 | 5.1 | 5.2 | 6.1 | 6.2 | 6.3 |
|---|---|---|---|---|---|---|---|---|---|---|---|
| Sea-level pressure (SLP) | **lag 1:**<br>**EDF=0.86,**<br>**p=0.025**<br>lag 6: | lag 8:<br>EDF=0.67,<br>p=0.094<br>**lag 6:** | **lag 9:**<br>**EDF=0.85,**<br>**p=0.011**<br>**lag 10:** | lag 6:<br>EDF=0.00,<br>p=0.557<br>**lag 10:** | lag 6:<br>EDF=0.83,<br>p=0.054<br>lag 9: | **lag 0:**<br>**EDF=3.74,**<br>**p=0.012**<br>lag 9: | **lag 1:**<br>**EDF=0.88,**<br>**p=0.020**<br>**lag 3:** | lag 1:<br>EDF=0.80,<br>p=0.104<br>**lag 2:** | **lag 6:**<br>**EDF=5.94,**<br>**p=0.006**<br>**lag 8:** | lag 7:<br>EDF=0.62,<br>p=0.102<br>lag 4: | lag 2:<br>EDF=0.28,<br>p=0.419<br>lag 1: |

| Atmospheric factor | 1.1 | 1.2 | 2.1 | 2.2 | 3.1 | 4.1 | 5.1 | 5.2 | 6.1 | 6.2 | 6.3 |
|---|---|---|---|---|---|---|---|---|---|---|---|
| | EDF=0.54, p=0.116<br>lag 2: EDF=0.58, p=0.119<br>lag 1: EDF=0.68, p=0.063 | **EDF=0.80, p=0.021**<br>**lag 4: EDF=0.78, p=0.031**<br>**lag 4: EDF=3.27, p=0.010** | **EDF=0.84, p=0.013**<br>lag 1: EDF=0.01, p=0.319<br>lag 7: EDF=0.35, p=0.199 | **EDF=0.76, p=0.038**<br>**lag 2: EDF=0.89, p=0.003**<br>lag 7: EDF=0.00, p=0.343 | EDF=0.00, p=0.560<br>lag 1: EDF=0.37, p=0.241<br>**lag 6: EDF=0.85, p=0.008** | EDF=0.63, p=0.078<br>**lag 3: EDF=0.82, p=0.038**<br>**lag 2: EDF=0.72, p=0.044** | **EDF=0.78, p=0.026**<br>**lag 3: EDF=0.88, p=0.006**<br>**lag 3: EDF=0.91, p=0.001** | **EDF=0.88, p=0.003**<br>lag 0: EDF=0.23, p=0.321<br>**lag 0: EDF=0.77, p=0.026** | **EDF=0.78, p=0.035**<br>lag 10: EDF=0.71, p=0.237<br>lag 6: EDF=0.00, p=0.400 | EDF=0.74, p=0.050<br>lag 4: EDF=0.48, p=0.138<br>**lag 0: EDF=0.84, p=0.012** | EDF=0.00, p=0.451<br>lag 0: EDF=0.73, p=0.115<br>lag 10: EDF=0.00, p=0.370 |
| Wind day | **lag 9: 0.75 (0.57–1.00), p=0.047**<br>lag 1: 0.93 (0.85–1.01), p=0.087<br>**lag 4: 1.21 (1.04–1.41), p=0.015**<br>**lag 1: 0.89 (0.81–0.98), p=0.018** | **lag 6: 1.23 (1.03–1.47), p=0.026**<br>lag 5: 1.06 (0.98–1.14), p=0.159<br>**lag 6: 1.17 (1.02–1.35), p=0.026**<br>lag 9: 1.06 (0.97–1.15), p=0.181 | **lag 9: 1.23 (1.06–1.44), p=0.008**<br>lag 6: 1.04 (0.99–1.10), p=0.142<br>**lag 9: 1.17 (1.04–1.32), p=0.011**<br>**lag 5: 0.93 (0.87–0.99), p=0.018** | lag 0: 1.09 (0.92–1.30), p=0.305<br>lag 7: 1.01 (0.95–1.07), p=0.694<br>lag 0: 0.88 (0.76–1.01), p=0.060<br>lag 5: 0.94 (0.89–1.01), p=0.088 | lag 9: 1.28 (0.99–1.67), p=0.060<br>lag 4: 1.07 (0.98–1.18), p=0.152<br>lag 1: 0.83 (0.65–1.05), p=0.113<br>**lag 5: 1.12 (1.01–1.23), p=0.027** | lag 10: 1.09 (0.81–1.47), p=0.575<br>**lag 8: 1.11 (1.02–1.22), p=0.023**<br>lag 10: 1.22 (0.99–1.50), p=0.060<br>**lag 8: 0.88 (0.79–0.99), p=0.032** | **lag 1: 1.42 (1.12–1.79), p=0.003**<br>**lag 7: 0.90 (0.83–0.98), p=0.017**<br>lag 1: 1.05 (0.89–1.24), p=0.573<br>lag 10: 1.07 (0.99–1.16), p=0.090 | **lag 2: 0.58 (0.38–0.90), p=0.016**<br>lag 8: 1.10 (0.99–1.22), p=0.088<br>**lag 10: 1.35 (1.08–1.69), p=0.008**<br>lag 0: 1.12 (1.00–1.26), p=0.056 | lag 10: 1.54 (0.89–2.67), p=0.120<br>lag 7: 1.11 (0.94–1.32), p=0.227<br>**lag 8: 1.69 (1.19–2.40), p=0.003**<br>**lag 9: 0.76 (0.59–0.96), p=0.023** | **lag 8: 1.26 (1.09–1.47), p=0.002**<br>lag 2: 1.05 (1.00–1.10), p=0.064<br>**lag 1: 0.84 (0.74–0.96), p=0.008**<br>lag 0: 0.98 (0.92–1.03), p=0.414 | **lag 3: 1.49 (1.08–2.06), p=0.015**<br>lag 5: 1.07 (0.95–1.22), p=0.272<br>**lag 4: 1.35 (1.05–1.72), p=0.019**<br>**lag 8: 1.22 (1.07–1.39), p=0.003** |
| Tropical night | lag 1: 1.45 (0.44–4.77), p=0.540<br>**lag 5: 0.51 (0.27–0.97), p=0.040**<br>lag 6: 1.33 (0.59–3.01), p=0.495<br>**lag 0: 0.30 (0.12–** | **lag 2: 2.13 (1.27–3.59), p=0.004**<br>lag 10: 0.74 (0.53–1.03), p=0.070<br>lag 10: 1.46 (0.95–2.24), p=0.084<br>lag 4: 1.27 (0.98– | **lag 4: 2.53 (1.03–6.18), p=0.042**<br>**lag 9: 0.37 (0.16–0.83), p=0.016**<br>**lag 8: 2.41 (1.23–4.73), p=0.011**<br>lag 10: 1.36 (0.89– | lag 0: 1.16 (0.69–1.93), p=0.580<br>lag 0: 1.17 (0.98–1.41), p=0.085<br>lag 7: 1.31 (0.92–1.85), p=0.130<br>lag 1: 1.16 (0.95– | - | lag 0: 3.24 (0.75–14.03), p=0.116<br>lag 7: 1.40 (0.56–3.55), p=0.474<br>lag 3: 2.74 (0.67–11.19), p=0.159<br>lag 3: 1.34 (0.43– | **lag 0: 5.64 (1.75–18.17), p=0.004**<br>lag 6: 1.71 (0.90–3.26), p=0.101<br>**lag 2: 3.36 (1.22–9.26), p=0.019**<br>lag 4: 1.59 (0.79– | lag 1: 3.33 (0.79–14.01), p=0.101<br>**lag 4: 2.21 (1.18–4.14), p=0.013**<br>lag 2: 2.38 (0.71–7.95), p=0.159<br>**lag 7: 2.27 (1.07–** | lag 0: 1.61 (0.17–15.22), p=0.678<br>lag 2: 2.17 (0.94–4.97), p=0.068<br>**lag 6: 4.21 (1.02–17.30), p=0.046**<br>lag 10: 2.24 (0.89– | **lag 8: 1.72 (1.00–2.97), p=0.050**<br>lag 9: 0.82 (0.65–1.04), p=0.099<br>lag 1: 1.33 (0.92–1.94), p=0.135<br>**lag 1: 0.71 (0.55–** | lag 9: 2.70 (0.84–8.60), p=0.094<br>lag 1: 1.50 (0.87–2.57), p=0.142<br>**lag 0: 2.37 (1.02–5.49), p=0.044**<br>**lag 8: 1.95 (1.06–** |

| Atmospheric factor | 1.1 | 1.2 | 2.1 | 2.2 | 3.1 | 4.1 | 5.1 | 5.2 | 6.1 | 6.2 | 6.3 |
|---|---|---|---|---|---|---|---|---|---|---|---|
| | **0.74), p=0.009** | 1.65), p=0.074 | 2.08), p=0.155 | 1.41), p=0.144 | | 4.15), p=0.615 | 3.19), p=0.191 | **4.82), p=0.034** | 5.59), p=0.085 | **0.93), p=0.012** | **3.60), p=0.032** |
| Hot day | **lag 10: 1.68 (1.05–2.68), p=0.029** | lag 10: 0.74 (0.50–1.08), p=0.114 | lag 10: 0.80 (0.59–1.09), p=0.160 | **lag 0: 1.30 (1.04–1.63), p=0.021** | lag 7: 1.58 (0.90–2.76), p=0.113 | **lag 1: 2.96 (1.26–6.92), p=0.012** | lag 5: 0.91 (0.55–1.52), p=0.726 | lag 0: 1.26 (0.74–2.15), p=0.397 | lag 2: 1.01 (0.32–3.16), p=0.988 | lag 3: 1.12 (0.88–1.42), p=0.366 | lag 0: 1.32 (0.75–2.29), p=0.333 |
| | **lag 6: 1.29 (1.11–1.50), p=<0.001** | lag 3: 1.08 (0.95–1.22), p=0.234 | lag 7: 0.93 (0.86–1.02), p=0.118 | lag 8: 0.94 (0.86–1.02), p=0.132 | **lag 6: 0.77 (0.61–0.99), p=0.039** | lag 3: 1.20 (0.79–1.84), p=0.390 | lag 1: 0.89 (0.75–1.05), p=0.162 | lag 4: 1.17 (0.97–1.42), p=0.093 | **lag 0: 1.43 (1.03–1.99), p=0.032** | lag 9: 0.96 (0.88–1.04), p=0.296 | **lag 0: 0.78 (0.62–0.99), p=0.038** |
| | lag 0: 1.02 (0.76–1.38), p=0.882 | lag 0: 1.00 (0.79–1.27), p=0.980 | lag 4: 1.16 (0.97–1.39), p=0.100 | lag 2: 1.13 (0.97–1.33), p=0.122 | **lag 7: 0.53 (0.29–0.98), p=0.043** | lag 10: 1.14 (0.47–2.82), p=0.768 | lag 0: 1.22 (0.88–1.69), p=0.230 | lag 4: 1.24 (0.80–1.92), p=0.338 | lag 7: 1.18 (0.49–2.87), p=0.715 | **lag 4: 1.26 (1.08–1.46), p=0.004** | lag 9: 1.15 (0.72–1.83), p=0.571 |
| | **lag 10: 1.27 (1.07–1.50), p=0.005** | **lag 6: 1.15 (1.02–1.30), p=0.021** | **lag 7: 1.11 (1.02–1.21), p=0.019** | lag 3: 0.94 (0.85–1.03), p=0.168 | lag 9: 0.84 (0.65–1.07), p=0.163 | lag 1: 1.48 (0.96–2.27), p=0.073 | lag 8: 0.86 (0.71–1.03), p=0.100 | lag 5: 1.13 (0.92–1.39), p=0.260 | lag 5: 1.35 (0.88–2.07), p=0.163 | lag 6: 1.06 (0.98–1.16), p=0.135 | lag 0: 1.06 (0.85–1.32), p=0.633 |
| Summer day | lag 6: 0.86 (0.68–1.08), p=0.190 | **lag 1: 0.83 (0.71–0.98), p=0.026** | **lag 10: 1.17 (1.03–1.33), p=0.015** | lag 8: 1.09 (0.96–1.23), p=0.202 | **lag 7: 1.31 (1.00–1.71), p=0.047** | **lag 3: 1.87 (1.36–2.57), p=<0.001** | lag 4: 1.21 (0.97–1.50), p=0.085 | **lag 0: 1.45 (1.10–1.92), p=0.009** | lag 1: 1.02 (0.52–2.02), p=0.946 | lag 9: 0.92 (0.82–1.04), p=0.186 | lag 6: 1.09 (0.82–1.45), p=0.541 |
| | **lag 5: 0.90 (0.84–0.97), p=0.008** | lag 8: 1.05 (0.99–1.11), p=0.129 | lag 2: 0.98 (0.94–1.02), p=0.301 | lag 4: 0.96 (0.92–1.00), p=0.075 | **lag 3: 0.91 (0.84–0.99), p=0.031** | lag 5: 1.09 (0.97–1.21), p=0.143 | lag 7: 1.05 (0.98–1.12), p=0.163 | lag 7: 1.03 (0.94–1.12), p=0.595 | lag 10: 0.92 (0.79–1.07), p=0.285 | lag 7: 0.99 (0.96–1.03), p=0.601 | **lag 10: 0.88 (0.80–0.98), p=0.020** |
| | lag 9: 1.12 (0.98–1.28), p=0.101 | lag 0: 1.08 (0.95–1.22), p=0.235 | lag 5: 1.00 (0.91–1.10), p=0.997 | lag 5: 1.04 (0.96–1.14), p=0.328 | lag 4: 1.09 (0.91–1.30), p=0.360 | lag 9: 1.13 (0.89–1.45), p=0.316 | lag 1: 1.14 (0.98–1.32), p=0.095 | lag 4: 1.07 (0.85–1.34), p=0.571 | lag 10: 0.97 (0.67–1.41), p=0.894 | lag 10: 1.08 (1.00–1.17), p=0.054 | **lag 5: 1.26 (1.03–1.53), p=0.025** |
| | lag 0: 1.05 (0.97–1.13), p=0.213 | lag 1: 1.06 (1.00–1.13), p=0.052 | **lag 0: 1.05 (1.00–1.09), p=0.033** | lag 10: 1.00 (0.96–1.05), p=0.864 | **lag 5: 0.87 (0.79–0.95), p=0.002** | lag 9: 0.89 (0.78–1.03), p=0.114 | lag 9: 0.94 (0.87–1.01), p=0.109 | lag 2: 0.90 (0.82–1.00), p=0.056 | lag 2: 0.88 (0.73–1.06), p=0.178 | **lag 5: 1.05 (1.01–1.09), p=0.022** | lag 6: 1.06 (0.95–1.18), p=0.281 |
| Daily mean temperature | lag 4: EDF=0.00, p=0.937 | lag 10: EDF=0.63, p=0.111 | **lag 4: EDF=0.81, p=0.015** | **lag 5: EDF=0.76, p=0.044** | **lag 10: EDF=0.94, p=0.009** | lag 0: EDF=0.70, p=0.147 | lag 1: EDF=0.00, p=0.588 | **lag 4: EDF=0.94, p=0.011** | lag 0: EDF=0.40, p=0.527 | lag 1: EDF=0.00, p=0.433 | lag 5: EDF=0.59, p=0.103 |
| | lag 3: | **lag 6:** | **lag 7:** | **lag 0:** | lag 3: | lag 2: | lag 1: | lag 2: | lag 2: | **lag 5:** | lag 0: |

| Atmospheric factor | 1.1 | 1.2 | 2.1 | 2.2 | 3.1 | 4.1 | 5.1 | 5.2 | 6.1 | 6.2 | 6.3 |
|---|---|---|---|---|---|---|---|---|---|---|---|
| | EDF=0.01, p=0.308<br>lag 1: EDF=0.31, p=0.206<br>lag 10: EDF=0.00, p=0.742 | **EDF=0.84, p=0.010**<br>lag 0: EDF=0.66, p=0.080<br>lag 0: EDF=0.00, p=0.916 | **EDF=0.92, p=<0.001** | **EDF=0.72, p=0.030**<br>lag 10: EDF=0.04, p=0.268<br>**lag 3: EDF=0.78, p=0.028** | EDF=0.00, p=0.461<br>lag 10: EDF=0.00, p=0.846 | EDF=0.00, p=0.334<br>lag 10: EDF=0.00, p=0.582<br>lag 7: EDF=0.57, p=0.092 | EDF=0.00, p=0.359<br>lag 9: EDF=0.00, p=0.830<br>lag 5: EDF=0.01, p=0.299 | EDF=0.00, p=0.496 | EDF=0.00, p=0.540<br>lag 8: EDF=0.00, p=0.903<br>lag 1: EDF=0.00, p=0.943 | **EDF=0.86, p=0.008**<br>**lag 10: EDF=0.91, p=0.002** | EDF=0.00, p=0.521<br>lag 0: EDF=0.00, p=0.403 |
| Heat stress wave intensity | lag 9: EDF=0.74, p=0.096<br>lag 9: EDF=0.00, p=0.306<br>lag 6: EDF=0.00, p=0.846<br>**lag 8: EDF=0.83, p=0.011** | lag 10: EDF=0.59, p=0.140<br>lag 3: EDF=0.50, p=0.131<br>lag 10: EDF=0.00, p=0.674<br>lag 0: EDF=0.00, p=0.672 | lag 8: EDF=0.74, p=0.051<br>lag 9: EDF=0.71, p=0.061<br>lag 10: EDF=0.47, p=0.134<br>**lag 9: EDF=0.80, p=0.021** | lag 9: EDF=0.22, p=0.257<br>lag 9: EDF=0.57, p=0.112<br>lag 1: EDF=0.00, p=0.467<br>lag 7: EDF=0.00, p=0.810 | lag 3: EDF=0.77, p=0.107<br>lag 3: EDF=0.00, p=0.412<br>lag 7: EDF=0.00, p=0.596<br>**lag 10: EDF=0.79, p=0.023** | **lag 9: EDF=0.91, p=0.013**<br>lag 7: EDF=0.00, p=0.476<br>lag 9: EDF=0.00, p=0.390<br>lag 5: EDF=0.57, p=0.109 | lag 5: EDF=2.32, p=0.056<br>lag 7: EDF=0.00, p=0.527<br>lag 0: EDF=0.00, p=0.823<br>lag 1: EDF=0.17, p=0.242 | lag 0: EDF=0.00, p=0.787<br>lag 7: EDF=0.29, p=0.199<br>lag 0: EDF=0.52, p=0.197<br>lag 3: EDF=0.08, p=0.262 | lag 0: EDF=5.04, p=0.330<br>lag 1: EDF=0.63, p=0.114<br>lag 6: EDF=0.62, p=0.282<br>**lag 4: EDF=0.80, p=0.040** | lag 2: EDF=0.00, p=0.730<br>**lag 1: EDF=0.89, p=0.002**<br>lag 3: EDF=0.00, p=0.324<br>lag 4: EDF=0.00, p=0.556 | lag 7: EDF=0.00, p=0.929<br>lag 1: EDF=0.00, p=0.510<br>lag 9: EDF=2.14, p=0.114<br>**lag 7: EDF=0.86, p=0.007** |
| Cold wave intensity | lag 9: EDF=0.69, p=0.057<br>lag 9: EDF=0.00, p=0.917 | lag 3: EDF=0.00, p=0.630<br>lag 0: EDF=0.00, p=0.650 | lag 8: EDF=0.35, p=0.198<br>lag 10: EDF=0.00, p=0.541 | lag 0: EDF=0.64, p=0.083<br>lag 6: EDF=0.00, p=0.602<br>lag 1: EDF=0.58, p=0.114 | lag 5: EDF=0.04, p=0.364 | **lag 0: EDF=0.88, p=0.032**<br>lag 1: EDF=0.00, p=0.476<br>lag 0: EDF=0.00, p=0.413<br>lag 0: EDF=0.23, p=0.208 | lag 10: EDF=0.71, p=0.114<br>lag 4: EDF=0.46, p=0.143<br>**lag 6: EDF=0.72, p=0.041** | lag 8: EDF=2.96, p=0.331<br>lag 10: EDF=0.00, p=0.570<br>lag 8: EDF=0.00, p=0.239 | lag 2: EDF=5.72, p=0.093<br>lag 0: EDF=0.65, p=0.101 | **lag 0: EDF=0.81, p=0.018**<br>**lag 0: EDF=0.85, p=0.005**<br>**lag 10: EDF=0.79, p=0.022** | **lag 0: EDF=0.93, p=0.018**<br>lag 0: EDF=0.00, p=0.365<br>**lag 7: EDF=0.87, p=0.014**<br>lag 0: EDF=0.00, p=0.394 |
| Cold stress wave duration | lag 0: EDF=0.45, p=0.250<br>lag 0: EDF=0.01, p=0.266 | lag 6: EDF=0.00, p=0.496<br>lag 10: EDF=0.00, p=0.389 | lag 10: EDF=0.00, p=0.495<br>lag 7: EDF=0.00, p=0.485 | lag 9: EDF=0.57, p=0.114<br>lag 8: EDF=0.00, p=0.675 | lag 8: EDF=0.73, p=0.095<br>lag 8: EDF=0.00, p=0.307 | lag 5: EDF=0.00, p=0.347<br>lag 10: EDF=0.00, p=0.701 | lag 9: EDF=0.00, p=0.860<br>lag 0: EDF=0.43, p=0.157 | **lag 5: EDF=0.86, p=0.049**<br>**lag 7: EDF=0.83, p=0.040** | lag 7: EDF=0.00, p=0.679<br>lag 9: EDF=0.34, p=0.215 | lag 5: EDF=0.72, p=0.060<br>lag 8: EDF=0.00, p=0.320 | lag 6: EDF=0.06, p=0.275<br>lag 10: EDF=0.66, p=0.155 |

| Atmospheric factor | 1.1 | 1.2 | 2.1 | 2.2 | 3.1 | 4.1 | 5.1 | 5.2 | 6.1 | 6.2 | 6.3 |
|---|---|---|---|---|---|---|---|---|---|---|---|
| | lag 3: EDF=0.00, p=0.529<br>lag 1: EDF=0.00, p=0.882 | lag 0: EDF=0.00, p=0.819 | lag 0: EDF=0.02, p=0.291 | | **lag 0: EDF=0.93, p=<0.001**<br>lag 0: EDF=0.00, p=0.893 | **lag 3: EDF=0.90, p=<0.001** | lag 0: EDF=0.00, p=0.465 | lag 1: EDF=0.00, p=0.352 | lag 9: EDF=0.00, p=0.388 | | lag 10: EDF=0.00, p=0.811 |
| Cold stress wave intensity | lag 0: EDF=0.00, p=0.677 | - | lag 4: EDF=0.62, p=0.116<br>**lag 1: EDF=0.79, p=0.017** | lag 0: EDF=0.00, p=0.342 | lag 6: EDF=0.73, p=0.121<br>lag 6: EDF=0.00, p=0.579<br>lag 6: EDF=0.00, p=0.904 | lag 9: EDF=0.00, p=0.400 | lag 2: EDF=0.00, p=0.718 | **lag 0: EDF=0.89, p=0.021**<br>lag 7: EDF=0.00, p=0.370<br>lag 10: EDF=0.52, p=0.130 | lag 6: EDF=2.33, p=0.094 | lag 1: EDF=0.00, p=0.854 | - |
| Ozone | **lag 4: EDF=3.04, p=0.001**<br>lag 1: EDF=0.00, p=0.716<br>**lag 4: EDF=0.76, p=0.033**<br>lag 3: EDF=0.00, p=0.636 | lag 2: EDF=0.00, p=0.527<br>lag 3: EDF=0.00, p=0.738<br>lag 4: EDF=0.00, p=0.932<br>lag 6: EDF=0.00, p=0.716 | **lag 7: EDF=2.22, p=0.007**<br>lag 1: EDF=0.00, p=0.327<br>**lag 5: EDF=0.74, p=0.040** | lag 3: EDF=0.38, p=0.175<br>lag 1: EDF=0.00, p=0.907<br>lag 0: EDF=0.00, p=0.527 | lag 6: EDF=0.00, p=0.831<br>lag 6: EDF=0.00, p=0.545<br>**lag 4: EDF=0.72, p=0.047**<br>**lag 6: EDF=0.76, p=0.033** | lag 1: EDF=0.35, p=0.326<br>lag 0: EDF=0.00, p=0.608<br>lag 4: EDF=0.00, p=0.425<br>lag 4: EDF=0.58, p=0.088 | lag 2: EDF=0.30, p=0.202<br>lag 0: EDF=0.00, p=0.495<br>lag 3: EDF=0.00, p=0.475 | **lag 3: EDF=0.74, p=0.037**<br>lag 4: EDF=0.37, p=0.268<br>lag 0: EDF=0.29, p=0.151 | lag 2: EDF=0.73, p=0.356<br>lag 6: EDF=0.00, p=0.465<br>lag 3: EDF=0.02, p=0.514<br>**lag 4: EDF=0.88, p=0.010** | **lag 2: EDF=0.81, p=0.016**<br>lag 2: EDF=0.41, p=0.161 | lag 0: EDF=0.08, p=0.473<br>lag 4: EDF=0.00, p=0.812<br>lag 1: EDF=0.54, p=0.225<br>lag 4: EDF=0.00, p=0.630 |
| Particulate matter ($PM_{10}$) | **lag 5: EDF=0.88, p=0.011**<br>lag 7: EDF=0.28, p=0.207<br>lag 2: EDF=0.21, p=0.257<br>lag 6: | lag 0: EDF=0.71, p=0.073<br>lag 2: EDF=0.30, p=0.210<br>**lag 1: EDF=0.78, p=0.029**<br>lag 5: | **lag 3: EDF=0.92, p=<0.001**<br>lag 7: EDF=0.51, p=0.144<br>lag 1: EDF=0.00, p=0.436<br>lag 0: | lag 0: EDF=0.00, p=0.474<br>lag 3: EDF=0.57, p=0.116<br>lag 5: EDF=0.00, p=0.606<br>lag 3: | **lag 3: EDF=0.87, p=0.026**<br>lag 2: EDF=0.00, p=0.351<br>lag 7: EDF=0.78, p=0.051<br>lag 1: | lag 0: EDF=0.14, p=0.406<br>lag 5: EDF=0.00, p=0.442<br>lag 4: EDF=0.66, p=0.119<br>lag 4: | **lag 7: EDF=3.91, p=0.021**<br>lag 6: EDF=0.26, p=0.216<br>lag 4: EDF=0.61, p=0.107<br>lag 3: | **lag 6: EDF=4.24, p=0.004**<br>**lag 4: EDF=0.86, p=0.005**<br>lag 3: EDF=0.24, p=0.311<br>lag 3: | **lag 7: 0.97 (0.94–0.99), p=0.004**<br>lag 2: EDF=0.42, p=0.193<br>lag 2: EDF=4.24, p=0.201<br>lag 2: | lag 2: EDF=0.66, p=0.080<br>lag 6: EDF=0.00, p=0.382<br>lag 4: EDF=0.00, p=0.960<br>lag 1: | **lag 0: EDF=0.92, p=0.023**<br>lag 4: EDF=0.00, p=0.415<br>lag 2: EDF=0.59, p=0.201<br>lag 1: |

| Atmospheric factor | 1.1 | 1.2 | 2.1 | 2.2 | 3.1 | 4.1 | 5.1 | 5.2 | 6.1 | 6.2 | 6.3 |
|---|---|---|---|---|---|---|---|---|---|---|---|
| | EDF=0.00, p=0.381 | EDF=0.48, p=0.146 | EDF=0.48, p=0.147 | EDF=0.00, p=0.480 | EDF=0.20, p=0.225 | EDF=0.69, p=0.056 | EDF=0.43, p=0.160 | EDF=0.63, p=0.077 | EDF=0.26, p=0.273 | EDF=0.00, p=0.449 | EDF=0.01, p=0.315 |

**Table S12.** Overview of time lags, effect sizes (effective degree of freedom (EDF) or relative risk (95% confidence interval)) and p-values per human-bioclimatic region, atmospheric factor and subgroup (pink = female <60 years, orange = females ≥60 years, green = males <60 years, blue = males ≥60 years) for ischemic stroke (stroke unit), significant p-values (≤ 0.05) are bold

| Atmospheric factor | 1.1 | 1.2 | 2.1 | 2.2 | 3.1 | 4.1 | 5.1 | 5.2 | 6.1 | 6.2 | 6.3 |
|---|---|---|---|---|---|---|---|---|---|---|---|
| Sea-level pressure difference (SLPD) | lag 6: 1.07 (0.96–1.19), p=0.234<br>lag 5: 1.03 (1.00–1.07), p=0.083<br>**lag 8: 0.91 (0.85–0.98), p=0.019**<br>**lag 7: 1.04 (1.00–1.08), p=0.050** | **lag 4: 1.12 (1.02–1.22), p=0.014**<br>lag 10: 0.98 (0.94–1.01), p=0.184<br>lag 10: 0.96 (0.90–1.02), p=0.185<br>lag 3: 1.03 (0.99–1.07), p=0.122 | **lag 0: 1.14 (1.01–1.28), p=0.036**<br>**lag 10: 1.05 (1.01–1.09), p=0.014**<br>**lag 7: 0.87 (0.79–0.94), p=0.001**<br>lag 8: 0.96 (0.92–1.00), p=0.055 | lag 9: 1.09 (0.97–1.23), p=0.168<br>lag 5: 1.02 (0.98–1.06), p=0.248<br>lag 5: 0.94 (0.86–1.03), p=0.168<br>lag 2: 0.97 (0.93–1.01), p=0.100 | **lag 5: 1.27 (1.02–1.57), p=0.030**<br>**lag 4: 1.09 (1.01–1.17), p=0.021**<br>lag 6: 1.09 (0.93–1.29), p=0.291<br>**lag 9: 0.91 (0.84–0.98), p=0.012** | lag 4: 1.12 (0.96–1.31), p=0.137<br>lag 4: 0.97 (0.92–1.02), p=0.238<br>lag 0: 1.10 (0.99–1.23), p=0.084<br>lag 0: 1.04 (0.98–1.10), p=0.170 | lag 10: 1.08 (0.94–1.26), p=0.279<br>**lag 0: 0.94 (0.89–0.98), p=0.006**<br>lag 6: 1.10 (1.00–1.22), p=0.056<br>lag 10: 0.96 (0.92–1.01), p=0.115 | lag 8: 1.14 (0.94–1.38), p=0.182<br>lag 4: 1.05 (0.99–1.12), p=0.119<br>lag 10: 0.89 (0.77–1.03), p=0.123<br>lag 6: 1.05 (0.98–1.12), p=0.161 | lag 7: 1.26 (0.93–1.71), p=0.135<br>lag 5: 1.06 (0.97–1.16), p=0.173<br>lag 2: 1.22 (0.99–1.50), p=0.059<br>lag 0: 0.93 (0.84–1.02), p=0.119 | **lag 10: 0.86 (0.78–0.96), p=0.007**<br>**lag 5: 1.04 (1.01–1.07), p=0.014**<br>**lag 4: 0.93 (0.86–1.00), p=0.041**<br>**lag 3: 1.04 (1.00–1.08), p=0.031** | lag 10: 1.16 (0.93–1.46), p=0.191<br>lag 2: 1.07 (1.00–1.16), p=0.062<br>lag 7: 1.11 (0.95–1.30), p=0.193<br>**lag 1: 0.88 (0.81–0.95), p=0.001** |
| Tropical night | lag 10: 1.38 (0.71–2.70), p=0.344<br>lag 1: 0.74 (0.54–1.01), p=0.057<br>lag 9: 1.21 (0.76–1.92), p=0.413<br>lag 7: 0.86 | lag 9: 1.27 (0.88–1.84), p=0.206<br>**lag 9: 0.81 (0.67–0.98), p=0.028**<br>**lag 8: 1.39 (1.08–1.78), p=0.010**<br>lag 2: 1.16 | lag 5: 1.46 (0.60–3.56), p=0.401<br>**lag 9: 0.52 (0.31–0.87), p=0.013**<br>lag 2: 1.72 (0.97–3.05), p=0.063<br>lag 9: 1.24 | lag 4: 1.30 (0.87–1.93), p=0.198<br>lag 3: 1.13 (0.98–1.29), p=0.082<br>lag 4: 1.14 (0.87–1.51), p=0.344<br>lag 4: 1.07 | - | lag 8: 2.59 (0.64–10.42), p=0.180<br>**lag 7: 1.94 (1.09–3.45), p=0.024**<br>lag 7: 1.49 (0.37–6.01), p=0.573<br>lag 9: 1.82 | lag 5: 1.63 (0.40–6.62), p=0.493<br>**lag 1: 0.21 (0.05–0.86), p=0.030**<br>lag 3: 1.77 (0.67–4.70), p=0.249<br>**lag 4: 1.70** | **lag 1: 5.54 (2.32–13.22), p=<0.001**<br>**lag 9: 1.68 (1.03–2.75), p=0.038**<br>**lag 6: 4.02 (1.89–8.59), p=<0.001**<br>lag 4: 1.70 | lag 5: 2.50 (0.60–10.41), p=0.208<br>lag 10: 1.62 (0.95–2.77), p=0.079<br>lag 6: 2.08 (0.51–8.47), p=0.307<br>lag 5: 1.57 | lag 4: 1.21 (0.77–1.90), p=0.401<br>lag 5: 0.89 (0.74–1.06), p=0.189<br>lag 1: 1.18 (0.86–1.62), p=0.295<br>lag 8: 1.10 | lag 3: 1.66 (0.38–7.16), p=0.499<br>lag 3: 1.10 (0.68–1.76), p=0.705<br>**lag 7: 2.46 (1.26–4.82), p=0.009**<br>**lag 9: 0.41** |

| Atmospheric factor | 1.1 | 1.2 | 2.1 | 2.2 | 3.1 | 4.1 | 5.1 | 5.2 | 6.1 | 6.2 | 6.3 |
|---|---|---|---|---|---|---|---|---|---|---|---|
| | (0.62–1.19), p=0.359 | (0.99–1.36), p=0.075 | (0.89–1.73), p=0.207 | (0.92–1.24), p=0.382 | | (0.98–3.36), p=0.057 | **(1.02–2.82), p=0.040** | (0.97–3.00), p=0.066 | (0.81–3.04), p=0.182 | (0.93–1.29), p=0.268 | **(0.18–0.92), p=0.030** |
| Wet-bulb globe temperature | lag 10: EDF=0.00, p=0.833<br>lag 7: EDF=0.00, p=0.430<br>**lag 0: EDF=0.64, p=0.042**<br>lag 0: EDF=0.52, p=0.119 | lag 9: EDF=0.67, p=0.059<br>lag 10: EDF=0.00, p=0.784<br>lag 0: EDF=0.66, p=0.069<br>lag 0: EDF=0.00, p=0.591 | lag 5: EDF=0.09, p=0.267<br>lag 5: EDF=0.00, p=0.823<br>**lag 4: EDF=0.88, p=0.004**<br>lag 0: EDF=0.14, p=0.272 | lag 10: EDF=0.52, p=0.122<br>lag 10: EDF=0.00, p=0.941<br>lag 4: EDF=0.53, p=0.092<br>**lag 0: EDF=0.84, p=0.012** | **lag 10: EDF=0.91, p=0.005**<br>**lag 2: EDF=0.84, p=0.011**<br>lag 4: EDF=0.00, p=0.988<br>lag 0: EDF=0.00, p=0.284 | **lag 0: EDF=0.87, p=0.015**<br>lag 0: EDF=0.00, p=0.708<br>**lag 10: EDF=0.79, p=0.028**<br>lag 0: EDF=0.00, p=0.807 | lag 2: EDF=0.38, p=0.201<br>**lag 3: EDF=0.92, p=0.001**<br>lag 4: EDF=0.00, p=0.919<br>lag 10: EDF=0.00, p=0.730 | lag 0: EDF=0.00, p=0.815<br>lag 5: EDF=0.00, p=0.540<br>lag 0: EDF=0.00, p=0.880<br>lag 0: EDF=0.00, p=0.977 | lag 0: EDF=0.00, p=0.893<br>lag 1: EDF=0.58, p=0.098<br>**lag 3: 0.98 (0.96–0.99), p=0.003**<br>lag 2: EDF=0.00, p=0.821 | lag 1: EDF=0.00, p=0.954<br>lag 8: EDF=0.70, p=0.061<br>**lag 0: EDF=0.85, p=0.008**<br>lag 0: EDF=0.00, p=0.438 | lag 0: EDF=0.37, p=0.323<br>lag 9: EDF=0.00, p=0.779<br>**lag 5: EDF=0.92, p=0.004**<br>lag 0: EDF=0.00, p=0.717 |
| Ice day | lag 8: 0.92 (0.74–1.15), p=0.467<br>lag 8: 1.01 (0.95–1.08), p=0.779<br>lag 1: 0.96 (0.84–1.10), p=0.560<br>lag 9: 0.97 (0.91–1.04), p=0.374 | lag 6: 0.83 (0.67–1.04), p=0.105<br>lag 9: 0.99 (0.92–1.06), p=0.703<br>lag 0: 0.97 (0.84–1.12), p=0.662<br>lag 8: 0.99 (0.92–1.08), p=0.878 | lag 5: 0.97 (0.81–1.17), p=0.780<br>lag 7: 0.96 (0.91–1.02), p=0.167<br>lag 8: 0.93 (0.81–1.06), p=0.267<br>lag 9: 0.98 (0.93–1.04), p=0.578 | lag 8: 0.82 (0.65–1.04), p=0.100<br>lag 1: 0.97 (0.91–1.02), p=0.227<br>lag 6: 0.89 (0.77–1.03), p=0.130<br>lag 10: 0.99 (0.92–1.05), p=0.677 | lag 1: 0.88 (0.67–1.14), p=0.329<br>lag 9: 1.01 (0.94–1.09), p=0.758<br>lag 8: 0.97 (0.80–1.20), p=0.806<br>lag 5: 0.97 (0.89–1.06), p=0.511 | lag 9: 0.75 (0.52–1.09), p=0.133<br>lag 10: 0.92 (0.84–1.01), p=0.091<br>**lag 1: 0.77 (0.62–0.97), p=0.026**<br>lag 3: 0.97 (0.89–1.06), p=0.532 | lag 4: 0.88 (0.73–1.06), p=0.176<br>lag 5: 0.99 (0.94–1.05), p=0.771<br>lag 1: 0.96 (0.85–1.09), p=0.516<br>lag 10: 0.97 (0.91–1.04), p=0.408 | lag 0: 0.91 (0.75–1.11), p=0.369<br>lag 5: 0.99 (0.93–1.05), p=0.760<br>lag 8: 1.04 (0.86–1.24), p=0.699<br>lag 1: 1.00 (0.93–1.08), p=0.924 | **lag 0: 0.60 (0.37–0.97), p=0.037**<br>lag 3: 1.13 (0.99–1.28), p=0.068<br>**lag 3: 0.66 (0.47–0.93), p=0.017**<br>**lag 5: 1.27 (1.11–1.46), p=<0.001** | lag 7: 1.03 (0.89–1.19), p=0.707<br>lag 8: 1.01 (0.97–1.06), p=0.564<br>lag 1: 0.98 (0.88–1.09), p=0.705<br>lag 2: 0.98 (0.93–1.03), p=0.387 | **lag 1: 0.49 (0.29–0.83), p=0.008**<br>lag 6: 0.99 (0.86–1.14), p=0.879<br>lag 4: 0.98 (0.75–1.28), p=0.908<br>lag 0: 1.00 (0.89–1.13), p=0.975 |
| Heat stress wave day | **lag 5: 1.15 (1.02–1.30), p=0.025**<br>lag 3: 1.03 (0.99– | lag 3: 1.05 (0.95–1.16), p=0.343<br>**lag 8: 0.95 (0.92–** | **lag 7: 1.26 (1.10–1.44), p=<0.001**<br>lag 1: 1.04 (1.00– | **lag 3: 0.85 (0.75–0.96), p=0.009**<br>lag 4: 0.99 (0.95– | lag 4: 1.16 (0.94–1.43), p=0.162<br>lag 3: 0.94 (0.87– | **lag 5: 1.20 (1.00–1.44), p=0.048**<br>lag 0: 1.03 (0.97– | **lag 2: 1.20 (1.04–1.39), p=0.014**<br>lag 9: 1.05 (1.00– | lag 7: 1.19 (0.99–1.42), p=0.062<br>lag 9: 1.03 (0.97– | lag 0: 1.35 (1.00–1.82), p=0.053<br>lag 10: 1.07 (0.97– | **lag 7: 1.19 (1.06–1.32), p=0.002**<br>**lag 0: 0.95 (0.92–** | **lag 7: 1.31 (1.02–1.68), p=0.032**<br>**lag 10: 1.09 (1.01–** |

| Atmospheric factor | 1.1 | 1.2 | 2.1 | 2.2 | 3.1 | 4.1 | 5.1 | 5.2 | 6.1 | 6.2 | 6.3 |
|---|---|---|---|---|---|---|---|---|---|---|---|
| | 1.08), p=0.119<br>**lag 8: 1.15 (1.06–1.24), p=<0.001**<br>**lag 4: 1.05 (1.00–1.09), p=0.040** | **0.99), p=0.019**<br>**lag 0: 1.16 (1.08–1.25), p=<0.001**<br>lag 0: 1.02 (0.98–1.07), p=0.302 | 1.07), p=0.061<br>lag 0: 1.06 (0.98–1.14), p=0.180<br>lag 6: 1.00 (0.95–1.05), p=0.999 | 1.04), p=0.727<br>lag 2: 1.02 (0.93–1.12), p=0.668<br>lag 4: 1.01 (0.97–1.06), p=0.642 | 1.01), p=0.105<br>**lag 0: 1.20 (1.04–1.39), p=0.011**<br>lag 0: 1.05 (0.98–1.13), p=0.134 | 1.09), p=0.290<br>lag 10: 0.97 (0.83–1.13), p=0.676<br>lag 7: 1.03 (0.97–1.09), p=0.406 | 1.10), p=0.059<br>lag 2: 1.09 (0.98–1.21), p=0.102<br>lag 9: 1.04 (0.99–1.09), p=0.164 | 1.10), p=0.344<br>lag 5: 1.09 (0.95–1.26), p=0.226<br>lag 8: 0.99 (0.91–1.08), p=0.880 | 1.17), p=0.199<br>lag 2: 0.85 (0.63–1.14), p=0.274<br>lag 1: 0.97 (0.87–1.08), p=0.543 | **0.99), p=0.007**<br>**lag 2: 1.12 (1.04–1.21), p=0.004**<br>lag 5: 1.04 (1.00–1.08), p=0.058 | **1.19), p=0.036**<br>lag 3: 1.08 (0.89–1.32), p=0.446<br>lag 4: 1.06 (0.97–1.16), p=0.181 |
| Heat stress wave intensity | **lag 1: EDF=0.78, p=0.030**<br>lag 3: EDF=0.00, p=0.742<br>lag 8: EDF=0.00, p=0.340<br>lag 8: EDF=0.22, p=0.249 | lag 0: EDF=0.59, p=0.093<br>lag 2: EDF=0.53, p=0.134<br>lag 0: EDF=0.00, p=0.431<br>lag 1: EDF=0.53, p=0.132 | **lag 7: EDF=0.91, p=<0.001**<br>lag 4: EDF=0.00, p=0.894<br>lag 9: EDF=0.41, p=0.160<br>**lag 6: EDF=0.85, p=0.007** | lag 10: EDF=0.05, p=0.268<br>lag 4: EDF=0.54, p=0.132<br>**lag 3: EDF=0.71, p=0.046**<br>lag 4: EDF=0.31, p=0.218 | lag 0: EDF=0.49, p=0.211<br>lag 2: EDF=0.42, p=0.167<br>lag 0: EDF=0.00, p=0.696<br>lag 0: EDF=0.00, p=0.898 | lag 2: EDF=0.00, p=0.676<br>lag 6: EDF=0.67, p=0.075<br>lag 10: EDF=0.50, p=0.134<br>lag 5: EDF=0.00, p=0.942 | lag 2: EDF=0.00, p=0.349<br>lag 9: EDF=0.00, p=0.891<br>lag 2: EDF=0.00, p=0.564<br>lag 1: EDF=0.00, p=0.418 | lag 0: EDF=0.00, p=0.990<br>lag 10: EDF=0.01, p=0.361<br>lag 5: EDF=0.02, p=0.310<br>lag 8: EDF=0.65, p=0.069 | lag 0: EDF=0.00, p=0.911<br>**lag 7: EDF=0.79, p=0.017**<br>lag 10: EDF=0.00, p=0.536<br>lag 4: EDF=0.03, p=0.288 | lag 0: EDF=0.03, p=0.292<br>**lag 5: EDF=0.75, p=0.043**<br>lag 2: EDF=0.00, p=0.442<br>lag 1: EDF=0.69, p=0.066 | **lag 1: EDF=0.89, p=0.023**<br>lag 10: EDF=0.00, p=0.859<br>lag 3: EDF=0.44, p=0.219<br>lag 10: EDF=0.23, p=0.216 |
| Cold wave duration | lag 6: EDF=0.00, p=0.589<br>lag 4: EDF=0.00, p=0.657 | - | lag 5: EDF=0.00, p=0.734<br>lag 4: EDF=0.00, p=0.859 | lag 10: EDF=0.43, p=0.168<br>lag 10: EDF=0.05, p=0.301<br>**lag 5: EDF=0.79, p=0.026** | lag 10: EDF=0.72, p=0.091<br>lag 2: EDF=0.47, p=0.149<br>lag 4: EDF=0.14, p=0.284 | lag 10: EDF=0.74, p=0.077<br>lag 8: EDF=0.00, p=0.572<br>lag 1: EDF=0.00, p=0.294 | lag 7: EDF=0.00, p=0.527<br>lag 8: EDF=0.01, p=0.305 | lag 5: EDF=0.00, p=0.705<br>lag 2: EDF=0.00, p=0.719 | **lag 9: EDF=0.83, p=0.047**<br>**lag 2: EDF=0.93, p=<0.001** | lag 8: EDF=0.41, p=0.173<br>lag 10: EDF=0.00, p=0.381 | - |
| Cold stress wave | lag 8: 0.89 (0.75–1.07), p=0.219<br>lag 3: 0.95 (0.90–1.01), | lag 2: 0.95 (0.82–1.12), p=0.562<br>lag 10: 0.96 (0.91–1.01), | lag 4: 1.15 (0.78–1.71), p=0.480<br>lag 5: 0.96 (0.92–1.02), | lag 7: 1.00 (0.82–1.22), p=0.991<br>lag 10: 0.97 (0.93–1.02), | lag 0: 1.01 (0.74–1.38), p=0.963<br>lag 10: 1.02 (0.90–1.16), | lag 7: 1.06 (0.76–1.47), p=0.749<br>lag 0: 0.92 (0.85–1.00), | lag 9: 0.94 (0.87–1.01), p=0.110<br>lag 10: 1.00 (0.88–1.14), | lag 8: 1.05 (0.97–1.13), p=0.231<br>lag 7: 0.85 (0.63–1.15), | lag 10: 1.08 (0.72–1.61), p=0.723<br>lag 0: 0.94 (0.82–1.06), | lag 9: 0.95 (0.81–1.11), p=0.487<br>**lag 4: 1.05 (1.01–1.10),** | lag 0: 1.60 (0.94–2.72), p=0.081<br>lag 5: 0.96 (0.84–1.09), |

| **Atmospheric factor** | **1.1** | **1.2** | **2.1** | **2.2** | **3.1** | **4.1** | **5.1** | **5.2** | **6.1** | **6.2** | **6.3** |
|---|---|---|---|---|---|---|---|---|---|---|---|
| | p=0.075<br>lag 7: 1.00 (0.90–1.11), p=0.984<br>lag 2: 1.00 (0.94–1.06), p=0.923 | p=0.134<br>lag 0: 0.95 (0.86–1.06), p=0.387<br>**lag 8: 0.93 (0.88–0.99), p=0.023** | p=0.182<br>lag 7: 1.00 (0.88–1.13), p=0.983<br>lag 7: 1.01 (0.96–1.06), p=0.743 | p=0.257<br>lag 0: 0.98 (0.88–1.11), p=0.782<br>lag 5: 1.01 (0.95–1.08), p=0.708 | p=0.743<br>lag 4: 0.83 (0.64–1.08), p=0.174<br>lag 9: 0.98 (0.89–1.08), p=0.680 | p=0.054<br>**lag 0: 1.42 (1.18–1.70), p=<0.001**<br>lag 6: 0.92 (0.83–1.02), p=0.118 | p=0.996<br>lag 3: 0.98 (0.91–1.05), p=0.486 | p=0.291<br>lag 9: 1.02 (0.94–1.11), p=0.593 | p=0.311<br>lag 4: 1.30 (0.95–1.77), p=0.098<br>**lag 8: 1.28 (1.13–1.46), p=<0.001** | **p=0.021**<br>lag 9: 0.97 (0.88–1.08), p=0.603<br>lag 10: 0.99 (0.94–1.03), p=0.564 | p=0.482<br>lag 8: 1.08 (0.87–1.35), p=0.495<br>lag 10: 1.05 (0.94–1.16), p=0.378 |
| Cold stress wave duration | lag 8: EDF=0.01, p=0.300<br>lag 3: EDF=0.00, p=0.498<br>lag 2: EDF=0.00, p=0.490 | lag 10: EDF=0.00, p=0.384<br>lag 0: EDF=0.00, p=0.534<br>lag 4: EDF=0.00, p=0.469 | **lag 4: EDF=2.10, p=0.019**<br>lag 0: EDF=0.00, p=0.757<br>lag 2: EDF=0.00, p=0.808 | lag 9: EDF=0.00, p=0.758<br>lag 2: EDF=0.00, p=0.581 | lag 10: EDF=0.52, p=0.132 | lag 0: EDF=0.35, p=0.197<br>**lag 6: EDF=0.86, p=0.005** | lag 2: EDF=0.00, p=0.527 | **lag 5: 1.04 (1.02–1.07), p=<0.001**<br>lag 8: EDF=0.00, p=0.377<br>lag 7: EDF=0.31, p=0.207<br>lag 1: EDF=0.00, p=0.917 | lag 7: EDF=0.00, p=0.639<br>lag 10: EDF=0.00, p=0.508 | lag 7: EDF=0.23, p=0.220<br>lag 5: EDF=0.00, p=0.467 | lag 0: EDF=0.83, p=0.068<br>lag 7: EDF=0.15, p=0.247<br>lag 9: EDF=0.00, p=0.940 |
| Cold stress wave intensity | lag 10: EDF=0.65, p=0.089<br>lag 8: EDF=0.07, p=0.266<br>lag 5: EDF=0.00, p=0.421 | lag 4: EDF=0.42, p=0.156<br>lag 1: EDF=0.00, p=0.890<br>lag 7: EDF=0.00, p=0.842<br>lag 3: EDF=0.00, p=0.895 | lag 0: EDF=0.00, p=0.952<br>lag 2: EDF=0.59, p=0.114<br>lag 6: EDF=0.00, p=0.448<br>lag 1: EDF=0.00, p=0.612 | lag 1: EDF=0.02, p=0.302 | **lag 4: EDF=2.98, p=0.002** | - | lag 9: EDF=0.66, p=0.075<br>lag 8: EDF=0.06, p=0.268<br>lag 4: EDF=0.46, p=0.164 | **lag 10: EDF=0.86, p=0.018**<br>lag 7: EDF=0.00, p=0.674<br>lag 10: EDF=0.62, p=0.100<br>lag 9: EDF=0.00, p=0.485 | lag 2: EDF=0.72, p=0.191<br>lag 6: EDF=0.00, p=0.287 | lag 10: EDF=0.00, p=0.673 | lag 10: EDF=0.00, p=0.977<br>lag 0: EDF=0.00, p=0.449<br>lag 8: EDF=0.00, p=0.379 |

**Table S13.** Overview of time lags, effect sizes (effective degree of freedom (EDF) or relative risk (95% confidence interval)) and p-values per human-bioclimatic region, atmospheric factor and subgroup (pink = female <60 years, orange = females ≥60 years, green = males <60 years, blue = males ≥60 years) for pulmonary artery embolism, significant p-values (≤ 0.05) are bold

| **Atmospheric factor** | **1.1** | **1.2** | **2.1** | **2.2** | **3.1** | **4.1** | **5.1** | **5.2** | **6.1** | **6.2** | **6.3** |
|---|---|---|---|---|---|---|---|---|---|---|---|
| Sea-level pressure (SLP) | lag 1: EDF=0.77, p=0.102<br>lag 0: EDF=0.00, p=0.423<br>lag 4: EDF=0.35, p=0.289<br>lag 10: EDF=0.66, p=0.098 | **lag 3: EDF=0.88, p=0.016**<br>**lag 0: EDF=0.88, p=0.005**<br>lag 2: EDF=0.63, p=0.129<br>lag 7: EDF=0.63, p=0.100 | **lag 4: EDF=0.85, p=0.026**<br>lag 4: EDF=0.66, p=0.074<br>lag 2: EDF=0.00, p=0.637<br>**lag 3: EDF=0.89, p=0.005** | lag 9: EDF=0.42, p=0.270<br>lag 3: EDF=0.44, p=0.160<br>lag 9: EDF=0.41, p=0.244<br>**lag 5: EDF=0.95, p=<0.001** | lag 5: EDF=0.11, p=0.483<br>lag 10: EDF=0.62, p=0.130<br>**lag 7: EDF=0.92, p=0.035**<br>**lag 7: EDF=0.92, p=0.009** | lag 0: EDF=0.63, p=0.297<br>**lag 3: EDF=0.95, p=0.002**<br>**lag 7: 0.98 (0.98–0.99), p=0.001**<br>lag 8: EDF=0.67, p=0.165 | lag 8: EDF=0.86, p=0.050<br>**lag 1: EDF=0.81, p=0.023**<br>lag 9: EDF=0.55, p=0.252<br>lag 7: EDF=0.40, p=0.220 | **lag 1: EDF=3.74, p=0.005**<br>lag 10: EDF=0.69, p=0.095<br>lag 8: EDF=0.88, p=0.067<br>**lag 9: EDF=0.93, p=0.003** | **lag 0: EDF=7.28, p=0.022**<br>lag 1: EDF=0.50, p=0.323<br>lag 9: EDF=0.00, p=0.705<br>**lag 1: 1.01 (1.00–1.02), p=0.011** | lag 4: EDF=0.48, p=0.244<br>lag 8: EDF=0.00, p=0.331<br>lag 10: EDF=0.79, p=0.055<br>**lag 1: EDF=0.86, p=0.010** | lag 4: EDF=6.68, p=0.081<br>lag 7: EDF=0.56, p=0.261<br>**lag 9: 1.02 (1.00–1.03), p=0.021**<br>lag 6: EDF=2.46, p=0.174 |
| Sea-level pressure difference (SLPD) | lag 6: 0.82 (0.66–1.02), p=0.076<br>**lag 2: 0.86 (0.76–0.97), p=0.013**<br>lag 4: 0.90 (0.74–1.10), p=0.298<br>lag 5: 1.08 (0.94–1.25), p=0.273 | lag 6: 1.15 (0.95–1.39), p=0.140<br>lag 3: 0.93 (0.84–1.03), p=0.147<br>lag 1: 0.86 (0.73–1.01), p=0.064<br>lag 1: 0.91 (0.80–1.03), p=0.124 | lag 3: 1.08 (0.87–1.35), p=0.485<br>lag 2: 1.07 (0.96–1.20), p=0.224<br>**lag 7: 1.23 (1.00–1.51), p=0.048**<br>lag 2: 1.08 (0.94–1.24), p=0.269 | lag 2: 0.83 (0.65–1.06), p=0.133<br>**lag 5: 0.88 (0.79–0.99), p=0.032**<br>lag 10: 0.86 (0.70–1.06), p=0.166<br>**lag 3: 0.81 (0.70–0.93), p=0.003** | **lag 5: 1.51 (1.04–2.20), p=0.030**<br>lag 2: 1.12 (0.92–1.36), p=0.256<br>**lag 2: 0.65 (0.45–0.93), p=0.018**<br>lag 6: 0.89 (0.69–1.15), p=0.370 | **lag 4: 0.67 (0.49–0.92), p=0.013**<br>lag 3: 0.93 (0.79–1.11), p=0.437<br>lag 9: 0.82 (0.60–1.12), p=0.211<br>lag 1: 0.82 (0.67–1.01), p=0.061 | lag 2: 0.80 (0.61–1.05), p=0.107<br>**lag 1: 0.83 (0.73–0.95), p=0.009**<br>lag 6: 1.25 (0.97–1.61), p=0.083<br>lag 9: 1.13 (0.96–1.33), p=0.135 | **lag 10: 0.71 (0.50–0.99), p=0.044**<br>lag 4: 1.10 (0.92–1.30), p=0.292<br>**lag 2: 0.71 (0.51–0.99), p=0.041**<br>**lag 4: 0.79 (0.65–0.98), p=0.028** | **lag 10: 1.92 (1.10–3.34), p=0.022**<br>**lag 9: 1.44 (1.10–1.88), p=0.008**<br>lag 7: 1.72 (0.99–2.97), p=0.053<br>lag 6: 0.74 (0.53–1.04), p=0.080 | lag 7: 0.82 (0.66–1.02), p=0.081<br>**lag 8: 0.90 (0.82–0.99), p=0.036**<br>lag 7: 1.14 (0.94–1.37), p=0.177<br>lag 8: 1.12 (0.99–1.26), p=0.077 | lag 4: 1.42 (0.88–2.27), p=0.147<br>lag 1: 1.26 (0.99–1.60), p=0.057<br>**lag 3: 0.59 (0.40–0.88), p=0.009**<br>lag 2: 1.30 (0.98–1.72), p=0.068 |
| Wind day | **lag 7: 1.42 (1.10–1.84), p=0.007**<br>**lag 3: 1.22** | lag 10: 0.78 (0.59–1.03), p=0.082<br>lag 5: 1.08 | **lag 7: 1.34 (1.08–1.67), p=0.008**<br>lag 1: 1.08 | **lag 8: 1.40 (1.11–1.78), p=0.005**<br>**lag 9: 1.19** | **lag 10: 1.52 (1.08–2.14), p=0.017**<br>lag 10: 1.21 | **lag 10: 1.74 (1.23–2.45), p=0.002**<br>lag 4: 1.14 | lag 9: 1.19 (0.87–1.62), p=0.284<br>**lag 3: 1.26** | lag 5: 1.30 (0.89–1.90), p=0.169<br>lag 4: 1.19 | **lag 4: 2.29 (1.21–4.34), p=0.011**<br>**lag 7: 1.46** | **lag 9: 1.31 (1.02–1.66), p=0.031**<br>lag 8: 1.05 | **lag 10: 2.58 (1.62–4.13), p=<0.001**<br>lag 10: 1.22 |

| Atmospheric factor | 1.1 | 1.2 | 2.1 | 2.2 | 3.1 | 4.1 | 5.1 | 5.2 | 6.1 | 6.2 | 6.3 |
|---|---|---|---|---|---|---|---|---|---|---|---|
| | **(1.05–1.41), p=0.008**<br>lag 4: 1.28 (1.00–1.64), p=0.052<br>lag 1: 1.03 (0.86–1.24), p=0.731 | (0.94–1.23), p=0.280<br>lag 1: 1.20 (0.98–1.46), p=0.081<br>**lag 3: 1.35 (1.16–1.56), p=<0.001** | (0.95–1.21), p=0.232<br>**lag 6: 1.24 (1.01–1.52), p=0.044**<br>**lag 8: 0.77 (0.65–0.92), p=0.003** | **(1.06–1.35), p=0.004**<br>lag 10: 1.10 (0.88–1.37), p=0.386<br>**lag 1: 0.81 (0.68–0.97), p=0.020** | (0.99–1.47), p=0.062<br>**lag 1: 1.92 (1.42–2.59), p=<0.001**<br>lag 0: 1.16 (0.91–1.48), p=0.227 | (0.92–1.41), p=0.247<br>lag 6: 0.61 (0.37–1.01), p=0.054<br>lag 9: 1.13 (0.87–1.48), p=0.366 | **(1.09–1.46), p=0.002**<br>lag 10: 1.29 (0.97–1.70), p=0.075<br>**lag 1: 1.28 (1.08–1.52), p=0.005** | (0.98–1.44), p=0.078<br>lag 6: 1.39 (0.99–1.96), p=0.061<br>lag 10: 1.15 (0.92–1.45), p=0.226 | **(1.06–2.01), p=0.020**<br>**lag 9: 2.52 (1.45–4.38), p=<0.001**<br>lag 2: 1.42 (0.94–2.12), p=0.092 | (0.94–1.19), p=0.381<br>**lag 1: 0.62 (0.47–0.82), p=<0.001**<br>lag 2: 1.09 (0.94–1.26), p=0.269 | (0.90–1.65), p=0.192<br>lag 8: 1.52 (0.95–2.42), p=0.080<br>lag 9: 1.33 (0.95–1.86), p=0.096 |
| Tropical night | lag 5: 2.28 (0.63–8.24), p=0.209<br>lag 1: 1.30 (0.60–2.81), p=0.509<br>lag 4: 1.40 (0.40–4.94), p=0.601<br>**lag 5: 2.28 (1.06–4.91), p=0.035** | lag 8: 1.75 (0.85–3.60), p=0.127<br>**lag 5: 1.63 (1.08–2.46), p=0.020**<br>lag 8: 1.55 (0.84–2.86), p=0.157<br>lag 9: 0.48 (0.20–1.16), p=0.103 | lag 8: 2.43 (0.67–8.74), p=0.176<br>**lag 4: 3.18 (1.68–6.03), p=<0.001**<br>**lag 6: 4.09 (1.11–15.06), p=0.034**<br>**lag 6: 2.43 (1.05–5.60), p=0.038** | lag 8: 1.49 (0.74–2.98), p=0.261<br>lag 6: 1.38 (0.93–2.04), p=0.113<br>lag 0: 0.37 (0.10–1.32), p=0.125<br>lag 2: 0.61 (0.29–1.28), p=0.192 | - | lag 9: 4.79 (0.54–42.26), p=0.158<br>**lag 4: 3.87 (1.03–14.55), p=0.045**<br>lag 7: 2.26 (0.21–24.71), p=0.503<br>**lag 1: 5.56 (1.06–29.17), p=0.043** | **lag 2: 14.93 (4.65–47.99), p=<0.001**<br>**lag 5: 3.83 (1.43–10.25), p=0.007**<br>lag 5: 3.73 (0.38–36.34), p=0.257<br>**lag 10: 5.99 (2.26–15.86), p=<0.001** | lag 8: 3.62 (0.37–35.72), p=0.270<br>**lag 5: 4.03 (1.65–9.86), p=0.002**<br>**lag 3: 17.25 (2.46–121.09), p=0.004**<br>**lag 3: 5.64 (1.59–20.01), p=0.007** | lag 10: 2.15 (0.13–36.07), p=0.594<br>lag 1: 2.82 (0.61–12.99), p=0.185<br>lag 10: 8.83 (0.57–137.29), p=0.120<br>lag 9: 2.69 (0.53–13.68), p=0.233 | **lag 10: 2.25 (1.19–4.26), p=0.012**<br>lag 2: 0.54 (0.28–1.06), p=0.075<br>lag 4: 1.82 (0.92–3.58), p=0.084<br>lag 10: 1.07 (0.62–1.84), p=0.803 | **lag 5: 8.16 (1.30–51.41), p=0.025**<br>lag 8: 3.03 (0.81–11.30), p=0.099<br>lag 3: 1.58 (0.16–15.31), p=0.694<br>lag 3: 2.76 (0.56–13.61), p=0.213 |
| Summer day | **lag 0: 0.73 (0.54–0.99), p=0.040**<br>**lag 2: 1.15 (1.00–1.33), p=0.047**<br>**lag 2: 0.77 (0.61–0.99), p=0.041** | lag 10: 1.12 (0.92–1.37), p=0.248<br>lag 1: 0.90 (0.79–1.03), p=0.127<br>lag 9: 1.04 (0.87–1.25), p=0.666 | lag 8: 1.08 (0.87–1.34), p=0.471<br>lag 2: 0.98 (0.89–1.08), p=0.705<br>lag 8: 0.93 (0.77–1.12), p=0.441 | lag 2: 0.82 (0.65–1.05), p=0.112<br>lag 0: 0.97 (0.87–1.07), p=0.494<br>lag 2: 0.89 (0.75–1.05), p=0.174 | lag 6: 0.61 (0.37–1.01), p=0.054<br>lag 2: 1.01 (0.80–1.27), p=0.934<br>**lag 9: 1.50 (1.06–2.14), p=0.023** | lag 2: 1.19 (0.66–2.14), p=0.563<br>lag 5: 0.77 (0.54–1.09), p=0.135<br>lag 10: 1.14 (0.67–1.93), p=0.633 | lag 2: 0.77 (0.55–1.07), p=0.123<br>**lag 5: 0.80 (0.68–0.94), p=0.008**<br>lag 0: 0.78 (0.56–1.06), p=0.116 | **lag 8: 0.45 (0.28–0.73), p=0.001**<br>lag 10: 0.94 (0.74–1.19), p=0.607<br>lag 3: 0.70 (0.41–1.22), p=0.211 | lag 4: 0.48 (0.11–2.09), p=0.330<br>lag 4: 0.73 (0.49–1.08), p=0.118<br>lag 1: 1.03 (0.44–2.42), p=0.949 | lag 9: 1.14 (0.94–1.38), p=0.186<br>lag 3: 0.93 (0.84–1.03), p=0.158<br>lag 5: 1.19 (0.99–1.42), p=0.057 | lag 1: 1.09 (0.56–2.12), p=0.804<br>lag 7: 1.05 (0.77–1.43), p=0.742<br>lag 7: 1.35 (0.83–2.19), p=0.226 |

| Atmospheric factor | 1.1 | 1.2 | 2.1 | 2.2 | 3.1 | 4.1 | 5.1 | 5.2 | 6.1 | 6.2 | 6.3 |
|---|---|---|---|---|---|---|---|---|---|---|---|
| | lag 1: 0.89 (0.72–1.10), p=0.280 | lag 10: 0.85 (0.71–1.02), p=0.088 | **lag 10: 0.89 (0.79–1.00), p=0.047** | **lag 10: 1.16 (1.04–1.31), p=0.010** | lag 9: 0.79 (0.60–1.04), p=0.089 | **lag 1: 1.91 (1.34–2.71), p=<0.001** | | lag 1: 0.81 (0.64–1.03), p=0.087 | lag 6: 0.71 (0.45–1.14), p=0.156 | lag 4: 0.92 (0.81–1.04), p=0.159 | lag 10: 0.92 (0.59–1.46), p=0.732 |
| Ice day | lag 3: 1.42 (0.85–2.35), p=0.177<br>lag 2: 1.07 (0.89–1.29), p=0.461<br>lag 0: 1.28 (0.95–1.71), p=0.103<br>lag 7: 1.14 (0.93–1.40), p=0.220 | lag 4: 1.35 (1.00–1.82), p=0.052<br>lag 9: 1.07 (0.90–1.27), p=0.453<br>lag 10: 1.29 (0.98–1.69), p=0.067<br>lag 6: 0.85 (0.66–1.10), p=0.214 | lag 7: 0.79 (0.60–1.05), p=0.102<br>lag 10: 1.07 (0.95–1.21), p=0.275<br>**lag 7: 0.65 (0.49–0.86), p=0.003**<br>lag 2: 1.06 (0.91–1.23), p=0.435 | lag 0: 0.95 (0.66–1.37), p=0.776<br>**lag 7: 1.25 (1.08–1.44), p=0.002**<br>**lag 4: 1.32 (1.02–1.71), p=0.033**<br>**lag 9: 1.28 (1.08–1.52), p=0.005** | lag 5: 0.66 (0.43–1.01), p=0.058<br>**lag 3: 0.76 (0.62–0.92), p=0.006**<br>lag 3: 0.98 (0.68–1.41), p=0.907<br>lag 10: 0.97 (0.72–1.29), p=0.814 | **lag 9: 1.59 (1.03–2.45), p=0.036**<br>lag 10: 1.05 (0.81–1.35), p=0.722<br>**lag 8: 3.28 (1.73–6.23), p=<0.001**<br>**lag 0: 1.36 (1.02–1.82), p=0.039** | lag 0: 0.75 (0.52–1.09), p=0.135<br>lag 6: 1.11 (0.96–1.28), p=0.177<br>lag 4: 1.25 (0.92–1.69), p=0.153<br>lag 9: 1.06 (0.88–1.27), p=0.546 | lag 7: 0.74 (0.49–1.12), p=0.157<br>lag 3: 1.07 (0.91–1.27), p=0.411<br>**lag 8: 0.65 (0.44–0.96), p=0.030**<br>**lag 6: 1.38 (1.14–1.66), p=<0.001** | **lag 5: 2.30 (1.20–4.42), p=0.013**<br>lag 5: 1.11 (0.78–1.56), p=0.561<br>lag 5: 1.47 (0.72–2.97), p=0.289<br>lag 2: 1.33 (0.90–1.97), p=0.149 | lag 8: 1.21 (0.93–1.59), p=0.161<br>lag 1: 1.07 (0.94–1.20), p=0.296<br>lag 1: 0.87 (0.66–1.14), p=0.301<br>lag 2: 1.07 (0.92–1.25), p=0.388 | **lag 9: 1.94 (1.06–3.56), p=0.033**<br>**lag 4: 0.55 (0.35–0.85), p=0.008**<br>lag 8: 0.55 (0.23–1.30), p=0.172<br>**lag 2: 0.46 (0.26–0.81), p=0.007** |
| Wind chill index | lag 3: EDF=3.65, p=0.087<br>lag 1: EDF=0.52, p=0.140<br>lag 10: EDF=0.00, p=0.463<br>**lag 0: EDF=0.87, p=0.012** | **lag 1: EDF=0.92, p=0.007**<br>lag 2: EDF=0.00, p=0.537<br>lag 7: EDF=0.64, p=0.096<br>**lag 8: EDF=0.86, p=0.011** | lag 9: EDF=0.00, p=0.876<br>lag 0: EDF=0.00, p=0.342 | lag 5: EDF=0.00, p=0.662<br>**lag 3: EDF=0.81, p=0.020**<br>lag 0: EDF=0.00, p=0.453<br>lag 0: EDF=0.41, p=0.138 | lag 1: EDF=0.00, p=0.503<br>**lag 0: EDF=1.14, p=0.004**<br>lag 10: EDF=2.26, p=0.139 | lag 1: EDF=0.90, p=0.068<br>lag 0: EDF=0.00, p=0.566<br>**lag 7: EDF=3.57, p=0.004**<br>**lag 10: EDF=0.83, p=0.049** | lag 4: EDF=0.66, p=0.192<br>lag 7: EDF=0.00, p=0.586<br>lag 7: EDF=0.82, p=0.086<br>lag 0: EDF=0.00, p=0.666 | lag 8: EDF=0.83, p=0.124<br>lag 10: EDF=0.00, p=0.861<br>**lag 2: EDF=0.90, p=0.030**<br>lag 0: EDF=0.61, p=0.157 | lag 6: EDF=0.54, p=0.558<br>lag 3: EDF=0.67, p=0.220<br>lag 10: EDF=4.93, p=0.109<br>lag 6: EDF=0.53, p=0.357 | lag 8: EDF=0.00, p=0.449<br>lag 0: EDF=0.00, p=0.525<br>lag 9: EDF=0.24, p=0.311<br>lag 4: EDF=0.00, p=0.984 | lag 1: 0.97 (0.93–1.00), p=0.063<br>lag 1: EDF=0.00, p=0.516<br>lag 10: EDF=0.13, p=0.566<br>lag 10: EDF=0.00, p=0.704 |
| Heat wave duration | lag 7: EDF=0.67, p=0.144<br>**lag 10: EDF=0.85,** | lag 10: EDF=0.37, p=0.230<br>lag 7: | lag 5: EDF=0.00, p=0.776<br>lag 7: EDF=0.42, | lag 10: EDF=0.63, p=0.156<br>lag 3: EDF=0.63, | lag 10: EDF=0.00, p=0.944<br>lag 8: EDF=0.00, | **lag 9: EDF=3.00, p=<0.001**<br>lag 8: EDF=0.01, | lag 6: EDF=0.00, p=0.548<br>**lag 3: EDF=0.84,** | - | lag 0: 1.05 (0.98–1.13), p=0.172<br>lag 7: | lag 2: EDF=0.00, p=0.435<br>**lag 3: EDF=0.95,** | **lag 0: 0.83 (0.71–0.98), p=0.025**<br>lag 10: |

| Atmospheric factor | 1.1 | 1.2 | 2.1 | 2.2 | 3.1 | 4.1 | 5.1 | 5.2 | 6.1 | 6.2 | 6.3 |
|---|---|---|---|---|---|---|---|---|---|---|---|
| | **p=0.025**<br>**lag 8: EDF=0.83, p=0.016** | EDF=0.00, p=0.527 | p=0.172<br>**lag 10: EDF=0.90, p=0.004** | p=0.090<br>lag 3: EDF=0.72, p=0.098<br>lag 2: EDF=0.73, p=0.060 | p=0.764<br>**lag 6: EDF=0.89, p=0.008** | p=0.524<br>lag 6: EDF=0.00, p=0.988 | **p=0.009**<br>lag 2: EDF=0.00, p=0.571 | | EDF=0.63, p=0.230<br>**lag 10: 1.08 (1.01–1.14), p=0.015**<br>lag 0: EDF=0.00, p=0.798 | **p=<0.001**<br>lag 9: EDF=0.00, p=0.700<br>**lag 1: EDF=0.88, p=0.002** | EDF=0.54, p=0.294<br>lag 4: EDF=0.00, p=0.696<br>lag 0: EDF=0.68, p=0.247 |
| Heat wave intensity | lag 0: EDF=0.00, p=0.491 | lag 1: EDF=0.00, p=0.429<br>lag 8: EDF=0.00, p=0.711<br>lag 6: EDF=0.00, p=0.439 | lag 8: EDF=0.23, p=0.305<br>lag 0: EDF=0.54, p=0.113 | lag 0: EDF=0.57, p=0.116<br>lag 9: EDF=0.00, p=0.831 | lag 0: EDF=0.72, p=0.213<br>lag 10: EDF=0.73, p=0.099 | lag 0: EDF=0.00, p=0.663<br>lag 6: EDF=3.99, p=0.056<br>lag 10: EDF=0.00, p=0.884 | lag 8: EDF=0.00, p=0.605<br>lag 5: EDF=0.00, p=0.427<br>lag 7: EDF=0.25, p=0.351<br>**lag 0: EDF=0.77, p=0.048** | **lag 10: EDF=4.51, p=<0.001**<br>lag 7: EDF=0.10, p=0.325<br>**lag 3: EDF=5.12, p=0.002**<br>lag 3: EDF=0.00, p=0.545 | **lag 3: EDF=0.90, p=0.017**<br>**lag 8: 0.87 (0.77–0.99), p=0.031** | lag 5: EDF=0.73, p=0.101<br>**lag 0: EDF=0.93, p=<0.001**<br>lag 3: EDF=0.00, p=0.444 | lag 3: EDF=0.88, p=0.150<br>lag 6: EDF=3.70, p=0.084<br>lag 10: EDF=0.38, p=0.349 |
| Heat stress wave day | **lag 8: 1.19 (1.04–1.36), p=0.011**<br>lag 10: 1.03 (0.81–1.31), p=0.809<br>lag 0: 0.92 (0.74–1.14), p=0.439 | lag 3: 0.83 (0.65–1.06), p=0.138<br>lag 0: 0.88 (0.76–1.02), p=0.088<br>lag 7: 1.13 (0.91–1.40), p=0.261<br>lag 10: 1.05 (0.86–1.28), p=0.624 | lag 8: 1.08 (0.83–1.42), p=0.566<br>lag 10: 1.08 (0.96–1.20), p=0.192<br>lag 3: 0.86 (0.69–1.08), p=0.190<br>lag 5: 1.10 (0.93–1.30), p=0.263 | lag 1: 0.82 (0.60–1.11), p=0.205<br>lag 1: 0.94 (0.81–1.08), p=0.362<br>lag 6: 1.16 (0.94–1.44), p=0.174<br>lag 1: 0.93 (0.79–1.09), p=0.364 | **lag 10: 1.58 (1.12–2.24), p=0.009**<br>lag 1: 1.02 (0.77–1.34), p=0.903<br>lag 10: 1.07 (0.73–1.56), p=0.740<br>**lag 1: 1.31 (1.04–1.66), p=0.023** | lag 1: 1.10 (0.68–1.77), p=0.692<br>**lag 9: 0.51 (0.35–0.74), p=<0.001**<br>lag 8: 0.95 (0.57–1.57), p=0.839<br>lag 7: 1.23 (0.95–1.59), p=0.108 | lag 9: 1.20 (0.89–1.61), p=0.242<br>**lag 2: 1.46 (1.05–2.02), p=0.024**<br>lag 2: 0.90 (0.71–1.16), p=0.421 | lag 0: 0.58 (0.34–1.01), p=0.052<br>lag 10: 1.09 (0.81–1.47), p=0.568<br>lag 3: 0.65 (0.31–1.39), p=0.268<br>**lag 4: 0.73 (0.56–0.96), p=0.024** | lag 3: 0.24 (0.04–1.39), p=0.110<br>lag 0: 0.76 (0.50–1.14), p=0.185<br>lag 0: 1.63 (0.68–3.88), p=0.273<br>**lag 2: 1.58 (1.07–2.32), p=0.022** | **lag 4: 1.33 (1.05–1.68), p=0.020**<br>**lag 4: 0.81 (0.70–0.93), p=0.003**<br>lag 6: 1.11 (0.89–1.39), p=0.349<br>**lag 10: 1.22 (1.06–1.40), p=0.005** | lag 4: 0.92 (0.64–1.33), p=0.669<br>**lag 5: 0.46 (0.21–0.97), p=0.042**<br>**lag 10: 0.47 (0.24–0.91), p=0.024** |

| Atmospheric factor | 1.1 | 1.2 | 2.1 | 2.2 | 3.1 | 4.1 | 5.1 | 5.2 | 6.1 | 6.2 | 6.3 |
|---|---|---|---|---|---|---|---|---|---|---|---|
| Heat stress wave intensity | lag 1: EDF=0.71, p=0.143<br>lag 1: EDF=0.42, p=0.173 | lag 0: EDF=0.00, p=0.759<br>lag 2: EDF=0.20, p=0.255 | lag 1: EDF=0.60, p=0.150<br>lag 3: EDF=0.01, p=0.336<br>lag 7: EDF=0.73, p=0.089<br>**lag 5: EDF=0.85, p=0.007** | lag 9: EDF=0.00, p=0.609<br>lag 9: EDF=0.30, p=0.290<br>lag 10: EDF=0.55, p=0.125 | **lag 3: EDF=2.70, p=0.008**<br>lag 1: EDF=0.76, p=0.061<br>**lag 4: EDF=0.89, p=0.041**<br>lag 1: EDF=0.00, p=0.749 | lag 1: EDF=0.00, p=0.724<br>**lag 0: EDF=0.90, p=0.015** | lag 4: EDF=0.00, p=0.518<br>lag 10: EDF=0.27, p=0.223<br>**lag 1: EDF=0.85, p=0.050**<br>lag 2: EDF=0.39, p=0.224 | lag 1: EDF=0.69, p=0.205<br>**lag 10: EDF=0.79, p=0.049**<br>lag 8: EDF=0.70, p=0.099 | lag 9: EDF=0.86, p=0.290<br>lag 5: EDF=0.81, p=0.101<br>lag 7: EDF=0.00, p=0.756<br>lag 8: EDF=0.54, p=0.336 | lag 9: EDF=0.00, p=0.368 | lag 10: EDF=0.70, p=0.355<br>lag 6: EDF=0.68, p=0.202<br>lag 3: EDF=0.04, p=0.584 |
| Ozone | lag 0: EDF=0.00, p=0.611<br>**lag 4: EDF=0.90, p=0.003**<br>**lag 4: 0.99 (0.98–1.00), p=0.005**<br>**lag 4: EDF=0.87, p=0.013** | lag 4: EDF=0.00, p=0.639<br>**lag 7: EDF=0.89, p=0.003**<br>**lag 7: 0.99 (0.98–1.00), p=<0.001**<br>lag 6: EDF=0.00, p=0.680 | **lag 4: 0.99 (0.98–1.00), p=0.004**<br>**lag 2: EDF=0.91, p=0.002**<br>**lag 7: EDF=0.87, p=0.019**<br>**lag 1: 0.99 (0.99–1.00), p=<0.001** | lag 0: EDF=0.00, p=0.467<br>lag 1: EDF=0.02, p=0.288<br>lag 2: EDF=0.40, p=0.158 | **lag 6: EDF=0.97, p=0.037**<br>**lag 6: EDF=0.79, p=0.026**<br>**lag 5: 0.98 (0.97–1.00), p=0.018**<br>**lag 0: 0.99 (0.99–1.00), p=0.004** | **lag 4: EDF=0.95, p=0.032**<br>lag 7: EDF=1.69, p=0.139<br>lag 7: EDF=0.01, p=0.516<br>**lag 2: EDF=0.87, p=0.033** | **lag 7: EDF=0.90, p=0.042**<br>**lag 7: EDF=0.83, p=0.021**<br>lag 1: EDF=0.87, p=0.058<br>lag 0: EDF=0.01, p=0.323 | **lag 3: EDF=3.68, p=0.005**<br>**lag 2: EDF=0.91, p=0.005**<br>lag 6: EDF=0.00, p=0.367 | lag 6: EDF=0.82, p=0.362<br>lag 1: EDF=0.01, p=0.454<br>lag 6: EDF=0.82, p=0.345<br>**lag 7: EDF=3.13, p=0.028** | lag 6: EDF=0.00, p=0.948<br>lag 1: EDF=0.15, p=0.236<br>**lag 0: EDF=2.29, p=<0.001**<br>**lag 6: 0.99 (0.99–1.00), p=<0.001** | lag 2: 0.98 (0.96–1.01), p=0.145<br>**lag 6: 0.99 (0.98–1.00), p=0.005**<br>**lag 6: EDF=3.88, p=0.033**<br>lag 1: EDF=0.60, p=0.242 |
| Particulate matter ($PM_{10}$) | lag 4: EDF=0.47, p=0.270<br>lag 4: EDF=0.58, p=0.119<br>lag 2: EDF=0.03, p=0.405<br>**lag 5: EDF=0.80, p=0.030** | lag 5: EDF=0.68, p=0.130<br>**lag 5: EDF=0.88, p=0.003**<br>lag 4: EDF=0.00, p=0.662<br>lag 4: EDF=0.00, p=0.907 | lag 4: EDF=0.78, p=0.057<br>lag 3: EDF=0.69, p=0.056<br>**lag 5: EDF=1.72, p=0.010**<br>**lag 6: EDF=2.90, p=<0.001** | lag 3: EDF=0.00, p=0.782<br>**lag 1: EDF=0.86, p=0.006**<br>lag 4: EDF=0.00, p=0.863<br>lag 1: EDF=0.50, p=0.155 | lag 7: EDF=0.66, p=0.249<br>lag 0: EDF=0.00, p=0.704<br>lag 2: EDF=0.00, p=0.647<br>**lag 5: EDF=2.50, p=0.012** | lag 5: EDF=0.00, p=0.647<br>lag 1: EDF=0.00, p=0.513<br>**lag 1: EDF=0.95, p=0.011**<br>**lag 1: EDF=0.86, p=0.033** | lag 1: EDF=2.64, p=0.183<br>**lag 6: EDF=0.80, p=0.025**<br>lag 3: EDF=0.43, p=0.308<br>**lag 1: EDF=0.80, p=0.032** | lag 7: EDF=0.87, p=0.067<br>lag 1: EDF=0.60, p=0.135<br>lag 0: EDF=0.30, p=0.420<br>lag 0: EDF=0.55, p=0.185 | **lag 0: EDF=3.83, p=0.005**<br>**lag 1: EDF=5.64, p=0.020**<br>**lag 5: EDF=2.64, p=0.027**<br>lag 6: EDF=4.99, p=0.107 | lag 0: EDF=0.75, p=0.096<br>lag 7: EDF=0.01, p=0.380<br>**lag 7: EDF=0.91, p=0.004**<br>lag 7: EDF=0.53, p=0.139 | **lag 5: EDF=6.44, p=0.035**<br>lag 7: EDF=0.76, p=0.130<br>lag 0: EDF=0.29, p=0.523<br>lag 6: EDF=0.84, p=0.094 |

**Table S14.** Overview of time lags, effect sizes (effective degree of freedom (EDF) or relative risk (95% confidence interval)) and p-values per human-bioclimatic region, atmospheric factor and subgroup (pink = female <60 years, orange = females ≥60 years, green = males <60 years, blue = males ≥60 years) for myocardial infarction (NSTEMI), significant p-values (≤ 0.05) are bold

| **Atmospheric factor** | **1.1** | **1.2** | **2.1** | **2.2** | **3.1** | **4.1** | **5.1** | **5.2** | **6.1** | **6.2** | **6.3** |
|---|---|---|---|---|---|---|---|---|---|---|---|
| Sea-level pressure (SLP) | lag 5: EDF=0.09, p=0.266<br>**lag 0: EDF=3.65, p=<0.001**<br>**lag 0: EDF=0.91, p=0.002**<br>lag 3: EDF=0.00, p=0.568 | lag 4: EDF=0.15, p=0.258<br>**lag 3: EDF=0.74, p=0.049**<br>**lag 0: EDF=0.92, p=0.001**<br>**lag 0: EDF=0.94, p=<0.001** | **lag 6: EDF=0.80, p=0.019**<br>lag 10: EDF=0.00, p=0.476<br>**lag 1: EDF=0.80, p=0.024**<br>lag 10: EDF=0.01, p=0.386 | **lag 1: EDF=0.83, p=0.013**<br>**lag 0: 1.00 (1.00–1.00), p=<0.001**<br>lag 6: EDF=0.00, p=0.524<br>**lag 3: EDF=0.94, p=<0.001** | **lag 3: 0.99 (0.99–0.99), p=<0.001**<br>lag 6: EDF=0.01, p=0.375<br>lag 4: EDF=0.01, p=0.307<br>lag 3: EDF=0.01, p=0.358 | lag 8: EDF=0.18, p=0.253<br>**lag 8: EDF=0.87, p=0.006**<br>lag 8: EDF=0.68, p=0.061<br>**lag 10: EDF=0.81, p=0.022** | lag 6: EDF=0.54, p=0.119<br>**lag 2: EDF=0.90, p=0.003**<br>**lag 10: EDF=0.80, p=0.023**<br>**lag 4: EDF=0.85, p=0.012** | lag 4: EDF=0.00, p=0.394<br>lag 1: EDF=0.46, p=0.159<br>lag 10: EDF=0.37, p=0.188<br>**lag 0: 1.00 (1.00–1.00), p=<0.001** | lag 10: EDF=3.92, p=0.159<br>**lag 0: EDF=0.77, p=0.030**<br>**lag 0: EDF=0.76, p=0.047**<br>**lag 0: EDF=0.83, p=0.013** | lag 9: EDF=0.03, p=0.283<br>**lag 1: EDF=0.89, p=0.004**<br>**lag 8: EDF=0.87, p=0.008**<br>**lag 0: EDF=0.93, p=<0.001** | **lag 3: EDF=2.62, p=0.001**<br>**lag 1: 1.00 (0.99–1.00), p=<0.001**<br>**lag 3: EDF=0.72, p=0.048**<br>lag 3: EDF=0.00, p=0.487 |
| Wind day | **lag 7: 1.12 (1.02–1.23), p=0.018**<br>lag 3: 0.98 (0.94–1.02), p=0.320<br>lag 1: 1.03 (0.98–1.09), p=0.274<br>lag 7: 1.04 (1.00–1.07), p=0.060 | lag 9: 1.06 (0.98–1.15), p=0.172<br>**lag 6: 0.94 (0.90–0.98), p=0.005**<br>**lag 6: 1.07 (1.02–1.12), p=0.007**<br>lag 6: 0.99 (0.96–1.03), p=0.674 | lag 8: 1.07 (0.98–1.18), p=0.126<br>**lag 7: 1.07 (1.03–1.11), p=<0.001**<br>lag 0: 1.03 (0.98–1.09), p=0.199<br>lag 7: 1.02 (0.99–1.06), p=0.159 | lag 4: 0.93 (0.85–1.03), p=0.158<br>lag 3: 0.97 (0.93–1.00), p=0.074<br>lag 3: 0.98 (0.93–1.04), p=0.542<br>lag 1: 1.01 (0.98–1.04), p=0.598 | lag 0: 1.08 (0.91–1.28), p=0.361<br>lag 3: 0.97 (0.90–1.04), p=0.384<br>**lag 7: 1.10 (1.00–1.20), p=0.046**<br>lag 0: 1.04 (0.98–1.10), p=0.209 | lag 4: 0.87 (0.74–1.02), p=0.082<br>lag 10: 1.03 (0.97–1.09), p=0.288<br>**lag 10: 1.09 (1.00–1.19), p=0.039**<br>lag 5: 1.03 (0.98–1.09), p=0.203 | lag 4: 1.09 (0.96–1.23), p=0.196<br>**lag 8: 1.07 (1.02–1.12), p=0.004**<br>lag 0: 0.94 (0.87–1.01), p=0.085<br>lag 5: 1.00 (0.96–1.04), p=0.880 | **lag 7: 0.75 (0.62–0.92), p=0.006**<br>**lag 4: 0.93 (0.87–0.99), p=0.021**<br>**lag 3: 0.89 (0.80–0.99), p=0.036**<br>lag 0: 1.03 (0.97–1.09), p=0.329 | **lag 7: 1.41 (1.08–1.83), p=0.011**<br>**lag 10: 1.10 (1.00–1.22), p=0.042**<br>**lag 6: 1.21 (1.03–1.42), p=0.018**<br>**lag 7: 1.09 (1.00–1.19), p=0.046** | lag 2: 0.95 (0.87–1.05), p=0.335<br>lag 7: 0.98 (0.95–1.02), p=0.321<br>lag 3: 0.95 (0.90–1.01), p=0.086<br>lag 1: 1.01 (0.98–1.05), p=0.412 | lag 0: 0.85 (0.69–1.05), p=0.139<br>lag 6: 0.95 (0.88–1.03), p=0.216<br>**lag 6: 1.15 (1.03–1.27), p=0.011**<br>**lag 3: 1.09 (1.02–1.17), p=0.015** |
| Frost day | **lag 6: 0.92 (0.85–0.98), p=0.012**<br>lag 2: 1.00 (0.98– | lag 8: 1.05 (0.99–1.11), p=0.132<br>lag 7: 1.03 (1.00– | lag 7: 1.05 (0.99–1.11), p=0.092<br>lag 7: 1.02 (1.00– | lag 10: 1.00 (0.95–1.06), p=0.901<br>**lag 5: 1.04 (1.02–** | lag 0: 1.05 (0.94–1.16), p=0.393<br>lag 6: 1.03 (0.99– | **lag 10: 1.11 (1.01–1.22), p=0.024**<br>**lag 9: 1.04 (1.00–** | lag 4: 0.93 (0.85–1.01), p=0.103<br>lag 9: 1.02 (0.99– | lag 7: 0.92 (0.84–1.00), p=0.056<br>**lag 2: 1.05 (1.01–** | lag 0: 0.83 (0.68–1.01), p=0.062<br>lag 5: 1.03 (0.96– | lag 5: 1.04 (0.98–1.10), p=0.188<br>**lag 9: 1.02 (1.00–** | lag 9: 1.11 (0.98–1.26), p=0.095<br>**lag 8: 1.06 (1.01–** |

| Atmospheric factor | 1.1 | 1.2 | 2.1 | 2.2 | 3.1 | 4.1 | 5.1 | 5.2 | 6.1 | 6.2 | 6.3 |
|---|---|---|---|---|---|---|---|---|---|---|---|
| | 1.03), p=0.770<br>lag 6: 1.01 (0.97–1.05), p=0.588<br>lag 7: 1.01 (0.98–1.03), p=0.508 | 1.06), p=0.054<br>lag 7: 1.01 (0.98–1.05), p=0.529<br>**lag 7: 1.03 (1.00–1.05), p=0.034** | 1.04), p=0.094<br>lag 7: 0.98 (0.95–1.02), p=0.334<br>lag 7: 1.01 (0.99–1.03), p=0.468 | **1.07), p=0.001**<br>lag 8: 1.02 (0.98–1.06), p=0.353<br>**lag 7: 1.04 (1.01–1.06), p=<0.001** | 1.07), p=0.186<br>lag 9: 1.01 (0.95–1.07), p=0.803<br>lag 0: 1.01 (0.97–1.04), p=0.713 | **1.09), p=0.038**<br>**lag 1: 1.10 (1.04–1.17), p=0.001**<br>**lag 3: 1.05 (1.02–1.09), p=0.004** | 1.05), p=0.157<br>lag 8: 1.02 (0.97–1.07), p=0.534<br>lag 2: 1.02 (0.99–1.04), p=0.209 | **1.09), p=0.006**<br>lag 9: 1.02 (0.96–1.08), p=0.544<br>lag 9: 1.02 (0.99–1.05), p=0.285 | 1.10), p=0.458<br>**lag 8: 1.15 (1.03–1.28), p=0.011**<br>**lag 3: 1.10 (1.05–1.16), p=<0.001** | **1.04), p=0.039**<br>lag 9: 1.02 (0.99–1.06), p=0.217<br>**lag 3: 1.02 (1.00–1.04), p=0.019** | **1.11), p=0.016**<br>lag 9: 1.03 (0.96–1.10), p=0.409<br>**lag 8: 1.07 (1.03–1.12), p=0.002** |
| Wind chill index | lag 1: EDF=0.01, p=0.392<br>lag 0: EDF=0.00, p=0.568<br>**lag 3: EDF=0.76, p=0.040**<br>lag 0: EDF=0.00, p=0.926 | lag 8: EDF=0.40, p=0.172<br>**lag 6: EDF=0.81, p=0.024**<br>lag 7: EDF=0.58, p=0.102<br>lag 0: EDF=0.00, p=0.910 | **lag 1: EDF=0.78, p=0.026**<br>**lag 6: EDF=0.85, p=0.015**<br>**lag 6: EDF=0.74, p=0.042**<br>**lag 0: EDF=0.93, p=0.004** | lag 7: EDF=0.00, p=0.649<br>**lag 7: EDF=0.92, p=0.002**<br>**lag 9: EDF=2.90, p=<0.001**<br>**lag 0: EDF=0.88, p=0.010** | lag 0: EDF=0.00, p=0.988<br>lag 7: EDF=0.48, p=0.149<br>**lag 6: EDF=1.87, p=0.003**<br>lag 4: EDF=0.02, p=0.300 | lag 10: EDF=0.00, p=0.371<br>lag 2: EDF=0.49, p=0.147<br>lag 0: EDF=0.00, p=0.595<br>lag 2: EDF=0.00, p=0.325 | **lag 6: EDF=0.77, p=0.030**<br>lag 0: EDF=0.00, p=0.543<br>**lag 8: EDF=0.88, p=0.006**<br>lag 1: EDF=0.12, p=0.280 | lag 3: EDF=0.00, p=0.725<br>lag 3: EDF=0.00, p=0.527<br>**lag 3: EDF=0.86, p=0.009**<br>**lag 6: EDF=0.80, p=0.024** | **lag 10: 0.97 (0.96–0.99), p=<0.001**<br>lag 5: EDF=0.62, p=0.068<br>lag 8: EDF=0.23, p=0.253<br>lag 3: EDF=0.00, p=0.394 | lag 0: EDF=0.00, p=0.871<br>lag 0: EDF=0.02, p=0.322<br>lag 6: EDF=0.68, p=0.075<br>lag 0: EDF=0.67, p=0.082 | lag 4: EDF=0.40, p=0.217<br>lag 8: EDF=0.00, p=0.651<br>lag 10: EDF=0.00, p=0.669<br>lag 6: EDF=0.00, p=0.419 |
| Heat wave | lag 1: 0.96 (0.85–1.09), p=0.575<br>**lag 0: 0.93 (0.88–0.99), p=0.013**<br>lag 5: 0.94 (0.88–1.00), p=0.069<br>lag 0: 0.95 (0.90–1.02), p=0.138 | lag 8: 1.00 (0.91–1.10), p=0.978<br>lag 0: 0.98 (0.92–1.04), p=0.454<br>lag 0: 0.98 (0.92–1.03), p=0.421<br>**lag 0: 0.92 (0.88–0.97), p=<0.001** | lag 2: 1.02 (0.90–1.15), p=0.785<br>lag 2: 0.97 (0.93–1.01), p=0.191<br>**lag 8: 0.92 (0.87–0.98), p=0.009**<br>**lag 0: 0.93 (0.89–0.98), p=0.008** | lag 9: 0.93 (0.85–1.03), p=0.178<br>**lag 0: 0.92 (0.88–0.96), p=<0.001**<br>lag 1: 0.97 (0.92–1.03), p=0.318<br>**lag 0: 0.92 (0.88–0.96), p=<0.001** | lag 7: 0.83 (0.67–1.02), p=0.071<br>**lag 2: 0.88 (0.82–0.96), p=0.002**<br>lag 7: 0.97 (0.87–1.08), p=0.568<br>lag 0: 0.94 (0.87–1.01), p=0.100 | lag 6: 0.93 (0.79–1.09), p=0.360<br>lag 0: 0.98 (0.91–1.05), p=0.548<br>lag 6: 1.06 (0.93–1.20), p=0.399<br>**lag 0: 0.93 (0.88–0.99), p=0.030** | lag 5: 0.91 (0.78–1.06), p=0.237<br>lag 3: 0.97 (0.90–1.04), p=0.354<br>lag 1: 0.96 (0.88–1.05), p=0.418<br>**lag 0: 0.92 (0.87–0.98), p=0.007** | lag 3: 0.90 (0.73–1.10), p=0.303<br>lag 0: 1.01 (0.92–1.10), p=0.851<br>lag 9: 0.89 (0.79–1.00), p=0.057<br>lag 0: 0.94 (0.87–1.02), p=0.115 | lag 3: 1.26 (0.89–1.77), p=0.191<br>**lag 7: 0.88 (0.78–0.98), p=0.027**<br>lag 8: 0.87 (0.71–1.07), p=0.189<br>lag 2: 1.02 (0.90–1.16), p=0.755 | lag 0: 1.05 (0.92–1.20), p=0.458<br>lag 0: 0.96 (0.91–1.02), p=0.162<br>lag 0: 0.97 (0.91–1.03), p=0.293<br>**lag 0: 0.94 (0.89–0.99), p=0.017** | **lag 8: 0.63 (0.46–0.87), p=0.006**<br>**lag 0: 0.86 (0.78–0.94), p=0.001**<br>**lag 0: 0.85 (0.72–0.99), p=0.041**<br>**lag 1: 0.90 (0.82–0.98), p=0.022** |

| **Atmospheric factor** | **1.1** | **1.2** | **2.1** | **2.2** | **3.1** | **4.1** | **5.1** | **5.2** | **6.1** | **6.2** | **6.3** |
|---|---|---|---|---|---|---|---|---|---|---|---|
| Heat stress wave duration | **lag 1: EDF=0.82, p=0.019**<br>lag 3: EDF=0.45, p=0.168<br>**lag 0: EDF=0.74, p=0.048** | lag 0: EDF=0.00, p=0.325<br>lag 1: EDF=0.12, p=0.280 | **lag 7: EDF=0.75, p=0.035**<br>lag 0: EDF=0.31, p=0.224<br>lag 7: EDF=0.00, p=0.729<br>lag 0: EDF=0.65, p=0.086 | lag 8: EDF=0.00, p=0.508<br>lag 0: EDF=0.00, p=0.757<br>lag 5: EDF=0.00, p=0.476 | lag 6: EDF=0.00, p=0.589<br>lag 2: EDF=0.00, p=0.730<br>lag 7: EDF=0.00, p=0.838<br>lag 1: EDF=0.59, p=0.111 | lag 8: EDF=0.00, p=0.660<br>lag 2: EDF=0.45, p=0.165<br>lag 6: EDF=0.65, p=0.077 | lag 9: EDF=0.31, p=0.209<br>lag 3: EDF=0.62, p=0.101<br>lag 0: EDF=0.40, p=0.191 | lag 10: EDF=0.39, p=0.184<br>lag 7: EDF=0.00, p=0.500 | lag 0: EDF=0.00, p=0.463<br>lag 4: EDF=0.00, p=0.611<br>lag 8: EDF=0.00, p=0.478<br>lag 1: EDF=0.00, p=0.999 | lag 0: EDF=0.63, p=0.096<br>lag 6: EDF=0.38, p=0.185<br>**lag 0: EDF=0.76, p=0.037** | **lag 8: EDF=0.86, p=0.009**<br>lag 0: EDF=0.00, p=0.838<br>lag 6: EDF=0.61, p=0.094<br>lag 1: EDF=0.00, p=0.812 |
| Heat stress wave intensity | lag 0: EDF=0.37, p=0.185<br>**lag 1: EDF=0.79, p=0.026** | **lag 4: EDF=0.79, p=0.023**<br>**lag 0: EDF=0.88, p=0.004**<br>lag 3: EDF=0.00, p=0.973<br>lag 0: EDF=0.27, p=0.235 | **lag 1: EDF=0.80, p=0.018**<br>lag 9: EDF=0.00, p=0.642 | lag 6: EDF=0.00, p=0.421<br>lag 0: EDF=0.35, p=0.207 | lag 1: EDF=0.00, p=0.534 | lag 6: EDF=0.00, p=0.970<br>lag 0: EDF=0.00, p=0.754<br>lag 8: EDF=0.00, p=0.676<br>lag 0: EDF=0.00, p=0.472 | lag 7: EDF=0.00, p=0.667<br>**lag 0: EDF=0.77, p=0.036**<br>lag 0: EDF=0.66, p=0.080 | lag 6: EDF=0.30, p=0.219<br>**lag 0: EDF=0.79, p=0.026**<br>lag 9: EDF=0.00, p=0.451<br>lag 0: EDF=0.53, p=0.138 | lag 3: EDF=0.47, p=0.257<br>lag 0: EDF=0.00, p=0.458<br>**lag 2: EDF=0.80, p=0.020** | **lag 0: EDF=0.85, p=0.006**<br>lag 5: EDF=0.16, p=0.244 | lag 6: EDF=0.10, p=0.311<br>**lag 2: EDF=0.84, p=0.006**<br>lag 0: EDF=0.52, p=0.134 |
| Particulate matter ($PM_{10}$) | **lag 4: EDF=0.85, p=0.007**<br>**lag 1: EDF=0.82, p=0.018**<br>**lag 0: EDF=0.93, p=<0.001**<br>lag 0: EDF=0.15, p=0.270 | **lag 6: EDF=0.85, p=0.007**<br>**lag 0: EDF=0.85, p=0.008**<br>**lag 5: EDF=0.75, p=0.041**<br>**lag 0: EDF=0.88, p=0.004** | lag 0: EDF=0.00, p=0.629<br>**lag 1: EDF=0.78, p=0.033**<br>lag 3: EDF=0.00, p=0.461<br>lag 0: EDF=0.00, p=0.785 | lag 4: EDF=0.01, p=0.323<br>**lag 0: EDF=0.87, p=0.005**<br>lag 0: EDF=0.00, p=0.815<br>lag 0: EDF=0.41, p=0.186 | lag 7: EDF=0.11, p=0.282<br>lag 0: EDF=0.10, p=0.273<br>lag 2: EDF=0.57, p=0.108<br>lag 0: EDF=0.50, p=0.144 | lag 0: EDF=0.68, p=0.067<br>lag 7: EDF=0.72, p=0.051<br>**lag 4: EDF=0.77, p=0.029**<br>lag 3: EDF=0.69, p=0.066 | lag 3: EDF=0.21, p=0.228<br>lag 4: EDF=0.34, p=0.213<br>lag 5: EDF=0.00, p=0.564<br>lag 6: EDF=0.00, p=0.784 | lag 2: EDF=0.35, p=0.217<br>lag 1: EDF=0.00, p=0.977<br>lag 4: EDF=0.01, p=0.302<br>lag 1: EDF=0.01, p=0.341 | lag 3: EDF=0.66, p=0.162<br>lag 2: EDF=0.00, p=0.663<br>**lag 2: EDF=4.00, p=0.015**<br>**lag 1: EDF=0.80, p=0.018** | lag 4: EDF=0.00, p=0.839<br>lag 4: EDF=0.01, p=0.355<br>lag 0: EDF=0.00, p=0.395<br>lag 2: EDF=0.00, p=0.765 | **lag 7: EDF=0.79, p=0.035**<br>lag 4: EDF=0.22, p=0.228<br>lag 6: EDF=0.00, p=0.953<br>lag 4: EDF=0.32, p=0.202 |

**Table S15.** Overview of time lags, effect sizes (effective degree of freedom (EDF) or relative risk (95% confidence interval)) and p-values per human-bioclimatic region, atmospheric factor and subgroup (pink = female <60 years, orange = females ≥60 years, green = males <60 years, blue = males ≥60 years) for non subdural hemorrhage, significant p-values (≤ 0.05) are bold

| Atmospheric factor | 1.1 | 1.2 | 2.1 | 2.2 | 3.1 | 4.1 | 5.1 | 5.2 | 6.1 | 6.2 | 6.3 |
|---|---|---|---|---|---|---|---|---|---|---|---|
| Sea-level pressure (SLP) | **lag 6: EDF=0.85, p=0.024**<br>**lag 8: EDF=0.85, p=0.010**<br>**lag 3: EDF=0.90, p=0.009**<br>lag 0: EDF=0.25, p=0.250 | lag 0: EDF=0.33, p=0.244<br>lag 9: EDF=0.00, p=0.798<br>**lag 3: EDF=0.85, p=0.015**<br>lag 9: EDF=0.00, p=0.978 | **lag 4: EDF=0.87, p=0.009**<br>lag 0: EDF=0.46, p=0.147<br>**lag 2: EDF=0.85, p=0.013**<br>**lag 6: EDF=0.82, p=0.015** | lag 2: EDF=0.70, p=0.086<br>lag 10: EDF=0.28, p=0.212<br>lag 3: EDF=0.01, p=0.342<br>lag 0: EDF=0.46, p=0.153 | lag 7: EDF=0.00, p=0.934<br>lag 10: EDF=0.50, p=0.172<br>**lag 2: EDF=0.90, p=0.022**<br>lag 6: EDF=0.00, p=0.430 | lag 8: EDF=0.00, p=0.556<br>lag 9: EDF=0.00, p=0.461<br>lag 2: EDF=0.64, p=0.201<br>lag 9: EDF=0.52, p=0.183 | lag 4: EDF=0.52, p=0.233<br>lag 0: EDF=0.48, p=0.161<br>lag 7: EDF=0.00, p=0.530<br>lag 5: EDF=0.64, p=0.093 | **lag 0: EDF=0.92, p=0.021**<br>lag 2: EDF=0.76, p=0.058<br>**lag 1: EDF=3.28, p=<0.001**<br>**lag 1: EDF=0.86, p=0.017** | lag 7: EDF=3.22, p=0.205<br>lag 1: EDF=0.71, p=0.173<br>**lag 10: 0.98 (0.96–0.99), p=0.006**<br>**lag 1: EDF=0.88, p=0.050** | lag 10: EDF=0.33, p=0.247<br>lag 5: EDF=0.00, p=0.436<br>lag 9: EDF=0.27, p=0.252<br>**lag 8: EDF=0.82, p=0.013** | lag 1: EDF=0.00, p=0.629<br>**lag 7: EDF=0.92, p=0.009**<br>lag 4: EDF=2.11, p=0.067<br>lag 2: EDF=1.87, p=0.140 |
| Sea-level pressure difference (SLPD) | **lag 4: 0.80 (0.67–0.95), p=0.013**<br>lag 10: 1.12 (0.99–1.26), p=0.066<br>**lag 7: 1.20 (1.01–1.42), p=0.035**<br>lag 9: 1.13 (1.00–1.29), p=0.055 | **lag 2: 1.18 (1.01–1.36), p=0.033**<br>**lag 3: 1.12 (1.01–1.25), p=0.036**<br>lag 8: 1.09 (0.95–1.25), p=0.234<br>lag 0: 0.90 (0.81–1.01), p=0.084 | lag 6: 1.15 (0.96–1.37), p=0.130<br>**lag 10: 0.89 (0.80–0.99), p=0.027**<br>lag 5: 0.91 (0.77–1.07), p=0.240<br>lag 3: 0.90 (0.80–1.01), p=0.073 | lag 8: 1.17 (0.98–1.39), p=0.089<br>**lag 2: 0.81 (0.73–0.90), p=<0.001**<br>lag 7: 0.88 (0.74–1.03), p=0.119<br>lag 7: 0.90 (0.80–1.01), p=0.077 | lag 7: 0.79 (0.57–1.08), p=0.139<br>**lag 6: 0.78 (0.65–0.94), p=0.008**<br>lag 6: 1.17 (0.88–1.57), p=0.283<br>lag 5: 0.90 (0.73–1.11), p=0.315 | lag 0: 1.24 (0.97–1.59), p=0.085<br>lag 10: 1.15 (0.99–1.34), p=0.071<br>lag 1: 1.09 (0.86–1.37), p=0.482<br>lag 5: 1.16 (0.99–1.36), p=0.063 | lag 0: 0.81 (0.65–1.02), p=0.077<br>**lag 10: 1.14 (1.00–1.30), p=0.044**<br>lag 5: 0.82 (0.66–1.02), p=0.077<br>lag 9: 1.09 (0.95–1.25), p=0.206 | lag 5: 1.23 (0.91–1.66), p=0.188<br>**lag 1: 0.81 (0.67–0.97), p=0.022**<br>lag 4: 0.79 (0.59–1.04), p=0.093<br>lag 1: 0.90 (0.74–1.08), p=0.258 | lag 5: 1.48 (0.84–2.58), p=0.172<br>lag 5: 0.85 (0.66–1.11), p=0.237<br>**lag 1: 1.69 (1.06–2.71), p=0.028**<br>lag 8: 0.81 (0.61–1.07), p=0.141 | lag 4: 0.89 (0.76–1.05), p=0.162<br>**lag 3: 1.10 (1.01–1.20), p=0.035**<br>**lag 1: 1.17 (1.02–1.35), p=0.026**<br>lag 1: 0.92 (0.84–1.01), p=0.071 | **lag 3: 1.42 (1.02–1.96), p=0.035**<br>lag 7: 0.89 (0.72–1.10), p=0.272<br>lag 3: 1.10 (0.79–1.51), p=0.580<br>**lag 5: 1.38 (1.11–1.73), p=0.005** |
| Wind day | lag 0: 0.91 (0.71–1.18), p=0.495<br>lag 0: 1.08 (0.93– | **lag 6: 1.28 (1.06–1.53), p=0.009**<br>lag 10: 1.07 (0.93– | **lag 7: 0.81 (0.66–1.00), p=0.045**<br>lag 7: 1.08 (0.97– | lag 3: 0.88 (0.71–1.09), p=0.236<br>lag 8: 1.10 (0.98– | lag 6: 1.25 (0.92–1.70), p=0.159<br>lag 5: 0.90 (0.73– | **lag 8: 1.43 (1.07–1.91), p=0.017**<br>lag 6: 1.21 (1.00– | lag 7: 1.20 (0.93–1.55), p=0.152<br>**lag 8: 1.17 (1.01–** | **lag 7: 1.52 (1.12–2.07), p=0.007**<br>lag 3: 0.79 (0.62– | **lag 10: 2.91 (1.77–4.77), p=<0.001**<br>lag 2: 1.29 (0.95– | lag 10: 1.18 (0.98–1.43), p=0.073<br>**lag 2: 0.87 (0.78–** | **lag 9: 2.25 (1.62–3.13), p=<0.001**<br>**lag 9: 1.49 (1.17–** |

| Atmospheric factor | 1.1 | 1.2 | 2.1 | 2.2 | 3.1 | 4.1 | 5.1 | 5.2 | 6.1 | 6.2 | 6.3 |
|---|---|---|---|---|---|---|---|---|---|---|---|
| | 1.25), p=0.318<br>**lag 10: 0.72 (0.56–0.93), p=0.011**<br>lag 2: 0.89 (0.75–1.06), p=0.205 | 1.23), p=0.353<br>**lag 2: 1.23 (1.03–1.47), p=0.020**<br>**lag 10: 1.25 (1.08–1.43), p=0.002** | 1.21), p=0.163<br>lag 9: 1.17 (0.99–1.39), p=0.069<br>**lag 2: 1.14 (1.01–1.29), p=0.028** | 1.23), p=0.114<br>lag 4: 1.09 (0.91–1.31), p=0.334<br>lag 0: 1.09 (0.96–1.23), p=0.188 | 1.10), p=0.292<br>**lag 4: 0.62 (0.42–0.90), p=0.013**<br>lag 7: 0.82 (0.64–1.04), p=0.103 | 1.46), p=0.053<br>lag 3: 1.31 (0.98–1.74), p=0.066<br>lag 4: 0.79 (0.62–1.01), p=0.059 | **1.35), p=0.036**<br>**lag 0: 0.72 (0.53–0.98), p=0.035**<br>lag 8: 1.15 (0.99–1.35), p=0.072 | 1.00), p=0.050<br>**lag 9: 1.44 (1.08–1.92), p=0.014**<br>lag 10: 1.15 (0.93–1.41), p=0.200 | 1.77), p=0.107<br>**lag 5: 2.17 (1.30–3.62), p=0.003**<br>**lag 3: 0.49 (0.29–0.83), p=0.007** | **0.98), p=0.018**<br>lag 8: 1.11 (0.94–1.32), p=0.222<br>lag 10: 1.08 (0.97–1.20), p=0.167 | **1.89), p=0.001**<br>lag 5: 1.30 (0.88–1.92), p=0.182<br>lag 10: 1.27 (0.96–1.67), p=0.096 |
| Tropical night | **lag 2: 2.61 (1.07–6.36), p=0.035**<br>lag 2: 0.26 (0.03–2.09), p=0.205<br>lag 6: 2.54 (0.95–6.82), p=0.065<br>lag 9: 1.86 (0.87–3.97), p=0.110 | lag 10: 0.44 (0.09–2.05), p=0.294<br>lag 8: 1.48 (0.93–2.36), p=0.101<br>**lag 10: 2.36 (1.43–3.89), p=<0.001**<br>lag 7: 1.50 (0.91–2.47), p=0.115 | **lag 3: 3.38 (1.37–8.32), p=0.008**<br>**lag 0: 2.90 (1.29–6.52), p=0.010**<br>lag 2: 2.80 (0.98–8.01), p=0.055<br>lag 6: 1.54 (0.61–3.89), p=0.366 | **lag 9: 2.24 (1.30–3.86), p=0.004**<br>lag 1: 0.59 (0.30–1.14), p=0.117<br>**lag 5: 1.84 (1.08–3.12), p=0.024**<br>lag 3: 0.80 (0.46–1.37), p=0.412 | - | **lag 10: 20.16 (4.06–100.12), p=<0.001**<br>lag 2: 1.67 (0.37–7.63), p=0.507<br>lag 1: 4.62 (0.57–37.70), p=0.154<br>lag 2: 4.12 (0.85–19.90), p=0.078 | lag 5: 4.45 (0.45–44.43), p=0.204<br>lag 7: 2.89 (0.87–9.58), p=0.083<br>**lag 3: 5.55 (1.54–19.94), p=0.009**<br>lag 1: 3.53 (1.00–12.48), p=0.050 | lag 2: 1.64 (0.17–16.30), p=0.671<br>lag 4: 2.45 (0.72–8.32), p=0.150<br>lag 3: 2.07 (0.24–17.80), p=0.509<br>lag 9: 1.47 (0.33–6.42), p=0.611 | **lag 8: 8.98 (1.51–53.30), p=0.016**<br>lag 4: 3.22 (0.35–29.63), p=0.301<br>lag 1: 4.60 (0.53–40.00), p=0.167<br>lag 8: 2.59 (0.57–11.79), p=0.219 | lag 9: 1.25 (0.61–2.56), p=0.536<br>**lag 1: 1.63 (1.12–2.38), p=0.012**<br>lag 5: 0.27 (0.06–1.20), p=0.085<br>**lag 10: 0.45 (0.21–0.96), p=0.040** | **lag 0: 5.03 (1.06–23.95), p=0.043**<br>**lag 9: 3.21 (1.19–8.64), p=0.021**<br>lag 10: 1.36 (0.15–12.00), p=0.780<br>lag 2: 2.42 (0.70–8.38), p=0.162 |
| Hot day | **lag 4: 1.65 (1.10–2.47), p=0.016**<br>lag 1: 0.92 (0.62–1.38), p=0.692<br>lag 2: 0.75 (0.45–1.27), p=0.284 | **lag 10: 0.61 (0.37–1.00), p=0.049**<br>**lag 9: 1.28 (1.02–1.61), p=0.030**<br>lag 3: 0.99 (0.67–1.46), p=0.962 | lag 3: 1.30 (0.99–1.70), p=0.059<br>lag 1: 0.88 (0.69–1.12), p=0.292<br>lag 1: 0.76 (0.55–1.06), p=0.110 | lag 7: 0.83 (0.58–1.20), p=0.329<br>lag 0: 0.90 (0.72–1.13), p=0.383<br>lag 2: 0.98 (0.69–1.38), p=0.887 | lag 5: 1.51 (0.85–2.68), p=0.161<br>lag 5: 0.73 (0.42–1.25), p=0.252<br>**lag 0: 0.11 (0.01–0.94), p=0.044** | lag 7: 2.20 (0.66–7.32), p=0.199<br>lag 3: 2.03 (1.00–4.16), p=0.052<br>lag 1: 1.84 (0.56–6.05), p=0.318 | lag 7: 0.64 (0.28–1.49), p=0.299<br>lag 0: 0.70 (0.45–1.10), p=0.125<br>lag 6: 0.52 (0.24–1.13), p=0.097 | lag 3: 1.30 (0.66–2.58), p=0.452<br>lag 0: 0.58 (0.31–1.07), p=0.079<br>lag 10: 0.59 (0.24–1.46), p=0.254 | lag 8: 1.91 (0.49–7.38), p=0.351<br>lag 4: 0.35 (0.10–1.28), p=0.113<br>lag 4: 1.88 (0.62–5.65), p=0.263 | lag 6: 0.72 (0.48–1.07), p=0.105<br>lag 0: 0.95 (0.78–1.16), p=0.604<br>lag 3: 0.78 (0.54–1.12), p=0.180 | **lag 6: 2.76 (1.51–5.02), p=<0.001**<br>lag 6: 0.53 (0.27–1.04), p=0.066<br>lag 7: 0.40 (0.10–1.55), p=0.186 |

| Atmospheric factor | 1.1 | 1.2 | 2.1 | 2.2 | 3.1 | 4.1 | 5.1 | 5.2 | 6.1 | 6.2 | 6.3 |
|---|---|---|---|---|---|---|---|---|---|---|---|
| | **lag 7: 0.65 (0.44–0.97), p=0.035** | **lag 6: 0.54 (0.39–0.76), p=<0.001** | lag 10: 0.88 (0.72–1.07), p=0.197 | lag 0: 0.83 (0.65–1.05), p=0.115 | lag 0: 0.68 (0.37–1.25), p=0.216 | lag 6: 1.53 (0.62–3.76), p=0.353 | lag 1: 0.95 (0.62–1.46), p=0.806 | **lag 5: 1.46 (1.00–2.13), p=0.050** | lag 10: 1.44 (0.72–2.86), p=0.303 | lag 4: 0.85 (0.67–1.07), p=0.171 | lag 1: 0.64 (0.32–1.28), p=0.203 |
| Summer day | lag 1: 0.94 (0.73–1.22), p=0.646<br>lag 1: 0.87 (0.72–1.04), p=0.123<br>lag 9: 0.86 (0.69–1.07), p=0.177<br>**lag 2: 0.81 (0.68–0.97), p=0.022** | lag 0: 0.85 (0.70–1.02), p=0.074<br>lag 5: 0.88 (0.77–1.00), p=0.051<br>**lag 0: 0.83 (0.71–0.97), p=0.020**<br>**lag 0: 0.83 (0.71–0.96), p=0.013** | lag 1: 0.90 (0.77–1.05), p=0.170<br>lag 0: 0.96 (0.87–1.07), p=0.509<br>lag 4: 0.90 (0.78–1.04), p=0.156<br>lag 0: 0.92 (0.82–1.03), p=0.163 | **lag 10: 0.78 (0.66–0.92), p=0.003**<br>lag 0: 0.92 (0.83–1.03), p=0.152<br>lag 4: 0.90 (0.78–1.04), p=0.159<br>lag 0: 0.91 (0.81–1.03), p=0.138 | lag 10: 1.11 (0.83–1.48), p=0.487<br>**lag 0: 0.81 (0.67–0.97), p=0.022**<br>lag 2: 0.84 (0.59–1.21), p=0.357<br>lag 1: 0.99 (0.81–1.22), p=0.930 | **lag 9: 0.44 (0.22–0.88), p=0.020**<br>lag 5: 1.11 (0.84–1.47), p=0.473<br>lag 4: 0.71 (0.42–1.21), p=0.212<br>lag 2: 0.81 (0.53–1.23), p=0.321 | lag 0: 0.87 (0.65–1.17), p=0.352<br>lag 2: 0.86 (0.71–1.04), p=0.116<br>**lag 0: 1.41 (1.09–1.81), p=0.008**<br>lag 2: 0.95 (0.78–1.15), p=0.608 | lag 9: 0.73 (0.51–1.03), p=0.073<br>lag 0: 0.98 (0.76–1.26), p=0.863<br>lag 3: 0.79 (0.59–1.07), p=0.132<br>lag 0: 0.83 (0.66–1.04), p=0.100 | **lag 2: 0.37 (0.15–0.91), p=0.030**<br>lag 0: 0.91 (0.64–1.28), p=0.580<br>lag 8: 0.69 (0.29–1.60), p=0.383<br>**lag 2: 0.62 (0.39–0.96), p=0.034** | **lag 6: 0.84 (0.71–0.99), p=0.040**<br>lag 0: 0.94 (0.85–1.04), p=0.260<br>**lag 2: 0.85 (0.73–0.99), p=0.038**<br>lag 0: 1.07 (0.95–1.20), p=0.284 | lag 10: 0.68 (0.41–1.14), p=0.147<br>lag 0: 0.89 (0.70–1.13), p=0.321<br>lag 7: 0.77 (0.48–1.22), p=0.266<br>lag 0: 0.78 (0.58–1.03), p=0.083 |
| Wet-bulb globe temperature | - | lag 0: EDF=0.66, p=0.100<br>**lag 0: EDF=0.88, p=0.004** | - | **lag 0: EDF=0.89, p=0.003** | lag 0: EDF=0.00, p=0.415 | lag 9: EDF=0.00, p=0.812 | - | **lag 9: EDF=4.39, p=0.017** | lag 8: EDF=0.00, p=0.727 | - | lag 0: EDF=0.00, p=0.353<br>lag 9: EDF=0.81, p=0.080 |
| Daily mean temperature | - | **lag 0: EDF=1.08, p=<0.001** | **lag 0: EDF=0.93, p=0.001** | - | - | lag 0: EDF=0.60, p=0.133<br>lag 1: EDF=0.71, p=0.078 | lag 0: EDF=0.73, p=0.116<br>**lag 0: EDF=0.90, p=0.015** | lag 9: EDF=0.69, p=0.195<br>**lag 0: EDF=2.38, p=0.007**<br>**lag 0: EDF=0.84, p=0.022** | lag 7: EDF=0.80, p=0.115 | **lag 0: 0.98 (0.98–0.99), p=<0.001** | lag 8: EDF=0.79, p=0.169<br>lag 3: EDF=0.68, p=0.250 |

| Atmospheric factor | 1.1 | 1.2 | 2.1 | 2.2 | 3.1 | 4.1 | 5.1 | 5.2 | 6.1 | 6.2 | 6.3 |
|---|---|---|---|---|---|---|---|---|---|---|---|
| Wind chill index | **lag 0: EDF=0.94, p=0.005**<br>**lag 0: EDF=0.84, p=0.013**<br>lag 3: EDF=0.42, p=0.220<br>lag 0: EDF=0.71, p=0.063 | lag 3: EDF=0.00, p=0.521 | **lag 0: EDF=0.77, p=0.045**<br>**lag 0: 0.98 (0.97–0.99), p=<0.001**<br>lag 7: EDF=0.62, p=0.103 | lag 9: EDF=0.00, p=0.259<br>lag 0: EDF=0.29, p=0.227 | - | **lag 0: EDF=0.89, p=0.030** | **lag 0: EDF=0.78, p=0.032**<br>**lag 0: EDF=2.61, p=<0.001** | - | lag 0: EDF=0.27, p=0.385<br>**lag 9: EDF=5.87, p=0.040** | lag 2: EDF=0.01, p=0.371<br>lag 0: EDF=0.00, p=0.516<br>lag 0: EDF=0.50, p=0.167<br>**lag 0: EDF=3.01, p=<0.001** | - |
| Heat wave | lag 0: 0.82 (0.57–1.17), p=0.277<br>lag 0: 0.94 (0.74–1.20), p=0.624<br>lag 3: 0.73 (0.51–1.03), p=0.076<br>lag 2: 0.79 (0.62–1.01), p=0.056 | lag 6: 0.95 (0.73–1.24), p=0.699<br>lag 5: 0.84 (0.69–1.01), p=0.062<br>**lag 2: 0.69 (0.52–0.91), p=0.008**<br>lag 1: 0.95 (0.78–1.15), p=0.602 | lag 10: 1.21 (1.00–1.46), p=0.053<br>lag 0: 1.04 (0.82–1.32), p=0.755<br>lag 9: 0.93 (0.71–1.22), p=0.599<br>lag 0: 1.02 (0.87–1.19), p=0.801 | lag 6: 0.97 (0.71–1.33), p=0.853<br>**lag 0: 0.72 (0.61–0.86), p=<0.001**<br>lag 0: 0.85 (0.60–1.19), p=0.332<br>lag 0: 0.96 (0.81–1.14), p=0.644 | lag 10: 1.07 (0.74–1.54), p=0.724<br>lag 1: 0.90 (0.70–1.16), p=0.434<br>lag 0: 1.10 (0.68–1.78), p=0.692<br>lag 8: 0.85 (0.66–1.10), p=0.229 | lag 8: 0.82 (0.49–1.36), p=0.436<br>lag 4: 1.14 (0.89–1.47), p=0.312<br>lag 3: 0.92 (0.59–1.44), p=0.723<br>lag 0: 0.74 (0.49–1.10), p=0.133 | lag 3: 0.82 (0.54–1.24), p=0.351<br>lag 0: 0.84 (0.62–1.13), p=0.255<br>**lag 0: 0.67 (0.47–0.96), p=0.028**<br>lag 0: 1.04 (0.81–1.35), p=0.740 | lag 0: 0.85 (0.53–1.35), p=0.489<br>lag 10: 0.96 (0.75–1.23), p=0.744<br>lag 8: 0.70 (0.41–1.19), p=0.185<br>**lag 9: 1.37 (1.05–1.81), p=0.023** | lag 7: 2.06 (0.41–10.34), p=0.382<br>lag 0: 0.93 (0.59–1.48), p=0.773<br>lag 7: 1.62 (0.27–9.82), p=0.599<br>lag 0: 1.09 (0.63–1.87), p=0.757 | **lag 9: 1.34 (1.08–1.65), p=0.007**<br>**lag 9: 0.85 (0.74–0.97), p=0.013**<br>lag 1: 1.12 (0.86–1.45), p=0.409<br>lag 2: 0.90 (0.76–1.06), p=0.218 | lag 7: 0.60 (0.21–1.71), p=0.339<br>lag 8: 0.87 (0.60–1.25), p=0.440<br>lag 5: 0.98 (0.47–2.02), p=0.951<br>lag 10: 0.90 (0.62–1.33), p=0.608 |
| Heat wave duration | lag 0: EDF=0.00, p=0.672<br>lag 1: EDF=0.00, p=0.951<br>lag 2: EDF=0.28, p=0.280<br>**lag 5:** | lag 9: EDF=0.53, p=0.178<br>lag 5: EDF=0.00, p=0.627<br>lag 0: EDF=0.00, p=0.351<br>**lag 9:** | lag 2: EDF=0.00, p=0.750<br>lag 0: EDF=0.50, p=0.142<br>lag 10: EDF=0.61, p=0.126<br>lag 5: | lag 6: EDF=0.10, p=0.324<br>lag 0: EDF=0.00, p=0.963<br>lag 0: EDF=0.42, p=0.213<br>lag 0: | lag 4: EDF=0.00, p=0.619<br>lag 2: EDF=0.00, p=0.618<br>lag 0: EDF=0.00, p=0.539<br>lag 1: | lag 10: EDF=0.00, p=0.857<br>lag 4: EDF=0.00, p=0.782<br>lag 3: EDF=0.00, p=0.617<br>lag 1: | lag 0: EDF=0.58, p=0.221<br>lag 0: EDF=0.37, p=0.216<br>lag 7: EDF=0.00, p=0.414<br>**lag 9:** | lag 4: EDF=0.00, p=0.960<br>lag 8: EDF=0.00, p=0.983<br>lag 8: EDF=0.00, p=0.778<br>lag 7: | **lag 8: EDF=7.98, p=<0.001**<br>lag 9: EDF=0.43, p=0.347<br>lag 7: EDF=0.86, p=0.235<br>lag 7: | lag 10: EDF=0.00, p=0.555<br>lag 0: EDF=0.00, p=0.806<br>lag 1: EDF=0.00, p=0.947<br>lag 2: | lag 7: EDF=0.46, p=0.436<br>lag 7: EDF=0.00, p=0.477<br>lag 0: EDF=0.35, p=0.438<br>lag 10: |

| **Atmospheric factor** | **1.1** | **1.2** | **2.1** | **2.2** | **3.1** | **4.1** | **5.1** | **5.2** | **6.1** | **6.2** | **6.3** |
|---|---|---|---|---|---|---|---|---|---|---|---|
| | **EDF=0.94, p=<0.001** | **EDF=0.82, p=0.014** | EDF=0.00, p=0.976 | EDF=0.00, p=0.479 | EDF=0.00, p=0.805 | EDF=0.26, p=0.299 | **EDF=0.82, p=0.014** | EDF=0.70, p=0.100 | EDF=0.00, p=0.560 | EDF=0.00, p=0.708 | EDF=0.00, p=0.908 |
| Cold wave intensity | lag 1: EDF=0.00, p=0.716<br>lag 5: EDF=0.00, p=0.893<br>lag 5: EDF=0.00, p=0.608 | lag 9: EDF=0.00, p=0.765<br>lag 0: EDF=0.00, p=0.901<br>lag 5: EDF=0.13, p=0.292<br>lag 6: EDF=0.48, p=0.162 | **lag 6: EDF=0.93, p=<0.001**<br>lag 0: EDF=0.00, p=0.519<br>lag 4: EDF=0.23, p=0.249<br>lag 9: EDF=0.18, p=0.227 | lag 0: EDF=0.01, p=0.408<br>lag 8: EDF=0.00, p=0.685<br>lag 9: EDF=0.53, p=0.132 | lag 5: EDF=0.00, p=0.821<br>lag 5: EDF=0.00, p=0.897 | lag 3: EDF=0.64, p=0.218<br>lag 5: EDF=0.26, p=0.281 | lag 5: EDF=0.00, p=0.917 | lag 0: EDF=0.52, p=0.150<br>**lag 0: EDF=2.53, p=0.002**<br>lag 10: EDF=0.00, p=0.502 | lag 1: EDF=0.91, p=0.149 | lag 0: EDF=0.00, p=0.843<br>lag 10: EDF=0.26, p=0.250<br>lag 6: EDF=0.05, p=0.272 | lag 10: EDF=0.83, p=0.160<br>**lag 0: EDF=0.88, p=0.021** |
| Cold stress wave day | lag 6: 1.13 (0.93–1.37), p=0.227<br>lag 1: 1.00 (0.87–1.16), p=0.949<br>lag 10: 1.14 (0.95–1.37), p=0.166<br>**lag 0: 1.18 (1.00–1.39), p=0.049** | lag 9: 1.10 (0.93–1.30), p=0.261<br>lag 0: 1.00 (0.87–1.14), p=0.943<br>**lag 1: 1.18 (1.01–1.37), p=0.039**<br>lag 3: 0.89 (0.77–1.03), p=0.127 | **lag 0: 1.24 (1.04–1.48), p=0.015**<br>lag 0: 0.94 (0.84–1.05), p=0.289<br>lag 7: 1.04 (0.88–1.23), p=0.660<br>lag 0: 1.10 (0.97–1.25), p=0.154 | lag 4: 1.19 (0.99–1.43), p=0.067<br>lag 0: 0.96 (0.86–1.07), p=0.483<br>**lag 3: 0.82 (0.68–0.97), p=0.025**<br>lag 7: 1.01 (0.89–1.13), p=0.919 | lag 7: 0.89 (0.65–1.23), p=0.486<br>**lag 0: 1.26 (1.08–1.46), p=0.003**<br>**lag 1: 0.60 (0.42–0.85), p=0.004**<br>**lag 4: 1.26 (1.05–1.50), p=0.011** | lag 1: 1.19 (0.85–1.66), p=0.319<br>**lag 9: 1.22 (1.03–1.46), p=0.021**<br>lag 0: 1.30 (0.94–1.79), p=0.117<br>lag 5: 1.12 (0.92–1.36), p=0.260 | **lag 2: 0.72 (0.54–0.96), p=0.025**<br>lag 0: 1.11 (0.96–1.29), p=0.173<br>lag 8: 1.25 (0.99–1.58), p=0.061<br>lag 5: 1.15 (1.00–1.33), p=0.057 | lag 0: 1.05 (0.77–1.44), p=0.736<br>lag 8: 1.05 (0.87–1.26), p=0.621<br>lag 10: 1.38 (0.96–1.98), p=0.084<br>lag 3: 1.15 (0.91–1.45), p=0.252 | **lag 8: 1.96 (1.00–3.84), p=0.048**<br>lag 3: 1.18 (0.86–1.62), p=0.308<br>lag 1: 0.55 (0.16–1.91), p=0.343<br>lag 2: 1.23 (0.90–1.69), p=0.186 | lag 0: 1.09 (0.92–1.29), p=0.305<br>lag 10: 1.07 (0.98–1.18), p=0.138<br>lag 9: 1.09 (0.93–1.27), p=0.301<br>lag 6: 1.06 (0.96–1.17), p=0.274 | lag 2: 0.77 (0.47–1.24), p=0.279<br>**lag 7: 1.31 (1.02–1.68), p=0.034**<br>**lag 9: 1.80 (1.19–2.73), p=0.005**<br>lag 4: 0.98 (0.74–1.29), p=0.868 |
| Cold stress wave intensity | lag 10: EDF=0.00, p=0.890<br>lag 10: EDF=0.00, p=0.701<br>lag 0: | lag 0: EDF=0.00, p=0.915<br>lag 10: EDF=0.23, p=0.228 | **lag 2: EDF=0.85, p=0.007** | **lag 0: EDF=0.90, p=0.004**<br>lag 10: EDF=0.69, p=0.062<br>lag 6: | lag 0: EDF=0.00, p=0.955<br>**lag 0: EDF=0.91, p=0.007** | lag 10: EDF=1.93, p=0.059<br>lag 9: EDF=0.00, p=0.481<br>**lag 8:** | lag 6: EDF=0.42, p=0.267<br>lag 0: EDF=0.29, p=0.223<br>**lag 0: EDF=3.69,** | lag 8: EDF=0.81, p=0.079<br>lag 10: EDF=0.52, p=0.291<br>**lag 3:** | lag 4: EDF=0.66, p=0.198<br>lag 2: EDF=3.98, p=0.106<br>lag 0: | lag 0: EDF=0.44, p=0.157 | lag 3: EDF=0.00, p=0.780<br>**lag 8: EDF=0.92, p=0.026**<br>lag 8: |

| Atmospheric factor | 1.1 | 1.2 | 2.1 | 2.2 | 3.1 | 4.1 | 5.1 | 5.2 | 6.1 | 6.2 | 6.3 |
|---|---|---|---|---|---|---|---|---|---|---|---|
| | EDF=0.44,<br>p=0.190 | | | EDF=0.00,<br>p=0.479 | | **EDF=0.86,<br>p=0.042** | **p=0.014**<br>lag 7:<br>EDF=0.00,<br>p=0.507 | **EDF=0.82,<br>p=0.037** | EDF=0.00,<br>p=0.816 | | EDF=0.00,<br>p=0.680 |
| Ozone | **lag 0:<br>EDF=0.94,<br>p=0.006**<br>lag 1:<br>EDF=0.00,<br>p=0.824<br>lag 0:<br>EDF=0.64,<br>p=0.122<br>lag 2:<br>EDF=0.00,<br>p=0.335 | lag 1:<br>EDF=0.30,<br>p=0.201<br>lag 0:<br>EDF=0.00,<br>p=0.895<br>lag 6:<br>EDF=0.00,<br>p=0.434 | lag 1:<br>EDF=0.00,<br>p=0.499<br>lag 1:<br>EDF=0.00,<br>p=0.371 | lag 0:<br>EDF=0.00,<br>p=0.483<br>**lag 0:<br>EDF=0.88,<br>p=0.005**<br>lag 7:<br>EDF=0.64,<br>p=0.096 | lag 7:<br>EDF=0.74,<br>p=0.141<br>**lag 7:<br>EDF=0.93,<br>p=0.012<br>lag 1:<br>EDF=0.90,<br>p=0.008** | lag 5:<br>EDF=0.61,<br>p=0.220<br>lag 7:<br>EDF=0.00,<br>p=0.414<br>lag 5:<br>EDF=2.19,<br>p=0.205<br>lag 1:<br>EDF=0.55,<br>p=0.154 | - | lag 3:<br>EDF=0.00,<br>p=0.632<br>**lag 5:<br>EDF=0.85,<br>p=0.025** | lag 0:<br>EDF=5.56,<br>p=0.412<br>lag 2:<br>EDF=0.00,<br>p=0.508<br>lag 0:<br>EDF=0.99,<br>p=0.090<br>lag 1:<br>EDF=0.00,<br>p=0.519 | lag 0:<br>EDF=0.00,<br>p=0.472<br>**lag 0:<br>EDF=1.69,<br>p=0.001** | lag 2:<br>EDF=0.73,<br>p=0.226<br>lag 5:<br>EDF=0.73,<br>p=0.128<br>lag 7:<br>EDF=0.00,<br>p=0.998<br>lag 3:<br>EDF=0.00,<br>p=0.660 |
| Particulate matter ($PM_{2.5}$) | **lag 0:<br>EDF=0.86,<br>p=0.017**<br>lag 0:<br>EDF=0.50,<br>p=0.150<br>lag 1:<br>EDF=0.67,<br>p=0.115<br>lag 2:<br>EDF=0.52,<br>p=0.141 | lag 4:<br>EDF=0.65,<br>p=0.104<br>lag 2:<br>EDF=0.00,<br>p=0.653<br>lag 3:<br>EDF=0.00,<br>p=0.683<br>**lag 4:<br>EDF=0.78,<br>p=0.029** | lag 1:<br>EDF=0.70,<br>p=0.077<br>**lag 0:<br>EDF=0.76,<br>p=0.030**<br>lag 0:<br>EDF=0.71,<br>p=0.063<br>lag 6:<br>EDF=0.00,<br>p=0.382 | **lag 2:<br>EDF=0.78,<br>p=0.042**<br>lag 6:<br>EDF=0.00,<br>p=0.630<br>**lag 6:<br>EDF=0.82,<br>p=0.022<br>lag 3:<br>EDF=0.87,<br>p=0.004** | lag 4:<br>EDF=0.84,<br>p=0.060<br>lag 5:<br>EDF=0.00,<br>p=0.613<br>lag 5:<br>EDF=0.74,<br>p=0.119<br>lag 5:<br>EDF=0.00,<br>p=0.914 | **lag 1:<br>EDF=0.92,<br>p=0.016<br>lag 2:<br>EDF=0.84,<br>p=0.018<br>lag 4:<br>EDF=0.85,<br>p=0.047**<br>lag 6:<br>EDF=0.76,<br>p=0.056 | lag 2:<br>EDF=0.43,<br>p=0.269<br>lag 7:<br>EDF=0.00,<br>p=0.890<br>**lag 7:<br>EDF=3.05,<br>p=0.006<br>lag 2:<br>EDF=0.90,<br>p=0.001** | lag 3:<br>EDF=0.81,<br>p=0.099<br>lag 2:<br>EDF=0.44,<br>p=0.209<br>lag 0:<br>EDF=0.85,<br>p=0.052<br>lag 7:<br>EDF=0.45,<br>p=0.215 | **lag 6: 0.96<br>(0.92–<br>0.99),<br>p=0.019**<br>lag 1:<br>EDF=0.56,<br>p=0.264<br>**lag 3:<br>EDF=4.15,<br>p=0.017**<br>lag 2:<br>EDF=0.40,<br>p=0.356 | **lag 0:<br>EDF=0.85,<br>p=0.015**<br>lag 5:<br>EDF=0.00,<br>p=0.747<br>**lag 4:<br>EDF=0.78,<br>p=0.036**<br>lag 1:<br>EDF=0.00,<br>p=0.861 | **lag 6:<br>EDF=7.89,<br>p=<0.001**<br>lag 3:<br>EDF=0.00,<br>p=0.955<br>lag 6:<br>EDF=2.38,<br>p=0.102<br>lag 3:<br>EDF=0.58,<br>p=0.228 |

**Table S16.** Overview of time lags, effect sizes (effective degree of freedom (EDF) or relative risk (95% confidence interval)) and p-values per human-bioclimatic region, atmospheric factor and subgroup (pink = female <60 years, orange = females ≥60 years, green = males <60 years, blue = males ≥60 years) for non subdural hemorrhage (stroke unit), significant p-values (≤ 0.05) are bold

| Atmospheric factor | 1.1 | 1.2 | 2.1 | 2.2 | 3.1 | 4.1 | 5.1 | 5.2 | 6.1 | 6.2 | 6.3 |
|---|---|---|---|---|---|---|---|---|---|---|---|
| Wind day | lag 7: 1.29<br>(0.86– | **lag 1: 0.50<br>(0.29–** | lag 4: 0.67<br>(0.43– | **lag 9: 1.44<br>(1.02–** | **lag 3: 2.33<br>(1.37–** | **lag 8: 2.63<br>(1.57–** | lag 10: 1.50<br>(0.97– | **lag 9: 1.80<br>(1.10–** | lag 10: 2.71<br>(0.89– | lag 8: 1.45<br>(0.99– | **lag 10: 3.06<br>(1.77–** |

| Atmospheric factor | 1.1 | 1.2 | 2.1 | 2.2 | 3.1 | 4.1 | 5.1 | 5.2 | 6.1 | 6.2 | 6.3 |
|---|---|---|---|---|---|---|---|---|---|---|---|
| | 1.91), p=0.214<br>lag 5: 1.14 (0.96–1.35), p=0.128<br>**lag 9: 1.49 (1.12–1.98), p=0.006**<br>**lag 8: 1.26 (1.07–1.48), p=0.005** | **0.88), p=0.015**<br>lag 5: 1.15 (0.97–1.37), p=0.099<br>lag 4: 1.26 (0.96–1.66), p=0.097<br>**lag 6: 1.26 (1.07–1.48), p=0.006** | 1.04), p=0.076<br>lag 0: 1.05 (0.91–1.22), p=0.465<br>lag 5: 1.22 (0.94–1.59), p=0.142<br>lag 0: 1.11 (0.96–1.28), p=0.176 | **2.04), p=0.040**<br>lag 8: 1.11 (0.95–1.29), p=0.180<br>lag 8: 1.23 (0.93–1.63), p=0.153<br>lag 1: 0.84 (0.71–1.01), p=0.061 | **3.94), p=0.002**<br>**lag 10: 1.28 (1.00–1.64), p=0.050**<br>lag 8: 1.57 (0.97–2.55), p=0.066<br>**lag 10: 1.32 (1.02–1.70), p=0.035** | **4.41), p=<0.001**<br>**lag 10: 1.47 (1.17–1.86), p=0.001**<br>lag 7: 1.53 (0.97–2.41), p=0.069<br>**lag 4: 0.68 (0.48–0.95), p=0.026** | 2.33), p=0.069<br>**lag 9: 1.23 (1.02–1.48), p=0.028**<br>**lag 9: 1.50 (1.06–2.12), p=0.023**<br>lag 8: 1.12 (0.93–1.36), p=0.233 | **2.96), p=0.020**<br>**lag 5: 0.70 (0.51–0.96), p=0.028**<br>**lag 7: 1.98 (1.27–3.09), p=0.003**<br>lag 8: 1.19 (0.90–1.56), p=0.217 | 8.20), p=0.078<br>**lag 7: 1.61 (1.09–2.37), p=0.016**<br>**lag 10: 2.83 (1.35–5.92), p=0.006**<br>**lag 2: 1.61 (1.05–2.46), p=0.029** | 2.12), p=0.057<br>**lag 9: 1.15 (1.01–1.32), p=0.039**<br>**lag 0: 0.54 (0.36–0.80), p=0.002**<br>lag 9: 1.13 (0.98–1.30), p=0.088 | **5.29), p=<0.001**<br>lag 6: 1.15 (0.83–1.60), p=0.403<br>lag 10: 1.43 (0.82–2.50), p=0.205<br>**lag 10: 1.49 (1.08–2.05), p=0.014** |
| Tropical night | lag 5: 1.37 (0.17–11.06), p=0.765<br>lag 3: 0.76 (0.19–3.14), p=0.709<br>**lag 4: 4.15 (1.34–12.91), p=0.014**<br>lag 6: 1.09 (0.40–3.00), p=0.866 | lag 0: 2.41 (0.74–7.83), p=0.145<br>lag 10: 1.65 (0.97–2.80), p=0.063<br>lag 2: 1.80 (0.68–4.77), p=0.241<br>lag 3: 0.49 (0.16–1.53), p=0.220 | **lag 7: 15.67 (3.38–72.78), p=<0.001**<br>lag 7: 1.66 (0.54–5.11), p=0.378<br>lag 8: 3.33 (0.82–13.41), p=0.091<br>lag 6: 2.24 (0.84–6.01), p=0.109 | **lag 10: 8.85 (3.32–23.62), p=<0.001**<br>lag 5: 0.62 (0.31–1.27), p=0.195<br>lag 1: 2.76 (0.86–8.85), p=0.087<br>lag 2: 0.85 (0.40–1.81), p=0.671 | - | lag 0: 0.00 (0.00–Inf), p=1.000<br>**lag 5: 7.54 (1.82–31.13), p=0.005**<br>lag 0: 0.00 (0.00–Inf), p=1.000<br>lag 2: 5.14 (0.68–38.65), p=0.112 | lag 5: 8.92 (0.99–80.71), p=0.052<br>lag 9: 2.74 (0.68–11.12), p=0.158<br>lag 0: 0.00 (0.00–Inf), p=1.000<br>lag 0: 2.05 (0.29–14.65), p=0.474 | lag 0: 0.00 (0.00–Inf), p=1.000<br>**lag 2: 5.60 (1.75–17.86), p=0.004**<br>lag 9: 4.18 (0.49–35.66), p=0.191<br>lag 3: 3.30 (0.43–25.20), p=0.251 | lag 0: 0.00 (0.00–Inf), p=1.000<br>lag 0: 0.00 (0.00–Inf), p=1.000<br>lag 0: 0.00 (0.00–Inf), p=1.000<br>lag 4: 3.11 (0.65–15.00), p=0.157 | lag 10: 2.32 (0.58–9.33), p=0.234<br>lag 10: 1.45 (0.87–2.41), p=0.159<br>lag 4: 2.07 (0.79–5.39), p=0.136<br>lag 6: 0.49 (0.16–1.56), p=0.227 | **lag 0: 13.60 (1.37–134.80), p=0.026**<br>**lag 9: 6.24 (2.23–17.44), p=<0.001**<br>**lag 10: 8.69 (1.14–66.27), p=0.037**<br>lag 2: 2.16 (0.48–9.66), p=0.316 |
| Hot day | lag 8: 1.76 (0.80–3.88), p=0.163<br>lag 5: 0.76 (0.51–1.14), p=0.191<br>lag 5: 0.96 | **lag 6: 0.18 (0.04–0.74), p=0.017**<br>**lag 7: 1.35 (1.04–1.76), p=0.025**<br>lag 7: 0.61 | lag 4: 0.58 (0.27–1.26), p=0.168<br>lag 1: 0.93 (0.72–1.20), p=0.560<br>lag 6: 1.33 | lag 0: 0.69 (0.32–1.47), p=0.336<br>**lag 1: 0.65 (0.47–0.91), p=0.011**<br>lag 0: 0.67 | **lag 3: 3.16 (1.09–9.17), p=0.034**<br>**lag 10: 2.10 (1.35–3.26), p=<0.001**<br>**lag 5: 2.56** | lag 0: 0.00 (0.00–Inf), p=1.000<br>lag 0: 2.62 (0.91–7.52), p=0.073<br>lag 6: 2.11 (0.30– | lag 4: 1.34 (0.49–3.65), p=0.574<br>lag 0: 0.80 (0.43–1.48), p=0.483<br>lag 2: 2.40 | lag 8: 1.20 (0.39–3.68), p=0.744<br>lag 8: 0.76 (0.39–1.49), p=0.422<br>lag 9: 2.18 | lag 0: 0.00 (0.00–Inf), p=1.000<br>lag 8: 1.38 (0.44–4.37), p=0.583<br>lag 7: 0.10 (0.00– | lag 4: 0.38 (0.12–1.18), p=0.094<br>**lag 3: 0.74 (0.57–0.97), p=0.031**<br>**lag 3: 0.46** | **lag 7: 4.05 (1.42–11.57), p=0.009**<br>lag 6: 0.64 (0.27–1.47), p=0.291<br>lag 6: 2.27 |

| Atmospheric factor | 1.1 | 1.2 | 2.1 | 2.2 | 3.1 | 4.1 | 5.1 | 5.2 | 6.1 | 6.2 | 6.3 |
|---|---|---|---|---|---|---|---|---|---|---|---|
| | (0.49–1.88), p=0.896 lag 6: 1.32 (0.95–1.84), p=0.103 | (0.30–1.24), p=0.174 **lag 5: 0.66 (0.45–0.97), p=0.034** | (0.86–2.07), p=0.206 lag 0: 0.84 (0.64–1.11), p=0.231 | (0.37–1.23), p=0.201 **lag 0: 0.74 (0.54–1.00), p=0.049** | **(1.03–6.36), p=0.043** lag 9: 1.24 (0.67–2.29), p=0.492 | 14.94), p=0.455 lag 9: 2.08 (0.65–6.63), p=0.215 | (0.98–5.87), p=0.056 lag 6: 0.79 (0.47–1.32), p=0.362 | (0.93–5.08), p=0.072 **lag 10: 1.65 (1.03–2.65), p=0.036** | 12402152.70), p=0.811 lag 4: 1.10 (0.41–2.97), p=0.850 | **(0.22–0.93), p=0.030** lag 6: 0.77 (0.57–1.05), p=0.096 | (0.88–5.82), p=0.089 lag 3: 1.38 (0.76–2.51), p=0.285 |
| Wet-bulb globe temperature | - | **lag 2: EDF=2.46, p=0.012 lag 1: 0.98 (0.97–0.99), p=0.001 lag 9: EDF=2.11, p=<0.001 lag 2: EDF=1.00, p=<0.001** | **lag 3: EDF=2.39, p=0.005 lag 0: EDF=1.02, p=<0.001** | **lag 1: EDF=0.93, p=<0.001** | - | **lag 0: EDF=3.22, p=0.009 lag 2: EDF=1.14, p=0.027 lag 3: 0.98 (0.97–0.99), p=0.008** | **lag 2: EDF=1.02, p=0.020** | **lag 1: EDF=2.33, p=<0.001 lag 0: 0.97 (0.95–1.00), p=0.017** | **lag 1: EDF=4.97, p=0.006 lag 10: EDF=1.35, p=0.008** lag 4: EDF=3.82, p=0.095 | lag 1: EDF=0.54, p=0.134 **lag 2: EDF=1.84, p=<0.001** | **lag 1: EDF=1.09, p=<0.001 lag 10: EDF=1.32, p=0.010** |
| Wind chill index | **lag 0: EDF=4.60, p=0.045 lag 0: EDF=0.85, p=0.011** lag 7: EDF=0.00, p=0.714 **lag 9: EDF=0.84, p=0.015** | **lag 3: EDF=0.81, p=0.027** | **lag 0: EDF=1.09, p=<0.001** | **lag 9: EDF=5.41, p=<0.001** lag 0: EDF=0.00, p=0.888 lag 4: EDF=0.00, p=0.525 | lag 5: 0.99 (0.94–1.03), p=0.506 **lag 2: EDF=0.88, p=0.020 lag 8: EDF=1.14, p=0.008 lag 7: EDF=0.84, p=0.046** | lag 5: EDF=4.29, p=0.208 lag 5: 1.03 (1.00–1.07), p=0.069 lag 3: EDF=0.00, p=0.576 | **lag 2: 0.98 (0.97–0.99), p=<0.001** | lag 2: EDF=1.89, p=0.063 **lag 10: EDF=1.05, p=0.018** lag 0: EDF=0.54, p=0.241 | lag 0: EDF=8.54, p=0.173 | **lag 1: 0.99 (0.98–1.00), p=0.039 lag 0: EDF=0.91, p=0.001** | **lag 5: EDF=7.12, p=0.007 lag 0: EDF=6.06, p=0.004** |
| Heat wave intensity | lag 6: EDF=3.34, p=0.081 lag 0: EDF=0.00, | lag 7: EDF=0.69, p=0.200 lag 0: EDF=0.55, | lag 7: EDF=0.87, p=0.055 lag 0: EDF=0.12, | **lag 7: EDF=0.92, p=0.021** lag 0: EDF=1.70, | lag 10: EDF=3.43, p=0.328 **lag 0: EDF=0.83,** | lag 3: EDF=0.84, p=0.332 lag 8: EDF=0.65, | lag 7: EDF=0.74, p=0.297 lag 0: EDF=0.72, | lag 7: EDF=2.27, p=0.356 lag 6: EDF=0.51, | lag 5: EDF=3.96, p=0.307 lag 0: EDF=0.87, | lag 4: EDF=0.00, p=0.656 lag 0: EDF=0.00, | lag 1: EDF=2.88, p=0.505 lag 1: EDF=2.11, |

| Atmospheric factor | 1.1 | 1.2 | 2.1 | 2.2 | 3.1 | 4.1 | 5.1 | 5.2 | 6.1 | 6.2 | 6.3 |
|---|---|---|---|---|---|---|---|---|---|---|---|
| | p=0.471<br>lag 4: EDF=0.35, p=0.336<br>lag 10: EDF=0.34, p=0.236 | p=0.150<br>lag 10: EDF=0.00, p=0.883<br>lag 0: EDF=0.00, p=0.885 | p=0.265<br>lag 5: EDF=0.55, p=0.205<br>lag 0: EDF=0.00, p=0.333 | p=0.072<br>**lag 0: EDF=0.89, p=0.022**<br>lag 0: EDF=0.72, p=0.064 | **p=0.048**<br>lag 10: EDF=0.85, p=0.145<br>lag 4: EDF=0.68, p=0.158 | p=0.180<br>lag 0: EDF=0.91, p=0.080<br>**lag 3: EDF=0.85, p=0.044** | p=0.090<br>lag 0: EDF=0.89, p=0.062<br>lag 5: EDF=0.00, p=0.553 | p=0.250<br>lag 0: EDF=0.81, p=0.231<br>lag 1: EDF=0.78, p=0.101 | p=0.130<br>**lag 9: EDF=6.57, p=0.002**<br>lag 9: EDF=0.79, p=0.193 | p=0.462<br>**lag 4: EDF=0.88, p=0.020**<br>**lag 1: EDF=0.83, p=0.015** | p=0.247<br>lag 0: EDF=0.95, p=0.053<br>lag 5: EDF=0.85, p=0.083 |
| Ozone | lag 2: EDF=0.00, p=0.666<br>lag 6: EDF=0.73, p=0.051<br>**lag 2: EDF=0.85, p=0.045**<br>**lag 6: EDF=0.82, p=0.024** | **lag 7: EDF=0.93, p=0.043**<br>lag 1: EDF=0.00, p=0.678<br>lag 0: EDF=0.29, p=0.348 | **lag 6: EDF=0.97, p=0.029**<br>lag 3: EDF=0.33, p=0.200 | lag 0: EDF=0.11, p=0.462<br>**lag 0: 0.99 (0.99–1.00), p=<0.001**<br>lag 2: EDF=0.77, p=0.115 | lag 5: EDF=1.15, p=0.323<br>lag 0: EDF=0.00, p=0.680<br>**lag 5: EDF=1.07, p=0.031**<br>lag 0: EDF=0.00, p=0.380 | **lag 0: EDF=1.14, p=0.015**<br>lag 1: EDF=0.29, p=0.309<br>lag 3: EDF=2.33, p=0.097<br>lag 2: EDF=1.83, p=0.201 | **lag 0: EDF=2.29, p=<0.001**<br>lag 6: EDF=0.84, p=0.118<br>lag 2: EDF=0.00, p=0.476 | lag 0: 1.01 (0.99–1.04), p=0.166<br>lag 1: EDF=2.74, p=0.062<br>lag 6: EDF=0.78, p=0.083 | lag 1: EDF=8.17, p=0.478<br>**lag 0: EDF=1.15, p=0.009**<br>lag 7: EDF=7.32, p=0.060<br>lag 3: EDF=2.79, p=0.343 | **lag 6: EDF=4.93, p=0.006**<br>**lag 1: 1.01 (1.01–1.02), p=0.002**<br>lag 0: EDF=1.82, p=0.051 | **lag 5: EDF=1.39, p=<0.001**<br>lag 4: EDF=2.10, p=0.145<br>lag 3: EDF=4.14, p=0.240<br>lag 3: EDF=0.10, p=0.451 |

**Table S17.** Overview of time lags, effect sizes (effective degree of freedom (EDF) or relative risk (95% confidence interval)) and p-values per human-bioclimatic region, atmospheric factor and subgroup (pink = female <60 years, orange = females ≥60 years, green = males <60 years, blue = males ≥60 years) for pneumonia, significant p-values (≤ 0.05) are bold

| Atmospheric factor | 1.1 | 1.2 | 2.1 | 2.2 | 3.1 | 4.1 | 5.1 | 5.2 | 6.1 | 6.2 | 6.3 |
|---|---|---|---|---|---|---|---|---|---|---|---|
| Sea-level pressure (SLP) | **lag 0: EDF=0.79, p=0.027**<br>lag 10: EDF=0.16, p=0.259<br>lag 1: EDF=0.00, p=0.985<br>lag 1: EDF=0.52, p=0.136 | lag 2: EDF=0.66, p=0.083<br>lag 7: EDF=0.15, p=0.267<br>lag 9: EDF=0.00, p=0.692<br>lag 7: EDF=0.70, p=0.061 | lag 8: EDF=0.00, p=0.763<br>lag 1: EDF=0.00, p=0.501<br>lag 9: EDF=0.01, p=0.327<br>**lag 2: EDF=5.23, p=0.004** | lag 9: EDF=0.00, p=0.467<br>lag 1: EDF=0.37, p=0.193<br>lag 4: EDF=0.00, p=0.621<br>lag 4: EDF=0.01, p=0.415 | lag 10: EDF=0.00, p=0.863<br>lag 2: EDF=0.00, p=0.876<br>lag 10: EDF=0.26, p=0.249<br>lag 10: EDF=0.00, p=0.673 | **lag 0: EDF=0.77, p=0.047**<br>**lag 0: EDF=0.93, p=<0.001**<br>lag 9: EDF=0.62, p=0.103<br>**lag 0: EDF=0.81, p=0.021** | lag 1: EDF=0.00, p=0.552<br>lag 2: EDF=0.00, p=0.445<br>**lag 6: EDF=0.93, p=<0.001**<br>**lag 8: EDF=0.77, p=0.033** | lag 3: EDF=0.00, p=0.522<br>lag 5: EDF=0.00, p=0.469<br>lag 3: EDF=0.00, p=0.961<br>lag 5: EDF=0.00, p=0.522 | **lag 4: EDF=0.91, p=0.016**<br>**lag 0: EDF=0.95, p=0.001**<br>**lag 7: EDF=0.84, p=0.032**<br>**lag 5: EDF=0.91, p=0.004** | **lag 8: EDF=2.17, p=0.010**<br>lag 1: EDF=0.00, p=0.973<br>**lag 4: EDF=2.99, p=0.002**<br>**lag 0: 0.99 (0.99–** | lag 0: EDF=0.32, p=0.292<br>**lag 2: EDF=0.86, p=0.022**<br>lag 5: EDF=0.62, p=0.126<br>**lag 4: EDF=0.84, p=0.020** |

| **Atmospheric factor** | **1.1** | **1.2** | **2.1** | **2.2** | **3.1** | **4.1** | **5.1** | **5.2** | **6.1** | **6.2** | **6.3** |
|---|---|---|---|---|---|---|---|---|---|---|---|
| | | | | | | | | | | **1.00), p=<0.001** | |
| Sea-level pressure difference (SLPD) | **lag 2: 1.13 (1.04–1.22), p=0.004**<br>lag 2: 0.95 (0.88–1.03), p=0.203<br>**lag 2: 1.08 (1.01–1.16), p=0.023**<br>**lag 8: 0.92 (0.86–0.98), p=0.007** | lag 6: 1.04 (0.98–1.11), p=0.214<br>lag 9: 0.95 (0.89–1.01), p=0.112<br>**lag 7: 1.07 (1.02–1.13), p=0.011**<br>lag 1: 1.05 (0.99–1.10), p=0.109 | lag 4: 0.93 (0.85–1.02), p=0.110<br>lag 9: 0.95 (0.88–1.03), p=0.187<br>**lag 7: 0.91 (0.85–0.98), p=0.018**<br>lag 3: 1.04 (0.97–1.12), p=0.230 | **lag 3: 1.09 (1.00–1.18), p=0.045**<br>lag 3: 0.95 (0.89–1.03), p=0.206<br>lag 2: 1.01 (0.95–1.09), p=0.698<br>**lag 0: 0.91 (0.85–0.97), p=0.002** | lag 5: 0.87 (0.75–1.00), p=0.056<br>lag 10: 1.11 (0.98–1.26), p=0.092<br>lag 7: 1.05 (0.92–1.19), p=0.467<br>**lag 3: 1.12 (1.00–1.26), p=0.042** | **lag 4: 0.85 (0.76–0.95), p=0.004**<br>lag 6: 0.90 (0.82–1.00), p=0.052<br>**lag 4: 0.87 (0.79–0.96), p=0.005**<br>lag 9: 0.93 (0.85–1.01), p=0.087 | **lag 1: 1.15 (1.04–1.27), p=0.007**<br>lag 5: 1.09 (1.00–1.18), p=0.053<br>lag 4: 1.03 (0.95–1.12), p=0.483<br>lag 1: 0.95 (0.89–1.02), p=0.171 | **lag 5: 1.23 (1.10–1.39), p=<0.001**<br>lag 4: 0.91 (0.81–1.03), p=0.138<br>lag 3: 1.05 (0.95–1.16), p=0.370<br>**lag 0: 0.88 (0.79–0.98), p=0.016** | lag 3: 1.06 (0.87–1.28), p=0.580<br>lag 4: 0.91 (0.77–1.07), p=0.232<br>lag 1: 1.12 (0.96–1.31), p=0.148<br>**lag 9: 0.80 (0.69–0.92), p=0.002** | lag 1: 0.94 (0.87–1.01), p=0.096<br>**lag 0: 0.92 (0.86–0.98), p=0.012**<br>lag 4: 1.05 (0.98–1.12), p=0.166<br>lag 7: 0.96 (0.91–1.02), p=0.210 | **lag 3: 1.21 (1.04–1.41), p=0.015**<br>lag 1: 1.14 (0.98–1.31), p=0.083<br>lag 6: 1.11 (0.98–1.26), p=0.102<br>lag 0: 0.88 (0.78–1.00), p=0.051 |
| Wind day | lag 1: 1.11 (1.00–1.24), p=0.056<br>lag 4: 1.07 (0.98–1.18), p=0.145<br>lag 2: 1.05 (0.96–1.14), p=0.291<br>**lag 4: 1.10 (1.02–1.19), p=0.016** | lag 1: 1.02 (0.93–1.11), p=0.731<br>lag 2: 1.01 (0.93–1.10), p=0.730<br>lag 0: 1.02 (0.95–1.10), p=0.505<br>lag 6: 1.03 (0.96–1.11), p=0.374 | lag 0: 1.01 (0.92–1.12), p=0.806<br>lag 1: 1.05 (0.97–1.14), p=0.248<br>lag 5: 1.06 (0.98–1.15), p=0.173<br>lag 0: 1.02 (0.95–1.10), p=0.617 | lag 1: 1.03 (0.94–1.13), p=0.504<br>**lag 10: 1.09 (1.01–1.18), p=0.033**<br>lag 7: 1.04 (0.96–1.12), p=0.310<br>lag 0: 0.98 (0.91–1.06), p=0.647 | lag 6: 1.13 (0.98–1.31), p=0.088<br>lag 8: 1.03 (0.90–1.18), p=0.648<br>lag 10: 1.08 (0.95–1.23), p=0.224<br>**lag 1: 1.16 (1.03–1.30), p=0.011** | **lag 10: 1.28 (1.11–1.48), p=<0.001**<br>lag 5: 1.07 (0.93–1.22), p=0.331<br>lag 9: 1.02 (0.90–1.17), p=0.726<br>**lag 8: 1.13 (1.01–1.26), p=0.033** | lag 0: 1.04 (0.92–1.18), p=0.558<br>lag 1: 1.04 (0.95–1.15), p=0.410<br>lag 0: 1.09 (0.99–1.20), p=0.095<br>lag 2: 1.00 (0.92–1.09), p=0.951 | lag 0: 1.02 (0.89–1.17), p=0.793<br>lag 8: 1.01 (0.88–1.17), p=0.860<br>lag 8: 1.04 (0.93–1.18), p=0.475<br>lag 2: 0.98 (0.87–1.12), p=0.801 | lag 9: 1.26 (0.99–1.59), p=0.062<br>lag 0: 0.95 (0.77–1.18), p=0.663<br>lag 10: 1.18 (0.97–1.45), p=0.096<br>lag 7: 1.12 (0.94–1.34), p=0.202 | lag 1: 0.95 (0.86–1.05), p=0.305<br>lag 2: 1.03 (0.95–1.11), p=0.528<br>lag 0: 1.00 (0.93–1.09), p=0.937<br>lag 0: 0.98 (0.91–1.06), p=0.675 | lag 10: 1.13 (0.93–1.38), p=0.212<br>lag 3: 0.98 (0.80–1.18), p=0.808<br>**lag 8: 1.17 (1.00–1.37), p=0.049**<br>**lag 10: 1.20 (1.03–1.40), p=0.021** |
| Tropical night | lag 9: 1.76 (0.96–3.23), p=0.067 | lag 3: 0.80 (0.46–1.36), p=0.405 | lag 2: 2.12 (0.76–5.91), p=0.153 | lag 7: 0.65 (0.35–1.19), p=0.163 | - | **lag 5: 5.09 (1.57–16.50), p=0.007** | **lag 2: 6.71 (2.51–17.91), p=<0.001** | lag 5: 1.81 (0.35–9.38), p=0.480 | lag 6: 3.63 (0.76–17.35), p=0.107 | **lag 3: 0.29 (0.09–0.89), p=0.031** | lag 3: 2.26 (0.80–6.37), p=0.122 |

| Atmospheric factor | 1.1 | 1.2 | 2.1 | 2.2 | 3.1 | 4.1 | 5.1 | 5.2 | 6.1 | 6.2 | 6.3 |
|---|---|---|---|---|---|---|---|---|---|---|---|
| | **lag 1: 2.31 (1.47–3.63), p=<0.001**<br>lag 6: 0.67 (0.33–1.38), p=0.279<br>lag 7: 0.79 (0.43–1.49), p=0.472 | lag 4: 1.30 (0.97–1.74), p=0.080<br>lag 8: 0.83 (0.58–1.18), p=0.290<br>**lag 4: 1.31 (1.03–1.67), p=0.030** | lag 3: 1.96 (0.94–4.09), p=0.074<br>**lag 9: 2.80 (1.39–5.64), p=0.004**<br>**lag 1: 2.74 (1.61–4.67), p=<0.001** | **lag 6: 1.30 (1.02–1.66), p=0.035**<br>**lag 7: 0.53 (0.33–0.86), p=0.010**<br>lag 4: 1.25 (0.99–1.57), p=0.065 | | **lag 3: 2.95 (1.15–7.59), p=0.025**<br>lag 4: 2.23 (0.56–8.81), p=0.255<br>**lag 3: 3.22 (1.26–8.23), p=0.015** | **lag 0: 3.30 (1.29–8.39), p=0.012**<br>lag 6: 2.83 (0.96–8.37), p=0.060<br>lag 3: 1.95 (0.82–4.64), p=0.129 | **lag 1: 4.16 (1.62–10.64), p=0.003**<br>**lag 7: 3.79 (1.41–10.23), p=0.008**<br>**lag 5: 3.82 (1.71–8.56), p=0.001** | **lag 1: 3.55 (1.32–9.51), p=0.012**<br>**lag 3: 4.12 (1.47–11.55), p=0.007**<br>**lag 5: 4.77 (2.41–9.45), p=<0.001** | lag 4: 0.87 (0.54–1.42), p=0.586<br>lag 5: 0.78 (0.48–1.25), p=0.302<br>**lag 1: 1.44 (1.08–1.93), p=0.013** | lag 1: 1.94 (0.91–4.14), p=0.086<br>lag 5: 1.87 (0.70–4.96), p=0.211<br>lag 8: 1.65 (0.76–3.60), p=0.209 |
| Daily mean temperature | lag 0: EDF=0.66, p=0.084<br>lag 0: EDF=0.04, p=0.290 | lag 0: EDF=0.00, p=0.730<br>lag 0: EDF=0.00, p=0.560 | **lag 0: EDF=0.83, p=0.022**<br>lag 0: EDF=0.00, p=0.488 | lag 0: EDF=0.72, p=0.061 | **lag 0: 0.98 (0.97–0.99), p=<0.001** | **lag 0: EDF=0.91, p=0.010**<br>lag 0: EDF=0.00, p=0.401 | - | lag 0: EDF=0.22, p=0.268<br>**lag 0: EDF=0.79, p=0.035**<br>lag 0: EDF=0.00, p=0.705 | **lag 10: EDF=0.94, p=0.032**<br>lag 9: EDF=0.77, p=0.075 | lag 0: 1.00 (0.99–1.00), p=0.384 | lag 0: EDF=0.00, p=0.526 |
| Frost day | **lag 2: 1.08 (1.00–1.17), p=0.049**<br>**lag 8: 1.08 (1.01–1.15), p=0.034**<br>**lag 0: 1.08 (1.01–1.16), p=0.017**<br>lag 0: 1.01 (0.94–1.08), p=0.835 | **lag 0: 1.08 (1.00–1.17), p=0.039**<br>lag 0: 1.03 (0.97–1.10), p=0.295<br>**lag 2: 1.09 (1.03–1.15), p=0.001**<br>lag 0: 1.06 (1.00–1.12), p=0.054 | lag 0: 1.01 (0.94–1.09), p=0.755<br>**lag 0: 1.13 (1.07–1.19), p=<0.001**<br>**lag 0: 1.07 (1.01–1.13), p=0.014**<br>**lag 0: 1.08 (1.03–1.14), p=0.001** | lag 0: 0.99 (0.93–1.07), p=0.870<br>lag 0: 1.07 (1.00–1.14), p=0.060<br>**lag 1: 1.09 (1.03–1.15), p=0.001**<br>**lag 0: 1.05 (1.00–1.11), p=0.046** | **lag 2: 1.14 (1.05–1.23), p=0.002**<br>lag 0: 1.03 (0.94–1.12), p=0.516<br>lag 0: 0.95 (0.85–1.07), p=0.408 | **lag 1: 1.15 (1.02–1.30), p=0.021**<br>lag 1: 1.05 (0.95–1.18), p=0.340<br>**lag 2: 1.10 (1.01–1.21), p=0.030**<br>lag 6: 1.09 (1.00–1.18), p=0.051 | **lag 2: 1.09 (1.01–1.17), p=0.036**<br>lag 0: 0.99 (0.93–1.06), p=0.880<br>lag 0: 1.04 (0.97–1.11), p=0.240<br>**lag 0: 1.06 (1.00–1.12), p=0.046** | lag 7: 1.03 (0.94–1.13), p=0.494<br>lag 1: 1.09 (1.00–1.19), p=0.055<br>lag 0: 1.02 (0.93–1.11), p=0.697<br>lag 0: 1.05 (0.97–1.13), p=0.199 | lag 10: 1.06 (0.87–1.29), p=0.576<br>lag 8: 1.10 (0.95–1.27), p=0.203<br>**lag 1: 1.21 (1.05–1.38), p=0.007**<br>lag 1: 0.93 (0.81–1.06), p=0.249 | lag 0: 1.05 (0.99–1.12), p=0.129<br>**lag 0: 1.09 (1.03–1.15), p=0.004**<br>lag 0: 1.05 (0.98–1.12), p=0.138<br>**lag 0: 1.09 (1.03–1.14), p=0.002** | **lag 2: 1.19 (1.03–1.38), p=0.016**<br>lag 7: 1.14 (1.00–1.30), p=0.052<br>lag 2: 1.05 (0.94–1.17), p=0.389<br>lag 6: 1.06 (0.95–1.19), p=0.295 |

| Atmospheric factor | 1.1 | 1.2 | 2.1 | 2.2 | 3.1 | 4.1 | 5.1 | 5.2 | 6.1 | 6.2 | 6.3 |
|---|---|---|---|---|---|---|---|---|---|---|---|
| Wind chill index | lag 0: EDF=0.00, p=0.612<br>lag 0: EDF=0.68, p=0.071 | **lag 0: EDF=0.93, p=0.003**<br>**lag 0: EDF=3.59, p=<0.001** | - | lag 0: 0.99 (0.99–1.00), p=0.102 | lag 0: EDF=0.00, p=0.355<br>**lag 0: EDF=3.44, p=0.002** | lag 0: EDF=0.00, p=0.587<br>lag 0: EDF=0.00, p=0.665 | lag 0: EDF=0.01, p=0.319<br>lag 0: EDF=0.00, p=0.855 | - | lag 0: EDF=0.40, p=0.239 | lag 0: EDF=0.01, p=0.341<br>lag 0: EDF=0.01, p=0.635 | **lag 5: EDF=1.01, p=<0.001**<br>lag 0: EDF=0.38, p=0.209 |
| Heat stress wave intensity | **lag 5: EDF=0.86, p=0.008**<br>**lag 4: EDF=0.77, p=0.034**<br>lag 2: EDF=0.14, p=0.266<br>**lag 7: EDF=0.73, p=0.049** | lag 0: EDF=0.00, p=0.474<br>**lag 0: EDF=3.86, p=<0.001**<br>lag 0: EDF=0.01, p=0.329<br>lag 0: EDF=0.00, p=0.609 | lag 2: EDF=0.19, p=0.258<br>lag 3: EDF=0.26, p=0.240<br>lag 0: EDF=0.00, p=0.624<br>lag 0: EDF=0.35, p=0.204 | **lag 7: EDF=0.76, p=0.042**<br>**lag 0: EDF=0.82, p=0.012**<br>lag 0: EDF=0.00, p=0.849<br>lag 0: EDF=0.18, p=0.253 | lag 0: EDF=0.00, p=0.479<br>lag 4: EDF=0.00, p=0.466<br>**lag 9: EDF=0.82, p=0.025**<br>lag 2: EDF=0.48, p=0.165 | lag 10: EDF=0.35, p=0.250<br>lag 5: EDF=0.66, p=0.095<br>lag 0: EDF=0.00, p=0.852<br>lag 6: EDF=0.00, p=0.923 | **lag 10: EDF=0.87, p=0.006**<br>lag 2: EDF=0.47, p=0.161<br>lag 0: EDF=0.00, p=0.909<br>lag 0: EDF=0.71, p=0.050 | lag 0: EDF=0.00, p=0.457<br>lag 10: EDF=0.75, p=0.058<br>lag 1: EDF=0.65, p=0.094<br>lag 2: EDF=0.00, p=0.987 | lag 10: EDF=0.00, p=0.455<br>lag 5: EDF=0.00, p=0.767<br>lag 8: EDF=0.00, p=0.895<br>lag 0: EDF=0.00, p=0.551 | lag 1: EDF=0.00, p=0.562<br>lag 0: EDF=0.19, p=0.248<br>lag 0: EDF=0.56, p=0.134<br>lag 0: EDF=0.50, p=0.139 | lag 5: EDF=0.00, p=0.865<br>lag 7: EDF=0.60, p=0.168<br>lag 2: EDF=0.78, p=0.052<br>lag 2: EDF=0.00, p=0.424 |
| Cold wave | lag 0: 1.00 (0.89–1.12), p=0.990<br>**lag 5: 1.11 (1.01–1.23), p=0.037**<br>**lag 0: 1.13 (1.03–1.25), p=0.008**<br>lag 0: 1.02 (0.93–1.13), p=0.644 | **lag 0: 1.21 (1.09–1.35), p=<0.001**<br>lag 0: 1.06 (0.97–1.17), p=0.200<br>lag 0: 1.03 (0.93–1.13), p=0.616<br>**lag 0: 1.15 (1.06–1.25), p=0.001** | **lag 3: 1.14 (1.01–1.27), p=0.029**<br>lag 0: 0.91 (0.79–1.06), p=0.225<br>**lag 0: 1.13 (1.03–1.23), p=0.006**<br>lag 0: 0.98 (0.87–1.11), p=0.753 | **lag 1: 1.18 (1.07–1.29), p=<0.001**<br>lag 0: 1.00 (0.91–1.10), p=0.986<br>**lag 3: 1.13 (1.05–1.22), p=0.002**<br>lag 0: 0.93 (0.84–1.04), p=0.196 | lag 0: 1.01 (0.85–1.19), p=0.947<br>lag 2: 1.10 (0.92–1.31), p=0.285<br>**lag 2: 1.24 (1.09–1.42), p=0.002**<br>lag 1: 1.04 (0.90–1.20), p=0.589 | lag 5: 1.14 (0.98–1.33), p=0.081<br>lag 0: 1.07 (0.91–1.26), p=0.412<br>**lag 10: 1.19 (1.04–1.36), p=0.012**<br>lag 9: 1.07 (0.95–1.21), p=0.264 | **lag 9: 1.24 (1.08–1.43), p=0.003**<br>lag 0: 1.05 (0.94–1.17), p=0.431<br>**lag 0: 1.13 (1.01–1.25), p=0.027**<br>lag 0: 1.06 (0.95–1.18), p=0.296 | **lag 10: 1.20 (1.03–1.41), p=0.022**<br>lag 1: 1.07 (0.92–1.24), p=0.407<br>**lag 0: 1.35 (1.15–1.58), p=<0.001**<br>lag 2: 1.05 (0.92–1.19), p=0.477 | lag 8: 0.90 (0.69–1.19), p=0.472<br>lag 6: 1.02 (0.80–1.30), p=0.866<br>lag 7: 1.02 (0.81–1.29), p=0.849<br>lag 1: 1.10 (0.90–1.33), p=0.358 | lag 1: 1.07 (0.97–1.18), p=0.161<br>lag 0: 0.96 (0.88–1.05), p=0.367<br>**lag 0: 1.18 (1.07–1.31), p=<0.001**<br>lag 0: 1.02 (0.94–1.11), p=0.627 | lag 0: 0.94 (0.74–1.19), p=0.612<br>lag 4: 1.18 (0.93–1.48), p=0.169<br>lag 10: 1.16 (0.95–1.40), p=0.140<br>lag 9: 1.17 (0.95–1.43), p=0.131 |
| Cold wave duration | lag 8: EDF=0.00, p=0.354<br>lag 5: | lag 0: EDF=0.39, p=0.186<br>**lag 1:** | **lag 4: EDF=0.71, p=0.045**<br>**lag 0:** | lag 4: EDF=0.69, p=0.053 | lag 6: EDF=0.00, p=0.398 | - | **lag 6: EDF=0.81, p=0.017** | **lag 6: EDF=0.84, p=0.013**<br>**lag 3:** | lag 9: EDF=0.60, p=0.172<br>lag 5: | lag 1: EDF=0.00, p=0.987<br>**lag 3:** | **lag 0: EDF=0.72, p=0.042** |

| Atmospheric factor | 1.1 | 1.2 | 2.1 | 2.2 | 3.1 | 4.1 | 5.1 | 5.2 | 6.1 | 6.2 | 6.3 |
|---|---|---|---|---|---|---|---|---|---|---|---|
| | EDF=0.00, p=0.758<br>lag 9: EDF=0.16, p=0.180 | **EDF=0.95, p=<0.001**<br>lag 7: EDF=0.56, p=0.123 | **EDF=0.86, p=0.006**<br>**lag 0: EDF=0.78, p=0.030** | | | | | **EDF=0.95, p=<0.001**<br>lag 2: EDF=0.01, p=0.324 | EDF=0.00, p=0.619 | **EDF=0.89, p=0.002** | |
| Cold wave intensity | lag 5: EDF=0.00, p=0.776<br>lag 1: EDF=0.00, p=0.706 | **lag 10: EDF=0.94, p=<0.001**<br>lag 9: EDF=0.00, p=0.359 | lag 9: EDF=0.67, p=0.057<br>lag 0: EDF=0.00, p=0.485 | - | lag 2: EDF=0.00, p=0.938 | lag 10: EDF=0.40, p=0.196<br>lag 1: EDF=0.70, p=0.062 | lag 6: EDF=0.00, p=0.897<br>**lag 3: EDF=0.78, p=0.026** | lag 0: EDF=0.11, p=0.293 | lag 6: EDF=0.00, p=0.492<br>lag 0: EDF=0.61, p=0.153<br>lag 0: EDF=0.69, p=0.124<br>lag 1: EDF=0.01, p=0.350 | lag 0: EDF=0.57, p=0.123 | lag 10: EDF=0.00, p=0.628<br>**lag 1: EDF=0.93, p=0.001**<br>lag 10: EDF=0.48, p=0.152 |
| Cold stress wave duration | lag 0: EDF=0.00, p=0.991<br>lag 10: EDF=0.62, p=0.077<br>lag 6: EDF=0.49, p=0.117 | lag 0: EDF=0.00, p=0.782 | - | lag 1: EDF=0.00, p=0.616<br>**lag 9: EDF=0.92, p=<0.001**<br>lag 1: EDF=0.00, p=0.335<br>**lag 0: EDF=0.72, p=0.046** | lag 10: EDF=0.71, p=0.070<br>**lag 1: EDF=2.63, p=<0.001**<br>**lag 0: EDF=0.86, p=0.005** | **lag 10: EDF=0.77, p=0.041**<br>lag 0: EDF=0.39, p=0.207<br>lag 10: EDF=0.00, p=0.623 | lag 8: EDF=0.70, p=0.061<br>**lag 10: EDF=0.78, p=0.024**<br>**lag 10: EDF=0.84, p=0.009**<br>lag 0: EDF=0.17, p=0.248 | lag 7: EDF=0.58, p=0.122 | - | lag 6: EDF=0.59, p=0.107<br>lag 8: EDF=0.00, p=0.972 | **lag 6: EDF=0.80, p=0.024** |
| Cold stress wave intensity | **lag 10: EDF=0.89, p=0.002**<br>lag 2: EDF=0.00, p=0.639<br>**lag 8: EDF=0.81, p=0.012** | **lag 7: EDF=0.89, p=0.002**<br>lag 5: EDF=0.00, p=0.365<br>lag 10: EDF=0.00, p=0.761 | **lag 7: EDF=0.89, p=0.001** | lag 7: EDF=0.44, p=0.149<br>lag 4: EDF=0.61, p=0.091<br>lag 1: EDF=0.63, p=0.081 | lag 8: EDF=0.00, p=0.350 | - | lag 8: EDF=0.00, p=0.355<br>**lag 10: EDF=0.82, p=0.011** | **lag 0: EDF=0.88, p=0.005** | lag 10: EDF=0.00, p=0.837<br>lag 7: EDF=0.00, p=0.552 | lag 10: EDF=0.00, p=0.455<br>**lag 4: EDF=2.01, p=0.016** | **lag 6: EDF=0.94, p=<0.001**<br>lag 9: EDF=0.55, p=0.166<br>lag 2: EDF=0.55, p=0.119 |

| Atmospheric factor | 1.1 | 1.2 | 2.1 | 2.2 | 3.1 | 4.1 | 5.1 | 5.2 | 6.1 | 6.2 | 6.3 |
|---|---|---|---|---|---|---|---|---|---|---|---|
| Ozone | **lag 0: EDF=4.20, p=0.004**<br>lag 0: EDF=0.00, p=0.698<br>lag 0: EDF=0.00, p=0.988<br>lag 0: EDF=0.01, p=0.390 | **lag 0: EDF=2.07, p=0.041**<br>lag 0: EDF=0.01, p=0.300<br>lag 0: EDF=0.00, p=0.815<br>lag 0: EDF=0.69, p=0.069 | lag 0: EDF=0.73, p=0.054<br>**lag 0: EDF=0.82, p=0.029** | lag 0: 1.00 (1.00–1.01), p=0.152<br>**lag 0: EDF=2.36, p=0.004**<br>lag 0: EDF=0.05, p=0.309<br>**lag 0: EDF=3.21, p=<0.001** | lag 7: EDF=0.00, p=0.699<br>lag 1: EDF=0.00, p=0.386<br>lag 0: EDF=0.72, p=0.070 | lag 1: EDF=0.00, p=0.475<br>**lag 0: EDF=0.93, p=0.003**<br>lag 4: EDF=2.74, p=0.067<br>lag 1: EDF=0.00, p=0.570 | **lag 0: EDF=2.16, p=0.035**<br>lag 0: EDF=0.00, p=0.373<br>lag 0: EDF=0.57, p=0.122<br>lag 0: EDF=0.00, p=0.387 | lag 0: EDF=0.00, p=0.464<br>lag 3: EDF=0.01, p=0.322 | lag 6: EDF=0.00, p=0.871<br>**lag 6: EDF=0.85, p=0.036**<br>lag 5: EDF=0.01, p=0.391<br>**lag 1: EDF=0.92, p=0.007** | **lag 0: EDF=4.47, p=<0.001**<br>lag 0: EDF=0.39, p=0.194<br>**lag 0: EDF=4.05, p=<0.001** | **lag 7: EDF=0.82, p=0.050**<br>**lag 0: EDF=0.87, p=0.022**<br>lag 5: EDF=0.75, p=0.058<br>lag 6: EDF=0.00, p=0.463 |
| Nitrogen dioxide | **lag 2: EDF=2.28, p=0.003**<br>lag 0: EDF=0.11, p=0.277<br>lag 0: EDF=0.54, p=0.125<br>lag 5: EDF=0.65, p=0.081 | lag 4: EDF=0.66, p=0.081<br>lag 7: EDF=0.23, p=0.234<br>lag 0: EDF=0.00, p=0.641<br>lag 7: EDF=0.24, p=0.232 | lag 6: EDF=0.53, p=0.134<br>lag 0: EDF=0.42, p=0.173<br>lag 0: EDF=0.66, p=0.080<br>lag 0: EDF=0.00, p=0.772 | lag 2: EDF=0.00, p=0.428<br>**lag 0: EDF=0.90, p=<0.001**<br>lag 0: EDF=0.42, p=0.177<br>**lag 5: EDF=0.89, p=0.003** | lag 7: EDF=0.00, p=0.953<br>lag 0: EDF=0.00, p=0.670<br>lag 1: EDF=0.00, p=0.959<br>lag 0: EDF=0.39, p=0.184 | **lag 0: EDF=0.79, p=0.034**<br>lag 1: EDF=0.00, p=0.406<br>lag 6: EDF=0.62, p=0.102<br>lag 5: EDF=0.23, p=0.239 | **lag 3: EDF=0.77, p=0.034**<br>lag 3: EDF=0.18, p=0.249<br>lag 3: EDF=0.48, p=0.151<br>lag 0: EDF=0.00, p=0.798 | **lag 7: EDF=0.87, p=0.008**<br>lag 1: EDF=0.00, p=0.461<br>lag 1: EDF=0.04, p=0.328<br>lag 5: EDF=0.53, p=0.138 | **lag 2: EDF=4.65, p=0.004**<br>lag 6: EDF=0.00, p=0.892<br>**lag 5: EDF=0.87, p=0.014**<br>lag 1: EDF=0.00, p=0.808 | lag 3: EDF=0.00, p=0.370<br>lag 0: EDF=0.00, p=0.801<br>lag 0: EDF=0.00, p=0.756<br>**lag 2: EDF=0.78, p=0.031** | lag 6: EDF=0.74, p=0.084<br>lag 0: EDF=0.00, p=0.913<br>lag 5: EDF=0.12, p=0.294<br>**lag 6: EDF=0.78, p=0.040** |

**Table S18.** Overview of time lags, effect sizes (effective degree of freedom (EDF) or relative risk (95% confidence interval)) and p-values per human-bioclimatic region, atmospheric factor and subgroup (pink = female <60 years, orange = females ≥60 years, green = males <60 years, blue = males ≥60 years) for myocardial infarction (STEMI), significant p-values (≤ 0.05) are bold

| Atmospheric factor | 1.1 | 1.2 | 2.1 | 2.2 | 3.1 | 4.1 | 5.1 | 5.2 | 6.1 | 6.2 | 6.3 |
|---|---|---|---|---|---|---|---|---|---|---|---|
| Sea-level pressure (SLP) | **lag 0: EDF=0.81, p=0.027**<br>lag 2: EDF=0.65, p=0.076 | lag 10: EDF=0.67, p=0.073<br>lag 4: EDF=0.23, p=0.234 | lag 7: EDF=0.46, p=0.165<br>**lag 3: EDF=0.80, p=0.020** | **lag 3: EDF=0.88, p=0.006**<br>lag 9: EDF=0.71, p=0.051 | lag 10: EDF=0.70, p=0.127<br>lag 0: EDF=0.00, p=0.532 | **lag 10: EDF=0.91, p=0.011**<br>lag 6: EDF=0.65, p=0.074 | **lag 10: EDF=0.80, p=0.040**<br>lag 1: EDF=0.58, p=0.101 | **lag 7: EDF=3.17, p=0.022**<br>lag 8: EDF=0.00, p=0.339 | lag 7: EDF=0.90, p=0.084<br>lag 7: EDF=0.01, p=0.424 | lag 6: EDF=0.20, p=0.257<br>**lag 2: EDF=0.85, p=0.009** | **lag 8: EDF=3.45, p=0.016**<br>**lag 5: EDF=0.92, p=0.001** |

| Atmospheric factor | 1.1 | 1.2 | 2.1 | 2.2 | 3.1 | 4.1 | 5.1 | 5.2 | 6.1 | 6.2 | 6.3 |
|---|---|---|---|---|---|---|---|---|---|---|---|
| | lag 3: EDF=0.59, p=0.105<br>**lag 9: EDF=0.87, p=0.006** | **lag 3: EDF=2.47, p=<0.001**<br>**lag 3: EDF=0.82, p=0.018** | lag 3: EDF=0.68, p=0.068<br>lag 2: EDF=0.58, p=0.112 | lag 0: EDF=0.59, p=0.104<br>lag 4: EDF=0.00, p=0.346 | **lag 3: EDF=0.74, p=0.042**<br>lag 0: EDF=0.59, p=0.093 | lag 0: EDF=0.55, p=0.115<br>**lag 6: EDF=0.75, p=0.035** | lag 7: EDF=0.68, p=0.060<br>**lag 2: EDF=0.82, p=0.016** | **lag 10: EDF=0.86, p=0.007**<br>lag 9: EDF=0.01, p=0.300 | **lag 9: EDF=0.88, p=0.020**<br>**lag 8: EDF=0.93, p=0.002** | **lag 6: EDF=0.86, p=0.007**<br>**lag 3: EDF=0.82, p=0.018** | lag 5: EDF=0.49, p=0.163<br>lag 5: EDF=0.00, p=0.351 |
| Tropical night | **lag 10: 2.29 (1.15–4.52), p=0.018**<br>lag 8: 1.40 (0.88–2.24), p=0.157<br>lag 8: 0.47 (0.21–1.04), p=0.064<br>lag 2: 0.55 (0.29–1.04), p=0.068 | lag 2: 0.56 (0.25–1.26), p=0.160<br>lag 0: 0.73 (0.50–1.06), p=0.098<br>lag 1: 0.80 (0.59–1.06), p=0.124<br>lag 0: 0.78 (0.58–1.05), p=0.099 | lag 2: 2.02 (0.73–5.59), p=0.174<br>lag 5: 1.47 (0.84–2.58), p=0.178<br>lag 0: 1.57 (0.91–2.73), p=0.106<br>lag 7: 0.51 (0.20–1.29), p=0.152 | lag 7: 0.64 (0.29–1.39), p=0.256<br>lag 2: 0.81 (0.58–1.13), p=0.215<br>lag 6: 0.87 (0.65–1.16), p=0.344<br>lag 2: 0.88 (0.67–1.16), p=0.377 | - | lag 6: 2.88 (0.37–22.09), p=0.310<br>lag 2: 2.47 (0.99–6.19), p=0.054<br>**lag 1: 3.04 (1.20–7.73), p=0.019**<br>lag 5: 1.77 (0.63–4.96), p=0.279 | **lag 0: 6.28 (1.90–20.78), p=0.003**<br>lag 5: 1.87 (0.83–4.21), p=0.129<br>**lag 3: 2.93 (1.54–5.59), p=0.001**<br>lag 2: 1.25 (0.57–2.74), p=0.582 | lag 8: 2.19 (0.28–16.96), p=0.455<br>lag 1: 1.40 (0.50–3.91), p=0.516<br>lag 1: 2.08 (0.81–5.36), p=0.130<br>**lag 1: 2.29 (1.14–4.59), p=0.020** | lag 9: 2.27 (0.28–18.04), p=0.440<br>lag 8: 1.49 (0.53–4.22), p=0.448<br>lag 5: 2.67 (0.92–7.75), p=0.071<br>**lag 3: 3.06 (1.40–6.68), p=0.005** | **lag 5: 1.84 (1.08–3.14), p=0.026**<br>**lag 10: 1.33 (1.03–1.72), p=0.031**<br>**lag 2: 1.49 (1.12–1.96), p=0.005**<br>**lag 3: 0.69 (0.50–0.95), p=0.022** | **lag 10: 3.25 (1.36–7.75), p=0.008**<br>**lag 3: 2.25 (1.20–4.22), p=0.011**<br>**lag 10: 2.51 (1.43–4.42), p=0.001**<br>lag 10: 0.40 (0.12–1.27), p=0.120 |
| Hot day | **lag 0: 0.58 (0.36–0.92), p=0.020**<br>lag 3: 0.95 (0.79–1.15), p=0.623<br>lag 8: 0.83 (0.68–1.01), p=0.063<br>**lag 10: 0.76 (0.64–0.90), p=0.001** | lag 3: 0.88 (0.67–1.17), p=0.392<br>lag 2: 0.89 (0.76–1.04), p=0.149<br>lag 8: 0.93 (0.82–1.06), p=0.270<br>lag 2: 0.95 (0.84–1.07), p=0.386 | lag 1: 0.85 (0.65–1.11), p=0.229<br>**lag 9: 0.83 (0.73–0.94), p=0.004**<br>lag 9: 0.90 (0.80–1.01), p=0.073<br>**lag 6: 0.89 (0.79–0.99), p=0.037** | lag 6: 1.01 (0.77–1.32), p=0.958<br>**lag 5: 0.80 (0.70–0.92), p=0.001**<br>lag 4: 0.91 (0.82–1.02), p=0.122<br>lag 2: 0.94 (0.84–1.05), p=0.282 | **lag 1: 0.21 (0.05–0.89), p=0.034**<br>lag 2: 0.73 (0.51–1.04), p=0.083<br>**lag 5: 0.68 (0.47–1.00), p=0.050**<br>lag 2: 0.91 (0.70–1.17), p=0.456 | lag 6: 2.03 (0.81–5.06), p=0.130<br>lag 4: 1.56 (0.98–2.50), p=0.061<br>lag 6: 0.48 (0.19–1.20), p=0.117<br>**lag 4: 1.57 (1.03–2.40), p=0.036** | lag 7: 0.70 (0.39–1.24), p=0.216<br>lag 5: 0.93 (0.73–1.17), p=0.521<br>lag 7: 0.79 (0.62–1.02), p=0.070<br>**lag 7: 0.72 (0.58–0.88), p=0.002** | lag 5: 0.45 (0.19–1.08), p=0.072<br>**lag 10: 0.68 (0.51–0.91), p=0.009**<br>lag 8: 1.24 (0.96–1.59), p=0.097<br>lag 4: 0.89 (0.70–1.13), p=0.338 | **lag 8: 2.74 (1.00–7.49), p=0.049**<br>**lag 3: 1.98 (1.36–2.87), p=<0.001**<br>lag 7: 1.53 (0.94–2.49), p=0.088<br>lag 8: 0.90 (0.55–1.46), p=0.662 | lag 3: 0.78 (0.57–1.06), p=0.110<br>lag 1: 0.90 (0.80–1.01), p=0.080<br>lag 6: 0.96 (0.84–1.10), p=0.543<br>lag 1: 0.99 (0.89–1.11), p=0.888 | **lag 1: 1.81 (1.16–2.81), p=0.008**<br>lag 4: 0.80 (0.58–1.10), p=0.175<br>lag 3: 0.95 (0.70–1.28), p=0.737<br>lag 1: 0.82 (0.61–1.09), p=0.174 |

| Atmospheric factor | 1.1 | 1.2 | 2.1 | 2.2 | 3.1 | 4.1 | 5.1 | 5.2 | 6.1 | 6.2 | 6.3 |
|---|---|---|---|---|---|---|---|---|---|---|---|
| Summer day | lag 1: 0.92 (0.78–1.09), p=0.335<br>**lag 9: 0.91 (0.84–0.99), p=0.036**<br>lag 8: 0.95 (0.87–1.03), p=0.193<br>**lag 4: 0.87 (0.81–0.93), p=<0.001** | lag 10: 0.98 (0.87–1.10), p=0.678<br>lag 2: 0.97 (0.90–1.05), p=0.466<br>lag 9: 0.95 (0.89–1.01), p=0.075<br>lag 2: 0.97 (0.91–1.03), p=0.288 | **lag 10: 0.86 (0.76–0.96), p=0.010**<br>**lag 1: 0.91 (0.85–0.96), p=0.001**<br>lag 5: 0.97 (0.92–1.02), p=0.263<br>**lag 7: 0.95 (0.90–1.00), p=0.044** | lag 8: 0.88 (0.77–1.01), p=0.068<br>lag 3: 0.95 (0.89–1.01), p=0.098<br>**lag 9: 0.95 (0.89–1.00), p=0.045**<br>lag 2: 0.95 (0.90–1.01), p=0.086 | lag 2: 0.82 (0.63–1.07), p=0.144<br>**lag 7: 0.88 (0.78–0.99), p=0.034**<br>**lag 6: 0.88 (0.78–0.99), p=0.032**<br>**lag 7: 0.88 (0.80–0.97), p=0.008** | **lag 2: 0.65 (0.44–0.95), p=0.025**<br>lag 2: 0.95 (0.81–1.11), p=0.510<br>**lag 2: 0.81 (0.68–0.97), p=0.020**<br>**lag 5: 0.79 (0.68–0.91), p=0.001** | **lag 3: 0.72 (0.57–0.91), p=0.006**<br>**lag 6: 0.90 (0.82–0.99), p=0.029**<br>lag 9: 0.93 (0.84–1.03), p=0.149<br>lag 2: 0.93 (0.86–1.01), p=0.082 | lag 6: 0.79 (0.60–1.05), p=0.108<br>lag 3: 0.92 (0.82–1.04), p=0.183<br>**lag 6: 0.84 (0.73–0.95), p=0.007**<br>lag 4: 0.92 (0.83–1.02), p=0.101 | **lag 8: 0.51 (0.28–0.92), p=0.024**<br>**lag 7: 0.80 (0.65–0.99), p=0.040**<br>lag 1: 0.79 (0.61–1.02), p=0.072<br>lag 7: 0.83 (0.68–1.01), p=0.070 | lag 3: 0.91 (0.79–1.04), p=0.175<br>**lag 4: 0.93 (0.88–0.98), p=0.013**<br>**lag 7: 0.94 (0.88–0.99), p=0.033**<br>lag 1: 0.97 (0.92–1.02), p=0.186 | lag 10: 1.24 (0.95–1.61), p=0.111<br>lag 1: 0.95 (0.83–1.09), p=0.472<br>**lag 6: 0.85 (0.74–0.98), p=0.028**<br>lag 2: 0.91 (0.81–1.02), p=0.119 |
| Cold wave duration | lag 0: EDF=0.00, p=0.643<br>lag 0: EDF=0.00, p=0.923<br>lag 7: EDF=0.00, p=0.643<br>lag 0: EDF=0.00, p=0.708 | lag 2: EDF=0.21, p=0.233<br>lag 5: EDF=0.01, p=0.335<br>lag 8: EDF=0.00, p=0.340<br>**lag 0: EDF=0.90, p=0.001** | **lag 0: EDF=0.85, p=0.008**<br>**lag 6: EDF=0.90, p=<0.001**<br>lag 0: EDF=0.00, p=0.533<br>lag 3: EDF=0.57, p=0.109 | lag 4: EDF=0.00, p=0.323<br>lag 10: EDF=0.00, p=0.738<br>lag 10: EDF=0.00, p=0.462<br>**lag 3: EDF=0.89, p=0.002** | lag 9: EDF=0.32, p=0.291<br>lag 3: EDF=0.00, p=0.313<br>lag 8: EDF=0.00, p=0.971<br>lag 4: EDF=0.00, p=0.694 | lag 4: EDF=0.03, p=0.388<br>lag 0: EDF=0.00, p=0.503<br>lag 10: EDF=0.00, p=0.803<br>lag 10: EDF=0.00, p=0.698 | **lag 10: EDF=0.83, p=0.027**<br>lag 0: EDF=0.64, p=0.071<br>lag 1: EDF=0.00, p=0.505<br>lag 2: EDF=0.28, p=0.206 | lag 8: EDF=0.83, p=0.053<br>lag 8: EDF=0.00, p=0.665<br>lag 6: EDF=0.56, p=0.112<br>lag 6: EDF=0.53, p=0.118 | **lag 0: EDF=0.94, p=0.017**<br>lag 4: EDF=0.01, p=0.391<br>**lag 9: EDF=0.86, p=0.019**<br>lag 9: EDF=0.50, p=0.171 | lag 10: EDF=0.00, p=0.852<br>lag 6: EDF=0.00, p=0.571<br>lag 4: EDF=0.38, p=0.177<br>lag 7: EDF=0.00, p=0.354 | lag 7: EDF=0.83, p=0.071<br>lag 10: EDF=0.00, p=0.425<br>lag 6: EDF=0.00, p=0.749<br>lag 0: EDF=0.00, p=0.548 |
| Ozone | **lag 3: EDF=0.78, p=0.040**<br>lag 6: EDF=0.00, p=0.727<br>lag 4: EDF=0.57, p=0.113<br>**lag 1:** | **lag 3: EDF=0.79, p=0.026**<br>**lag 5: EDF=0.84, p=0.009**<br>lag 7: EDF=0.03, p=0.294<br>lag 1: | lag 0: EDF=0.00, p=0.542<br>lag 6: EDF=0.00, p=0.363<br>lag 5: EDF=0.00, p=0.833<br>**lag 3:** | **lag 6: EDF=0.92, p=0.002**<br>lag 0: EDF=0.53, p=0.091<br>lag 7: EDF=0.00, p=0.647<br>**lag 1:** | lag 7: EDF=0.61, p=0.178<br>lag 3: EDF=0.64, p=0.085<br>lag 7: EDF=0.00, p=0.571<br>lag 7: | lag 5: EDF=0.00, p=0.921<br>lag 1: EDF=0.64, p=0.057<br>**lag 1: EDF=0.76, p=0.033**<br>lag 3: | **lag 4: EDF=0.82, p=0.016**<br>lag 5: EDF=0.70, p=0.051<br>lag 6: EDF=0.00, p=0.865<br>lag 1: | lag 7: EDF=0.00, p=0.577<br>**lag 5: EDF=0.75, p=0.037**<br>lag 2: EDF=0.00, p=0.479<br>lag 7: | lag 3: EDF=0.00, p=0.753<br>lag 7: EDF=0.12, p=0.304<br>**lag 5: EDF=0.80, p=0.048**<br>**lag 7:** | **lag 2: EDF=0.69, p=0.043**<br>**lag 4: EDF=0.86, p=0.008**<br>lag 1: EDF=0.00, p=0.437<br>lag 7: | **lag 7: 0.98 (0.97–1.00), p=0.006**<br>lag 0: EDF=0.00, p=0.503<br>lag 6: EDF=0.24, p=0.198<br>lag 7: |

| Atmospheric factor | 1.1 | 1.2 | 2.1 | 2.2 | 3.1 | 4.1 | 5.1 | 5.2 | 6.1 | 6.2 | 6.3 |
|---|---|---|---|---|---|---|---|---|---|---|---|
| | **EDF=0.67, p=0.043** | EDF=0.00, p=0.722 | **EDF=1.96, p=0.026** | **EDF=0.77, p=0.018** | EDF=0.17, p=0.238 | EDF=0.35, p=0.148 | EDF=0.64, p=0.078 | EDF=0.00, p=0.463 | **EDF=0.81, p=0.033** | EDF=0.19, p=0.238 | EDF=0.00, p=0.786 |

**Table S19.** Overview of time lags, effect sizes (effective degree of freedom (EDF) or relative risk (95% confidence interval)) and p-values per human-bioclimatic region, atmospheric factor and subgroup (pink = female <60 years, orange = females ≥60 years, green = males <60 years, blue = males ≥60 years) for sepsis, significant p-values (≤ 0.05) are bold

| Atmospheric factor | 1.1 | 1.2 | 2.1 | 2.2 | 3.1 | 4.1 | 5.1 | 5.2 | 6.1 | 6.2 | 6.3 |
|---|---|---|---|---|---|---|---|---|---|---|---|
| Sea-level pressure (SLP) | lag 4: EDF=0.65, p=0.089<br>lag 5: EDF=0.00, p=0.301<br>lag 8: EDF=0.69, p=0.061<br>lag 4: EDF=0.01, p=0.487 | **lag 8: EDF=0.91, p=0.001**<br>lag 7: EDF=0.00, p=0.873<br>**lag 1: EDF=0.80, p=0.021**<br>lag 6: EDF=0.43, p=0.175 | lag 3: EDF=0.35, p=0.194<br>lag 10: EDF=0.00, p=0.770<br>**lag 9: EDF=0.88, p=0.003**<br>lag 6: EDF=0.23, p=0.244 | **lag 3: EDF=0.84, p=0.011**<br>lag 3: EDF=0.00, p=0.380<br>lag 0: EDF=0.00, p=0.866<br>lag 8: EDF=0.00, p=0.676 | **lag 6: EDF=0.86, p=0.019**<br>**lag 0: EDF=0.88, p=0.004**<br>lag 4: EDF=0.00, p=0.414<br>lag 6: EDF=0.00, p=0.892 | lag 5: EDF=0.16, p=0.315<br>lag 10: EDF=0.57, p=0.111<br>lag 7: EDF=0.00, p=0.373<br>lag 2: EDF=0.00, p=0.512 | lag 4: EDF=0.65, p=0.094<br>lag 4: EDF=0.68, p=0.064<br>**lag 4: EDF=0.78, p=0.030**<br>lag 3: EDF=0.02, p=0.302 | lag 5: EDF=0.53, p=0.183<br>lag 1: EDF=0.60, p=0.096<br>**lag 6: EDF=0.81, p=0.021**<br>**lag 0: EDF=0.86, p=0.006** | lag 0: EDF=0.66, p=0.237<br>lag 1: EDF=0.01, p=0.441<br>lag 6: EDF=0.32, p=0.307<br>**lag 0: EDF=0.94, p=<0.001** | lag 9: EDF=0.00, p=0.726<br>**lag 7: EDF=0.90, p=0.003**<br>lag 4: EDF=0.00, p=0.627<br>**lag 0: EDF=0.88, p=0.005** | lag 10: EDF=0.59, p=0.211<br>lag 6: EDF=0.00, p=0.575<br>lag 10: EDF=0.62, p=0.139<br>**lag 9: EDF=0.78, p=0.034** |
| Sea-level pressure difference (SLPD) | lag 7: 1.08 (0.97–1.21), p=0.151<br>**lag 7: 0.93 (0.87–0.99), p=0.021**<br>lag 8: 0.94 (0.87–1.02), p=0.157<br>**lag 1: 0.93 (0.88–0.98), p=0.008** | **lag 1: 0.88 (0.81–0.96), p=0.004**<br>**lag 6: 0.95 (0.90–1.00), p=0.040**<br>**lag 1: 0.92 (0.86–0.98), p=0.010**<br>**lag 1: 0.94 (0.90–0.98), p=0.007** | **lag 5: 0.85 (0.76–0.95), p=0.006**<br>lag 3: 0.96 (0.90–1.02), p=0.174<br>lag 0: 0.95 (0.87–1.04), p=0.256<br>**lag 9: 0.93 (0.89–0.99), p=0.014** | lag 3: 0.96 (0.86–1.08), p=0.521<br>**lag 1: 0.90 (0.84–0.96), p=<0.001**<br>**lag 5: 0.89 (0.81–0.96), p=0.005**<br>lag 1: 0.96 (0.91–1.01), p=0.127 | lag 9: 0.84 (0.68–1.04), p=0.118<br>lag 5: 1.08 (0.97–1.21), p=0.157<br>lag 1: 0.85 (0.72–1.00), p=0.054<br>**lag 0: 0.91 (0.83–0.99), p=0.036** | lag 10: 0.91 (0.78–1.06), p=0.232<br>**lag 10: 0.91 (0.83–0.99), p=0.031**<br>lag 5: 0.91 (0.82–1.02), p=0.108<br>lag 3: 1.04 (0.97–1.12), p=0.303 | **lag 1: 1.18 (1.02–1.36), p=0.023**<br>lag 5: 0.94 (0.88–1.02), p=0.129<br>lag 4: 0.91 (0.82–1.01), p=0.087<br>lag 10: 0.95 (0.89–1.01), p=0.106 | lag 6: 1.19 (0.99–1.43), p=0.057<br>**lag 4: 0.89 (0.81–0.97), p=0.012**<br>**lag 0: 1.15 (1.01–1.30), p=0.032**<br>lag 2: 1.06 (0.98–1.15), p=0.127 | **lag 7: 0.72 (0.55–0.95), p=0.022**<br>**lag 2: 1.16 (1.02–1.32), p=0.026**<br>lag 7: 0.83 (0.68–1.01), p=0.057<br>**lag 9: 0.85 (0.76–0.95), p=0.006** | lag 9: 1.06 (0.97–1.16), p=0.206<br>lag 7: 0.98 (0.93–1.03), p=0.377<br>lag 10: 0.95 (0.88–1.01), p=0.115<br>lag 1: 1.04 (1.00–1.08), p=0.053 | **lag 7: 0.78 (0.63–0.97), p=0.025**<br>**lag 9: 1.13 (1.00–1.27), p=0.047**<br>lag 2: 1.13 (0.97–1.32), p=0.120<br>lag 4: 0.90 (0.81–1.00), p=0.060 |

| Atmospheric factor | 1.1 | 1.2 | 2.1 | 2.2 | 3.1 | 4.1 | 5.1 | 5.2 | 6.1 | 6.2 | 6.3 |
|---|---|---|---|---|---|---|---|---|---|---|---|
| Wind day | lag 7: 1.08 (0.94–1.24), p=0.273<br>**lag 3: 1.08 (1.00–1.17), p=0.047**<br>**lag 6: 1.11 (1.00–1.22), p=0.044**<br>lag 1: 1.03 (0.96–1.10), p=0.428 | lag 9: 1.07 (0.96–1.20), p=0.229<br>**lag 6: 1.07 (1.00–1.15), p=0.049**<br>lag 9: 1.08 (0.99–1.17), p=0.094<br>lag 8: 1.03 (0.97–1.09), p=0.347 | lag 3: 1.08 (0.96–1.22), p=0.216<br>lag 2: 1.06 (1.00–1.14), p=0.065<br>**lag 7: 1.18 (1.08–1.29), p=<0.001**<br>lag 10: 1.04 (0.98–1.10), p=0.186 | lag 0: 1.07 (0.95–1.21), p=0.265<br>lag 9: 1.06 (0.99–1.13), p=0.104<br>**lag 7: 1.12 (1.03–1.23), p=0.010**<br>lag 0: 1.02 (0.96–1.08), p=0.502 | lag 10: 1.20 (0.97–1.48), p=0.095<br>lag 5: 1.11 (1.00–1.24), p=0.059<br>**lag 5: 1.18 (1.01–1.38), p=0.041**<br>**lag 3: 1.11 (1.01–1.22), p=0.023** | lag 8: 1.13 (0.93–1.37), p=0.210<br>**lag 0: 0.88 (0.78–0.99), p=0.036**<br>lag 7: 0.88 (0.75–1.03), p=0.113<br>lag 10: 1.03 (0.94–1.14), p=0.498 | lag 9: 1.15 (0.98–1.34), p=0.087<br>lag 8: 1.03 (0.95–1.12), p=0.488<br>**lag 10: 1.13 (1.00–1.26), p=0.043**<br>lag 8: 1.05 (0.98–1.13), p=0.181 | lag 6: 1.18 (0.95–1.45), p=0.129<br>lag 9: 1.06 (0.96–1.18), p=0.240<br>lag 9: 1.15 (1.00–1.32), p=0.052<br>lag 1: 1.04 (0.95–1.13), p=0.397 | lag 4: 1.27 (0.91–1.77), p=0.160<br>**lag 4: 0.81 (0.68–0.98), p=0.031**<br>**lag 10: 1.34 (1.07–1.68), p=0.010**<br>lag 7: 1.13 (0.98–1.30), p=0.083 | **lag 3: 1.15 (1.04–1.28), p=0.009**<br>lag 10: 0.95 (0.90–1.01), p=0.083<br>lag 7: 1.02 (0.94–1.10), p=0.666<br>lag 9: 1.03 (0.98–1.08), p=0.274 | **lag 6: 1.32 (1.02–1.71), p=0.032**<br>lag 8: 1.08 (0.93–1.27), p=0.314<br>lag 5: 1.13 (0.93–1.38), p=0.222<br>lag 9: 1.14 (0.99–1.31), p=0.065 |
| Summer day | lag 4: 1.05 (0.89–1.23), p=0.543<br>lag 2: 1.01 (0.92–1.11), p=0.853<br>lag 0: 1.09 (0.97–1.23), p=0.138<br>lag 1: 1.02 (0.94–1.10), p=0.631 | lag 0: 1.00 (0.90–1.12), p=0.975<br>**lag 0: 1.09 (1.03–1.15), p=0.004**<br>lag 7: 1.00 (0.92–1.08), p=0.909<br>lag 0: 1.02 (0.96–1.08), p=0.495 | **lag 10: 0.91 (0.83–1.00), p=0.047**<br>lag 2: 1.03 (0.97–1.09), p=0.404<br>**lag 0: 1.09 (1.02–1.16), p=0.013**<br>**lag 0: 1.06 (1.02–1.11), p=0.005** | **lag 10: 0.89 (0.81–0.98), p=0.013**<br>**lag 2: 1.10 (1.05–1.16), p=<0.001**<br>**lag 0: 1.15 (1.06–1.25), p=<0.001**<br>lag 9: 1.04 (1.00–1.08), p=0.074 | lag 1: 0.84 (0.68–1.04), p=0.112<br>lag 7: 0.96 (0.86–1.07), p=0.455<br>lag 2: 1.14 (0.95–1.37), p=0.154<br>lag 4: 1.05 (0.98–1.14), p=0.183 | lag 6: 1.16 (0.87–1.53), p=0.309<br>**lag 10: 0.83 (0.72–0.97), p=0.015**<br>lag 8: 1.15 (0.98–1.36), p=0.095<br>lag 6: 1.05 (0.94–1.18), p=0.377 | lag 5: 1.16 (0.97–1.39), p=0.114<br>**lag 0: 1.10 (1.00–1.21), p=0.049**<br>lag 10: 1.03 (0.90–1.17), p=0.681<br>lag 2: 1.05 (0.99–1.12), p=0.118 | lag 8: 0.82 (0.66–1.03), p=0.085<br>lag 1: 1.08 (0.99–1.17), p=0.077<br>lag 10: 1.04 (0.90–1.20), p=0.621<br>**lag 3: 1.11 (1.03–1.19), p=0.004** | lag 0: 1.18 (0.80–1.75), p=0.398<br>**lag 0: 1.22 (1.06–1.40), p=0.005**<br>lag 1: 0.80 (0.58–1.12), p=0.190<br>lag 7: 0.89 (0.75–1.05), p=0.181 | lag 3: 0.95 (0.87–1.03), p=0.205<br>lag 9: 1.03 (0.99–1.07), p=0.214<br>lag 5: 1.05 (0.99–1.11), p=0.110<br>**lag 1: 1.08 (1.04–1.12), p=<0.001** | **lag 8: 0.73 (0.56–0.94), p=0.015**<br>**lag 0: 1.14 (1.01–1.29), p=0.040**<br>lag 8: 0.93 (0.77–1.13), p=0.472<br>lag 7: 1.11 (1.00–1.23), p=0.050 |
| Wet-bulb globe temperature | **lag 10: 0.99 (0.98–0.99), p=<0.001**<br>lag 8: | lag 10: EDF=0.72, p=0.052 | - | lag 7: EDF=0.00, p=0.434<br>lag 0: EDF=0.44, p=0.144 | lag 10: EDF=0.00, p=0.603<br>lag 8: EDF=0.35, p=0.193 | lag 1: EDF=0.00, p=0.361<br>**lag 9: EDF=0.84, p=0.011**<br>lag 0: | lag 7: EDF=0.69, p=0.080<br>**lag 6: EDF=0.81, p=0.016**<br>**lag 10:** | lag 6: EDF=3.27, p=0.070<br>lag 3: EDF=0.02, p=0.310<br>**lag 7:** | **lag 10: 0.98 (0.96–1.00), p=0.016** | lag 7: EDF=0.00, p=0.498<br>**lag 1: EDF=0.84, p=0.014** | lag 9: EDF=0.70, p=0.100<br>lag 10: EDF=0.01, p=0.392 |

| Atmospheric factor | 1.1 | 1.2 | 2.1 | 2.2 | 3.1 | 4.1 | 5.1 | 5.2 | 6.1 | 6.2 | 6.3 |
|---|---|---|---|---|---|---|---|---|---|---|---|
| | EDF=0.00, p=0.678 | | | | | EDF=0.41, p=0.191 | **EDF=0.76, p=0.037** | **EDF=0.80, p=0.023** | | | |
| Daily mean temperature | lag 10: EDF=0.00, p=0.557<br>lag 6: EDF=0.00, p=0.927<br>lag 0: EDF=0.00, p=0.899 | **lag 6: EDF=0.85, p=0.009** | lag 8: EDF=0.00, p=0.824 | **lag 2: EDF=0.90, p=0.002** | lag 5: EDF=0.18, p=0.323<br>lag 1: EDF=0.00, p=0.427 | - | lag 2: EDF=0.00, p=0.414 | lag 2: EDF=0.00, p=0.889 | lag 10: EDF=0.00, p=0.642<br>lag 10: EDF=0.62, p=0.171<br>**lag 3: EDF=0.87, p=0.008** | lag 2: EDF=0.00, p=0.477<br>lag 6: EDF=0.00, p=0.617<br>lag 5: EDF=0.35, p=0.208 | **lag 10: EDF=2.39, p=0.041** |
| Frost day | lag 7: 0.91 (0.82–1.01), p=0.085<br>**lag 8: 0.94 (0.88–1.00), p=0.037**<br>lag 9: 1.01 (0.94–1.08), p=0.832<br>lag 6: 1.02 (0.97–1.06), p=0.501 | lag 9: 0.96 (0.88–1.04), p=0.282<br>lag 2: 0.96 (0.92–1.01), p=0.123<br>lag 1: 1.00 (0.94–1.06), p=0.989<br>lag 5: 1.01 (0.97–1.06), p=0.574 | lag 1: 1.06 (0.99–1.13), p=0.123<br>**lag 0: 1.04 (1.00–1.09), p=0.043**<br>**lag 6: 1.09 (1.03–1.16), p=0.004**<br>lag 10: 0.98 (0.95–1.01), p=0.251 | lag 0: 0.92 (0.84–1.00), p=0.058<br>**lag 0: 1.05 (1.00–1.10), p=0.031**<br>lag 1: 1.06 (0.99–1.13), p=0.083<br>lag 7: 1.00 (0.96–1.04), p=0.892 | **lag 6: 1.13 (1.00–1.28), p=0.043**<br>lag 5: 1.05 (0.99–1.11), p=0.128<br>lag 7: 0.97 (0.90–1.06), p=0.542<br>lag 8: 1.04 (0.98–1.10), p=0.173 | lag 3: 0.97 (0.84–1.12), p=0.642<br>lag 0: 0.95 (0.88–1.03), p=0.249<br>lag 0: 1.11 (1.00–1.23), p=0.056<br>lag 9: 1.02 (0.95–1.09), p=0.661 | **lag 6: 1.12 (1.01–1.25), p=0.034**<br>**lag 2: 1.07 (1.02–1.13), p=0.009**<br>**lag 9: 1.14 (1.05–1.23), p=<0.001**<br>**lag 2: 0.96 (0.92–1.00), p=0.041** | lag 6: 1.11 (0.94–1.30), p=0.215<br>lag 3: 1.04 (0.98–1.10), p=0.169<br>**lag 1: 1.12 (1.04–1.21), p=0.003**<br>lag 6: 1.05 (1.00–1.11), p=0.069 | **lag 6: 0.74 (0.56–0.97), p=0.033**<br>**lag 0: 0.85 (0.76–0.95), p=0.006**<br>lag 1: 1.08 (0.92–1.26), p=0.354<br>**lag 2: 1.17 (1.06–1.29), p=0.002** | **lag 2: 1.18 (1.11–1.27), p=<0.001**<br>lag 7: 1.00 (0.97–1.04), p=0.811<br>**lag 10: 0.95 (0.90–1.00), p=0.050**<br>lag 5: 1.00 (0.96–1.03), p=0.794 | **lag 7: 1.25 (1.02–1.53), p=0.030**<br>lag 1: 1.09 (0.98–1.21), p=0.100<br>**lag 6: 0.86 (0.74–1.00), p=0.047**<br>lag 9: 1.03 (0.94–1.13), p=0.483 |
| Wind chill index | **lag 6: 0.99 (0.99–1.00), p=<0.001**<br>lag 3: EDF=0.00, p=0.318 | lag 1: EDF=0.01, p=0.354<br>lag 6: EDF=0.00, p=0.506<br>lag 8: EDF=0.00, p=0.707 | lag 10: EDF=0.39, p=0.172<br>lag 0: EDF=0.00, p=0.436<br>**lag 5: EDF=0.83, p=0.013** | lag 8: EDF=0.00, p=0.420<br>lag 9: EDF=0.00, p=0.660 | lag 5: EDF=0.00, p=0.585 | lag 0: EDF=0.00, p=0.613<br>**lag 2: EDF=0.88, p=0.003** | - | lag 10: EDF=0.00, p=0.709 | lag 0: EDF=0.00, p=0.670 | lag 7: EDF=0.00, p=0.995 | lag 8: EDF=0.21, p=0.257 |

| Atmospheric factor | 1.1 | 1.2 | 2.1 | 2.2 | 3.1 | 4.1 | 5.1 | 5.2 | 6.1 | 6.2 | 6.3 |
|---|---|---|---|---|---|---|---|---|---|---|---|
| Heat wave day | lag 4: 1.11 (0.94–1.31), p=0.210<br>**lag 2: 1.10 (1.00–1.21), p=0.044**<br>lag 0: 0.99 (0.88–1.12), p=0.887<br>lag 1: 1.07 (0.99–1.16), p=0.102 | lag 0: 1.11 (0.98–1.25), p=0.089<br>**lag 1: 1.09 (1.03–1.16), p=0.003**<br>**lag 7: 1.10 (1.00–1.20), p=0.042**<br>**lag 0: 1.09 (1.02–1.16), p=0.008** | **lag 5: 1.15 (1.04–1.28), p=0.008**<br>lag 2: 1.02 (0.95–1.10), p=0.554<br>lag 4: 1.04 (0.95–1.12), p=0.398<br>**lag 2: 1.06 (1.01–1.12), p=0.014** | **lag 2: 1.13 (1.00–1.27), p=0.045**<br>lag 1: 1.06 (1.00–1.13), p=0.060<br>lag 0: 1.05 (0.95–1.15), p=0.332<br>**lag 3: 1.07 (1.02–1.12), p=0.008** | lag 5: 0.84 (0.67–1.06), p=0.149<br>lag 6: 0.94 (0.83–1.06), p=0.280<br>lag 2: 0.99 (0.81–1.20), p=0.890<br>lag 9: 0.93 (0.86–1.02), p=0.130 | lag 5: 1.07 (0.86–1.33), p=0.568<br>**lag 0: 1.11 (1.01–1.22), p=0.034**<br>lag 1: 1.10 (0.96–1.25), p=0.161<br>**lag 7: 1.13 (1.04–1.23), p=0.005** | lag 5: 1.02 (0.84–1.23), p=0.868<br>lag 0: 1.07 (0.97–1.18), p=0.157<br>lag 9: 1.09 (0.96–1.23), p=0.193<br>**lag 0: 1.09 (1.02–1.17), p=0.007** | lag 6: 0.88 (0.67–1.16), p=0.369<br>lag 4: 1.08 (0.99–1.18), p=0.095<br>lag 9: 1.16 (0.99–1.34), p=0.061<br>**lag 10: 1.11 (1.03–1.19), p=0.006** | lag 0: 1.20 (0.80–1.79), p=0.380<br>**lag 10: 1.17 (1.02–1.34), p=0.028**<br>lag 1: 0.85 (0.61–1.20), p=0.354<br>lag 7: 0.95 (0.80–1.12), p=0.520 | **lag 8: 1.13 (1.03–1.24), p=0.011**<br>lag 7: 1.04 (0.99–1.09), p=0.135<br>lag 10: 1.07 (0.99–1.14), p=0.072<br>**lag 4: 1.07 (1.03–1.12), p=<0.001** | lag 1: 0.83 (0.63–1.10), p=0.189<br>lag 10: 0.87 (0.75–1.01), p=0.068<br>lag 9: 0.91 (0.73–1.13), p=0.397<br>**lag 0: 1.18 (1.05–1.32), p=0.004** |
| Cold stress wave intensity | lag 1: EDF=0.00, p=0.633<br>lag 7: EDF=0.00, p=0.397<br>lag 0: EDF=0.00, p=0.579<br>lag 6: EDF=0.14, p=0.264 | lag 8: EDF=0.00, p=0.717<br>lag 2: EDF=0.00, p=0.646<br>lag 7: EDF=0.00, p=0.817<br>lag 4: EDF=0.72, p=0.053 | lag 1: EDF=0.46, p=0.141<br>lag 4: EDF=0.00, p=0.363<br>lag 0: EDF=0.00, p=0.426<br>lag 6: EDF=0.48, p=0.152 | lag 1: EDF=0.00, p=0.822<br>**lag 0: EDF=0.90, p=<0.001**<br>lag 2: EDF=0.62, p=0.096<br>lag 4: EDF=0.00, p=0.764 | lag 6: EDF=0.63, p=0.163<br>lag 1: EDF=0.62, p=0.098<br>lag 8: EDF=0.00, p=0.961<br>lag 2: EDF=0.00, p=0.584 | lag 8: EDF=0.00, p=0.882<br>lag 1: EDF=0.00, p=0.484<br>lag 2: EDF=0.00, p=0.853<br>lag 10: EDF=0.00, p=0.711 | lag 10: EDF=0.00, p=0.587<br>lag 10: EDF=0.00, p=0.845<br>lag 0: EDF=0.00, p=0.976<br>lag 7: EDF=0.00, p=0.735 | **lag 3: EDF=0.92, p=<0.001**<br>**lag 2: EDF=0.72, p=0.041**<br>lag 10: EDF=0.00, p=0.369<br>lag 9: EDF=0.00, p=0.990 | lag 2: EDF=1.97, p=0.267<br>lag 3: EDF=0.67, p=0.093<br>**lag 8: EDF=0.88, p=0.017**<br>lag 10: EDF=0.00, p=0.972 | lag 9: EDF=0.00, p=0.581<br>lag 4: EDF=0.00, p=0.802<br>lag 10: EDF=0.11, p=0.272<br>lag 7: EDF=0.24, p=0.244 | **lag 0: EDF=0.85, p=0.046**<br>lag 3: EDF=0.00, p=0.742<br>lag 6: EDF=0.01, p=0.375<br>lag 10: EDF=0.46, p=0.161 |

**Table S20.** Overview of time lags, effect sizes (effective degree of freedom (EDF) or relative risk (95% confidence interval)) and p-values per human-bioclimatic region, atmospheric factor and subgroup (pink = female <60 years, orange = females ≥60 years, green = males <60 years, blue = males ≥60 years) for transient ischemic attack, significant p-values (≤ 0.05) are bold

| Atmospheric factor | 1.1 | 1.2 | 2.1 | 2.2 | 3.1 | 4.1 | 5.1 | 5.2 | 6.1 | 6.2 | 6.3 |
|---|---|---|---|---|---|---|---|---|---|---|---|
| Sea-level pressure difference (SLPD) | lag 7: 1.26 (1.00–1.59), | **lag 8: 1.24 (1.02–1.50),** | lag 1: 1.20 (0.97–1.48), | **lag 9: 0.68 (0.55–0.84),** | **lag 5: 0.67 (0.45–0.99),** | lag 4: 1.29 (0.89–1.87), | **lag 2: 1.45 (1.10–1.92),** | lag 9: 0.70 (0.46–1.07), | **lag 2: 0.51 (0.27–0.94),** | **lag 3: 1.26 (1.01–1.58),** | **lag 9: 0.61 (0.42–0.88),** |

| **Atmospheric factor** | **1.1** | **1.2** | **2.1** | **2.2** | **3.1** | **4.1** | **5.1** | **5.2** | **6.1** | **6.2** | **6.3** |
|---|---|---|---|---|---|---|---|---|---|---|---|
| | p=0.050<br>**lag 10: 0.88 (0.79–0.97), p=0.013**<br>**lag 9: 0.78 (0.63–0.97), p=0.025**<br>**lag 3: 0.87 (0.77–0.99), p=0.031** | **p=0.033**<br>lag 7: 1.08 (0.98–1.20), p=0.106<br>lag 1: 0.90 (0.75–1.07), p=0.217<br>lag 8: 0.92 (0.82–1.03), p=0.170 | p=0.087<br>lag 10: 1.08 (1.00–1.18), p=0.065<br>lag 2: 1.18 (0.98–1.41), p=0.078<br>**lag 6: 1.11 (1.01–1.23), p=0.032** | **p=<0.001**<br>**lag 4: 1.11 (1.02–1.22), p=0.014**<br>**lag 4: 1.23 (1.02–1.48), p=0.034**<br>lag 7: 1.09 (0.98–1.20), p=0.111 | **p=0.046**<br>**lag 8: 0.85 (0.73–1.00), p=0.049**<br>lag 2: 1.26 (0.91–1.75), p=0.162<br>**lag 8: 1.36 (1.14–1.62), p=<0.001** | p=0.172<br>**lag 7: 1.19 (1.03–1.37), p=0.016**<br>lag 6: 0.73 (0.54–1.00), p=0.051<br>lag 0: 1.13 (0.96–1.33), p=0.130 | **p=0.009**<br>**lag 3: 0.83 (0.74–0.93), p=0.001**<br>lag 3: 1.12 (0.88–1.41), p=0.369<br>lag 5: 1.11 (0.97–1.27), p=0.137 | p=0.099<br>**lag 2: 1.18 (1.00–1.39), p=0.044**<br>**lag 3: 0.61 (0.42–0.88), p=0.008**<br>**lag 0: 0.72 (0.59–0.88), p=<0.001** | **p=0.032**<br>lag 3: 1.18 (0.92–1.51), p=0.184<br>lag 6: 1.42 (0.78–2.58), p=0.250<br>**lag 2: 1.42 (1.04–1.93), p=0.029** | **p=0.044**<br>**lag 4: 0.92 (0.85–1.00), p=0.044**<br>lag 5: 0.90 (0.75–1.08), p=0.263<br>**lag 7: 0.90 (0.82–0.99), p=0.029** | **p=0.008**<br>lag 5: 0.86 (0.73–1.01), p=0.061<br>lag 0: 1.24 (0.89–1.71), p=0.201<br>**lag 9: 0.73 (0.60–0.88), p=0.001** |
| Tropical night | lag 9: 2.29 (0.70–7.46), p=0.169<br>lag 2: 1.73 (0.92–3.24), p=0.086<br>lag 3: 1.51 (0.47–4.89), p=0.488<br>lag 6: 1.74 (0.82–3.69), p=0.151 | lag 7: 1.18 (0.55–2.54), p=0.664<br>**lag 10: 0.44 (0.22–0.90), p=0.024**<br>lag 9: 1.66 (0.87–3.18), p=0.125<br>lag 7: 1.33 (0.80–2.20), p=0.266 | lag 8: 2.59 (0.84–8.04), p=0.099<br>lag 9: 1.47 (0.76–2.84), p=0.254<br>lag 1: 2.16 (0.69–6.77), p=0.187<br>lag 4: 1.62 (0.76–3.42), p=0.211 | lag 7: 1.30 (0.67–2.50), p=0.436<br>lag 9: 0.80 (0.56–1.15), p=0.227<br>**lag 4: 0.18 (0.04–0.73), p=0.016**<br>lag 0: 0.77 (0.50–1.17), p=0.220 | - | **lag 10: 15.19 (3.26–70.85), p=<0.001**<br>lag 1: 2.31 (0.31–17.23), p=0.414<br>**lag 6: 11.23 (2.55–49.44), p=0.001**<br>lag 10: 1.66 (0.23–12.23), p=0.618 | lag 2: 3.66 (0.51–26.12), p=0.196<br>lag 2: 2.06 (0.66–6.46), p=0.216<br>lag 8: 4.84 (0.70–33.73), p=0.111<br>lag 2: 2.26 (0.72–7.06), p=0.162 | lag 8: 4.36 (0.59–31.95), p=0.148<br>lag 4: 1.64 (0.51–5.24), p=0.405<br>**lag 1: 7.42 (1.81–30.42), p=0.005**<br>lag 6: 2.91 (0.72–11.83), p=0.136 | **lag 10: 10.69 (1.35–84.66), p=0.025**<br>lag 7: 1.78 (0.44–7.24), p=0.421<br>lag 8: 3.88 (0.47–31.88), p=0.207<br>lag 3: 3.44 (0.79–14.95), p=0.099 | lag 4: 1.86 (0.90–3.85), p=0.094<br>lag 2: 0.67 (0.37–1.20), p=0.174<br>lag 4: 1.40 (0.69–2.85), p=0.356<br>**lag 1: 0.42 (0.20–0.87), p=0.020** | lag 7: 3.46 (0.84–14.23), p=0.085<br>**lag 6: 3.17 (1.74–5.76), p=<0.001**<br>**lag 9: 3.05 (1.08–8.56), p=0.035**<br>lag 3: 1.59 (0.64–3.93), p=0.318 |
| Summer day | lag 5: 0.82 (0.61–1.10), p=0.190<br>**lag 1: 0.85 (0.75–0.97), p=0.015**<br>lag 4: 1.14 | lag 4: 1.15 (0.95–1.40), p=0.155<br>lag 3: 0.90 (0.81–1.00), p=0.054<br>**lag 7: 0.69** | lag 5: 1.07 (0.93–1.24), p=0.356<br>lag 3: 0.94 (0.88–1.01), p=0.093<br>**lag 8: 0.84** | lag 7: 1.06 (0.88–1.28), p=0.539<br>**lag 0: 0.90 (0.84–0.96), p=0.003**<br>lag 5: 1.13 | lag 10: 1.33 (0.94–1.87), p=0.103<br>**lag 10: 0.85 (0.73–0.99), p=0.035**<br>lag 5: 1.21 | lag 5: 1.29 (0.69–2.42), p=0.423<br>**lag 0: 0.61 (0.46–0.81), p=<0.001**<br>lag 5: 1.38 | **lag 10: 1.63 (1.21–2.20), p=0.001**<br>lag 1: 0.87 (0.75–1.00), p=0.057<br>lag 5: 0.85 | lag 3: 1.43 (0.95–2.16), p=0.084<br>lag 6: 1.14 (0.96–1.35), p=0.133<br>lag 0: 1.09 | lag 0: 0.25 (0.06–1.10), p=0.066<br>lag 9: 1.15 (0.86–1.52), p=0.342<br>**lag 0: 2.46** | lag 6: 1.18 (0.96–1.45), p=0.117<br>**lag 4: 0.90 (0.84–0.98), p=0.009**<br>**lag 9: 1.27** | **lag 8: 1.54 (1.05–2.25), p=0.025**<br>lag 1: 0.85 (0.70–1.04), p=0.112<br>lag 0: 1.29 |

| Atmospheric factor | 1.1 | 1.2 | 2.1 | 2.2 | 3.1 | 4.1 | 5.1 | 5.2 | 6.1 | 6.2 | 6.3 |
|---|---|---|---|---|---|---|---|---|---|---|---|
| | (0.91–1.42), p=0.257<br>**lag 2: 0.74 (0.62–0.89), p=0.001** | **(0.56–0.86), p=<0.001**<br>**lag 8: 0.87 (0.77–0.98), p=0.024** | **(0.71–1.00), p=0.045**<br>lag 3: 0.93 (0.87–1.01), p=0.076 | (0.98–1.32), p=0.098<br>lag 7: 0.93 (0.86–1.02), p=0.122 | (0.90–1.63), p=0.205<br>**lag 3: 0.81 (0.67–0.99), p=0.037** | (0.80–2.36), p=0.244<br>**lag 8: 0.62 (0.45–0.85), p=0.003** | (0.63–1.14), p=0.284<br>lag 8: 0.95 (0.81–1.12), p=0.572 | (0.79–1.52), p=0.589<br>lag 10: 0.87 (0.71–1.08), p=0.215 | **(1.19–5.11), p=0.016**<br>**lag 2: 0.63 (0.41–0.99), p=0.045** | **(1.10–1.47), p=0.001**<br>lag 7: 0.94 (0.86–1.02), p=0.148 | (0.97–1.72), p=0.079<br>lag 6: 1.13 (0.92–1.39), p=0.231 |
| Wet-bulb globe temperature | lag 3: EDF=0.79, p=0.091<br>lag 5: EDF=0.31, p=0.201<br>lag 1: EDF=0.00, p=0.992<br>lag 2: EDF=0.00, p=0.569 | lag 7: EDF=0.00, p=0.899<br>**lag 7: EDF=0.81, p=0.013**<br>lag 4: EDF=0.00, p=0.892<br>lag 0: EDF=0.00, p=0.400 | lag 6: EDF=0.00, p=0.519<br>lag 5: EDF=0.00, p=0.693<br>lag 0: EDF=0.00, p=0.473<br>lag 6: EDF=0.00, p=0.552 | **lag 9: EDF=0.86, p=0.016**<br>lag 2: EDF=0.00, p=0.904<br>lag 9: EDF=0.71, p=0.086<br>lag 7: EDF=0.00, p=0.695 | **lag 6: EDF=5.58, p=0.006**<br>**lag 5: EDF=0.81, p=0.031** | **lag 9: 0.97 (0.94–1.00), p=0.034**<br>**lag 0: EDF=0.87, p=0.010**<br>lag 1: EDF=0.00, p=0.529<br>lag 4: EDF=0.00, p=0.561 | lag 6: EDF=0.10, p=0.439<br>lag 5: EDF=0.66, p=0.068<br>lag 4: EDF=0.31, p=0.327<br>lag 2: EDF=0.50, p=0.147 | lag 2: EDF=0.74, p=0.257<br>lag 1: EDF=0.71, p=0.072<br>lag 5: EDF=0.47, p=0.212 | lag 0: EDF=0.73, p=0.472<br>lag 9: EDF=0.00, p=0.695<br>lag 2: EDF=6.28, p=0.233<br>lag 6: EDF=0.89, p=0.065 | lag 0: EDF=0.76, p=0.100<br>lag 1: EDF=0.00, p=0.786<br>lag 8: EDF=0.00, p=0.747 | lag 10: EDF=0.91, p=0.095<br>lag 2: EDF=2.28, p=0.056<br>lag 0: EDF=0.00, p=0.542<br>lag 4: EDF=0.00, p=0.796 |
| Heat wave | lag 9: 1.21 (0.87–1.70), p=0.262<br>lag 4: 0.89 (0.75–1.05), p=0.167<br>lag 8: 1.15 (0.87–1.54), p=0.325<br>lag 0: 0.99 (0.79–1.24), p=0.943 | lag 5: 1.20 (0.93–1.55), p=0.171<br>lag 10: 1.11 (0.97–1.27), p=0.138<br>**lag 3: 1.33 (1.00–1.76), p=0.048**<br>lag 1: 1.07 (0.91–1.26), p=0.415 | lag 9: 1.16 (0.94–1.44), p=0.166<br>**lag 1: 0.86 (0.77–0.96), p=0.007**<br>lag 5: 1.15 (0.90–1.46), p=0.256<br>lag 8: 0.90 (0.80–1.01), p=0.069 | lag 5: 1.01 (0.77–1.33), p=0.950<br>**lag 2: 0.84 (0.75–0.93), p=0.002**<br>lag 0: 1.10 (0.88–1.36), p=0.394<br>lag 5: 0.97 (0.85–1.10), p=0.605 | lag 6: 1.09 (0.66–1.80), p=0.730<br>lag 3: 0.86 (0.70–1.04), p=0.125<br>lag 5: 1.11 (0.77–1.61), p=0.578<br>lag 0: 0.89 (0.69–1.15), p=0.360 | lag 0: 1.02 (0.56–1.84), p=0.955<br>**lag 6: 0.76 (0.60–0.95), p=0.015**<br>lag 2: 1.12 (0.68–1.84), p=0.669<br>**lag 4: 1.41 (1.12–1.78), p=0.004** | **lag 9: 0.46 (0.28–0.74), p=0.002**<br>lag 0: 0.90 (0.75–1.08), p=0.252<br>lag 0: 0.69 (0.47–1.03), p=0.067<br>**lag 5: 0.75 (0.61–0.93), p=0.008** | lag 5: 1.18 (0.71–1.97), p=0.531<br>lag 3: 1.11 (0.89–1.39), p=0.362<br>lag 7: 1.28 (0.85–1.93), p=0.228<br>lag 5: 0.80 (0.59–1.08), p=0.146 | lag 0: 0.44 (0.10–2.01), p=0.291<br>lag 9: 1.21 (0.86–1.70), p=0.267<br>lag 1: 2.61 (0.89–7.63), p=0.080<br>lag 1: 1.19 (0.72–1.98), p=0.498 | lag 5: 1.12 (0.83–1.51), p=0.463<br>**lag 0: 0.80 (0.70–0.91), p=<0.001**<br>lag 0: 1.10 (0.88–1.39), p=0.395<br>lag 9: 0.88 (0.77–1.01), p=0.062 | **lag 0: 0.40 (0.18–0.85), p=0.018**<br>lag 2: 1.26 (0.98–1.62), p=0.075<br>lag 10: 1.21 (0.80–1.83), p=0.362<br>lag 3: 1.03 (0.76–1.39), p=0.859 |
| Cold wave | lag 9: 1.03 (0.89– | lag 0: 0.73 (0.52– | lag 3: 1.05 (0.95– | lag 8: 0.77 (0.58– | lag 7: 0.73 (0.43– | lag 2: 1.59 (0.87– | lag 4: 0.85 (0.62– | lag 1: 0.62 (0.37– | **lag 6: 2.47 (1.27–** | lag 6: 0.87 (0.64– | **lag 9: 1.28 (1.03–** |

| **Atmospheric factor** | **1.1** | **1.2** | **2.1** | **2.2** | **3.1** | **4.1** | **5.1** | **5.2** | **6.1** | **6.2** | **6.3** |
|---|---|---|---|---|---|---|---|---|---|---|---|
| | 1.20), p=0.674 | 1.01), p=0.060<br>**lag 3: 0.85 (0.73–0.99), p=0.032** | 1.16), p=0.311<br>lag 8: 0.94 (0.72–1.23), p=0.666<br>lag 2: 1.03 (0.91–1.17), p=0.630 | 1.03), p=0.082<br>lag 10: 0.95 (0.83–1.09), p=0.484<br>lag 9: 0.90 (0.70–1.16), p=0.404 | 1.26), p=0.258<br>lag 3: 0.85 (0.54–1.33), p=0.468 | 2.92), p=0.133<br>lag 5: 0.98 (0.78–1.24), p=0.895<br>**lag 3: 1.74 (1.07–2.82), p=0.025**<br>lag 8: 1.00 (0.75–1.34), p=0.979 | 1.16), p=0.305 | 1.04), p=0.071 | **4.83), p=0.008**<br>lag 6: 0.76 (0.46–1.26), p=0.294 | 1.19), p=0.391<br>lag 3: 1.02 (0.91–1.15), p=0.737<br>lag 2: 0.81 (0.59–1.10), p=0.174<br>lag 10: 1.09 (0.98–1.21), p=0.120 | **1.58), p=0.028**<br>lag 2: 0.91 (0.66–1.24), p=0.543 |
| Cold stress wave | lag 10: 0.72 (0.51–1.04), p=0.077<br>lag 10: 0.86 (0.63–1.17), p=0.332 | lag 1: 0.78 (0.59–1.03), p=0.075 | lag 5: 0.93 (0.68–1.26), p=0.622<br>lag 8: 1.02 (0.91–1.14), p=0.690 | lag 9: 0.95 (0.83–1.09), p=0.440<br>lag 0: 0.90 (0.80–1.01), p=0.074 | **lag 1: 0.81 (0.68–0.98), p=0.032**<br>**lag 3: 1.39 (1.14–1.69), p=0.001** | lag 5: 0.55 (0.25–1.20), p=0.132<br>lag 4: 1.00 (0.72–1.37), p=0.981 | **lag 7: 1.19 (1.03–1.37), p=0.016** | lag 0: 0.98 (0.78–1.23), p=0.848<br>lag 4: 1.15 (0.93–1.44), p=0.204 | lag 2: 0.82 (0.53–1.25), p=0.355 | lag 0: 0.99 (0.88–1.12), p=0.914<br>lag 10: 0.95 (0.72–1.26), p=0.734 | lag 0: 1.24 (0.96–1.60), p=0.102<br>lag 1: 0.75 (0.44–1.26), p=0.276 |
| Cold stress wave day | lag 0: 1.11 (0.98–1.24), p=0.089<br>lag 0: 0.86 (0.65–1.14), p=0.288 | lag 3: 0.89 (0.78–1.02), p=0.099 | **lag 3: 0.80 (0.65–0.99), p=0.043**<br>lag 7: 1.03 (0.95–1.12), p=0.461<br>lag 3: 0.83 (0.66–1.04), p=0.112<br>lag 5: 1.09 (0.97–1.22), p=0.137 | lag 4: 1.09 (1.00–1.19), p=0.050<br>lag 3: 0.91 (0.73–1.12), p=0.365 | lag 1: 0.86 (0.54–1.37), p=0.531<br>lag 9: 0.85 (0.58–1.23), p=0.385 | lag 10: 0.82 (0.45–1.50), p=0.524<br>lag 0: 1.00 (0.80–1.24), p=0.973<br>lag 8: 0.67 (0.40–1.10), p=0.112<br>lag 1: 0.93 (0.72–1.20), p=0.561 | lag 7: 0.76 (0.54–1.06), p=0.106<br>**lag 1: 0.87 (0.76–1.00), p=0.045**<br>lag 0: 0.89 (0.77–1.04), p=0.150 | **lag 0: 1.66 (1.10–2.51), p=0.016**<br>lag 4: 1.12 (0.92–1.36), p=0.277<br>lag 10: 1.43 (0.99–2.06), p=0.054 | lag 6: 0.94 (0.65–1.37), p=0.763<br>lag 10: 0.32 (0.10–1.06), p=0.063 | lag 6: 0.91 (0.68–1.23), p=0.537 | lag 5: 0.53 (0.27–1.02), p=0.059<br>lag 8: 0.79 (0.59–1.05), p=0.102 |

| Atmospheric factor | 1.1 | 1.2 | 2.1 | 2.2 | 3.1 | 4.1 | 5.1 | 5.2 | 6.1 | 6.2 | 6.3 |
|---|---|---|---|---|---|---|---|---|---|---|---|
| Ozone | lag 2: EDF=0.00, p=0.714<br>lag 7: EDF=0.01, p=0.290<br>lag 2: EDF=0.00, p=0.521<br>**lag 0: EDF=0.78, p=0.030** | lag 3: EDF=0.23, p=0.318<br>**lag 1: EDF=0.70, p=0.047**<br>lag 5: EDF=2.26, p=0.114<br>lag 2: EDF=0.00, p=0.431 | lag 2: EDF=0.00, p=0.676<br>lag 0: EDF=0.00, p=0.845 | lag 0: EDF=0.00, p=0.663<br>lag 0: EDF=0.00, p=0.838<br>lag 0: EDF=0.01, p=0.313 | lag 5: EDF=0.70, p=0.063<br>lag 6: EDF=0.72, p=0.172 | lag 0: EDF=0.83, p=0.169<br>lag 7: EDF=0.40, p=0.218<br>lag 6: EDF=0.74, p=0.217<br>lag 7: EDF=0.00, p=0.427 | lag 3: EDF=0.01, p=0.284 | lag 7: EDF=0.00, p=0.659 | lag 7: 0.98 (0.95–1.01), p=0.253<br>lag 7: EDF=0.81, p=0.090<br>lag 6: EDF=6.02, p=0.153 | lag 1: EDF=0.00, p=0.642<br>lag 0: EDF=0.00, p=0.696 | **lag 6: 0.98 (0.97–0.99), p=0.004**<br>lag 7: EDF=0.00, p=0.409<br>lag 7: EDF=0.00, p=0.693<br>lag 3: EDF=0.00, p=0.792 |

**Table S21.** Overview of time lags, effect sizes (effective degree of freedom (EDF) or relative risk (95% confidence interval)) and p-values per human-bioclimatic region, atmospheric factor and subgroup (pink = female <60 years, orange = females ≥60 years, green = males <60 years, blue = males ≥60 years) for transient ischemic attack (stroke unit), significant p-values (≤ 0.05) are bold

| Atmospheric factor | 1.1 | 1.2 | 2.1 | 2.2 | 3.1 | 4.1 | 5.1 | 5.2 | 6.1 | 6.2 | 6.3 |
|---|---|---|---|---|---|---|---|---|---|---|---|
| Sea-level pressure (SLP) | **lag 2: EDF=0.92, p=0.002**<br>lag 3: EDF=0.66, p=0.073<br>lag 7: EDF=0.01, p=0.363<br>lag 5: EDF=0.00, p=0.384 | lag 7: EDF=0.04, p=0.290<br>lag 2: EDF=0.00, p=0.310<br>**lag 1: EDF=2.55, p=0.009**<br>lag 0: EDF=0.69, p=0.055 | lag 8: EDF=0.00, p=0.599<br>lag 1: EDF=0.00, p=0.464<br>lag 10: EDF=0.67, p=0.073<br>lag 10: EDF=0.65, p=0.078 | lag 10: EDF=0.57, p=0.126<br>lag 3: EDF=0.00, p=0.594<br>lag 10: EDF=0.72, p=0.052<br>**lag 2: EDF=0.85, p=0.008** | lag 3: EDF=0.76, p=0.108<br>**lag 7: EDF=0.79, p=0.023**<br>lag 8: EDF=0.00, p=0.418<br>lag 7: EDF=0.71, p=0.053 | lag 1: EDF=0.84, p=0.052<br>lag 7: EDF=0.63, p=0.078<br>lag 9: EDF=0.28, p=0.307<br>lag 6: EDF=0.59, p=0.102 | lag 10: EDF=0.06, p=0.352<br>lag 5: EDF=0.00, p=0.447<br>lag 8: EDF=0.00, p=0.673<br>lag 4: EDF=0.70, p=0.052 | lag 0: EDF=0.82, p=0.073<br>lag 4: EDF=0.51, p=0.127<br>lag 4: EDF=0.65, p=0.158<br>**lag 3: EDF=0.83, p=0.013** | **lag 9: 0.99 (0.97–1.00), p=0.038**<br>lag 9: EDF=0.73, p=0.059<br>lag 2: EDF=3.37, p=0.166<br>**lag 8: EDF=0.85, p=0.023** | **lag 8: EDF=2.63, p=0.017**<br>**lag 3: EDF=0.91, p=0.001**<br>lag 0: EDF=0.39, p=0.193<br>**lag 4: EDF=0.88, p=0.004** | lag 9: EDF=3.77, p=0.071<br>lag 1: EDF=0.24, p=0.236<br>lag 2: EDF=0.29, p=0.369<br>**lag 3: EDF=0.78, p=0.036** |
| Wind day | **lag 0: 0.82 (0.68–0.99), p=0.041**<br>lag 7: 1.07 | lag 1: 1.08 (0.93–1.25), p=0.324<br>**lag 8: 1.08** | lag 4: 0.87 (0.72–1.04), p=0.120<br>lag 2: 1.02 | **lag 6: 1.32 (1.13–1.53), p=<0.001**<br>lag 7: 1.07 | lag 5: 1.15 (0.87–1.53), p=0.326<br>lag 5: 1.02 | **lag 5: 1.34 (1.03–1.73), p=0.027**<br>**lag 8: 1.12** | lag 8: 1.22 (0.99–1.51), p=0.064<br>lag 9: 1.05 | lag 8: 1.24 (0.93–1.65), p=0.151<br>lag 2: 1.10 | lag 9: 1.18 (0.71–1.97), p=0.527<br>lag 10: 1.13 | **lag 1: 0.80 (0.65–0.99), p=0.038**<br>lag 1: 1.03 | **lag 9: 1.47 (1.07–2.02), p=0.018**<br>**lag 1: 1.18** |

| Atmospheric factor | 1.1 | 1.2 | 2.1 | 2.2 | 3.1 | 4.1 | 5.1 | 5.2 | 6.1 | 6.2 | 6.3 |
|---|---|---|---|---|---|---|---|---|---|---|---|
| | (1.00–1.15), p=0.053<br>**lag 5: 0.85 (0.73–1.00), p=0.048**<br>lag 7: 1.08 (0.99–1.17), p=0.081 | **(1.00–1.16), p=0.043**<br>lag 6: 0.87 (0.76–1.00), p=0.056<br>lag 3: 1.04 (0.95–1.13), p=0.408 | (0.96–1.09), p=0.477<br>**lag 9: 1.16 (1.01–1.34), p=0.039**<br>lag 3: 1.06 (0.98–1.14), p=0.138 | (1.00–1.15), p=0.053<br>lag 2: 1.13 (0.98–1.30), p=0.094<br>lag 2: 1.04 (0.97–1.13), p=0.269 | (0.91–1.16), p=0.703<br>lag 1: 1.21 (0.95–1.56), p=0.126<br>**lag 0: 0.80 (0.69–0.93), p=0.004** | **(1.00–1.25), p=0.041**<br>lag 8: 1.24 (0.99–1.55), p=0.066<br>**lag 10: 1.22 (1.08–1.38), p=0.002** | (0.96–1.14), p=0.282<br>lag 4: 1.19 (1.00–1.42), p=0.051<br>**lag 10: 1.11 (1.00–1.22), p=0.042** | (0.97–1.23), p=0.128<br>lag 4: 0.89 (0.65–1.21), p=0.453<br>lag 10: 1.09 (0.95–1.25), p=0.220 | (0.95–1.34), p=0.176<br>**lag 4: 1.55 (1.03–2.31), p=0.033**<br>lag 8: 0.92 (0.72–1.18), p=0.518 | (0.96–1.10), p=0.403<br>**lag 9: 1.22 (1.06–1.42), p=0.007**<br>lag 4: 1.07 (0.99–1.16), p=0.088 | **(1.02–1.36), p=0.026**<br>lag 4: 0.69 (0.47–1.01), p=0.055<br>**lag 9: 1.23 (1.04–1.45), p=0.013** |
| Summer day | lag 7: 1.04 (0.88–1.24), p=0.627<br>**lag 1: 0.92 (0.86–0.98), p=0.012**<br>lag 8: 1.02 (0.90–1.16), p=0.796<br>**lag 3: 0.90 (0.83–0.98), p=0.013** | **lag 4: 1.14 (1.02–1.28), p=0.019**<br>lag 3: 0.95 (0.90–1.01), p=0.108<br>lag 8: 1.09 (0.99–1.21), p=0.073<br>lag 9: 0.97 (0.90–1.03), p=0.320 | lag 6: 1.11 (0.98–1.25), p=0.105<br>**lag 1: 0.95 (0.90–0.99), p=0.024**<br>lag 6: 1.03 (0.93–1.14), p=0.553<br>**lag 0: 0.93 (0.88–0.98), p=0.006** | **lag 8: 1.16 (1.03–1.30), p=0.013**<br>**lag 0: 0.94 (0.90–0.99), p=0.020**<br>lag 1: 1.03 (0.93–1.14), p=0.523<br>**lag 10: 0.93 (0.88–0.98), p=0.012** | lag 8: 1.11 (0.85–1.44), p=0.442<br>**lag 5: 0.86 (0.76–0.96), p=0.008**<br>**lag 2: 1.35 (1.06–1.71), p=0.014**<br>**lag 3: 0.88 (0.78–1.00), p=0.042** | lag 5: 1.29 (0.91–1.84), p=0.155<br>lag 2: 0.88 (0.74–1.05), p=0.149<br>**lag 3: 1.29 (1.01–1.66), p=0.044**<br>lag 6: 1.06 (0.91–1.23), p=0.473 | lag 6: 1.06 (0.85–1.32), p=0.600<br>**lag 1: 0.90 (0.83–0.99), p=0.027**<br>**lag 10: 0.79 (0.66–0.95), p=0.014**<br>**lag 1: 0.88 (0.80–0.96), p=0.006** | lag 10: 1.21 (0.95–1.56), p=0.127<br>**lag 1: 0.86 (0.78–0.96), p=0.006**<br>lag 3: 1.25 (1.00–1.56), p=0.054<br>lag 8: 1.04 (0.91–1.18), p=0.601 | lag 7: 1.41 (0.93–2.14), p=0.106<br>lag 10: 0.87 (0.74–1.01), p=0.071<br>lag 0: 1.09 (0.72–1.64), p=0.689<br>**lag 1: 0.78 (0.63–0.97), p=0.023** | lag 10: 1.12 (0.98–1.29), p=0.092<br>**lag 2: 0.94 (0.89–0.99), p=0.023**<br>**lag 9: 1.20 (1.07–1.35), p=0.002**<br>**lag 9: 0.94 (0.88–1.00), p=0.049** | **lag 2: 0.56 (0.41–0.78), p=<0.001**<br>**lag 0: 0.84 (0.73–0.96), p=0.009**<br>lag 0: 1.27 (1.00–1.62), p=0.053<br>**lag 5: 0.83 (0.71–0.96), p=0.014** |
| Wet-bulb globe temperature | lag 3: EDF=0.55, p=0.109 | lag 3: EDF=0.00, p=0.630<br>**lag 5: EDF=0.82, p=0.011** | lag 5: EDF=0.00, p=0.841 | lag 9: EDF=0.00, p=0.322 | lag 6: EDF=0.75, p=0.119<br>lag 10: EDF=0.00, p=0.539 | **lag 3: EDF=2.53, p=0.031**<br>lag 3: EDF=0.34, p=0.186<br>lag 6: EDF=0.00, p=0.717 | lag 5: EDF=0.40, p=0.242<br>lag 2: EDF=0.37, p=0.175 | lag 7: EDF=0.65, p=0.190 | lag 3: EDF=7.11, p=0.061<br>**lag 6: 1.01 (1.01–1.02), p=<0.001**<br>lag 0: EDF=0.00, p=0.771<br>lag 3: | lag 4: EDF=0.47, p=0.169<br>lag 5: EDF=0.19, p=0.252<br>lag 9: EDF=0.00, p=0.370 | **lag 7: EDF=0.90, p=0.044** |

| Atmospheric factor | 1.1 | 1.2 | 2.1 | 2.2 | 3.1 | 4.1 | 5.1 | 5.2 | 6.1 | 6.2 | 6.3 |
|---|---|---|---|---|---|---|---|---|---|---|---|
| | | | | | | | | | EDF=0.58, p=0.163 | | |
| Frost day | lag 3: 0.91 (0.81–1.03), p=0.130<br>lag 1: 0.96 (0.91–1.01), p=0.109<br>lag 5: 0.92 (0.83–1.02), p=0.130<br>**lag 0: 0.94 (0.88–1.00), p=0.042** | lag 3: 0.96 (0.86–1.08), p=0.492<br>lag 0: 0.97 (0.92–1.02), p=0.226<br>lag 1: 0.94 (0.86–1.03), p=0.169<br>lag 3: 0.96 (0.90–1.02), p=0.222 | **lag 0: 0.88 (0.80–0.97), p=0.011**<br>lag 8: 0.98 (0.95–1.02), p=0.372<br>**lag 4: 0.91 (0.83–0.99), p=0.024**<br>**lag 3: 0.93 (0.89–0.97), p=0.001** | **lag 4: 0.83 (0.74–0.94), p=0.003**<br>**lag 7: 0.95 (0.90–0.99), p=0.019**<br>lag 9: 0.91 (0.82–1.01), p=0.086<br>lag 1: 0.98 (0.92–1.04), p=0.482 | **lag 6: 0.71 (0.59–0.87), p=<0.001**<br>lag 9: 0.95 (0.89–1.01), p=0.129<br>lag 7: 0.95 (0.83–1.09), p=0.470<br>lag 0: 0.95 (0.88–1.02), p=0.125 | lag 9: 0.89 (0.70–1.12), p=0.307<br>**lag 1: 0.91 (0.84–0.99), p=0.030**<br>lag 0: 0.87 (0.73–1.03), p=0.113<br>**lag 5: 0.91 (0.83–1.00), p=0.043** | **lag 6: 0.85 (0.73–0.98), p=0.031**<br>lag 8: 0.99 (0.93–1.04), p=0.601<br>**lag 5: 0.88 (0.78–0.99), p=0.028**<br>lag 10: 0.97 (0.92–1.03), p=0.374 | **lag 6: 1.27 (1.05–1.54), p=0.015**<br>lag 4: 0.97 (0.89–1.05), p=0.422<br>**lag 8: 0.84 (0.71–0.99), p=0.043**<br>lag 9: 1.03 (0.96–1.10), p=0.471 | lag 8: 0.96 (0.64–1.44), p=0.854<br>**lag 8: 1.18 (1.05–1.34), p=0.007**<br>**lag 2: 0.64 (0.42–0.98), p=0.042**<br>lag 9: 0.87 (0.75–1.02), p=0.087 | **lag 4: 0.86 (0.75–0.99), p=0.036**<br>**lag 2: 0.96 (0.92–1.00), p=0.041**<br>lag 0: 0.95 (0.86–1.05), p=0.318<br>lag 4: 1.00 (0.94–1.05), p=0.873 | lag 2: 0.86 (0.66–1.13), p=0.280<br>lag 0: 0.99 (0.87–1.12), p=0.875<br>**lag 2: 1.28 (1.03–1.58), p=0.024**<br>lag 4: 0.96 (0.84–1.09), p=0.508 |
| Wind chill index | lag 3: EDF=0.00, p=0.728<br>lag 9: EDF=0.00, p=0.650<br>lag 1: EDF=0.53, p=0.129<br>lag 6: EDF=0.00, p=0.976 | lag 1: EDF=0.00, p=0.569<br>lag 1: EDF=0.00, p=0.307<br>lag 10: EDF=0.03, p=0.271 | lag 1: EDF=0.00, p=0.831<br>lag 10: EDF=0.00, p=0.468<br>lag 0: EDF=0.23, p=0.240<br>lag 3: EDF=0.00, p=0.468 | lag 7: EDF=0.07, p=0.279<br>**lag 3: EDF=0.77, p=0.035**<br>**lag 0: EDF=2.03, p=0.029** | lag 9: EDF=0.00, p=0.951<br>lag 1: EDF=0.00, p=0.793 | lag 2: EDF=0.00, p=0.668 | lag 7: EDF=0.00, p=0.660<br>lag 5: EDF=0.00, p=0.640<br>lag 0: EDF=0.03, p=0.287 | lag 4: EDF=0.61, p=0.087<br>lag 6: EDF=0.00, p=0.455<br>lag 6: EDF=0.00, p=0.436 | **lag 3: 1.07 (1.02–1.12), p=0.011**<br>lag 5: EDF=0.00, p=0.982<br>**lag 0: 1.04 (1.01–1.07), p=0.022** | lag 3: EDF=0.00, p=0.605<br>**lag 1: EDF=0.73, p=0.043** | **lag 0: EDF=0.93, p=0.001**<br>lag 10: EDF=2.39, p=0.149<br>**lag 2: EDF=0.91, p=0.004** |
| Heat stress wave | lag 5: 1.14 (0.90–1.43), p=0.274<br>**lag 5: 0.88 (0.81–0.97), p=0.011** | lag 5: 1.11 (0.95–1.30), p=0.203<br>lag 0: 0.93 (0.85–1.01), p=0.090 | lag 3: 1.08 (0.89–1.31), p=0.425<br>lag 10: 1.06 (0.98–1.14), p=0.127 | lag 4: 1.16 (0.97–1.38), p=0.111<br>lag 9: 1.00 (0.92–1.08), p=0.922 | lag 4: 1.17 (0.81–1.69), p=0.390<br>lag 1: 0.90 (0.76–1.06), p=0.199 | lag 0: 1.27 (0.89–1.83), p=0.190<br>lag 1: 0.87 (0.74–1.02), p=0.076 | lag 3: 1.13 (0.85–1.50), p=0.388<br>lag 0: 0.91 (0.81–1.03), p=0.135 | lag 5: 1.04 (0.72–1.51), p=0.824<br>**lag 10: 0.82 (0.70–0.96), p=0.014** | lag 3: 0.70 (0.36–1.36), p=0.288<br>lag 2: 1.02 (0.84–1.25), p=0.833 | lag 6: 1.23 (1.00–1.51), p=0.054<br>**lag 0: 0.86 (0.79–0.95), p=0.002** | **lag 9: 1.54 (1.09–2.18), p=0.015**<br>lag 3: 1.03 (0.86–1.23), p=0.747 |

| Atmospheric factor | 1.1 | 1.2 | 2.1 | 2.2 | 3.1 | 4.1 | 5.1 | 5.2 | 6.1 | 6.2 | 6.3 |
|---|---|---|---|---|---|---|---|---|---|---|---|
| | lag 3: 1.08 (0.91–1.28), p=0.401<br>**lag 0: 0.85 (0.75–0.95), p=0.004** | lag 4: 0.95 (0.82–1.11), p=0.524<br>lag 1: 0.93 (0.84–1.03), p=0.165 | lag 8: 1.11 (0.95–1.30), p=0.189<br>**lag 9: 0.88 (0.81–0.96), p=0.005** | lag 7: 1.14 (0.99–1.33), p=0.076<br>lag 5: 0.93 (0.85–1.02), p=0.137 | lag 1: 0.92 (0.65–1.30), p=0.626<br>lag 9: 1.14 (0.98–1.33), p=0.101 | lag 8: 1.06 (0.92–1.22), p=0.449 | lag 4: 1.23 (0.99–1.52), p=0.062<br>**lag 10: 0.86 (0.76–0.98), p=0.019** | lag 3: 0.92 (0.66–1.30), p=0.641<br>**lag 7: 0.66 (0.53–0.83), p=<0.001** | **lag 5: 0.45 (0.21–0.96), p=0.040**<br>lag 4: 0.78 (0.59–1.05), p=0.102 | lag 9: 1.03 (0.86–1.25), p=0.724<br>lag 9: 0.91 (0.82–1.02), p=0.105 | lag 4: 1.33 (0.92–1.92), p=0.128<br>**lag 0: 0.76 (0.59–0.96), p=0.023** |
| Cold stress wave duration | lag 10: EDF=0.42, p=0.203<br>lag 1: EDF=0.00, p=0.293<br>lag 0: EDF=0.00, p=0.317<br>lag 0: EDF=0.29, p=0.196 | **lag 0: EDF=0.90, p=0.001**<br>lag 0: EDF=0.00, p=0.925<br>lag 0: EDF=0.00, p=0.372<br>lag 3: EDF=0.00, p=0.534 | lag 10: EDF=0.36, p=0.214<br>lag 10: EDF=0.00, p=0.655<br>lag 4: EDF=0.00, p=0.385<br>lag 0: EDF=0.00, p=0.764 | lag 3: EDF=0.65, p=0.096<br>lag 5: EDF=0.00, p=0.355<br>lag 2: EDF=0.00, p=0.688<br>lag 0: EDF=0.00, p=0.948 | lag 7: EDF=0.00, p=0.910<br>lag 6: EDF=0.00, p=0.766<br>lag 10: EDF=0.71, p=0.137<br>lag 4: EDF=0.55, p=0.128 | lag 4: EDF=0.00, p=0.778<br>lag 0: EDF=0.00, p=0.497<br>lag 0: EDF=0.00, p=0.485<br>lag 0: EDF=0.00, p=0.903 | lag 1: EDF=0.23, p=0.291<br>**lag 0: EDF=0.76, p=0.032**<br>lag 0: EDF=0.00, p=0.355<br>lag 2: EDF=0.00, p=0.682 | lag 8: EDF=0.01, p=0.437<br>lag 2: EDF=0.00, p=0.382<br>**lag 9: EDF=0.93, p=0.002**<br>lag 6: EDF=0.00, p=0.461 | lag 2: EDF=0.84, p=0.177<br>lag 1: EDF=0.00, p=0.675<br>**lag 0: EDF=2.64, p=0.007**<br>**lag 7: EDF=0.89, p=0.005** | lag 2: EDF=0.00, p=0.817<br>**lag 8: EDF=0.85, p=0.007**<br>lag 8: EDF=0.64, p=0.097<br>lag 0: EDF=0.39, p=0.178 | lag 5: EDF=0.64, p=0.259<br>lag 3: EDF=0.00, p=0.344<br>lag 1: EDF=0.08, p=0.418<br>lag 7: EDF=0.00, p=0.568 |
| Nitrogen dioxide | lag 4: EDF=0.00, p=0.761<br>lag 5: EDF=0.54, p=0.119<br>lag 7: EDF=0.23, p=0.238<br>lag 2: EDF=0.00, p=0.820 | lag 1: EDF=0.63, p=0.089<br>lag 4: EDF=0.00, p=0.857<br>lag 2: EDF=0.02, p=0.296<br>**lag 3: EDF=0.77, p=0.026** | lag 1: EDF=0.45, p=0.175<br>lag 7: EDF=0.64, p=0.084<br>lag 5: EDF=0.00, p=0.576<br>lag 3: EDF=0.00, p=0.792 | lag 3: EDF=0.39, p=0.195<br>lag 7: EDF=0.28, p=0.217<br>**lag 7: EDF=0.81, p=0.019**<br>lag 1: EDF=0.00, p=0.421 | lag 1: EDF=0.00, p=0.595<br>lag 0: EDF=0.00, p=0.710<br>lag 1: EDF=0.00, p=0.714<br>lag 7: EDF=0.00, p=0.585 | lag 0: EDF=0.00, p=0.840<br>lag 4: EDF=0.65, p=0.069<br>lag 0: EDF=0.00, p=0.845<br>lag 7: EDF=0.00, p=0.966 | lag 7: EDF=0.00, p=0.749<br>lag 4: EDF=0.00, p=0.355<br>lag 3: EDF=0.04, p=0.331<br>lag 3: EDF=0.00, p=0.862 | **lag 1: EDF=0.92, p=0.006**<br>lag 2: EDF=0.00, p=0.785<br>**lag 7: EDF=0.93, p=0.002**<br>**lag 0: EDF=0.91, p=<0.001** | **lag 4: EDF=3.81, p=0.001**<br>lag 2: EDF=0.71, p=0.064<br>lag 1: EDF=0.38, p=0.440<br>lag 4: EDF=0.00, p=0.441 | lag 2: EDF=0.47, p=0.181<br>lag 3: EDF=0.66, p=0.075<br>lag 1: EDF=0.24, p=0.238<br>lag 4: EDF=0.00, p=0.541 | lag 6: EDF=0.85, p=0.069<br>lag 1: EDF=0.00, p=0.563<br>lag 7: EDF=0.01, p=0.460<br>lag 1: EDF=0.00, p=0.871 |
| Particulate matter ($PM_{10}$) | lag 1: EDF=0.00, p=0.794<br>lag 6: EDF=0.06, p=0.279 | lag 6: EDF=0.00, p=0.525<br>lag 2: EDF=0.00, p=0.499 | lag 4: EDF=0.63, p=0.097<br>lag 2: EDF=0.00, p=0.391 | lag 6: EDF=0.00, p=0.488<br>lag 4: EDF=0.00, p=0.724 | **lag 1: EDF=2.84, p=0.037**<br>lag 6: EDF=0.00, p=0.963 | lag 5: EDF=0.69, p=0.155<br>lag 1: EDF=0.00, p=0.825 | lag 7: EDF=0.00, p=0.660<br>lag 6: EDF=0.01, p=0.299 | lag 1: EDF=0.00, p=0.757<br>lag 1: EDF=0.00, p=0.857 | **lag 2: 1.02 (1.01–1.04), p=0.006**<br>lag 6: EDF=0.07, | **lag 5: EDF=0.80, p=0.031**<br>lag 2: EDF=0.00, p=0.846 | lag 6: EDF=0.21, p=0.420<br>lag 0: EDF=0.00, p=0.818 |

| Atmospheric factor | 1.1 | 1.2 | 2.1 | 2.2 | 3.1 | 4.1 | 5.1 | 5.2 | 6.1 | 6.2 | 6.3 |
|---|---|---|---|---|---|---|---|---|---|---|---|
| | lag 7: EDF=0.00, p=0.376<br>lag 1: EDF=0.00, p=0.913 | **lag 2: EDF=1.88, p=0.015**<br>**lag 2: EDF=3.14, p=0.001** | lag 5: EDF=0.72, p=0.051<br>**lag 5: EDF=0.88, p=0.003** | lag 2: EDF=0.00, p=0.866<br>lag 0: EDF=0.00, p=0.844 | lag 6: EDF=0.45, p=0.262<br>lag 1: EDF=0.07, p=0.275 | lag 2: EDF=0.00, p=0.430<br>lag 0: EDF=0.00, p=0.493 | lag 2: EDF=0.50, p=0.173<br>lag 3: EDF=0.65, p=0.069 | lag 2: EDF=0.78, p=0.073<br>lag 3: EDF=0.00, p=0.607 | p=0.297<br>**lag 7: EDF=0.93, p=0.038**<br>**lag 1: EDF=0.85, p=0.018** | lag 1: EDF=0.00, p=0.463<br>lag 3: EDF=0.00, p=0.533 | lag 5: EDF=0.77, p=0.121<br>lag 1: EDF=0.00, p=0.797 |

**Table S22.** Overview of time lags, effect sizes (effective degree of freedom (EDF) or relative risk (95% confidence interval)) and p-values per human-bioclimatic region, atmospheric factor and subgroup (pink = female <60 years, orange = females ≥60 years, green = males <60 years, blue = males ≥60 years) for tachycardia, significant p-values (≤ 0.05) are bold

| Atmospheric factor | 1.1 | 1.2 | 2.1 | 2.2 | 3.1 | 4.1 | 5.1 | 5.2 | 6.1 | 6.2 | 6.3 |
|---|---|---|---|---|---|---|---|---|---|---|---|
| Sea-level pressure (SLP) | **lag 0: EDF=0.93, p=<0.001**<br>lag 9: EDF=0.00, p=0.604<br>**lag 9: EDF=0.77, p=0.027**<br>lag 10: EDF=0.00, p=0.339 | lag 9: EDF=0.48, p=0.155<br>lag 0: EDF=0.54, p=0.121<br>lag 10: EDF=0.57, p=0.104<br>**lag 1: EDF=0.83, p=0.013** | **lag 1: EDF=0.89, p=0.003**<br>**lag 0: EDF=0.78, p=0.029**<br>**lag 10: EDF=0.86, p=0.006**<br>**lag 3: EDF=0.83, p=0.014** | lag 0: EDF=0.40, p=0.182<br>**lag 0: EDF=0.88, p=0.003**<br>lag 2: EDF=0.60, p=0.093<br>**lag 0: EDF=0.86, p=0.008** | lag 1: EDF=0.72, p=0.092<br>lag 5: EDF=0.69, p=0.058<br>**lag 1: EDF=2.01, p=0.018**<br>**lag 2: EDF=0.79, p=0.021** | lag 1: EDF=0.80, p=0.068<br>lag 2: EDF=0.71, p=0.056<br>lag 1: EDF=0.29, p=0.261<br>**lag 10: EDF=0.88, p=0.005** | lag 0: EDF=0.65, p=0.107<br>**lag 2: EDF=0.84, p=0.009**<br>lag 2: EDF=0.00, p=0.464<br>lag 7: EDF=0.01, p=0.297 | **lag 2: EDF=0.89, p=0.019**<br>lag 10: EDF=0.69, p=0.059<br>**lag 3: EDF=0.82, p=0.030**<br>lag 4: EDF=0.66, p=0.073 | **lag 1: EDF=2.22, p=0.005**<br>lag 6: EDF=0.00, p=0.396<br>lag 8: EDF=0.68, p=0.259<br>lag 5: EDF=0.70, p=0.110 | lag 4: EDF=0.71, p=0.069<br>lag 6: EDF=0.00, p=0.322<br>**lag 1: EDF=0.77, p=0.029**<br>lag 0: EDF=0.49, p=0.137 | lag 7: EDF=0.00, p=0.487<br>lag 4: EDF=0.71, p=0.065<br>lag 8: EDF=0.83, p=0.052<br>**lag 2: EDF=1.93, p=0.041** |
| Wind day | **lag 3: 0.82 (0.70–0.96), p=0.016**<br>**lag 9: 1.11 (1.03–1.20), p=0.006**<br>**lag 2: 0.86 (0.77–0.98), p=0.018** | lag 9: 1.07 (0.91–1.25), p=0.403<br>**lag 4: 0.91 (0.84–0.99), p=0.028**<br>lag 5: 1.07 (0.96–1.20), p=0.213 | lag 10: 0.88 (0.76–1.03), p=0.107<br>lag 8: 1.03 (0.96–1.10), p=0.446<br>lag 4: 0.90 (0.81–1.01), p=0.076 | **lag 0: 1.29 (1.13–1.48), p=<0.001**<br>**lag 2: 0.92 (0.85–0.99), p=0.021**<br>**lag 4: 0.85 (0.75–0.97), p=0.016** | **lag 9: 1.29 (1.03–1.60), p=0.026**<br>lag 4: 0.93 (0.83–1.05), p=0.233<br>lag 10: 0.84 (0.69–1.01), p=0.070 | lag 7: 1.14 (0.88–1.48), p=0.305<br>**lag 10: 1.13 (1.00–1.29), p=0.049**<br>**lag 10: 1.21 (1.01–1.46), p=0.036** | lag 8: 1.18 (0.98–1.42), p=0.074<br>**lag 3: 1.11 (1.02–1.20), p=0.014**<br>**lag 2: 0.82 (0.70–0.96), p=0.014** | **lag 0: 1.30 (1.00–1.69), p=0.049**<br>**lag 8: 0.87 (0.75–1.00), p=0.048**<br>**lag 3: 0.77 (0.61–0.98), p=0.036** | **lag 8: 1.68 (1.03–2.76), p=0.039**<br>lag 3: 0.86 (0.68–1.08), p=0.197<br>**lag 6: 1.59 (1.12–2.27), p=0.010** | lag 4: 0.83 (0.68–1.02), p=0.073<br>lag 0: 0.93 (0.87–1.00), p=0.062<br>**lag 5: 0.87 (0.76–0.99), p=0.038** | **lag 7: 1.52 (1.13–2.06), p=0.006**<br>lag 3: 0.88 (0.73–1.05), p=0.159<br>lag 7: 0.86 (0.63–1.18), p=0.347 |

| Atmospheric factor | 1.1 | 1.2 | 2.1 | 2.2 | 3.1 | 4.1 | 5.1 | 5.2 | 6.1 | 6.2 | 6.3 |
|---|---|---|---|---|---|---|---|---|---|---|---|
| | lag 0: 0.94 (0.87–1.03), p=0.184 | **lag 8: 1.16 (1.07–1.26), p=<0.001** | lag 1: 1.07 (0.99–1.14), p=0.080 | lag 9: 0.97 (0.90–1.05), p=0.499 | **lag 7: 1.12 (1.01–1.25), p=0.038** | lag 0: 1.11 (0.98–1.26), p=0.113 | lag 8: 1.07 (0.97–1.17), p=0.167 | lag 7: 1.12 (0.98–1.28), p=0.111 | lag 0: 0.78 (0.60–1.03), p=0.081 | lag 5: 1.04 (0.97–1.13), p=0.271 | **lag 8: 1.29 (1.09–1.52), p=0.003** |
| Tropical night | lag 7: 1.47 (0.73–2.99), p=0.284<br>**lag 2: 1.96 (1.39–2.77), p=<0.001**<br>lag 5: 0.14 (0.02–1.03), p=0.054<br>lag 6: 0.62 (0.29–1.33), p=0.221 | lag 9: 1.31 (0.77–2.21), p=0.322<br>lag 1: 1.31 (0.99–1.72), p=0.056<br>lag 5: 1.19 (0.80–1.78), p=0.394<br>lag 6: 1.01 (0.72–1.43), p=0.939 | **lag 8: 3.04 (1.33–6.94), p=0.008**<br>lag 5: 1.13 (0.66–1.92), p=0.666<br>lag 9: 1.61 (0.79–3.29), p=0.186<br>lag 10: 1.41 (0.79–2.49), p=0.242 | lag 10: 0.67 (0.34–1.31), p=0.238<br>**lag 9: 1.25 (1.01–1.53), p=0.037**<br>lag 5: 1.33 (0.95–1.86), p=0.094<br>lag 8: 0.83 (0.61–1.13), p=0.244 | - | lag 8: 2.08 (0.26–16.75), p=0.493<br>lag 4: 1.91 (0.59–6.19), p=0.280<br>lag 9: 2.22 (0.53–9.18), p=0.273<br>lag 6: 1.93 (0.61–6.14), p=0.265 | lag 7: 3.04 (0.75–12.28), p=0.119<br>lag 9: 1.89 (0.97–3.69), p=0.063<br>**lag 7: 4.55 (1.37–15.04), p=0.013**<br>**lag 2: 2.11 (1.05–4.27), p=0.037** | lag 1: 2.23 (0.52–9.53), p=0.281<br>**lag 2: 2.30 (1.08–4.89), p=0.031**<br>lag 6: 2.42 (0.76–7.70), p=0.135<br>**lag 9: 2.35 (1.12–4.96), p=0.025** | lag 0: 0.00 (0.00–Inf), p=1.000<br>lag 4: 1.59 (0.50–5.03), p=0.434<br>lag 2: 2.34 (0.48–11.37), p=0.290<br>lag 0: 1.84 (0.46–7.41), p=0.390 | **lag 7: 2.71 (1.50–4.88), p=<0.001**<br>lag 4: 0.74 (0.53–1.05), p=0.092<br>lag 10: 1.24 (0.78–1.98), p=0.356<br>lag 0: 0.77 (0.51–1.17), p=0.216 | lag 5: 3.49 (0.82–14.80), p=0.090<br>lag 6: 1.67 (0.86–3.26), p=0.132<br>lag 10: 2.02 (0.77–5.30), p=0.151<br>lag 0: 1.61 (0.75–3.44), p=0.220 |
| Hot day | lag 0: 1.15 (0.89–1.50), p=0.280<br>lag 1: 0.92 (0.78–1.08), p=0.303<br>**lag 7: 0.75 (0.57–0.98), p=0.034**<br>lag 8: 0.95 (0.80–1.12), p=0.510 | lag 5: 0.84 (0.61–1.14), p=0.263<br>lag 1: 0.97 (0.84–1.12), p=0.663<br>lag 5: 1.19 (0.97–1.46), p=0.089<br>lag 9: 0.98 (0.85–1.13), p=0.756 | **lag 7: 0.78 (0.61–1.00), p=0.050**<br>**lag 4: 0.87 (0.78–0.97), p=0.015**<br>lag 8: 0.92 (0.78–1.09), p=0.351<br>lag 1: 0.90 (0.79–1.02), p=0.091 | lag 9: 0.85 (0.66–1.10), p=0.224<br>lag 0: 0.94 (0.85–1.04), p=0.218<br>lag 10: 0.94 (0.76–1.16), p=0.549<br>lag 8: 1.00 (0.88–1.14), p=0.990 | lag 8: 1.17 (0.72–1.89), p=0.522<br>**lag 8: 0.70 (0.52–0.95), p=0.023**<br>lag 10: 0.61 (0.36–1.03), p=0.066<br>lag 8: 1.01 (0.76–1.34), p=0.961 | lag 7: 1.80 (0.63–5.10), p=0.272<br>lag 1: 0.41 (0.15–1.12), p=0.082<br>**lag 4: 1.89 (1.01–3.57), p=0.048**<br>**lag 2: 0.11 (0.02–0.81), p=0.030** | **lag 10: 0.49 (0.29–0.83), p=0.007**<br>lag 3: 1.15 (0.98–1.35), p=0.092<br>lag 4: 0.75 (0.53–1.08), p=0.123<br>**lag 3: 1.27 (1.05–1.52), p=0.013** | **lag 1: 1.59 (1.02–2.48), p=0.040**<br>lag 6: 0.79 (0.59–1.07), p=0.127<br>lag 8: 1.33 (0.95–1.87), p=0.098<br>**lag 1: 0.67 (0.48–0.94), p=0.019** | lag 5: 1.35 (0.31–5.89), p=0.692<br>lag 0: 0.62 (0.35–1.11), p=0.109<br>lag 2: 1.25 (0.56–2.79), p=0.590<br>lag 8: 0.60 (0.28–1.31), p=0.200 | lag 7: 0.75 (0.54–1.04), p=0.080<br>lag 6: 0.89 (0.79–1.00), p=0.058<br>lag 5: 0.83 (0.68–1.01), p=0.067<br>**lag 1: 0.87 (0.76–0.99), p=0.038** | lag 8: 0.39 (0.14–1.06), p=0.064<br>lag 5: 1.14 (0.88–1.46), p=0.324<br>**lag 10: 1.97 (1.28–3.03), p=0.002**<br>**lag 7: 0.71 (0.51–1.00), p=0.047** |
| Wind chill index | **lag 8: EDF=0.95,** | lag 5: EDF=0.00, | **lag 9: EDF=0.74,** | lag 7: EDF=0.00, | **lag 3: EDF=0.83,** | lag 6: EDF=0.00, | lag 7: EDF=0.00, | lag 8: EDF=0.72, | lag 9: 0.97 (0.94– | lag 2: EDF=0.62, | lag 4: EDF=0.00, |

| **Atmospheric factor** | **1.1** | **1.2** | **2.1** | **2.2** | **3.1** | **4.1** | **5.1** | **5.2** | **6.1** | **6.2** | **6.3** |
|---|---|---|---|---|---|---|---|---|---|---|---|
| | **p=<0.001** lag 2: EDF=0.00, p=0.451 lag 9: EDF=0.40, p=0.165 lag 0: EDF=0.49, p=0.122 | p=0.811 lag 1: EDF=0.00, p=0.854 **lag 0: EDF=0.89, p=0.002** **lag 6: 0.99 (0.99–1.00), p=<0.001** | **p=0.043** lag 2: EDF=0.00, p=0.643 lag 3: EDF=0.46, p=0.141 | p=0.524 lag 7: EDF=0.69, p=0.052 lag 8: EDF=0.00, p=0.542 | **p=0.031** lag 6: EDF=0.00, p=0.241 lag 10: EDF=0.29, p=0.202 | p=0.886 **lag 4: EDF=0.85, p=0.011** lag 3: EDF=0.00, p=0.662 lag 9: EDF=0.00, p=0.937 | p=0.643 **lag 5: EDF=0.76, p=0.028** lag 0: EDF=0.00, p=0.932 lag 6: EDF=0.38, p=0.178 | p=0.091 **lag 0: EDF=0.84, p=0.004** lag 5: EDF=0.60, p=0.116 lag 2: EDF=0.00, p=0.710 | 1.00), p=0.092 lag 7: EDF=0.41, p=0.226 lag 7: EDF=0.00, p=0.615 lag 1: EDF=0.00, p=0.597 | p=0.101 lag 6: EDF=0.00, p=0.509 lag 0: EDF=0.00, p=0.371 lag 10: EDF=0.51, p=0.121 | p=0.829 **lag 3: EDF=0.75, p=0.047** lag 3: EDF=0.77, p=0.079 **lag 5: EDF=0.89, p=0.005** |
| Heat wave day | lag 7: 1.13 (0.98–1.29), p=0.093 lag 10: 0.98 (0.91–1.06), p=0.606 lag 5: 0.92 (0.83–1.03), p=0.152 **lag 4: 0.90 (0.83–0.97), p=0.005** | lag 4: 0.89 (0.76–1.05), p=0.166 lag 0: 0.96 (0.90–1.04), p=0.323 lag 3: 0.95 (0.84–1.07), p=0.399 **lag 3: 0.93 (0.86–1.00), p=0.048** | **lag 10: 0.81 (0.70–0.94), p=0.005** lag 1: 1.00 (0.94–1.07), p=0.960 lag 2: 0.93 (0.84–1.03), p=0.156 lag 2: 0.95 (0.89–1.02), p=0.171 | lag 10: 0.93 (0.80–1.09), p=0.374 lag 1: 0.94 (0.88–1.01), p=0.078 lag 10: 0.89 (0.78–1.01), p=0.078 lag 7: 0.95 (0.87–1.02), p=0.165 | lag 3: 1.09 (0.88–1.36), p=0.426 **lag 10: 0.88 (0.79–0.98), p=0.015** lag 6: 0.88 (0.73–1.06), p=0.172 **lag 4: 0.86 (0.76–0.97), p=0.011** | **lag 1: 0.75 (0.57–0.99), p=0.045** **lag 6: 0.84 (0.73–0.96), p=0.011** **lag 1: 1.21 (1.02–1.43), p=0.026** **lag 5: 0.85 (0.74–0.98), p=0.022** | lag 7: 1.16 (0.97–1.38), p=0.096 lag 8: 0.96 (0.88–1.04), p=0.281 lag 3: 0.99 (0.85–1.14), p=0.858 lag 8: 0.93 (0.86–1.02), p=0.111 | **lag 9: 0.68 (0.48–0.96), p=0.030** lag 3: 0.95 (0.83–1.08), p=0.432 **lag 0: 1.28 (1.06–1.54), p=0.009** lag 1: 0.98 (0.86–1.12), p=0.742 | lag 2: 0.54 (0.26–1.11), p=0.095 lag 4: 0.87 (0.71–1.06), p=0.178 lag 5: 1.34 (0.91–1.95), p=0.135 lag 6: 0.93 (0.72–1.19), p=0.550 | **lag 0: 0.84 (0.70–1.00), p=0.049** **lag 0: 0.93 (0.88–1.00), p=0.040** lag 7: 0.90 (0.80–1.02), p=0.090 lag 8: 0.95 (0.88–1.02), p=0.179 | lag 3: 0.73 (0.50–1.08), p=0.115 lag 1: 0.99 (0.85–1.15), p=0.901 **lag 8: 0.56 (0.41–0.78), p=<0.001** lag 0: 1.09 (0.93–1.28), p=0.298 |
| Heat wave intensity | lag 0: EDF=0.00, p=0.870 lag 8: EDF=0.34, p=0.200 lag 5: EDF=0.00, p=0.747 lag 4: EDF=0.00, p=0.931 | lag 3: EDF=0.00, p=0.750 lag 7: EDF=0.08, p=0.273 **lag 3: EDF=0.77, p=0.030** lag 10: EDF=0.21, p=0.240 | lag 3: EDF=0.00, p=0.494 lag 3: EDF=0.00, p=0.413 lag 2: EDF=0.00, p=0.406 lag 1: EDF=0.00, p=0.581 | lag 10: EDF=0.00, p=0.667 lag 0: EDF=0.00, p=0.377 lag 10: EDF=0.00, p=0.562 lag 8: EDF=0.00, p=0.359 | lag 9: EDF=0.10, p=0.342 lag 5: EDF=0.00, p=0.442 lag 7: EDF=0.44, p=0.188 **lag 0: EDF=0.83, p=0.009** | lag 1: EDF=0.00, p=0.757 **lag 9: EDF=0.79, p=0.022** lag 7: EDF=0.71, p=0.085 **lag 7: EDF=0.87, p=0.004** | lag 9: EDF=0.70, p=0.071 lag 7: EDF=0.32, p=0.203 lag 6: EDF=0.00, p=0.426 **lag 0: EDF=0.87, p=0.004** | **lag 10: EDF=0.84, p=0.038** **lag 4: EDF=0.79, p=0.023** lag 3: EDF=0.00, p=0.743 lag 1: EDF=0.00, p=0.685 | **lag 10: EDF=0.94, p=0.047** lag 0: EDF=0.00, p=0.631 lag 10: EDF=0.76, p=0.200 lag 8: EDF=0.80, p=0.058 | lag 0: EDF=0.00, p=0.836 lag 0: EDF=0.00, p=0.518 lag 5: EDF=0.00, p=0.974 lag 8: EDF=0.39, p=0.181 | lag 4: EDF=0.00, p=0.669 lag 2: EDF=0.00, p=0.459 lag 7: EDF=0.00, p=0.753 lag 9: EDF=0.00, p=0.569 |

| Atmospheric factor | 1.1 | 1.2 | 2.1 | 2.2 | 3.1 | 4.1 | 5.1 | 5.2 | 6.1 | 6.2 | 6.3 |
|---|---|---|---|---|---|---|---|---|---|---|---|
| Cold wave duration | **lag 6: EDF=0.86, p=0.006**<br>**lag 6: EDF=0.85, p=0.006**<br>lag 1: EDF=0.00, p=0.652<br>lag 0: EDF=0.30, p=0.206 | lag 10: EDF=0.65, p=0.075<br>lag 5: EDF=0.00, p=0.591<br>lag 10: EDF=0.63, p=0.084<br>lag 1: EDF=0.00, p=0.325 | lag 7: EDF=0.00, p=0.624<br>**lag 7: EDF=0.81, p=0.016**<br>lag 10: EDF=0.58, p=0.090<br>lag 0: EDF=0.47, p=0.149 | lag 6: EDF=0.00, p=0.744<br>lag 1: EDF=0.00, p=0.560<br>lag 0: EDF=0.67, p=0.055<br>lag 6: EDF=0.00, p=0.617 | lag 2: EDF=0.59, p=0.171<br>**lag 9: EDF=0.71, p=0.044**<br>**lag 6: EDF=0.75, p=0.039**<br>**lag 5: EDF=0.72, p=0.044** | **lag 10: EDF=0.82, p=0.044**<br>lag 3: EDF=0.00, p=0.442<br>lag 10: EDF=0.00, p=0.487<br>lag 10: EDF=0.00, p=0.714 | lag 3: EDF=0.23, p=0.268<br>lag 10: EDF=0.00, p=0.333<br>lag 8: EDF=0.00, p=0.806<br>lag 10: EDF=0.00, p=0.337 | lag 5: EDF=0.76, p=0.080<br>**lag 0: EDF=0.69, p=0.050**<br>lag 9: EDF=0.24, p=0.273<br>lag 9: EDF=0.60, p=0.094 | lag 8: EDF=0.00, p=0.881<br>lag 10: EDF=0.00, p=0.486<br>lag 0: EDF=0.81, p=0.125<br>lag 10: EDF=0.40, p=0.242 | lag 10: EDF=0.63, p=0.114<br>lag 10: EDF=0.00, p=0.774<br>lag 0: EDF=0.49, p=0.143<br>lag 7: EDF=0.27, p=0.206 | lag 2: EDF=0.21, p=0.409<br>lag 7: EDF=0.45, p=0.183<br>lag 0: EDF=0.18, p=0.359<br>**lag 5: EDF=0.76, p=0.040** |
| Ozone | lag 7: EDF=0.00, p=0.441<br>**lag 1: EDF=0.89, p=0.002**<br>lag 6: EDF=0.00, p=0.769<br>lag 7: EDF=0.16, p=0.227 | lag 6: EDF=0.00, p=0.766<br>lag 0: EDF=0.63, p=0.080<br>lag 6: EDF=0.55, p=0.091<br>lag 6: EDF=0.00, p=0.919 | lag 6: EDF=0.00, p=0.346<br>**lag 1: 1.00 (0.99–1.00), p=0.001**<br>lag 7: EDF=0.00, p=0.547 | lag 3: EDF=0.00, p=0.682<br>lag 3: EDF=0.00, p=0.479<br>lag 2: EDF=0.00, p=0.378<br>lag 4: EDF=0.34, p=0.195 | **lag 1: EDF=0.89, p=0.013**<br>lag 4: EDF=0.54, p=0.115<br>lag 1: EDF=0.67, p=0.083<br>lag 0: EDF=0.01, p=0.304 | **lag 0: EDF=0.84, p=0.040**<br>**lag 1: EDF=0.86, p=0.009**<br>lag 2: EDF=0.24, p=0.274<br>lag 2: EDF=0.17, p=0.251 | lag 3: EDF=0.00, p=0.451<br>lag 6: EDF=0.00, p=0.274<br>**lag 3: EDF=0.87, p=0.006**<br>lag 7: EDF=0.00, p=0.587 | lag 6: EDF=0.54, p=0.199<br>lag 6: EDF=0.53, p=0.103<br>**lag 1: EDF=0.89, p=0.007**<br>lag 0: EDF=0.00, p=0.511 | lag 0: EDF=6.29, p=0.209<br>lag 5: EDF=0.00, p=0.483<br>lag 7: EDF=4.52, p=0.069<br>lag 0: EDF=0.62, p=0.151 | **lag 0: EDF=0.72, p=0.050**<br>lag 6: EDF=0.66, p=0.074<br>lag 7: EDF=0.44, p=0.155<br>lag 0: EDF=0.00, p=0.540 | lag 2: EDF=0.19, p=0.424<br>lag 4: EDF=0.00, p=0.448<br>**lag 4: EDF=0.93, p=0.014**<br>lag 1: EDF=0.75, p=0.053 |

**Table S23.** Overview of time lags, effect sizes (effective degree of freedom (EDF) or relative risk (95% confidence interval)) and p-values per human-bioclimatic region, atmospheric factor and subgroup (pink = female <60 years, orange = females ≥60 years, green = males <60 years, blue = males ≥60 years) for trauma, significant p-values (≤ 0.05) are bold

| Atmospheric factor | 1.1 | 1.2 | 2.1 | 2.2 | 3.1 | 4.1 | 5.1 | 5.2 | 6.1 | 6.2 | 6.3 |
|---|---|---|---|---|---|---|---|---|---|---|---|
| Sea-level pressure (SLP) | **lag 0: EDF=0.93, p=<0.001**<br>**lag 1: EDF=0.85, p=0.013**<br>lag 8: | **lag 4: EDF=0.83, p=0.018**<br>lag 10: EDF=0.00, p=0.587<br>lag 9: | **lag 0: 1.00 (1.00–1.00), p=<0.001**<br>**lag 3: EDF=0.92, p=0.002** | **lag 0: EDF=0.89, p=0.004**<br>**lag 1: EDF=0.92, p=0.002**<br>lag 1: | lag 9: EDF=0.00, p=0.726<br>lag 7: EDF=0.00, p=0.515<br>**lag 9:** | lag 8: EDF=0.39, p=0.180<br>**lag 2: EDF=0.83, p=0.018**<br>lag 10: | lag 6: EDF=0.51, p=0.147<br>**lag 0: EDF=0.90, p=0.003**<br>lag 0: | lag 9: EDF=0.72, p=0.053<br>**lag 4: EDF=0.75, p=0.047**<br>lag 10: | lag 0: EDF=0.57, p=0.105<br>**lag 0: EDF=0.89, p=0.003**<br>**lag 3:** | lag 2: EDF=0.00, p=0.610<br>lag 8: EDF=0.58, p=0.121<br>lag 10: | lag 4: EDF=0.10, p=0.263<br>lag 1: EDF=0.65, p=0.088<br>lag 1: |

| Atmospheric factor | 1.1 | 1.2 | 2.1 | 2.2 | 3.1 | 4.1 | 5.1 | 5.2 | 6.1 | 6.2 | 6.3 |
|---|---|---|---|---|---|---|---|---|---|---|---|
| | EDF=0.62, p=0.102<br>lag 1: EDF=0.42, p=0.185 | EDF=0.52, p=0.149<br>lag 6: EDF=0.68, p=0.074 | **lag 0: EDF=0.88, p=0.005**<br>lag 4: EDF=0.00, p=0.518 | EDF=0.03, p=0.309<br>lag 7: EDF=0.02, p=0.313 | **EDF=0.78, p=0.033**<br>lag 5: EDF=0.00, p=0.991 | EDF=0.52, p=0.141<br>lag 7: EDF=0.23, p=0.251 | EDF=0.00, p=0.526<br>lag 7: EDF=0.40, p=0.194 | EDF=0.37, p=0.201<br>**lag 10: EDF=0.76, p=0.041** | **EDF=0.74, p=0.039**<br>**lag 10: EDF=0.83, p=0.014** | EDF=0.62, p=0.102<br>**lag 1: EDF=0.86, p=0.009** | EDF=0.48, p=0.149<br>**lag 8: EDF=0.77, p=0.035** |
| Tropical night | lag 5: 0.93 (0.67–1.27), p=0.641<br>lag 8: 1.15 (0.99–1.32), p=0.061<br>lag 9: 0.93 (0.76–1.15), p=0.510<br>lag 7: 0.91 (0.77–1.08), p=0.295 | lag 8: 0.89 (0.76–1.05), p=0.173<br>lag 3: 1.00 (0.91–1.10), p=0.967<br>lag 3: 0.94 (0.84–1.05), p=0.287<br>lag 1: 1.08 (0.99–1.17), p=0.100 | **lag 8: 1.54 (1.15–2.07), p=0.004**<br>lag 7: 0.82 (0.65–1.03), p=0.094<br>lag 9: 1.24 (0.99–1.57), p=0.067<br>lag 7: 0.92 (0.73–1.17), p=0.507 | **lag 8: 1.15 (1.01–1.31), p=0.033**<br>lag 1: 0.95 (0.88–1.03), p=0.221<br>lag 6: 0.95 (0.86–1.06), p=0.357<br>lag 9: 0.97 (0.90–1.05), p=0.515 | - | **lag 7: 2.58 (1.50–4.46), p=<0.001**<br>lag 3: 0.54 (0.29–1.02), p=0.059<br>lag 1: 1.38 (0.87–2.18), p=0.173<br>lag 7: 0.83 (0.49–1.39), p=0.472 | lag 6: 0.60 (0.26–1.42), p=0.245<br>lag 5: 1.26 (0.97–1.65), p=0.088<br>lag 0: 0.69 (0.40–1.18), p=0.174<br>lag 7: 0.95 (0.65–1.41), p=0.816 | lag 3: 0.32 (0.10–1.05), p=0.061<br>lag 7: 0.65 (0.41–1.02), p=0.062<br>lag 7: 1.38 (0.93–2.05), p=0.111<br>**lag 7: 1.45 (1.05–1.99), p=0.023** | **lag 10: 2.31 (1.28–4.17), p=0.005**<br>lag 7: 0.70 (0.44–1.12), p=0.136<br>**lag 3: 1.66 (1.04–2.65), p=0.034**<br>**lag 3: 1.52 (1.09–2.11), p=0.014** | lag 7: 1.12 (0.95–1.32), p=0.170<br>lag 6: 1.00 (0.91–1.09), p=0.963<br>lag 7: 1.10 (0.98–1.24), p=0.096<br>lag 6: 0.97 (0.88–1.07), p=0.525 | **lag 1: 1.58 (1.07–2.33), p=0.021**<br>lag 4: 1.13 (0.89–1.43), p=0.316<br>lag 8: 1.36 (0.99–1.89), p=0.060<br>lag 0: 1.21 (0.95–1.52), p=0.117 |
| Hot day | lag 5: 0.95 (0.86–1.05), p=0.283<br>lag 9: 0.97 (0.92–1.03), p=0.306<br>lag 7: 0.97 (0.91–1.04), p=0.405<br>**lag 5: 0.93 (0.89–0.98), p=0.009** | **lag 5: 0.93 (0.87–1.00), p=0.043**<br>lag 0: 0.98 (0.93–1.02), p=0.350<br>**lag 0: 0.93 (0.88–0.98), p=0.012**<br>lag 3: 0.97 (0.93–1.01), p=0.100 | lag 7: 0.97 (0.91–1.03), p=0.293<br>lag 0: 1.00 (0.96–1.04), p=0.889<br>lag 9: 1.00 (0.95–1.05), p=0.986<br>**lag 6: 0.96 (0.93–1.00), p=0.042** | **lag 0: 0.93 (0.86–1.00), p=0.042**<br>lag 0: 1.01 (0.97–1.05), p=0.718<br>lag 4: 0.98 (0.94–1.02), p=0.411<br>lag 0: 0.98 (0.95–1.02), p=0.390 | lag 7: 0.96 (0.81–1.13), p=0.608<br>lag 0: 0.98 (0.90–1.07), p=0.696<br>**lag 2: 0.89 (0.79–0.99), p=0.038**<br>lag 0: 0.95 (0.87–1.03), p=0.221 | **lag 7: 1.75 (1.35–2.26), p=<0.001**<br>lag 8: 1.15 (0.98–1.35), p=0.093<br>**lag 7: 0.70 (0.52–0.93), p=0.015**<br>**lag 2: 1.28 (1.10–1.49), p=0.001** | lag 3: 0.97 (0.86–1.10), p=0.682<br>lag 1: 0.95 (0.90–1.01), p=0.085<br>lag 9: 1.07 (0.98–1.15), p=0.120<br>**lag 6: 0.93 (0.88–0.99), p=0.021** | **lag 5: 0.80 (0.68–0.93), p=0.005**<br>lag 8: 0.98 (0.91–1.05), p=0.516<br>lag 8: 1.10 (1.00–1.20), p=0.052<br>**lag 7: 0.91 (0.85–0.98), p=0.013** | lag 1: 1.26 (0.99–1.61), p=0.057<br>lag 7: 0.93 (0.81–1.06), p=0.267<br>lag 2: 0.89 (0.71–1.11), p=0.293<br>**lag 0: 0.83 (0.72–0.96), p=0.013** | lag 0: 0.96 (0.89–1.03), p=0.273<br>lag 1: 0.98 (0.95–1.01), p=0.167<br>lag 2: 1.05 (0.99–1.11), p=0.077<br>lag 8: 0.99 (0.96–1.02), p=0.442 | lag 7: 0.89 (0.74–1.06), p=0.198<br>lag 10: 1.04 (0.96–1.13), p=0.289<br>lag 5: 0.89 (0.79–1.01), p=0.076<br>lag 8: 0.99 (0.90–1.09), p=0.831 |

| Atmospheric factor | 1.1 | 1.2 | 2.1 | 2.2 | 3.1 | 4.1 | 5.1 | 5.2 | 6.1 | 6.2 | 6.3 |
|---|---|---|---|---|---|---|---|---|---|---|---|
| Wet-bulb globe temperature | **lag 0: EDF=4.73, p=<0.001**<br>lag 0: EDF=0.00, p=0.653<br>lag 5: EDF=1.98, p=0.066<br>lag 0: EDF=0.00, p=0.643 | **lag 0: EDF=2.06, p=<0.001**<br>**lag 0: EDF=0.91, p=0.010** | lag 1: EDF=0.00, p=0.444<br>**lag 0: EDF=4.17, p=<0.001**<br>**lag 0: EDF=2.98, p=<0.001** | lag 6: EDF=0.00, p=0.523 | **lag 0: EDF=1.20, p=<0.001**<br>lag 1: EDF=0.00, p=0.647 | lag 1: EDF=0.28, p=0.214 | lag 4: EDF=0.00, p=0.416<br>**lag 0: EDF=2.32, p=<0.001**<br>**lag 0: EDF=1.02, p=<0.001** | **lag 0: EDF=0.96, p=<0.001** | **lag 0: EDF=0.72, p=0.046**<br>lag 0: EDF=0.00, p=0.806<br>lag 1: EDF=0.00, p=0.715 | **lag 0: EDF=1.65, p=<0.001**<br>lag 0: EDF=0.64, p=0.093 | lag 6: EDF=0.60, p=0.092 |
| Daily mean temperature | - | - | lag 0: EDF=0.00, p=0.766 | - | - | lag 0: EDF=0.00, p=0.563<br>**lag 10: EDF=2.46, p=0.049** | lag 0: EDF=0.37, p=0.202 | lag 0: EDF=0.00, p=0.445 | - | **lag 0: 0.99 (0.99–1.00), p=0.011** | lag 0: EDF=0.63, p=0.081 |
| Ice day | lag 4: 1.02 (0.96–1.08), p=0.534<br>lag 8: 1.02 (0.99–1.06), p=0.189<br>lag 5: 1.01 (0.96–1.07), p=0.593<br>lag 10: 1.01 (0.97–1.05), p=0.610 | **lag 8: 1.07 (1.01–1.13), p=0.012**<br>**lag 9: 1.04 (1.01–1.07), p=0.014**<br>lag 9: 1.00 (0.96–1.05), p=0.825<br>**lag 0: 1.05 (1.01–1.08), p=0.015** | lag 5: 1.03 (0.99–1.08), p=0.142<br>lag 0: 1.01 (0.99–1.04), p=0.321<br>lag 0: 0.98 (0.94–1.03), p=0.525 | lag 4: 1.05 (1.00–1.11), p=0.053<br>lag 9: 1.02 (1.00–1.05), p=0.091<br>lag 4: 1.00 (0.96–1.04), p=0.948<br>**lag 7: 1.05 (1.03–1.08), p=<0.001** | lag 0: 1.06 (0.99–1.13), p=0.072<br>lag 6: 1.03 (1.00–1.06), p=0.070<br>lag 2: 0.99 (0.94–1.04), p=0.667<br>lag 10: 1.03 (1.00–1.06), p=0.091 | lag 5: 1.04 (0.96–1.12), p=0.391<br>lag 7: 1.04 (1.00–1.09), p=0.070<br>lag 0: 1.04 (0.98–1.10), p=0.239<br>lag 6: 1.04 (0.99–1.09), p=0.116 | lag 8: 1.06 (1.00–1.12), p=0.065<br>**lag 10: 1.03 (1.01–1.06), p=0.014**<br>**lag 10: 1.05 (1.01–1.10), p=0.017**<br>lag 9: 1.03 (1.00–1.05), p=0.062 | lag 2: 1.01 (0.94–1.07), p=0.850<br>lag 0: 0.99 (0.95–1.02), p=0.530<br>lag 8: 0.96 (0.92–1.01), p=0.106<br>lag 6: 1.03 (0.99–1.06), p=0.101 | **lag 9: 1.15 (1.01–1.31), p=0.033**<br>lag 3: 1.03 (0.97–1.10), p=0.263<br>**lag 4: 1.10 (1.00–1.20), p=0.043**<br>lag 8: 1.04 (0.98–1.11), p=0.174 | lag 4: 1.04 (0.99–1.08), p=0.122<br>**lag 10: 1.02 (1.00–1.04), p=0.038**<br>lag 4: 0.97 (0.93–1.00), p=0.064<br>**lag 8: 1.02 (1.00–1.05), p=0.036** | **lag 8: 1.23 (1.10–1.37), p=<0.001**<br>**lag 1: 0.92 (0.86–0.98), p=0.006**<br>lag 7: 0.98 (0.90–1.07), p=0.700<br>**lag 5: 0.93 (0.87–0.98), p=0.010** |
| Frost day | **lag 5: 1.04 (1.01–1.08), p=0.015** | **lag 0: 1.04 (1.01–1.08), p=0.019** | lag 0: 1.02 (0.99–1.05), p=0.139 | lag 2: 1.00 (0.97–1.03), p=0.874 | lag 6: 1.04 (1.00–1.08), p=0.075 | lag 1: 1.02 (0.97–1.08), p=0.370 | lag 9: 1.02 (0.99–1.06), p=0.248 | lag 5: 1.03 (0.99–1.07), p=0.110 | lag 7: 1.03 (0.95–1.12), p=0.514 | lag 2: 1.01 (0.98–1.04), p=0.507 | **lag 4: 1.11 (1.04–1.18), p=0.001** |

| Atmospheric factor | 1.1 | 1.2 | 2.1 | 2.2 | 3.1 | 4.1 | 5.1 | 5.2 | 6.1 | 6.2 | 6.3 |
|---|---|---|---|---|---|---|---|---|---|---|---|
| | lag 0: 1.01 (0.99–1.03), p=0.246<br>lag 0: 1.00 (0.97–1.02), p=0.944<br>lag 0: 1.00 (0.98–1.02), p=0.710 | lag 0: 1.00 (0.98–1.02), p=0.849<br>lag 0: 1.02 (0.99–1.04), p=0.178<br>**lag 7: 1.03 (1.01–1.05), p=<0.001** | **lag 0: 1.02 (1.00–1.03), p=0.020**<br>lag 9: 1.02 (1.00–1.03), p=0.071<br>lag 0: 0.99 (0.98–1.01), p=0.395 | lag 0: 1.01 (1.00–1.03), p=0.168<br>lag 6: 1.01 (0.99–1.03), p=0.413<br>**lag 0: 0.98 (0.97–1.00), p=0.024** | lag 0: 1.00 (0.98–1.03), p=0.820<br>lag 0: 1.02 (0.98–1.06), p=0.286<br>lag 0: 0.99 (0.97–1.02), p=0.585 | lag 5: 1.01 (0.98–1.03), p=0.694<br>**lag 2: 1.04 (1.01–1.09), p=0.026**<br>lag 1: 1.00 (0.97–1.03), p=0.922 | lag 0: 0.99 (0.97–1.01), p=0.168<br>lag 0: 1.00 (0.97–1.04), p=0.778<br>lag 0: 1.01 (0.99–1.03), p=0.312 | lag 0: 1.02 (0.99–1.04), p=0.142<br>lag 2: 0.98 (0.95–1.01), p=0.126<br>lag 0: 1.00 (0.97–1.02), p=0.854 | lag 4: 1.03 (0.99–1.07), p=0.120<br>lag 1: 1.01 (0.95–1.07), p=0.761<br>lag 1: 1.02 (0.99–1.06), p=0.204 | lag 0: 1.00 (0.99–1.02), p=0.839<br>lag 1: 0.99 (0.97–1.01), p=0.340<br>lag 0: 1.01 (1.00–1.03), p=0.103 | **lag 0: 1.04 (1.00–1.08), p=0.043**<br>lag 8: 0.95 (0.91–1.00), p=0.060<br>**lag 8: 1.04 (1.00–1.07), p=0.030** |
| Wind chill index | - | **lag 0: EDF=0.90, p=0.007**<br>lag 0: EDF=0.12, p=0.286 | - | lag 0: EDF=0.00, p=0.925<br>lag 0: EDF=0.00, p=0.681 | lag 0: EDF=0.23, p=0.224 | lag 0: EDF=0.57, p=0.120 | lag 0: EDF=0.21, p=0.255 | - | lag 10: EDF=0.00, p=0.395<br>lag 0: EDF=0.00, p=0.465 | lag 0: EDF=0.00, p=0.505<br>lag 3: EDF=0.00, p=0.498 | **lag 9: EDF=0.72, p=0.044**<br>lag 0: EDF=0.00, p=0.839 |
| Heat wave | **lag 9: 0.94 (0.91–0.98), p=0.005**<br>lag 0: 0.97 (0.94–1.01), p=0.190 | lag 0: 1.01 (0.98–1.05), p=0.450 | lag 8: 0.99 (0.95–1.04), p=0.746<br>lag 2: 1.00 (0.97–1.03), p=0.860 | **lag 0: 0.94 (0.91–0.96), p=<0.001**<br>lag 0: 0.98 (0.95–1.01), p=0.131 | lag 7: 0.92 (0.84–1.02), p=0.121<br>lag 0: 0.95 (0.90–1.01), p=0.095<br>**lag 4: 0.94 (0.89–0.99), p=0.024**<br>**lag 0: 0.96 (0.92–1.00), p=0.046** | lag 8: 0.92 (0.83–1.02), p=0.113<br>lag 8: 1.00 (0.94–1.08), p=0.901<br>lag 0: 0.97 (0.93–1.01), p=0.143 | lag 0: 0.97 (0.90–1.03), p=0.288<br>lag 0: 1.02 (0.98–1.06), p=0.339<br>lag 9: 1.01 (0.97–1.06), p=0.589<br>**lag 0: 0.95 (0.92–0.99), p=0.004** | lag 4: 1.04 (0.94–1.15), p=0.476<br>lag 1: 0.97 (0.91–1.03), p=0.315<br>lag 0: 0.98 (0.94–1.03), p=0.476 | lag 10: 0.83 (0.69–1.00), p=0.052<br>lag 0: 0.97 (0.89–1.05), p=0.451<br>lag 6: 0.96 (0.84–1.10), p=0.545<br>**lag 7: 0.94 (0.88–1.00), p=0.049** | lag 4: 0.99 (0.95–1.04), p=0.697<br>lag 10: 1.01 (0.98–1.05), p=0.448<br>lag 0: 0.99 (0.96–1.02), p=0.356 | lag 10: 1.11 (0.99–1.24), p=0.062<br>lag 2: 1.03 (0.97–1.09), p=0.326<br>lag 5: 0.98 (0.89–1.07), p=0.581<br>lag 2: 0.99 (0.93–1.05), p=0.688 |
| Heat wave day | lag 0: 0.97 (0.91–1.04), p=0.439 | lag 0: 0.99 (0.95–1.03), p=0.566 | lag 1: 0.99 (0.95–1.04), p=0.800 | **lag 8: 0.97 (0.94–1.00), p=0.022** | lag 9: 0.99 (0.92–1.07), p=0.840 | lag 9: 0.98 (0.90–1.06), p=0.576 | lag 3: 0.98 (0.93–1.05), p=0.609 | lag 5: 0.98 (0.90–1.06), p=0.616 | lag 8: 0.95 (0.85–1.06), p=0.328 | lag 0: 0.99 (0.95–1.03), p=0.647 | lag 6: 0.94 (0.85–1.04), p=0.241 |

| Atmospheric factor | 1.1 | 1.2 | 2.1 | 2.2 | 3.1 | 4.1 | 5.1 | 5.2 | 6.1 | 6.2 | 6.3 |
|---|---|---|---|---|---|---|---|---|---|---|---|
| | lag 0: 1.00 (0.98–1.03), p=0.744<br>lag 1: 0.98 (0.95–1.01), p=0.186<br>**lag 3: 0.97 (0.95–1.00), p=0.031** | lag 0: 0.98 (0.95–1.00), p=0.082<br>**lag 0: 0.97 (0.95–1.00), p=0.038** | lag 0: 0.98 (0.95–1.00), p=0.073<br>lag 10: 1.00 (0.98–1.03), p=0.836<br>lag 0: 0.98 (0.95–1.01), p=0.116 | | **lag 3: 0.94 (0.91–0.97), p=<0.001**<br>lag 0: 0.98 (0.93–1.03), p=0.402<br>**lag 9: 0.96 (0.93–0.99), p=0.008** | lag 0: 0.98 (0.94–1.01), p=0.181<br>lag 10: 0.96 (0.90–1.02), p=0.188 | | lag 0: 0.99 (0.96–1.02), p=0.558<br>lag 3: 0.95 (0.89–1.02), p=0.166<br>lag 2: 0.99 (0.96–1.03), p=0.642 | **lag 0: 0.91 (0.83–1.00), p=0.041** | lag 0: 0.98 (0.96–1.00), p=0.077<br>lag 6: 1.01 (0.98–1.03), p=0.689 | lag 0: 1.04 (0.99–1.10), p=0.133<br>lag 9: 0.95 (0.88–1.03), p=0.207 |
| Heat wave intensity | lag 1: EDF=0.64, p=0.094<br>lag 7: EDF=0.00, p=0.423<br>lag 9: EDF=0.00, p=0.918<br>lag 3: EDF=0.29, p=0.232 | lag 0: EDF=0.00, p=0.888<br>**lag 3: EDF=0.82, p=0.019**<br>lag 10: EDF=0.37, p=0.206<br>**lag 4: EDF=0.87, p=0.006** | lag 3: EDF=0.00, p=0.453<br>lag 0: EDF=0.00, p=0.336<br>lag 9: EDF=0.00, p=0.870<br>lag 4: EDF=0.00, p=0.394 | lag 0: EDF=0.01, p=0.342<br>lag 0: EDF=0.00, p=0.715<br>lag 8: EDF=0.00, p=0.396<br>lag 0: EDF=0.35, p=0.208 | lag 7: EDF=0.01, p=0.313<br>lag 0: EDF=0.00, p=0.333<br>lag 5: EDF=0.00, p=0.531<br>lag 0: EDF=0.00, p=0.363 | lag 8: EDF=0.01, p=0.333<br>lag 0: EDF=0.02, p=0.308<br>**lag 4: EDF=1.71, p=0.006**<br>lag 0: EDF=0.00, p=0.372 | lag 10: EDF=0.00, p=0.382<br>**lag 0: EDF=0.84, p=0.013**<br>lag 3: EDF=0.43, p=0.179<br>lag 0: EDF=0.00, p=0.928 | lag 5: EDF=0.00, p=0.690<br>lag 6: EDF=0.00, p=0.405<br>**lag 3: EDF=1.95, p=0.031**<br>lag 0: EDF=0.00, p=0.450 | lag 10: EDF=0.00, p=0.546<br>lag 4: EDF=0.00, p=0.506<br>lag 0: EDF=0.00, p=0.813<br>**lag 0: EDF=2.55, p=0.002** | **lag 7: EDF=0.82, p=0.016**<br>lag 0: EDF=0.00, p=0.765<br>lag 9: EDF=0.00, p=0.772<br>lag 0: EDF=0.06, p=0.301 | lag 1: EDF=0.66, p=0.066<br>lag 3: EDF=0.00, p=0.739<br>**lag 8: EDF=2.38, p=0.008**<br>lag 3: EDF=0.62, p=0.099 |
| Heat stress wave intensity | lag 5: 0.97 (0.93–1.01), p=0.165<br>lag 0: 1.00 (0.97–1.03), p=0.831 | lag 1: 1.01 (0.97–1.05), p=0.804<br>lag 1: 0.98 (0.95–1.01), p=0.163 | lag 2: 0.99 (0.95–1.04), p=0.780<br>lag 9: 1.02 (0.99–1.05), p=0.210<br>lag 7: 0.99 (0.97–1.01), p=0.168 | lag 0: 1.02 (0.97–1.07), p=0.391<br>lag 0: 1.01 (0.98–1.04), p=0.427 | lag 1: 0.94 (0.88–1.01), p=0.071<br>lag 0: 1.03 (0.98–1.07), p=0.254 | lag 7: 1.00 (0.95–1.06), p=0.987 | lag 4: 1.00 (0.94–1.06), p=0.995<br>lag 0: 1.00 (0.96–1.05), p=0.911 | lag 8: 0.94 (0.88–1.01), p=0.077<br>lag 6: 0.97 (0.94–1.01), p=0.146<br>**lag 6: 0.93 (0.89–0.98), p=0.005**<br>lag 3: 0.97 (0.94– | lag 10: 0.95 (0.82–1.10), p=0.475<br>lag 0: 0.95 (0.89–1.02), p=0.142<br>**lag 0: 0.88 (0.81–0.97), p=0.007**<br>lag 1: 0.97 (0.91– | lag 2: 1.00 (0.96–1.03), p=0.866<br>lag 0: 1.00 (0.98–1.03), p=0.842 | lag 8: 1.10 (1.00–1.21), p=0.061<br>**lag 3: 1.11 (1.04–1.18), p=0.002**<br>lag 8: 0.96 (0.91–1.01), p=0.153 |

| Atmospheric factor | 1.1 | 1.2 | 2.1 | 2.2 | 3.1 | 4.1 | 5.1 | 5.2 | 6.1 | 6.2 | 6.3 |
|---|---|---|---|---|---|---|---|---|---|---|---|
| | | | | | | | | 1.01),<br>p=0.161 | 1.03),<br>p=0.263 | | |
| Cold wave duration | lag 9:<br>EDF=0.00,<br>p=0.662<br>lag 5:<br>EDF=0.00,<br>p=0.826<br>**lag 8:<br>EDF=0.73,<br>p=0.050** | **lag 10:<br>EDF=0.93,<br>p=<0.001**<br>**lag 4: 1.01<br>(1.00–<br>1.01),<br>p=<0.001**<br>**lag 0:<br>EDF=0.76,<br>p=0.037** | **lag 0:<br>EDF=0.74,<br>p=0.048**<br>**lag 0:<br>EDF=0.74,<br>p=0.047**<br>lag 2:<br>EDF=0.00,<br>p=0.330<br>**lag 0: 1.01<br>(1.00–<br>1.01),<br>p=<0.001** | **lag 1:<br>EDF=0.86,<br>p=0.006**<br>**lag 0:<br>EDF=2.34,<br>p=<0.001**<br>lag 0:<br>EDF=0.00,<br>p=0.865<br>lag 0:<br>EDF=0.00,<br>p=0.731 | lag 1:<br>EDF=0.00,<br>p=0.393<br>**lag 0:<br>EDF=0.89,<br>p=0.001**<br>lag 0:<br>EDF=0.20,<br>p=0.257 | lag 3:<br>EDF=0.00,<br>p=0.431<br>lag 9:<br>EDF=0.56,<br>p=0.117<br>lag 6:<br>EDF=1.65,<br>p=0.081 | **lag 7:<br>EDF=0.74,<br>p=0.048**<br>lag 0:<br>EDF=0.71,<br>p=0.061<br>**lag 6:<br>EDF=0.82,<br>p=0.017**<br>lag 0:<br>EDF=0.29,<br>p=0.229 | **lag 0:<br>EDF=0.85,<br>p=0.006**<br>**lag 0:<br>EDF=0.79,<br>p=0.026**<br>**lag 1:<br>EDF=0.91,<br>p=<0.001**<br>lag 0:<br>EDF=0.34,<br>p=0.202 | lag 7:<br>EDF=0.00,<br>p=0.966<br>lag 4:<br>EDF=0.00,<br>p=0.515 | **lag 0:<br>EDF=0.92,<br>p=<0.001**<br>lag 10:<br>EDF=0.17,<br>p=0.266 | lag 7:<br>EDF=0.00,<br>p=0.324<br>lag 3:<br>EDF=0.26,<br>p=0.232 |
| Cold stress wave intensity | lag 10:<br>EDF=0.00,<br>p=0.463<br>lag 0:<br>EDF=0.47,<br>p=0.166<br>lag 10:<br>EDF=1.55,<br>p=0.119 | lag 0:<br>EDF=0.01,<br>p=0.316<br>lag 0:<br>EDF=0.09,<br>p=0.290<br>lag 4:<br>EDF=0.00,<br>p=0.796 | lag 10:<br>EDF=0.33,<br>p=0.222<br>lag 0:<br>EDF=0.70,<br>p=0.061 | lag 5:<br>EDF=0.28,<br>p=0.226<br>lag 4:<br>EDF=0.67,<br>p=0.077 | **lag 3:<br>EDF=0.77,<br>p=0.029**<br>lag 0:<br>EDF=0.00,<br>p=0.694<br>lag 5:<br>EDF=0.49,<br>p=0.145 | **lag 10:<br>EDF=0.78,<br>p=0.026**<br>lag 1:<br>EDF=0.00,<br>p=0.896<br>lag 1:<br>EDF=0.00,<br>p=0.609 | lag 1:<br>EDF=0.00,<br>p=0.839<br>**lag 9:<br>EDF=0.75,<br>p=0.042** | lag 5:<br>EDF=0.25,<br>p=0.230 | lag 6:<br>EDF=0.00,<br>p=0.408<br>lag 1:<br>EDF=0.00,<br>p=0.496<br>lag 1:<br>EDF=0.00,<br>p=0.759<br>**lag 0:<br>EDF=0.89,<br>p=0.002** | lag 8:<br>EDF=0.41,<br>p=0.188<br>lag 0:<br>EDF=0.00,<br>p=0.535<br>lag 8:<br>EDF=0.00,<br>p=0.467 | **lag 0:<br>EDF=0.93,<br>p=<0.001**<br>lag 1:<br>EDF=0.00,<br>p=0.489<br>lag 10:<br>EDF=0.12,<br>p=0.266 |
| Ozone | lag 0:<br>EDF=0.00,<br>p=0.771<br>**lag 0:<br>EDF=2.52,<br>p=0.021**<br>lag 4:<br>EDF=0.00,<br>p=0.795<br>lag 0: | - | - | lag 0:<br>EDF=0.52,<br>p=0.147<br>**lag 0:<br>EDF=3.35,<br>p=<0.001**<br>lag 3:<br>EDF=0.00,<br>p=0.405<br>lag 0: | lag 7:<br>EDF=0.00,<br>p=0.819<br>lag 0:<br>EDF=0.00,<br>p=0.797<br>**lag 0: 1.00<br>(1.00–<br>1.01),<br>p=<0.001**<br>lag 0: | lag 0:<br>EDF=0.00,<br>p=0.813<br>lag 0:<br>EDF=0.00,<br>p=0.651<br>lag 7:<br>EDF=0.69,<br>p=0.069<br>lag 0: | lag 0:<br>EDF=0.00,<br>p=0.502<br>lag 2:<br>EDF=0.66,<br>p=0.084 | lag 6:<br>EDF=0.00,<br>p=0.497<br>**lag 0:<br>EDF=0.79,<br>p=0.032** | lag 6:<br>EDF=0.00,<br>p=0.882<br>lag 1:<br>EDF=0.07,<br>p=0.282<br>lag 5:<br>EDF=0.00,<br>p=0.417<br>lag 1: | lag 0:<br>EDF=0.12,<br>p=0.280<br>lag 0:<br>EDF=0.01,<br>p=0.417<br>**lag 3:<br>EDF=0.94,<br>p=0.002** | lag 6:<br>EDF=0.30,<br>p=0.188<br>**lag 0:<br>EDF=3.36,<br>p=<0.001**<br>lag 5:<br>EDF=0.01,<br>p=0.343<br>**lag 2:** |

| Atmospheric factor | 1.1 | 1.2 | 2.1 | 2.2 | 3.1 | 4.1 | 5.1 | 5.2 | 6.1 | 6.2 | 6.3 |
|---|---|---|---|---|---|---|---|---|---|---|---|
| | EDF=0.00, p=0.610 | | | EDF=0.00, p=0.411 | EDF=0.68, p=0.065 | EDF=0.00, p=0.436 | | | EDF=0.00, p=0.682 | | **EDF=0.73, p=0.049** |
| Nitrogen dioxide | lag 0: EDF=0.09, p=0.284<br>lag 0: EDF=0.04, p=0.305<br>lag 0: EDF=0.70, p=0.067<br>**lag 0: EDF=0.75, p=0.043** | lag 0: EDF=0.00, p=0.381<br>lag 0: EDF=0.00, p=0.690<br>lag 0: EDF=0.00, p=0.727<br>lag 4: EDF=0.60, p=0.099 | lag 0: EDF=0.71, p=0.059<br>lag 0: EDF=0.00, p=0.583<br>lag 0: EDF=0.00, p=0.740<br>lag 0: EDF=0.00, p=0.613 | lag 7: EDF=0.00, p=0.914<br>lag 0: EDF=0.68, p=0.069<br>**lag 0: EDF=1.81, p=<0.001**<br>lag 0: EDF=0.00, p=0.691 | lag 6: EDF=0.00, p=0.979<br>lag 0: EDF=0.00, p=0.841<br>**lag 7: EDF=0.87, p=0.005**<br>**lag 0: EDF=0.82, p=0.018** | lag 0: EDF=0.63, p=0.083<br>lag 0: EDF=0.00, p=0.506<br>**lag 0: EDF=2.16, p=0.031**<br>lag 0: EDF=0.72, p=0.058 | **lag 0: EDF=0.91, p=0.001**<br>lag 0: EDF=0.00, p=0.446<br>lag 2: EDF=0.00, p=0.512<br>lag 0: EDF=0.00, p=0.533 | **lag 2: EDF=0.76, p=0.034**<br>lag 0: EDF=0.00, p=0.493<br>lag 0: EDF=0.65, p=0.086<br>**lag 0: EDF=0.79, p=0.026** | lag 0: EDF=0.13, p=0.250<br>lag 2: EDF=0.00, p=0.590<br>**lag 5: EDF=0.72, p=0.043**<br>lag 6: EDF=0.61, p=0.097 | lag 0: EDF=0.00, p=0.560<br>**lag 0: EDF=0.81, p=0.020**<br>**lag 4: EDF=3.08, p=0.038**<br>lag 0: EDF=0.46, p=0.164 | lag 4: EDF=0.01, p=0.362<br>lag 0: EDF=0.46, p=0.160<br>lag 7: EDF=0.34, p=0.199<br>lag 3: EDF=0.43, p=0.172 |
| Particulate matter ($PM_{10}$) | lag 7: EDF=0.00, p=0.496 | - | **lag 6: EDF=0.90, p=0.002**<br>**lag 6: EDF=1.24, p=0.005** | lag 2: EDF=0.00, p=0.566<br>lag 0: EDF=0.00, p=0.801 | lag 0: EDF=0.00, p=0.801 | **lag 5: EDF=0.92, p=<0.001**<br>lag 6: EDF=0.01, p=0.325 | lag 2: EDF=0.00, p=0.671<br>lag 0: EDF=0.00, p=0.881<br>lag 3: EDF=0.69, p=0.072 | lag 4: EDF=0.00, p=0.814<br>lag 0: EDF=0.00, p=0.572<br>lag 3: EDF=0.44, p=0.174 | lag 6: EDF=0.44, p=0.157<br>lag 2: EDF=0.44, p=0.168<br>lag 4: EDF=0.00, p=0.653<br>lag 4: EDF=0.00, p=0.531 | **lag 3: EDF=0.87, p=0.006**<br>lag 5: EDF=0.37, p=0.205 | **lag 5: EDF=0.78, p=0.025** |
| Particulate matter ($PM_{2.5}$) | **lag 0: EDF=2.20, p=0.029**<br>**lag 6: EDF=0.83, p=0.015**<br>lag 3: EDF=0.39, p=0.197 | **lag 0: EDF=0.90, p=0.002**<br>lag 3: EDF=0.62, p=0.100<br>lag 4: EDF=0.00, p=0.636<br>lag 4: EDF=0.32, p=0.206 | **lag 6: 0.99 (0.98–1.00), p=0.009**<br>lag 7: EDF=0.00, p=0.391<br>lag 7: EDF=0.68, p=0.074 | lag 0: EDF=0.00, p=0.568<br>lag 4: EDF=0.00, p=0.770 | **lag 4: EDF=0.90, p=0.001**<br>lag 4: EDF=0.47, p=0.165<br>lag 7: EDF=0.44, p=0.170<br>**lag 6: EDF=0.79, p=0.028** | lag 0: EDF=0.00, p=0.344<br>**lag 3: EDF=0.81, p=0.023** | **lag 2: EDF=1.94, p=0.006** | **lag 6: EDF=0.88, p=0.005**<br>lag 0: EDF=0.00, p=0.678 | - | lag 0: EDF=0.00, p=0.868<br>lag 6: EDF=0.00, p=0.580 | lag 7: EDF=0.60, p=0.110<br>lag 7: EDF=0.00, p=0.886<br>lag 3: EDF=0.00, p=0.423 |

**Table S24.1.** Sex- and age-specific significant associations between daily intensive care unit admissions and atmospheric factors in Germany per cardiovascular outcome type. Significant percentage *(%)* of all 22 possible models (2 sex/age subgroups x 11 human-bioclimatic regions), order of the presented values: Females, males, <60 years, ≥60 years

| Atmospheric factor | Myocardial infarction (STEMI) | Myocardial infarction (NSTEMI) | Heart failure | Pulmonary artery embolism | Tachycardia |
|---|---|---|---|---|---|
| Sea-level pressure (SLP) | 40.9 \| 50.0 \| 50.0 \| 40.9 | 54.5 \| 63.6 \| 50.0 \| 68.2 | 40.9 \| 45.5 \| 36.4 \| 50.0 | 31.8 \| 40.9 \| 31.8 \| 40.9 | 31.8 \| 50.0 \| 40.9 \| 40.9 |
| Sea-level pressure difference (SLPD) | - | - | - | 40.9 \| 27.3 \| 36.4 \| 31.8 | - |
| Wind day | - | 36.4 \| 36.4 \| 40.9 \| 31.8 | - | 54.5 \| 36.4 \| 54.5 \| 36.4 | 54.5 \| 45.5 \| 59.1 \| 40.9 |
| Tropical night | 31.8 \| 36.4 \| 40.9 \| 27.3 | - | 22.7 \| 27.3 \| 31.8 \| 18.2 | 40.9 \| 36.4 \| 27.3 \| 50.0 | 27.3 \| 18.2 \| 18.2 \| 27.3 |
| Hot day | 36.4 \| 22.7 \| 22.7 \| 36.4 | - | - | - | 22.7 \| 36.4 \| 27.3 \| 31.8 |
| Summer day | 45.5 \| 45.5 \| 45.5 \| 45.5 | - | 31.8 \| 59.1 \| 31.8 \| 59.1 | 18.2 \| 22.7 \| 18.2 \| 22.7 | - |
| Wet-bulb globe temperature | - | - | 9.1 \| 13.6 \| 9.1 \| 13.6 | - | - |
| Daily mean temperature | - | - | 18.2 \| 13.6 \| 22.7 \| 9.1 | - | - |
| Ice day | - | - | - | 27.3 \| 36.4 \| 31.8 \| 31.8 | - |
| Frost day | - | - | - | - | - |
| Wind chill index | - | 27.3 \| 40.9 \| 40.9 \| 27.3 | - | 9.1 \| 27.3 \| 18.2 \| 18.2 | 31.8 \| 13.6 \| 18.2 \| 27.3 |
| Heat wave | - | 27.3 \| 40.9 \| 13.6 \| 54.5 | - | - | - |

| | | | | | |
|---|---|---|---|---|---|
| Heat wave day | - | - | - | - | 31.8 \| 31.8 \| 31.8 \| 31.8 |
| Heat wave duration | - | - | 31.8 \| 45.5 \| 22.7 \| 54.5 | 18.2 \| 27.3 \| 18.2 \| 27.3 | - |
| Heat wave intensity | - | - | - | 13.6 \| 13.6 \| 13.6 \| 13.6 | 18.2 \| 18.2 \| 13.6 \| 22.7 |
| Heat stress wave | - | - | - | - | - |
| Heat stress wave day | - | - | - | 22.7 \| 31.8 \| 18.2 \| 36.4 | - |
| Heat stress wave duration | - | 13.6 \| 9.1 \| 9.1 \| 13.6 | - | - | - |
| Heat stress wave intensity | - | 27.3 \| 13.6 \| 18.2 \| 22.7 | - | 9.1 \| 18.2 \| 13.6 \| 13.6 | - |
| Cold wave | - | - | - | - | - |
| Cold wave day | - | - | - | - | - |
| Cold wave duration | 18.2 \| 13.6 \| 18.2 \| 13.6 | - | - | - | 27.3 \| 13.6 \| 13.6 \| 27.3 |
| Cold wave intensity | - | - | - | - | - |
| Cold stress wave | - | - | - | - | - |
| Cold stress wave day | - | - | 18.2 \| 9.1 \| 22.7 \| 4.5 | - | - |
| Cold stress wave duration | - | - | - | - | - |

| | | | | | |
|---|---|---|---|---|---|
| Cold stress wave intensity | - | - | 9.1 \| 18.2 \| 9.1 \| 18.2 | - | - |
| Ozone | 40.9 \| 27.3 \| 36.4 \| 31.8 | - | - | 54.5 \| 54.5 \| 50.0 \| 59.1 | 27.3 \| 13.6 \| 27.3 \| 13.6 |
| Nitrogen dioxide ($NO_2$) | - | - | - | - | - |
| Particular matter ($PM_{10}$) | - | 31.8 \| 27.3 \| 31.8 \| 27.3 | 13.6 \| 22.7 \| 27.3 \| 9.1 | 27.3 \| 40.9 \| 27.3 \| 40.9 | - |
| Particular matter ($PM_{2.5}$) | - | - | - | - | - |

**Table S24.2.** Sex- and age-specific significant associations between daily intensive care unit admissions and atmospheric factors in Germany per neurological outcome type. Significant percentage *(%)* of all 22 possible models (2 sex/age subgroups x 11 human-bioclimatic regions), order of the presented values: Females, males, <60 years, ≥60 years

| Atmospheric factor | Ischemic stroke | Ischemic stroke (stroke unit) | Non subdural hemorrhage | Non subdural hemorrhage (stroke unit) | Transient ischemic attack | Transient ischemic attack (stroke unit) | Epilepsy |
|---|---|---|---|---|---|---|---|
| Sea-level pressure (SLP) | 50.0 \| 45.5 \| 40.9 \| 54.5 | - | 22.7 \| 45.5 \| 40.9 \| 27.3 | - | - | 22.7 \| 27.3 \| 18.2 \| 31.8 | - |
| Sea-level pressure difference (SLPD) | - | 36.4 \| 31.8 \| 31.8 \| 36.4 | 45.5 \| 18.2 \| 27.3 \| 36.4 | - | 63.6 \| 45.5 \| 45.5 \| 63.6 | - | 40.9 \| 36.4 \| 45.5 \| 31.8 |
| Wind day | 40.9 \| 59.1 \| 63.6 \| 36.4 | - | 40.9 \| 40.9 \| 54.5 \| 27.3 | 54.5 \| 50.0 \| 50.0 \| 54.5 | - | 36.4 \| 36.4 \| 40.9 \| 31.8 | 40.9 \| 40.9 \| 54.5 \| 27.3 |
| Tropical night | 36.4 \| 40.9 \| 40.9 \| 36.4 | 31.8 \| 27.3 \| 22.7 \| 36.4 | 45.5 \| 22.7 \| 45.5 \| 22.7 | 31.8 \| 13.6 \| 27.3 \| 18.2 | 22.7 \| 27.3 \| 31.8 \| 18.2 | - | - |
| Hot day | 31.8 \| 22.7 \| 22.7 \| 31.8 | - | 18.2 \| 18.2 \| 18.2 \| 18.2 | 31.8 \| 22.7 \| 22.7 \| 31.8 | - | - | - |
| Summer day | 36.4 \| 18.2 \| 27.3 \| 27.3 | - | 22.7 \| 27.3 \| 31.8 \| 18.2 | - | 31.8 \| 40.9 \| 27.3 \| 45.5 | 50.0 \| 54.5 \| 31.8 \| 72.7 | - |
| Wet-bulb globe temperature | - | 18.2 \| 31.8 \| 36.4 \| 13.6 | 0.0 \| 13.6 \| 4.5 \| 9.1 | 31.8 \| 50.0 \| 36.4 \| 45.5 | 22.7 \| 4.5 \| 13.6 \| 13.6 | 13.6 \| 4.5 \| 9.1 \| 9.1 | 4.5 \| 13.6 \| 9.1 \| 9.1 |
| Daily mean temperature | 27.3 \| 18.2 \| 18.2 \| 27.3 | - | 13.6 \| 13.6 \| 4.5 \| 22.7 | - | - | - | - |
| Ice day | - | 9.1 \| 13.6 \| 18.2 \| 4.5 | - | - | - | - | - |
| Frost day | - | - | - | - | - | - | - |

| | | | | | | | |
|---|---|---|---|---|---|---|---|
| Wind chill index | - | - | 22.7 \| 18.2 \| 18.2 \| 22.7 | 36.4 \| 31.8 \| 22.7 \| 45.5 | - | 9.1 \| 22.7 \| 13.6 \| 18.2 | - |
| Heat wave | - | - | 13.6 \| 13.6 \| 13.6 \| 13.6 | - | 27.3 \| 13.6 \| 13.6 \| 27.3 | - | - |
| Heat wave day | - | - | - | - | - | - | 27.3 \| 40.9 \| 4.5 \| 63.6 |
| Heat wave duration | - | - | 4.5 \| 13.6 \| 4.5 \| 13.6 | - | - | - | - |
| Heat wave intensity | - | - | - | 9.1 \| 22.7 \| 18.2 \| 13.6 | - | - | - |
| Heat stress wave | - | - | - | - | - | 18.2 \| 27.3 \| 9.1 \| 36.4 | - |
| Heat stress wave day | - | 45.5 \| 22.7 \| 50.0 \| 18.2 | - | - | - | - | - |
| Heat stress wave duration | - | - | - | - | - | - | - |
| Heat stress wave intensity | 9.1 \| 22.7 \| 4.5 \| 27.3 | 22.7 \| 9.1 \| 18.2 \| 13.6 | - | - | - | - | 9.1 \| 9.1 \| 9.1 \| 9.1 |
| Cold wave | - | - | - | - | 13.6 \| 4.5 \| 9.1 \| 9.1 | - | - |
| Cold wave day | - | - | - | - | - | - | - |
| Cold wave duration | - | 0.0 \| 13.6 \| 4.5 \| 9.1 | - | - | - | - | 9.1 \| 9.1 \| 13.6 \| 4.5 |
| Cold wave intensity | 13.6 \| 18.2 \| 22.7 \| 9.1 | - | 9.1 \| 4.5 \| 9.1 \| 4.5 | - | - | - | - |

| | | | | | | | |
|---|---|---|---|---|---|---|---|
| Cold stress wave | - | 4.5 \| 13.6 \| 4.5 \| 13.6 | - | - | 9.1 \| 4.5 \| 0.0 \| 13.6 | - | 22.7 \| 18.2 \| 22.7 \| 18.2 |
| Cold stress wave day | - | - | 27.3 \| 27.3 \| 31.8 \| 22.7 | - | 13.6 \| 0.0 \| 9.1 \| 4.5 | - | - |
| Cold stress wave duration | 4.5 \| 13.6 \| 13.6 \| 4.5 | 9.1 \| 4.5 \| 9.1 \| 4.5 | - | - | - | 13.6 \| 13.6 \| 13.6 \| 13.6 | - |
| Cold stress wave intensity | 4.5 \| 4.5 \| 9.1 \| 0.0 | 4.5 \| 4.5 \| 4.5 \| 4.5 | 4.5 \| 27.3 \| 22.7 \| 9.1 | - | - | - | 9.1 \| 9.1 \| 9.1 \| 9.1 |
| Ozone | 13.6 \| 27.3 \| 22.7 \| 18.2 | - | 9.1 \| 18.2 \| 9.1 \| 18.2 | 36.4 \| 18.2 \| 36.4 \| 18.2 | 9.1 \| 4.5 \| 4.5 \| 9.1 | - | 18.2 \| 18.2 \| 22.7 \| 13.6 |
| Nitrogen dioxide ($NO_2$) | - | - | - | - | - | 9.1 \| 18.2 \| 18.2 \| 9.1 | - |
| Particular matter ($PM_{10}$) | 36.4 \| 4.5 \| 36.4 \| 4.5 | - | - | - | - | 13.6 \| 22.7 \| 22.7 \| 13.6 | - |
| Particular matter ($PM_{2.5}$) | - | - | 36.4 \| 36.4 \| 50.0 \| 22.7 | - | - | - | 22.7 \| 31.8 \| 22.7 \| 31.8 |

**Table S24.3.** Sex- and age-specific significant associations between daily intensive care unit admissions and atmospheric factors in Germany per respiratory, internal and traumatic outcome type. Significant percentage *(%)* of all 22 possible models (2 sex/age subgroups x 11 human-bioclimatic regions), order of the presented values: Females, males, <60 years, ≥60 years

| Atmospheric factor | Pneumonia | Chronic obstructive pulmonary disease (COPD) | Sepsis | Trauma |
|---|---|---|---|---|
| Sea-level pressure (SLP) | 31.8 \| 40.9 \| 31.8 \| 40.9 | 36.4 \| 22.7 \| 22.7 \| 36.4 | 22.7 \| 36.4 \| 31.8 \| 27.3 | 50.0 \| 31.8 \| 31.8 \| 50.0 |
| Sea-level pressure difference (SLPD) | 31.8 \| 40.9 \| 45.5 \| 27.3 | 50.0 \| 45.5 \| 45.5 \| 50.0 | 54.5 \| 36.4 \| 36.4 \| 54.5 | - |
| Wind day | 9.1 \| 22.7 \| 9.1 \| 22.7 | - | 27.3 \| 31.8 \| 36.4 \| 22.7 | - |
| Tropical night | 45.5 \| 50.0 \| 36.4 \| 59.1 | - | - | 27.3 \| 18.2 \| 31.8 \| 13.6 |
| Hot day | - | - | - | 18.2 \| 40.9 \| 31.8 \| 27.3 |
| Summer day | - | - | 40.9 \| 22.7 \| 22.7 \| 40.9 | - |
| Wet-bulb globe temperature | - | 18.2 \| 22.7 \| 27.3 \| 13.6 | 18.2 \| 13.6 \| 18.2 \| 13.6 | 9.1 \| 40.9 \| 36.4 \| 13.6 |
| Daily mean temperature | 18.2 \| 4.5 \| 22.7 \| 0.0 | 18.2 \| 4.5 \| 9.1 \| 13.6 | 13.6 \| 4.5 \| 13.6 \| 4.5 | 0.0 \| 9.1 \| 9.1 \| 0.0 |
| Ice day | - | 22.7 \| 22.7 \| 22.7 \| 22.7 | - | 31.8 \| 27.3 \| 22.7 \| 36.4 |
| Frost day | - | - | - | - |
| Wind chill index | 9.1 \| 9.1 \| 13.6 \| 4.5 | - | 4.5 \| 9.1 \| 4.5 \| 9.1 | 9.1 \| 0.0 \| 9.1 \| 0.0 |
| Heat wave | - | - | - | 4.5 \| 22.7 \| 9.1 \| 18.2 |

| | | | | |
|---|---|---|---|---|
| Heat wave day | - | - | 31.8 \| 40.9 \| 18.2 \| 54.5 | 4.5 \| 22.7 \| 4.5 \| 22.7 |
| Heat wave duration | - | - | - | - |
| Heat wave intensity | - | 9.1 \| 22.7 \| 9.1 \| 22.7 | - | 13.6 \| 22.7 \| 18.2 \| 18.2 |
| Heat stress wave | - | 36.4 \| 31.8 \| 31.8 \| 36.4 | - | - |
| Heat stress wave day | - | - | - | 0.0 \| 13.6 \| 13.6 \| 0.0 |
| Heat stress wave duration | - | - | - | - |
| Heat stress wave intensity | 27.3 \| 9.1 \| 18.2 \| 18.2 | - | - | - |
| Cold wave | 27.3 \| 40.9 \| 59.1 \| 9.1 | 22.7 \| 27.3 \| 31.8 \| 18.2 | - | - |
| Cold wave day | - | - | - | - |
| Cold wave duration | 13.6 \| 27.3 \| 18.2 \| 22.7 | 13.6 \| 13.6 \| 9.1 \| 18.2 | - | 36.4 \| 36.4 \| 50.0 \| 22.7 |
| Cold wave intensity | 9.1 \| 4.5 \| 4.5 \| 9.1 | 13.6 \| 9.1 \| 9.1 \| 13.6 | - | - |
| Cold stress wave | - | 31.8 \| 9.1 \| 18.2 \| 22.7 | - | - |
| Cold stress wave day | - | - | - | - |
| Cold stress wave duration | 18.2 \| 18.2 \| 9.1 \| 27.3 | - | - | - |
| Cold stress wave intensity | 18.2 \| 18.2 \| 18.2 \| 18.2 | 13.6 \| 4.5 \| 9.1 \| 9.1 | 18.2 \| 4.5 \| 13.6 \| 9.1 | 13.6 \| 9.1 \| 18.2 \| 4.5 |
| Ozone | 40.9 \| 18.2 \| 22.7 \| 36.4 | 9.1 \| 9.1 \| 9.1 \| 9.1 | - | 13.6 \| 18.2 \| 9.1 \| 22.7 |

| | | | | |
|---|---|---|---|---|
| Nitrogen dioxide ($NO_2$) | 27.3 \| 18.2 \| 27.3 \| 18.2 | - | - | 13.6 \| 36.4 \| 31.8 \| 18.2 |
| Particular matter ($PM_{10}$) | - | 18.2 \| 9.1 \| 22.7 \| 4.5 | - | 22.7 \| 0.0 \| 9.1 \| 13.6 |
| Particular matter ($PM_{2.5}$) | - | - | - | 22.7 \| 18.2 \| 18.2 \| 22.7 |

**Table S25.1.** Weighted explained deviance (WED, %) across sex-, age-specific subgroups per human-bioclimatic (HBC) region and cardiovascular outcome type

| HBC-region | Myocardial infarction (STEMI) | Myocardial infarction (NSTEMI) | Heart failure | Pulmonary artery embolism | Tachycardia |
|---|---|---|---|---|---|
| 1.1 | 6 | 40 | 8 | 2 | 15 |
| 1.2 | 8 | 46 | 9 | 3 | 16 |
| 2.1 | 6 | 46 | 15 | 3 | 22 |
| 2.2 | 7 | 45 | 14 | 3 | 20 |
| 3.1 | 2 | 27 | 7 | 3 | 9 |
| 4.1 | 4 | 29 | 5 | 3 | 8 |
| 5.1 | 7 | 34 | 9 | 3 | 12 |
| 5.2 | 3 | 29 | 6 | 3 | 7 |
| 6.1 | 4 | 10 | 4 | 5 | 4 |
| 6.2 | 12 | 49 | 13 | 3 | 18 |
| 6.3 | 2 | 15 | 3 | 3 | 5 |

**Table S25.2.** Weighted explained deviance (WED, %) across subgroups per human-bioclimatic (HBC) region and neurological outcome type

| HBC-region | Ischemic stroke | Ischemic stroke (stroke unit) | Epilepsy | Non subdural hemorrhage | Non subdural hemorrhage (stroke unit) | Transient ischemic attack | Transient ischemic attack (stroke unit) |
|---|---|---|---|---|---|---|---|
| 1.1 | 3 | 12 | 2 | 2 | 1 | 2 | 7 |
| 1.2 | 5 | 17 | 2 | 2 | 2 | 3 | 10 |
| 2.1 | 9 | 23 | 3 | 3 | 3 | 4 | 14 |
| 2.2 | 7 | 16 | 2 | 3 | 2 | 2 | 9 |
| 3.1 | 4 | 14 | 1 | 2 | 2 | 2 | 6 |
| 4.1 | 4 | 5 | 1 | 2 | 2 | 2 | 5 |
| 5.1 | 4 | 8 | 2 | 3 | 2 | 2 | 6 |
| 5.2 | 3 | 6 | 2 | 2 | 2 | 2 | 3 |
| 6.1 | 3 | 2 | 2 | 3 | 5 | 4 | 3 |
| 6.2 | 9 | 14 | 2 | 5 | 2 | 4 | 9 |
| 6.3 | 4 | 5 | 2 | 2 | 3 | 5 | 3 |

**Table S25.3.** Weighted explained deviance (WED, %) across subgroups per human-bioclimatic (HBC) region and respiratory, internal and traumatic outcome type

| HBC-region | Pneumonia | Chronic obstructive pulmonary disease (COPD) | Sepsis | Trauma |
|---|---|---|---|---|
| 1.1 | 16 | 12 | 13 | 27 |
| 1.2 | 25 | 20 | 15 | 33 |
| 2.1 | 25 | 13 | 16 | 41 |
| 2.2 | 23 | 16 | 14 | 38 |
| 3.1 | 13 | 8 | 6 | 23 |
| 4.1 | 8 | 8 | 7 | 19 |
| 5.1 | 19 | 12 | 9 | 26 |
| 5.2 | 14 | 5 | 6 | 18 |
| 6.1 | 5 | 6 | 4 | 11 |
| 6.2 | 26 | 14 | 14 | 38 |
| 6.3 | 8 | 6 | 5 | 12 |

**Table S261** Explained deviance (ED, %) across sex-, age-specific subgroups per human-bioclimatic (HBC) region and cardiovascular outcome type, order of the presented values: Female <60 years, Female ≥60 years, Male <60 years, Male ≥60 years

| HBC-region | Myocardial infarction (STEMI) | Myocardial infarction (NSTEMI) | Heart failure | Pulmonary artery embolism | Tachycardia |
|---|---|---|---|---|---|
| 1.1 | 1 \| 10 \| 2 \| 7 | 10 \| 42 \| 25 \| 49 | 1 \| 11 \| 2 \| 6 | 3 \| 2 \| 2 \| 2 | 4 \| 19 \| 7 \| 18 |
| 1.2 | 2 \| 12 \| 5 \| 8 | 17 \| 49 \| 33 \| 54 | 2 \| 11 \| 4 \| 9 | 3 \| 4 \| 3 \| 3 | 4 \| 21 \| 7 \| 17 |
| 2.1 | 1 \| 1 \| 1 \| 8 | 11 \| 44 \| 32 \| 57 | 2 \| 16 \| 4 \| 16 | 2 \| 4 \| 3 \| 4 | 6 \| 26 \| 11 \| 26 |
| 2.2 | 1 \| 10 \| 2 \| 9 | 1 \| 48 \| 26 \| 54 | 2 \| 16 \| 4 \| 13 | 2 \| 4 \| 2 \| 4 | 5 \| 25 \| 8 \| 24 |
| 3.1 | 1 \| 3 \| 1 \| 2 | 5 \| 24 \| 13 \| 35 | 3 \| 7 \| 3 \| 7 | 3 \| 3 \| 4 \| 3 | 3 \| 10 \| 5 \| 12 |
| 4.1 | 1 \| 6 \| 1 \| 5 | 5 \| 29 \| 13 \| 36 | 2 \| 5 \| 3 \| 5 | 2 \| 3 \| 3 \| 3 | 3 \| 10 \| 4 \| 9 |
| 5.1 | 1 \| 9 \| 3 \| 8 | 5 \| 34 \| 18 \| 42 | 3 \| 10 \| 3 \| 9 | 2 \| 3 \| 4 \| 3 | 3 \| 14 \| 5 \| 14 |
| 5.2 | 1 \| 4 \| 2 \| 3 | 4 \| 30 \| 14 \| 35 | 2 \| 8 \| 3 \| 5 | 5 \| 3 \| 3 \| 3 | 3 \| 9 \| 4 \| 8 |
| 6.1 | 3 \| 5 \| 2 \| 4 | 2 \| 10 \| 6 \| 12 | 3 \| 6 \| 3 \| 3 | 8 \| 4 \| 7 \| 3 | 4 \| 4 \| 2 \| 4 |
| 6.2 | 2 \| 17 \| 2 \| 14 | 9 \| 56 \| 25 \| 56 | 2 \| 17 \| 4 \| 11 | 2 \| 5 \| 3 \| 2 | 3 \| 22 \| 9 \| 21 |
| 6.3 | 2 \| 3 \| 1 \| 2 | 3 \| 15 \| 8 \| 20 | 5 \| 3 \| 3 \| 4 | 7 \| 2 \| 5 \| 3 | 2 \| 5 \| 3 \| 6 |

**Table S262** Explained deviance (ED, %) across subgroups per human-bioclimatic (HBC) region and neurological outcome type, order of the presented values: Female <60 years, Female ≥60 years, Male <60 years, Male ≥60 years

| HBC-region | Ischemic stroke | Ischemic stroke (stroke unit) | Epilepsy | Non subdural hemorrhage | Non subdural hemorrhage (stroke unit) | Transient ischemic attack | Transient ischemic attack (stroke unit) |
|---|---|---|---|---|---|---|---|
| 1.1 | 2 \| 3 \| 2 \| 4 | 3 \| 10 \| 7 \| 16 | 2 \| 1 \| 4 \| 1 | 2 \| 2 \| 2 \| 2 | 2 \| 1 \| 2 \| 1 | 2 \| 2 \| 3 \| 2 | 3 \| 9 \| 7 \| 7 |
| 1.2 | 3 \| 5 \| 5 \| 6 | 4 \| 16 \| 11 \| 22 | 2 \| 2 \| 2 \| 3 | 1 \| 2 \| 1 \| 2 | 2 \| 2 \| 2 \| 2 | 4 \| 3 \| 4 \| 3 | 5 \| 11 \| 8 \| 10 |
| 2.1 | 3 \| 1 \| 4 \| 10 | 4 \| 23 \| 10 \| 28 | 2 \| 2 \| 4 \| 3 | 2 \| 4 \| 2 \| 3 | 2 \| 4 \| 1 \| 3 | 2 \| 5 \| 3 \| 4 | 4 \| 16 \| 6 \| 15 |
| 2.2 | 2 \| 7 \| 4 \| 9 | 3 \| 15 \| 8 \| 21 | 2 \| 2 \| 2 \| 1 | 2 \| 4 \| 1 \| 2 | 4 \| 3 \| 2 \| 2 | 3 \| 3 \| 3 \| 2 | 5 \| 11 \| 6 \| 9 |
| 3.1 | 3 \| 4 \| 2 \| 5 | 3 \| 14 \| 5 \| 16 | 1 \| 2 \| 2 \| 1 | 1 \| 2 \| 2 \| 2 | 3 \| 2 \| 2 \| 1 | 3 \| 2 \| 2 \| 2 | 3 \| 7 \| 5 \| 6 |
| 4.1 | 4 \| 3 \| 2 \| 5 | 2 \| 5 \| 3 \| 6 | 2 \| 1 \| 1 \| 2 | 2 \| 2 \| 1 \| 2 | 6 \| 2 \| 2 \| 2 | 3 \| 2 \| 3 \| 2 | 3 \| 6 \| 3 \| 4 |
| 5.1 | 2 \| 5 \| 2 \| 5 | 2 \| 7 \| 4 \| 11 | 2 \| 1 \| 3 \| 1 | 1 \| 4 \| 2 \| 2 | 1 \| 2 \| 2 \| 1 | 3 \| 2 \| 3 \| 2 | 3 \| 6 \| 5 \| 5 |
| 5.2 | 3 \| 3 \| 2 \| 4 | 3 \| 5 \| 2 \| 8 | 2 \| 1 \| 2 \| 2 | 2 \| 2 \| 3 \| 2 | 2 \| 2 \| 2 \| 2 | 3 \| 2 \| 3 \| 2 | 3 \| 3 \| 3 \| 3 |
| 6.1 | 5 \| 3 \| 2 \| 3 | 1 \| 3 \| 1 \| 2 | 3 \| 2 \| 2 \| 2 | 9 \| 2 \| 6 \| 2 | 22 \| 4 \| 12 \| 3 | 8 \| 4 \| 8 \| 3 | 6 \| 2 \| 4 \| 2 |
| 6.2 | 2 \| 1 \| 4 \| 10 | 3 \| 15 \| 7 \| 15 | 2 \| 2 \| 3 \| 2 | 2 \| 6 \| 2 \| 5 | 2 \| 2 \| 2 \| 2 | 2 \| 5 \| 2 \| 2 | 4 \| 11 \| 5 \| 6 |
| 6.3 | 2 \| 3 \| 3 \| 6 | 2 \| 5 \| 3 \| 5 | 2 \| 1 \| 2 \| 2 | 4 \| 1 \| 2 \| 1 | 9 \| 2 \| 2 \| 2 | 6 \| 5 \| 4 \| 4 | 3 \| 3 \| 4 \| 3 |

**Table S263** Explained deviance (ED, %) across subgroups per human-bioclimatic (HBC) region and respiratory, internal and traumatic outcome type, order of the presented values: Female <60 years, Female ≥60 years, Male <60 years, Male ≥60 years

| HBC-region | Pneumonia | Chronic obstructive pulmonary disease (COPD) | Sepsis | Trauma |
|---|---|---|---|---|
| 1.1 | 18 \| 13 \| 18 \| 15 | 5 \| 17 \| 7 \| 11 | 5 \| 9 \| 9 \| 19 | 12 \| 34 \| 13 \| 35 |
| 1.2 | 27 \| 20 \| 3 \| 22 | 6 \| 25 \| 8 \| 22 | 6 \| 13 \| 8 \| 22 | 20 \| 36 \| 21 \| 43 |
| 2.1 | 24 \| 22 \| 28 \| 24 | 3 \| 16 \| 5 \| 13 | 4 \| 14 \| 8 \| 22 | 19 \| 48 \| 21 \| 51 |
| 2.2 | 23 \| 2 \| 25 \| 24 | 5 \| 20 \| 5 \| 18 | 4 \| 11 \| 8 \| 2 | 17 \| 45 \| 18 \| 47 |
| 3.1 | 15 \| 12 \| 16 \| 10 | 4 \| 9 \| 5 \| 8 | 2 \| 5 \| 3 \| 9 | 7 \| 27 \| 9 \| 30 |
| 4.1 | 11 \| 6 \| 9 \| 6 | 3 \| 10 \| 4 \| 8 | 2 \| 6 \| 4 \| 9 | 5 \| 21 \| 8 \| 27 |
| 5.1 | 18 \| 17 \| 19 \| 2 | 3 \| 15 \| 5 \| 13 | 2 \| 8 \| 4 \| 12 | 7 \| 32 \| 7 \| 35 |
| 5.2 | 18 \| 11 \| 16 \| 13 | 4 \| 6 \| 2 \| 4 | 3 \| 5 \| 3 \| 9 | 5 \| 24 \| 4 \| 23 |
| 6.1 | 8 \| 4 \| 6 \| 4 | 5 \| 6 \| 5 \| 6 | 2 \| 4 \| 3 \| 5 | 2 \| 15 \| 5 \| 12 |
| 6.2 | 23 \| 24 \| 27 \| 28 | 6 \| 16 \| 6 \| 16 | 5 \| 12 \| 8 \| 20 | 13 \| 48 \| 16 \| 45 |
| 6.3 | 9 \| 8 \| 8 \| 7 | 4 \| 7 \| 5 \| 6 | 3 \| 4 \| 3 \| 6 | 5 \| 14 \| 3 \| 16 |

#### Supplementary Figures

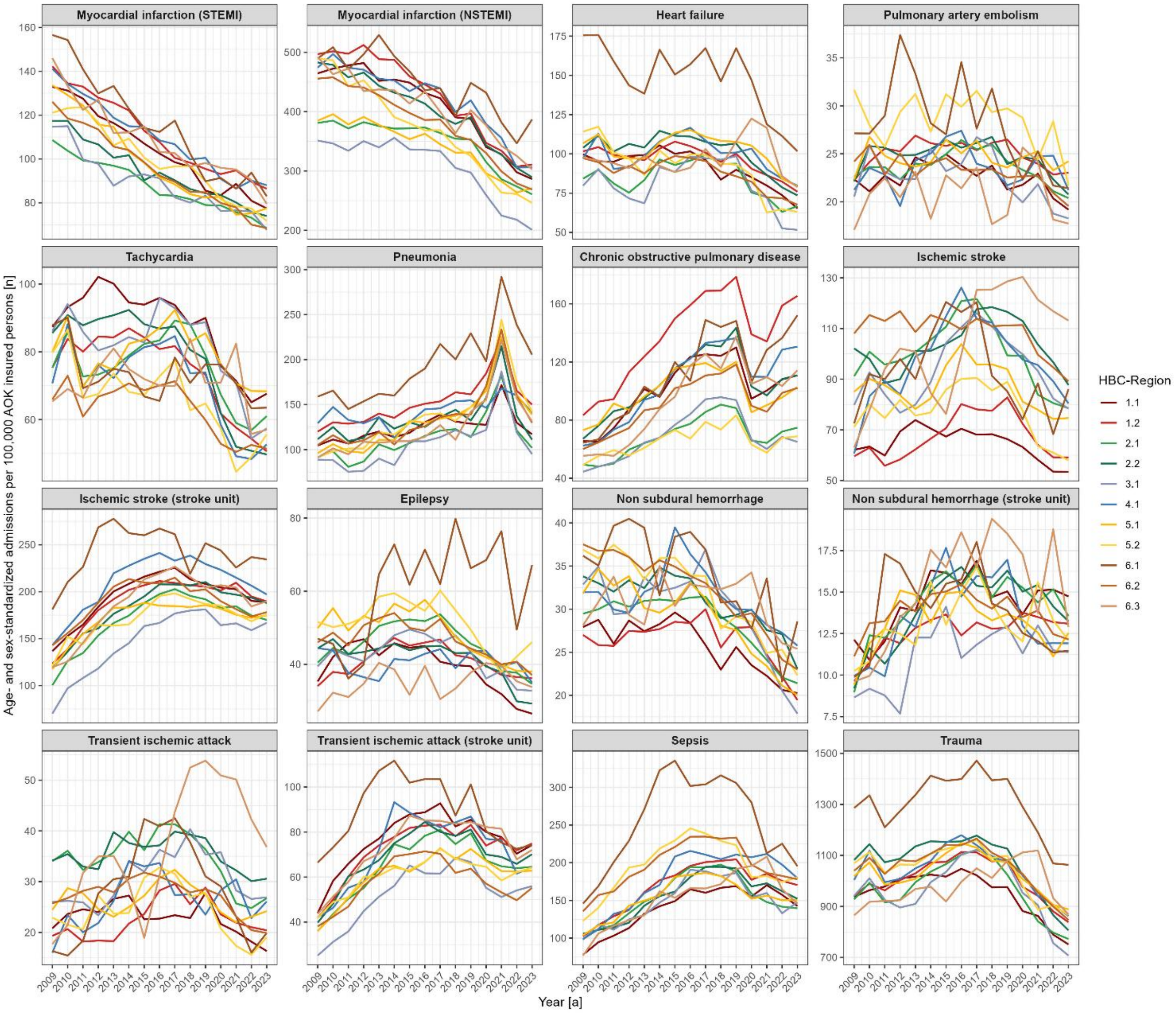


**Figure S1.** Annual intensive care unit (ICU) admissions per 100,000 AOK insured persons by outcome type and human-bioclimatic (HBC) region, 2009–2023, standardized for age and sex to the AOK insured population. Each panel shows one outcome type, lines represent HBC-regions.

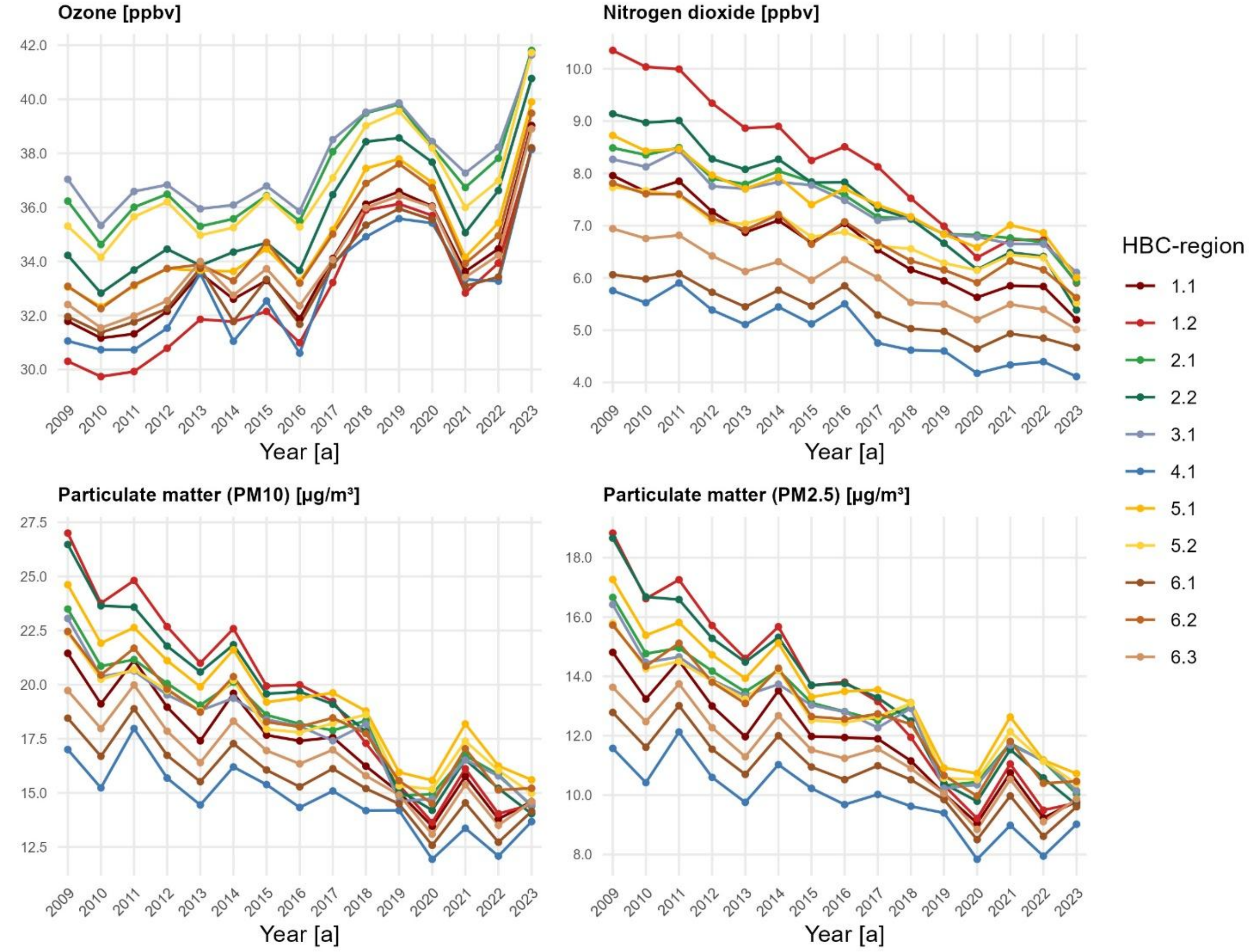


**Figure S2.** Annually air pollution factors per human-bioclimatic (HBC) region

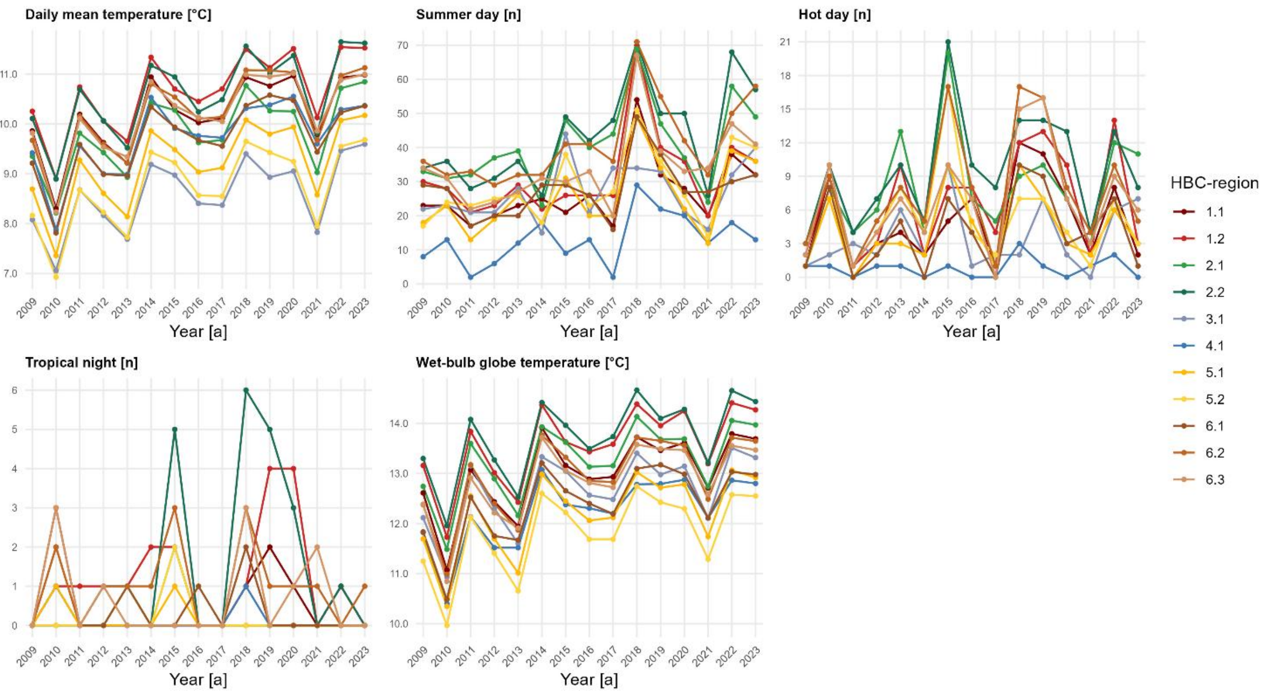


**Figure S3.** Annually heat-related factors (raw, binary or indices) per human-bioclimatic (HBC) region

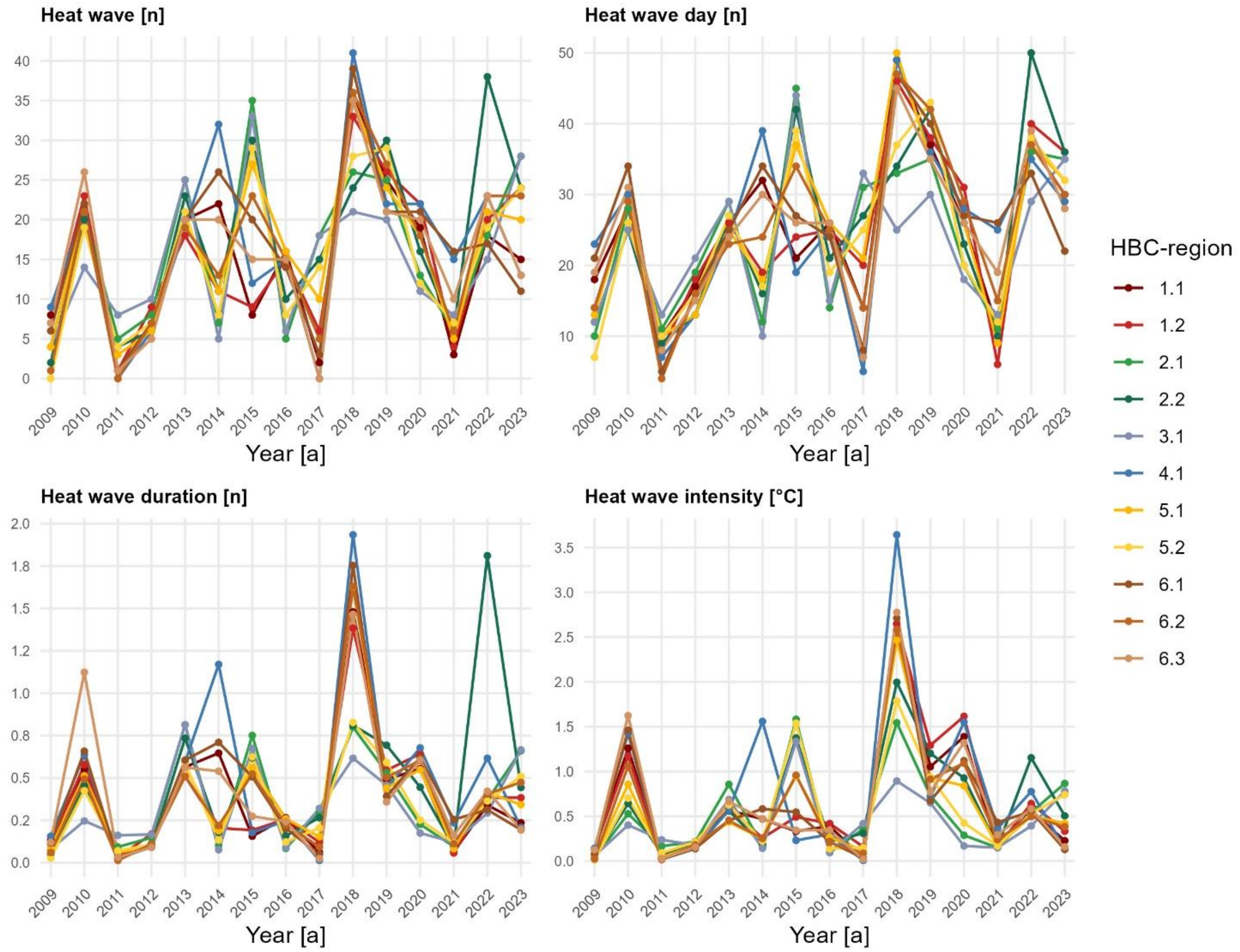


**Figure S4.** Annually heat wave-related factors per human-bioclimatic (HBC) region

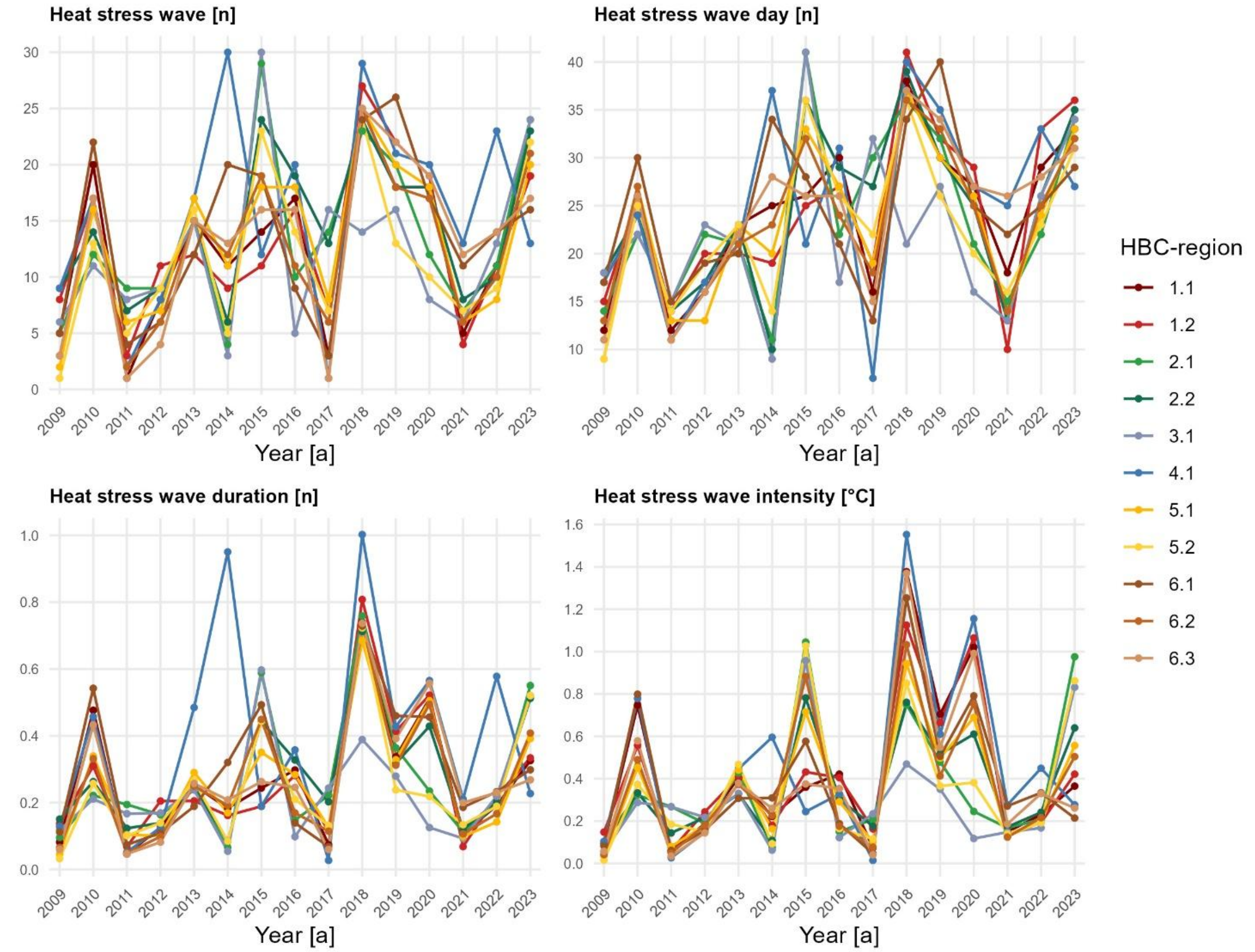


**Figure S5.** Annually heat stress wave-related factors per human-bioclimatic (HBC) region

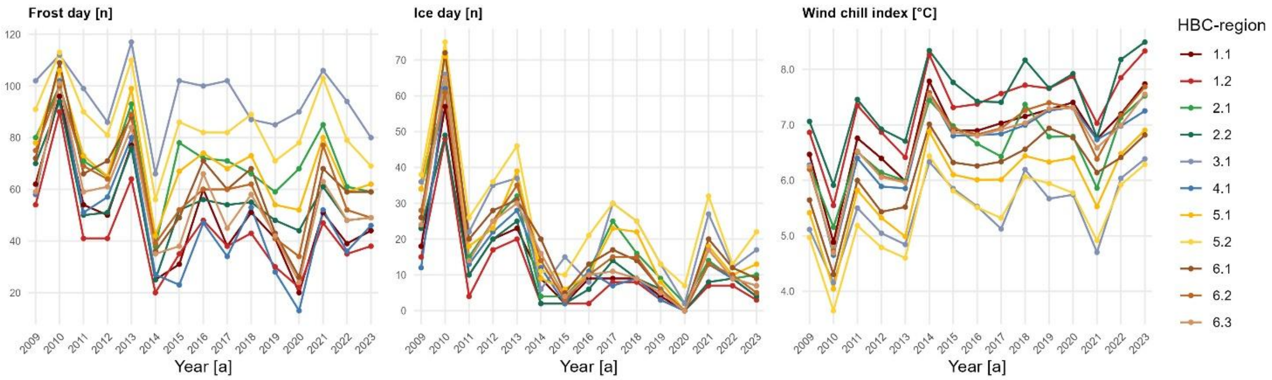


**Figure S6.** Annually cold-related factors (binary or indices) per human-bioclimatic (HBC) region

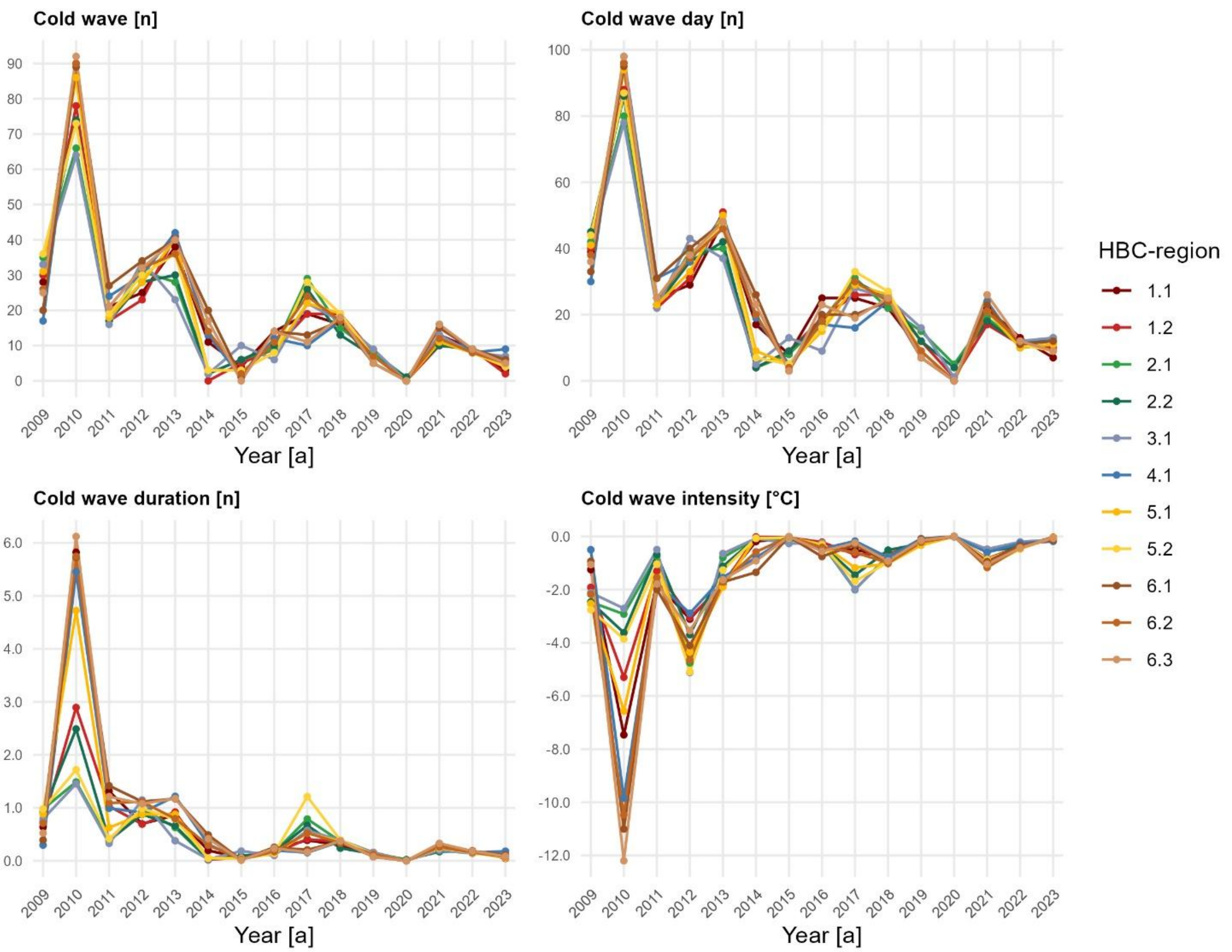


**Figure S7.** Annually cold wave-related factors per human-bioclimatic (HBC) region

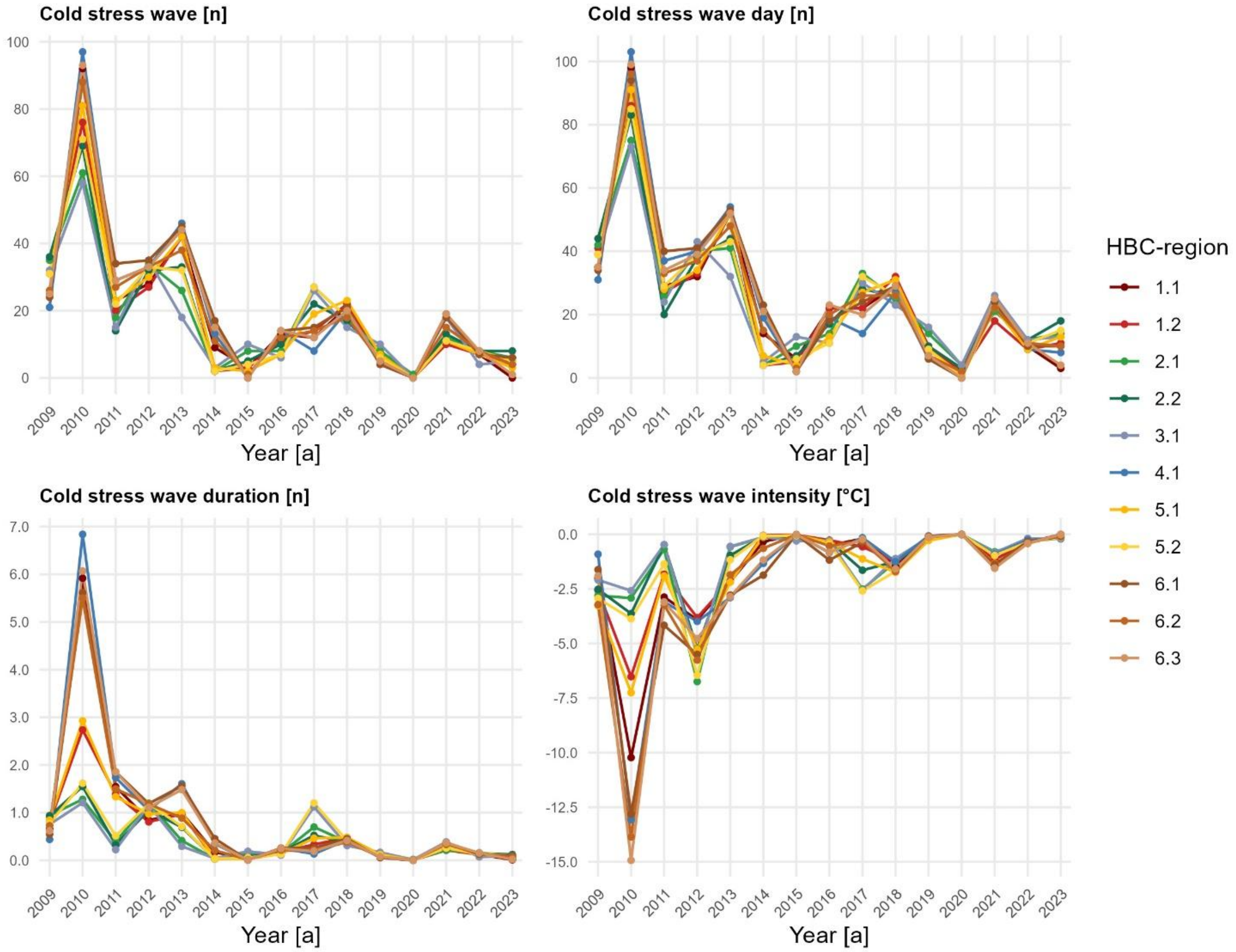


**Figure S8.** Annually cold stress wave-related factors per human-bioclimatic (HBC) region

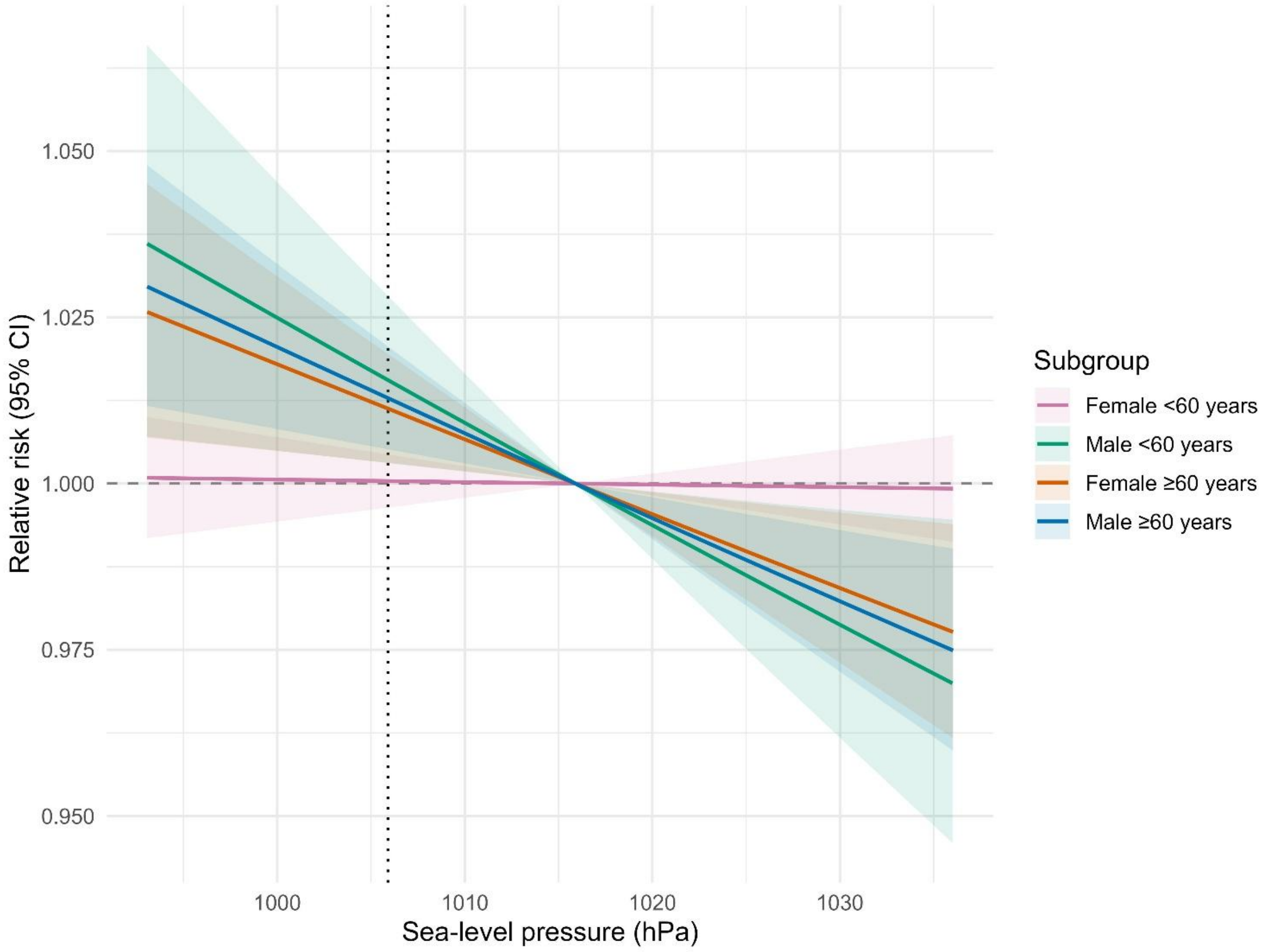


**Figure S9.** Subgroup-specific exposure-response curve between sea-level pressure (SLP) and myocardial infarction (NSTEMI) intensive care unit admissions in human-bioclimatic (HBC) region 6.2, dashed line = point of comparison (relative risk calculation)

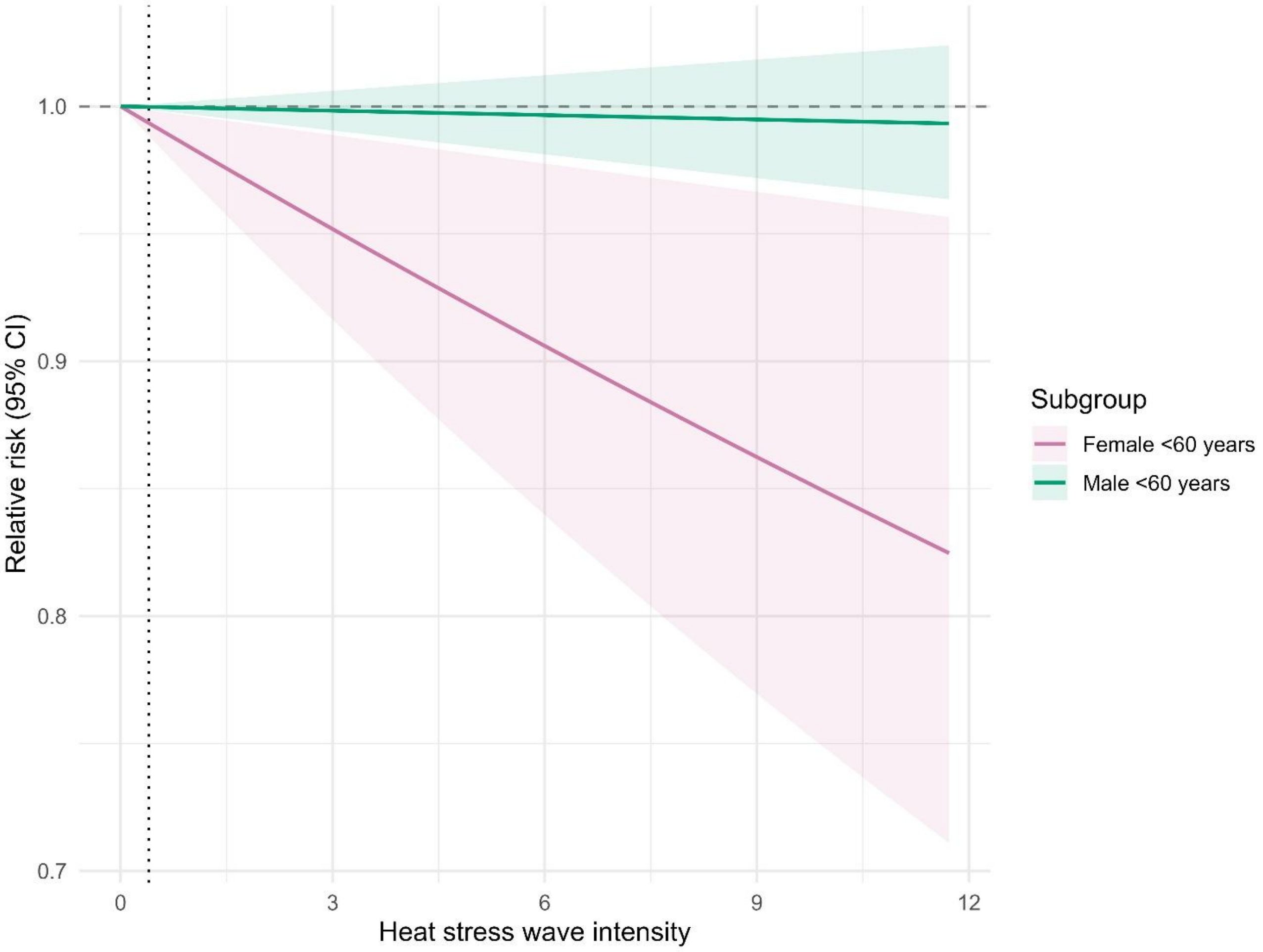


**Figure S10.** Subgroup-specific exposure-response curve between heat stress wave intensity (°C) and myocardial infarction (NSTEMI) intensive care unit admissions in human-bioclimatic (HBC) region 6.2, dashed line = point of comparison (relative risk calculation)

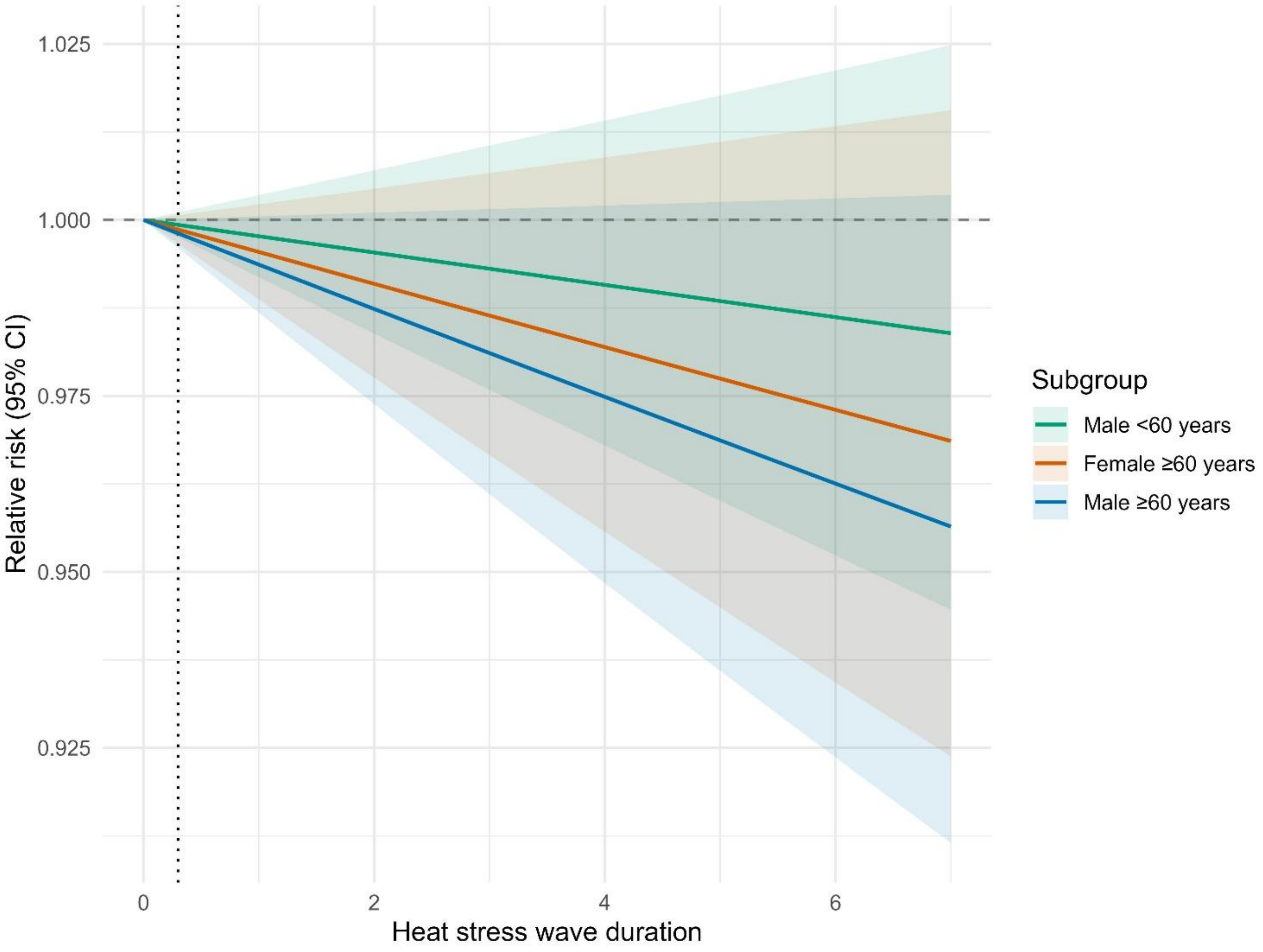


**Figure S11.** Subgroup-specific exposure-response curve between heat stress wave duration (days) and myocardial infarction (NSTEMI) intensive care unit admissions in human-bioclimatic (HBC) region 6.2, dashed line = point of comparison (relative risk calculation)

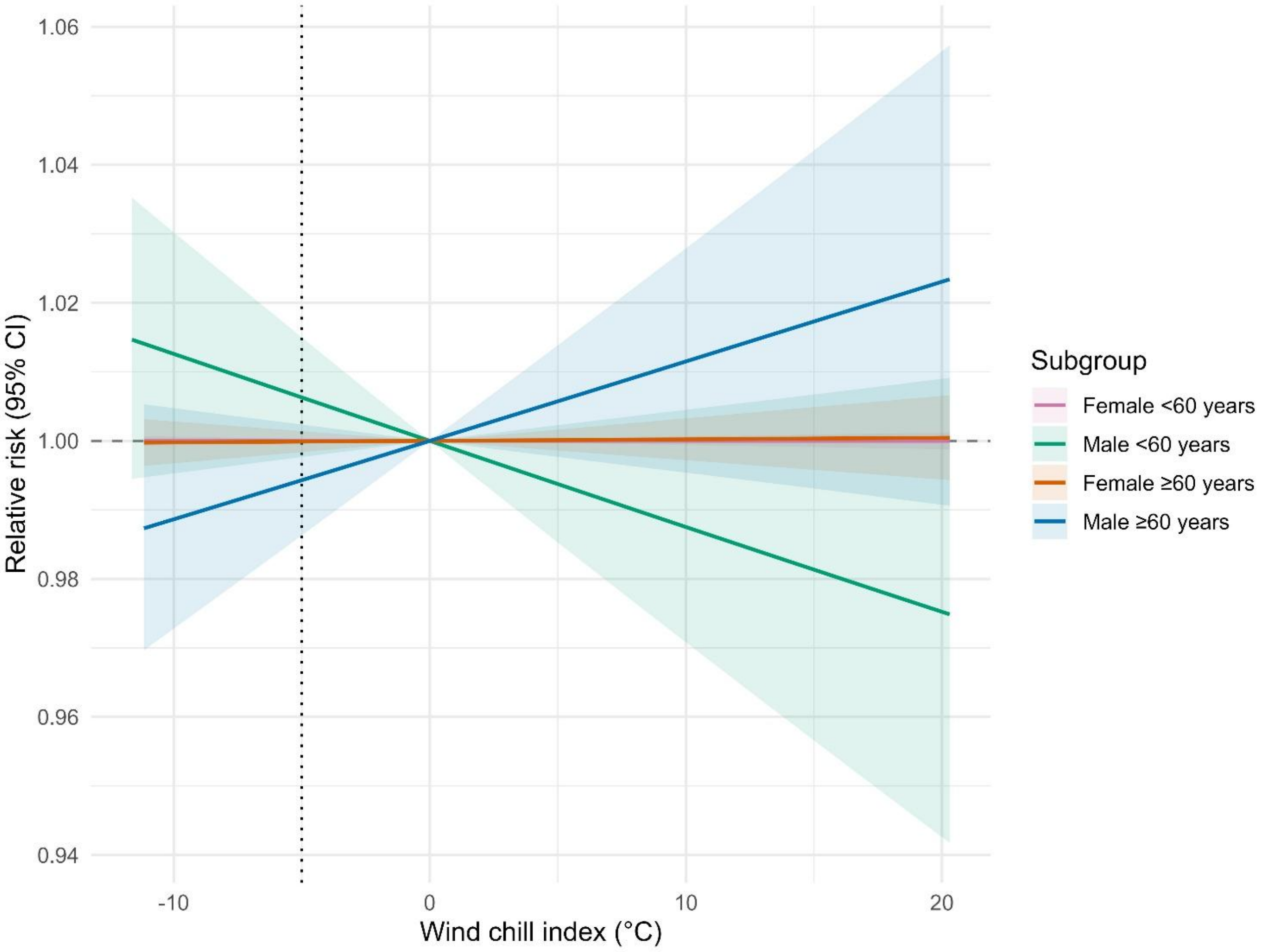


**Figure S12.** Subgroup-specific exposure-response curve between Wind chill index (WCI) and myocardial infarction (NSTEMI) intensive care unit admissions in human-bioclimatic (HBC) region 6.2, dashed line = point of comparison (relative risk calculation)

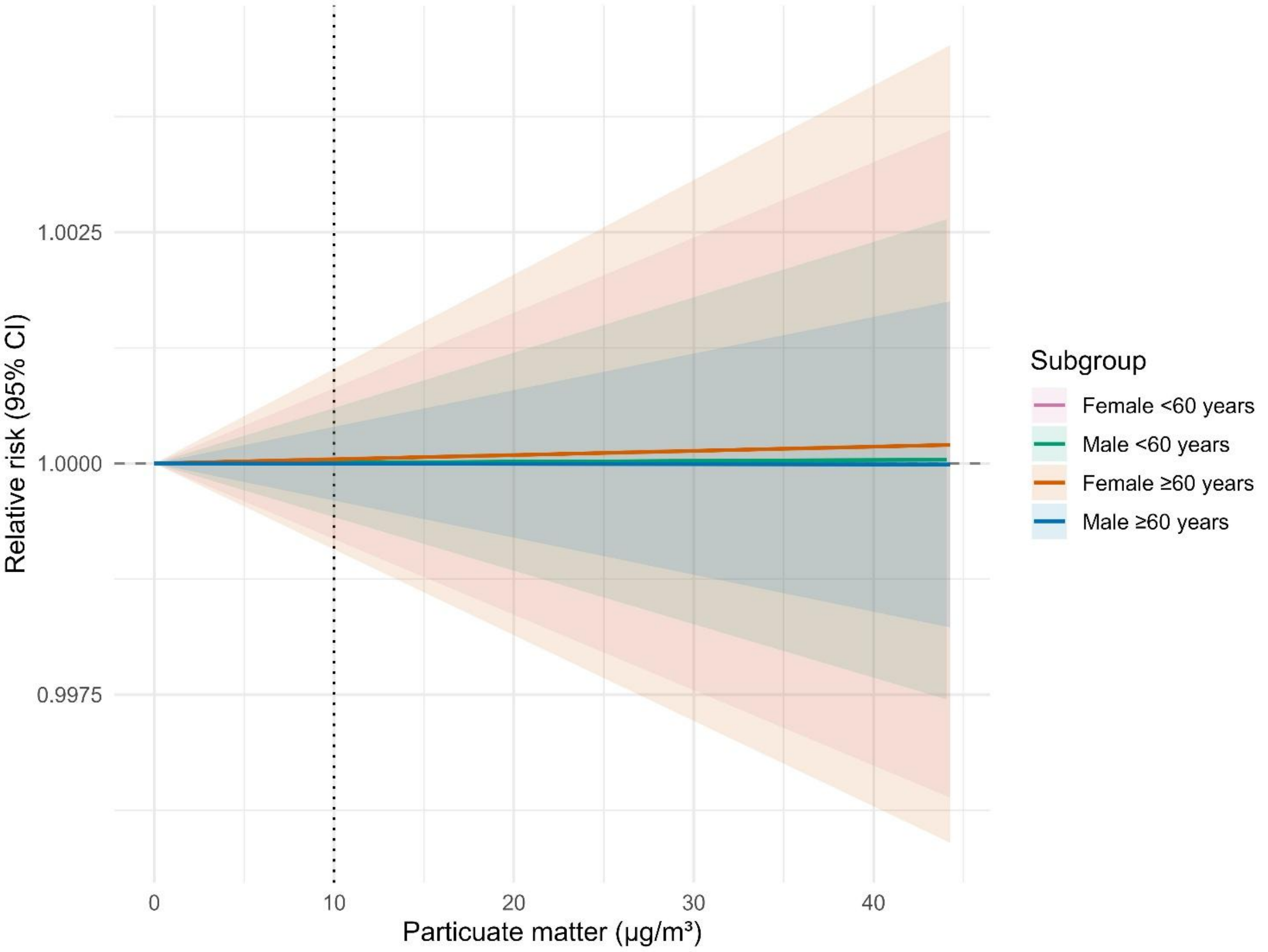


**Figure S13.** Subgroup-specific exposure-response curve between Particulate matter ($PM_{10}$) and myocardial infarction (NSTEMI) intensive care unit admissions in human-bioclimatic (HBC) region 6.2, dashed line = point of comparison (relative risk calculation)
